# Commercial or industrial use of mental health data for research: primer and best-practice guidelines from the DATAMIND patient/public Lived Experience Advisory Group

**DOI:** 10.1101/2025.11.05.25339575

**Authors:** Linda A. Jones, Corrine Barber, Naomi Clements-Brod, Jan Davies, Marcos DelPozo-Banos, Andrea Hughes, Matthew H. Iveson, Diksha Jain, Ann John, Aideen Maguire, Clara Martins de Barros, Andrew M. McIntosh, Michael McTernan, Jan Speechley, Robert J. Stewart, Jane Taylor, Louise Ting, Rudolf N. Cardinal

**Author notes:** Pseudonym; identity available to editor (Diksha Jain).

## Abstract

1

**BACKGROUND:** Routinely collected health data, such as that held by United Kingdom (UK) national health services (NHS), has important research uses. However, its use requires public trust and transparency. Access by commercial/industrial organisations is especially sensitive for the public, as is mental health (MH) data. Although existing MH data science guidelines emphasise patient/public involvement (PPI), they do not cover commercial uses specifically.

**OBJECTIVES:** To develop patient- and public-led guidelines for the commercial and industrial use of MH data for research. Though UK-focused, their principles may apply internationally.

**METHODS:** A PPI lived experience advisory group (LEAG) was created within DATAMIND, a UK data hub for MH informatics. Initial discussion yielded a requirement for definitions and explanations of concepts relating to MH data research, developed iteratively. Subsequently, the LEAG developed guidelines via a qualitative quasi-Delphi approach. The agreed scope excluded data provided for research with informed consent, data processing arrangements (e.g. companies hosting electronic systems on the instruction of health services), and compliance with legal minimum requirements. The scope included the use of routinely collected MH data for research by commercial/industrial organisations without explicit consent, and aspects of industry-led MH data collection conducted with consent.

**RESULTS:** Alongside the primer in MH data research concepts, the LEAG provide best-practice guidelines relating to commercial/industrial research use of MH data, for organisations controlling MH data (such as NHS bodies) and for commercial applicants seeking access. Core principles include transparency, patient rights, meaningful PPI, stringent governance, and statistical disclosure control. The guidelines recommend a risk–benefit approach to assessing data access applications, within limits that include avoiding the export of unconsented patient-level data outside NHS-controlled secure data environments, and not providing commercial applicants with access to unconsented free-text MH data. Further recommendations for NHS executive and regulatory bodies relate to public choice and transparency, clarity of guidance to research-active NHS organisations, and support for de-identification.

**CONCLUSIONS:** MH data research requires patient/public involvement and understanding. These guidelines reflect the views of people with personal or family experience of mental ill health. We hope they are useful to the MH research community and increase public transparency and trust.

**PLAIN-LANGUAGE SUMMARY:** The NHS collects health information that can help with research. This includes information about people’s mental health. But people often worry about how this data is used, especially if companies (not just doctors or university researchers) want to use it.

A group of patients, carers, and members of the public worked together with mental health researchers to make new guidelines. These guidelines help make sure mental health data is used safely and fairly, especially by companies doing research.

The group learned about how mental health data is collected and used. Then they wrote guidelines based on what they thought was right and fair. These guidelines say that:

- People should be told clearly how their data is being used.
- Companies should not get private notes (text) unless people have agreed.
- Patients’ mental health data should stay safe in NHS systems.
- Patients and the public should help decide how data is used.

They also gave advice to NHS leaders to help protect people’s privacy and make things clearer.

These rules were written by people with real experience of mental health problems. The aim is to protect people’s rights and privacy and to build trust while still helping important research to happen.

## 3 INTRODUCTION

Data collected and created as part of routine health care has an important part to play in health research, alongside data collected specifically for research purposes. Many health systems have mechanisms to use routinely collected data for research, as permitted by relevant legislation and research governance structures. In the United Kingdom (UK), the National Health Services (NHS) in England, Scotland, and Wales and Health and Social Care (HSC) in Northern Ireland (collectively “NHS”) strongly support the use of anonymised, routinely collected health data for research without explicit consent (1,2), and the legal basis for such work is clear (3–6). A great deal of research is conducted in this way [e.g. (7)], and the importance of this field is likely to grow. For example, the UK Government has emphasised health data in its science strategy (8) and has been reviewing data protection legislation to support the analysis of data (9).

However, appropriate use of routinely collected health data also requires public trust, or a “social licence” (10). The processes governing research must be transparent and reflect patients’ views (11–13). “Data science” can be complex, emphasising the importance of publicly accessible information about health data and how it is used (14).

One particular area of controversy or sensitivity is around commercial use of routinely collected health data. Such uses, and proposals for use, feature regularly in the UK media [e.g. (15)], and the UK government is seeking to develop a pricing model for NHS data (16). There is public wariness about commercial access to healthcare data. This has been linked in part to limited public awareness about how data is typically used and safeguarded (17), though that is not the only reason (6). There is also considerable variation in what is meant by “commercial use”. In the present work, we exclude the many common situations where the NHS pays a company to store or handle data, such as by providing computers, e-mail systems, or electronic health records systems, but where the NHS remains solely responsible for how the data is used. We consider instead situations where companies have active interests in the data for research. Even here, the spectrum is broad (discussed later). Past use of NHS data for research beyond healthcare has been particularly controversial, such as the use of de-identified hospital episode data, together with “geodemographic” profiling based on postcode, for actuarial (insurance) research (18–20). Our previous work has demonstrated strong UK public support for research using their de-identified health data by the NHS, academic institutions, and health charities; ambivalence around such use by profit-making companies researching treatments; and strong net opposition to access by other kinds of profit-making companies (6). The UK’s national institute for data science, Health Data Research UK (HDR UK), supports several research hubs covering different disease areas, including the mental health (MH) hub, DATAMIND (DATA Hub for Mental health INformatics research Development), funded by the UK Medical Research Council (MRC). HDR UK has consulted the public about commercial access to data (21), set out plans to develop a commercial model for data access (22,23), and provided generic advice for considering commercial requests (24,25). There are operational commercial services for some HDR UK hubs (26).

The question of commercial access to data also relates to national data “pooling”: assembling and linking data from many parts of a health service to improve service evaluation and/or research. In the UK, national data pooling was successful in 2020 onwards to support the response to the COVID-19 pandemic, relying on emergency regulations for the handling of data [e.g. (27)]. However, before this, the England-wide data pooling initiative “care.data” foundered (10,28) and another, General Practice Data for Planning and Research (GPDPR), has been paused (29–31). Concerns and uncertainty about plans for commercial access to NHS data played a significant role in the stopping of these initiatives (10,28,29), and soon after “care.data” was abandoned, Understanding Patient Data was set up as an independent body funded by a medical research charity, UK research councils, and government (32). The 2022 Goldacre review (33) recommended a small number of NHS trusted research environments (TREs) for hosting sensitive data, advocated a frank conversation about commercial use of NHS data for innovation, and noted the strong privacy safeguards that well-designed TREs can bring. From 2023, a number of UK regional NHS trusted research environments (TREs) or secure data environments (SDEs) are being set up (34), with plans for a national “federated” data platform (FDP) in England (35,36). The Faculty of Clinical Informatics have called for transparency about the FDP (37) in particular, given its scope. Data and Analytics Research Environments UK (DARE UK) recommended, after public consultation, that it should not matter who is using sensitive data (e.g. people working for academia, industry, or government) so long as the research is in the public interest—a judgement that should be made involving the public—and the end-to-end process is transparent, with strong governance processes and rigorous vetting and monitoring of researchers (21). However, this consultation did not focus on MH data and we consider some other limitations in the present study. A related concept connected to TREs is that of Data Trusts, which are organisations designed to manage data for the benefits of the data subjects (38). The potential for Data Trusts is being explored within healthcare (39): patients do not legally “own” data about them held by services such as the NHS (40), but do have rights relating to the data (3).

There are reasons to consider MH data specifically when contemplating the commercialisation of routinely collected data or health data more broadly. MH data is considered more sensitive than some other forms of health data, slightly reducing (on average) people’s willingness for it to be shared (6,41), and consistent with patients’ experience of stigma attached to MH illness (42). Preferences are strongly influenced by the kinds of organisations that might receive data, and by how detailed the data is (6). Personal experience of MH illness is, however, associated with greater willingness (on average) to share MH data for research (6), and mental health research has been underfunded historically compared to many areas of physical health research (43). Best practice for MH data science involves incorporating the views of people with relevant personal experience (44,45). Therefore, in this study, we develop the views of DATAMIND’s patient/public advisors into guidelines applicable to the commercial use of MH data. In doing so, we set out background information concerning health data research, particularly in the context of UK health services, as a primer that the patient/public advisors considered necessary in order to form their recommendations.

## 4 MATERIALS AND METHODS

### 4.1 Research advisory group formation

Funding for DATAMIND was awarded by the UK Medical Research Council (MRC) in May 2021 and commenced in December 2021. National advertising for the DATAMIND supranational (Super) Research Advisory Group (SRAG) membership was launched in November 2021 with a closing date in December 2021; the group was renamed the DATAMIND Lived Experience Advisory Group (LEAG) in 2025. Participants were sought with experience of mental ill health either personally or through a family member/friend, an interest in MH research using health data, and a desire to use their experience to shape research. Prior experience was not required; recruitment proactively sought a range of experience levels and emphasised that all perspectives were welcome. Interviews were held in January 2022, and 13 members were appointed, with 12 proceeding to the guideline development work. No applications were received from Northern Ireland, but all other UK nations were represented. LEAG members spontaneously reported diverse backgrounds, levels of research experience, knowledge about and views on secondary uses of data, personal experience of MH conditions and services, and protected characteristics (46). LEAG members were remunerated for their time following National Institute for Health and Care Research (NIHR) guidelines (47).

### 4.2 Ethics approval

Ethics approval was not applicable (48).

### 4.3 Scope and definitions

The LEAG’s focus was on the use of MH data by industry, given the particular sensitivity of this aspect of health data (6). The LEAG noted potential additional MH considerations around consent and vulnerability, and that relevant principles around the use of health data may frequently apply more generally, beyond the MH context. Recognising that individuals may in general consent to any use of their data that they so choose (3,4), if properly informed (49) and possessing capacity to decide (50), the LEAG did not focus on uses of health service data used for research with explicit subject consent, or data collected with consent within ethically approved research studies. Instead, they elected to consider primarily the use, without consent, of routinely collected MH data for research (e.g. research using NHS MH data), but also to consider issues relating to MH data collected directly by industry with consent, such as the nature of that consent process.

We use the terms “industry” and “commercial” to refer to for-profit organisations, typically companies, and any collaborations involving such organisations.

We did not consider truly anonymous data; under UK data protection law, this is not personal data, and by definition it is impossible to re-identify any person from it (3,4).

We did not consider companies involved in processing MH data but only as “data processors”. Under European Union (EU) and UK law (3,4), data processors do not make decisions about the uses of data, but perform tasks as they are instructed by “data controllers”, who are the “owners” and custodians of the data. For example, a company providing computing facilities, e-mail services, or electronic health record (EHR) systems to the NHS is likely to be a data processor handling sensitive confidential (fully identifiable) patient data. While such a company would be responsible for keeping the data secure, it would not be permitted to use the data for its own purposes or research.

While many of the principles, risks, and benefits discussed here would apply to data access by other “third-party” organisations, such as universities, medical charities, and other non-profit organisations such as social enterprises, there is also evidence that such organisations are seen more favourably in this respect by the public (6), and such entities were not the focus of the present work.

We also excluded all considerations of compliance with data protection legislation, applicable information governance rules, or patient/public rights in law, as these are a minimum mandatory standard.

### 4.4 Guideline development

Development was via a qualitative iterative quasi-Delphi approach (51). The DATAMIND academic team provided the LEAG with background materials and an induction programme (March 2022) that included teaching and question/answer sessions on types of health data, examples of the use of health data for research, existing MH data science guidelines, examples of prior work on public opinion on health data sharing, examples of industrial/commercial healthcare research, and examples of real-world controversies in health data use. This contributed to the iterative development of a glossary of relevant terms (see **Supplementary Methods**). The LEAG contributed to decisions around methods from this point. LEAG members were then embedded in the regular DATAMIND leadership and management meetings, with additional patient/public involvement (PPI) in the Strategic Advisory Board overseeing DATAMIND.

In May 2022, guideline development commenced. Academic and industry representatives briefed on the background and their perspectives. LEAG members contributed their views and perspectives to a recorded discussion; the academic team circulated minutes to the LEAG. The LEAG joined the regular DATAMIND Industry Forum (January 2023) to gain and give further perspective on the work of industry and, separately, advised on creation of a data literacy course by DATAMIND and McPin for the public. Some LEAG members attended additional meetings, such as with DARE UK and Research Data Scotland, bringing points and perspectives into development of these guidelines. The LEAG coalesced their views into a 1,950-word short-form note document summarising their considerations for sharing MH data with industry (with anonymity of all specific contributions). The main areas were (1) background knowledge and definitions; (2) the societal benefit of research versus risk of harm and potential mistrust of industry; (3) considerations on consent; (4) particular risks of negative impact on MH patients; (5) processes and other measures to control access to data; (6) security/privacy and identifiability issues; (7) data accuracy; (8) the process of guideline development by the LEAG; (9) routes to output for the LEAG’s work in this area. A knowledge exchange session was delivered by the academic team based on LEAG-identified learning needs (October 2023). The academic team expanded these notes and views into the present paper (being one of the routes to output designed by the LEAG) with draft recommendations based on the LEAG views to date, definitions and explanations of terms as requested by the LEAG, and background information (including regarding health data science and regulations). A LEAG member reviewed existing industry-related health data guidelines, including those in the “grey” literature (e.g. web-based sources) (see **Supplementary Methods**).

The LEAG iteratively edited and revised this paper including the recommendations (from July 2023 to April 2025). Discussions involving the LEAG and industry members were held at the DATAMIND Industry Forum (8 December 2023), focusing in particular on how commercial/industrial research projects/researchers are treated by NHS organisations holding data, and how approvals for UK commercial/industrial MH research are and should be managed outside the public-sector health system. Alongside LEAG development of guidelines for organisations controlling data and for commercial applicants, additional recommendations for NHS executive and regulatory bodies were drafted by academic members of the team with experience in health data research. Separately, the DATAMIND Business Development and Sustainability workstream produced a general guide to the MH data landscape for industry (52). A set of LEAG workshops were held on 8–9 July 2024 as part of finalising the guidelines. The LEAG decided not to rate/score individual guidelines; all were agreed, and they decided not to seek to prioritise some over others. After this process, all authors reviewed and edited the final manuscript, with further revisions discussed and agreed by e-mail, prior to submission.

## 5 RESULTS

### 5.1 Glossary of relevant concepts, background knowledge, and previous guidelines

The glossary of terms that was created is provided in the **Supplementary Results**. Some of the definitions thus created also formed part of the basis for the evolving online DATAMIND Glossary (https://datamind.org.uk/glossary).

The following topics of background information were considered of particular relevance and are presented here:

- What kinds of research is health data (including MH data) used for (**Table 1**)?
- How does data typically flow from health services to research ( **Error: Reference source not found**)?
- What sorts of research do companies do (**Table 2**)?

**Table 1:**
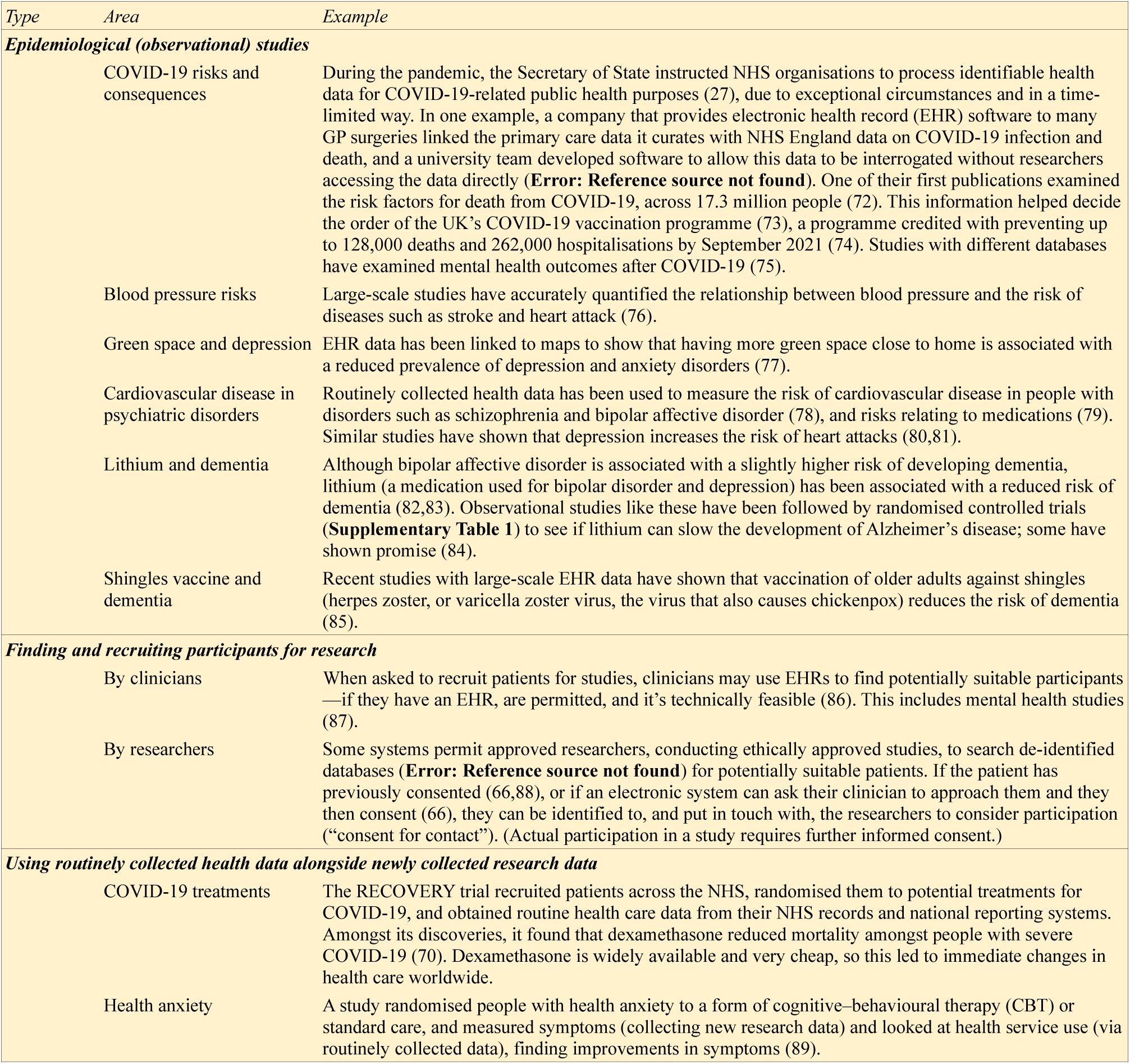
What kinds of research is routinely collected health data (including mental health data) used for? The table shows a few examples, but many studies use these techniques (71). For abbreviations, see **Abbreviations**.

| Type | Area | Example |
| --- | --- | --- |
| **Epidemiological (observational) studies** |  |  |
|  | COVID-19 risks and consequences | During the pandemic, the Secretary of State instructed NHS organisations to process identifiable health data for COVID-19-related public health purposes (27), due to exceptional circumstances and in a time-limited way. In one example, a company that provides electronic health record (EHR) software to many GP surgeries linked the primary care data it curates with NHS England data on COVID-19 infection and death, and a university team developed software to allow this data to be interrogated without researchers accessing the data directly (**Error: Reference source not found**). One of their first publications examined the risk factors for death from COVID-19, across 17.3 million people (72). This information helped decide the order of the UK's COVID-19 vaccination programme (73), a programme credited with preventing up to 128,000 deaths and 262,000 hospitalisations by September 2021 (74). Studies with different databases have examined mental health outcomes after COVID-19 (75). |
|  | Blood pressure risks | Large-scale studies have accurately quantified the relationship between blood pressure and the risk of diseases such as stroke and heart attack (76). |
|  | Green space and depression | EHR data has been linked to maps to show that having more green space close to home is associated with a reduced prevalence of depression and anxiety disorders (77). |
|  | Cardiovascular disease in psychiatric disorders | Routinely collected health data has been used to measure the risk of cardiovascular disease in people with disorders such as schizophrenia and bipolar affective disorder (78), and risks relating to medications (79). Similar studies have shown that depression increases the risk of heart attacks (80,81). |
|  | Lithium and dementia | Although bipolar affective disorder is associated with a slightly higher risk of developing dementia, lithium (a medication used for bipolar disorder and depression) has been associated with a reduced risk of dementia (82,83). Observational studies like these have been followed by randomised controlled trials (**Supplementary Table 1**) to see if lithium can slow the development of Alzheimer's disease; some have shown promise (84). |
|  | Shingles vaccine and dementia | Recent studies with large-scale EHR data have shown that vaccination of older adults against shingles (herpes zoster, or varicella zoster virus, the virus that also causes chickenpox) reduces the risk of dementia (85). |
| **Finding and recruiting participants for research** |  |  |
|  | By clinicians | When asked to recruit patients for studies, clinicians may use EHRs to find potentially suitable participants—if they have an EHR, are permitted, and it's technically feasible (86). This includes mental health studies (87). |
|  | By researchers | Some systems permit approved researchers, conducting ethically approved studies, to search de-identified databases (**Error: Reference source not found**) for potentially suitable patients. If the patient has previously consented (66,88), or if an electronic system can ask their clinician to approach them and they then consent (66), they can be identified to, and put in touch with, the researchers to consider participation ("consent for contact"). (Actual participation in a study requires further informed consent.) |
| **Using routinely collected health data alongside newly collected research data** |  |  |
|  | COVID-19 treatments | The RECOVERY trial recruited patients across the NHS, randomised them to potential treatments for COVID-19, and obtained routine health care data from their NHS records and national reporting systems. Amongst its discoveries, it found that dexamethasone reduced mortality amongst people with severe COVID-19 (70). Dexamethasone is widely available and very cheap, so this led to immediate changes in health care worldwide. |
|  | Health anxiety | A study randomised people with health anxiety to a form of cognitive-behavioural therapy (CBT) or standard care, and measured symptoms (collecting new research data) and looked at health service use (via routinely collected data), finding improvements in symptoms (89). |

**Table 2:** Examples of types of industrial or commercial research, meaning research conducted by or with companies or paid for by companies, relating to health or healthcare. Some examples involve companies acting independently and some involve companies collaborating with, or funding, universities or the NHS. For abbreviations, see **Abbreviations**.

| *Topic* | *Comments* |
| --- | --- |
| **Market research and internal research** | Much research by companies is nothing to do with health care or the NHS. Companies might do simple “research” among their staff (e.g. seeing if they like a new mobile phone app), or market research (e.g. asking the public how much they would pay to use an app), without any requirement for ethical or regulatory approvals. There are industry standards for market research, however, including informed consent (90,91). |
| **“Basic science” research** | Similarly, they might do sophisticated biological or engineering research (e.g. on molecules or cells, in animals, or to build devices), some of which might need regulatory approvals, but which does not involve patients or the NHS. |
| **Direct relationship with consumer** | Companies may obtain data from people directly. For example, a company might invent a mobile phone app that allows users to record their mood. They might ask their users for consent to do research using their data, e.g. to see whether phone usage activity predicts sleep or mood. (By “consumer”, we mean a user of a commercial product, including products provided “for free” by commercial companies.) Sometimes, one company might pay another to collect data. |
| **Pharmaceutical and physical medical device research: clinical trials** | When a pharmaceutical company invents a new medicine (e.g. a drug for cancer) or device (e.g. a heart pacemaker), it normally collaborates with healthcare (e.g. NHS) organisations, to test the invention in consenting volunteers. Typically, patients who wish to participate must consent to the company being able to see some of their health data for this, or the trial couldn’t be done safely (because the company has to help oversee safety, including noticing if an unexpected side effect is happening across multiple sites). |
| **Feasibility questions** | Setting up a medicine trial is a lot of work, so pharmaceutical companies often ask multiple NHS organisations how feasible it would be to work with them, before committing. Part of that involves asking how many patients they see with the relevant condition. NHS organisations would typically provide a very basic answer to that question that cannot identify anyone (e.g. “50 per year”). |
| **Recruitment to clinical trials** | If a commercial clinical trial starts, it needs to recruit participants. Sometimes the criteria are complex, e.g. “people aged 18–45 with depression plus a certain genetic marker who have tried at least three antidepressants and do not have epilepsy”. Therefore, NHS organisations might use EHR data to “shortlist” patients to approach and ask about participation. |
| **Computational health research: artificial intelligence and machine learning** | A technology company developing software to “read” chest X-rays or to predict the risk of mental health crisis might want to train (teach) the software or test (validate) it using NHS data. |
| **Epidemiological research** | A company might want to know if a particular condition or problem is common (“how many people with bipolar affective disorder develop diabetes?”), or look for an unusual side effect (“how many people taking this antidepressant get serotonin syndrome and what are the risk factors?”). They might pay a university or NHS organisation to find out and publish the results. An EHR provider company might support NHS/academic research by enabling research using data that it looks after on behalf of NHS organisations (see COVID-19 example in **Table 1**). |

Reviews of some existing guidelines were provided by a LEAG member, noting both strengths and limitations of content and the approach to guideline development, such as the level of patient/public involvement and of industry involvement (**Supplementary Results**). A common theme of existing guidelines was that MH data, and unstructured or free-text data, was often not considered explicitly (see **Supplementary Results**).

### 5.2 LEAG perspectives noted during guideline development

The LEAG noted some existing and potential benefits of health data research in general, alongside the principle of altruism via data sharing and the benefits and importance of de-identification/anonymisation. These included the potential for healthcare benefits via research, the potential for cost-effective research, and the potential for widening participation in research through the use of routinely collected data. They noted some existing/potential benefits of commercial/industrial health data research, including the potential for treatment developments and financial benefits to the NHS (**Table 3**). They also noted a number of concerns about this area, including concerns about health data research in general (such as data accuracy, concerns around privacy and data security, security concerns, the need for trust and transparency, and the potential for misuse); additional concerns relating to MH data (such as additional sensitivity, the added potential at times for vulnerability when asked for consent, and a particular need for trust and confidentiality in therapeutic relationships); additional concerns relating to free-text MH data; and additional concerns relating to commercial access to data (such as diversity in companies’ motivation and public opinion, previous examples raising public concern, and potential for additional difficulties in overseeing and regulating use) (**Table 4**). This list is not exhaustive (see e.g. **Supplementary Results**).

**Table 3:** Potential strengths and societal benefits noted by the LEAG of commercial/industrial health data research. Some points are not specific to commercial/industrial use and relate also to principles of data use in general. For abbreviations, see **Abbreviations**.

| *Topic* | *Comments* |
| --- | --- |
| **Altruism and helping others** | “If it could help someone then I would share.” |
| **Industry and industrial/NHS collaborations can create healthcare benefits** | The LEAG noted the long history of companies creating health benefits, such as via drug discovery, the development or improvement of medical imaging technology, medical device development. These types of innovation may require healthcare data, sometimes from consenting participants. Companies have also worked to create healthcare benefits through the use of routinely collected data. These include decision support systems in EHRs, such as sepsis alerts (92,93), though these are not foolproof (94,95). De-identified data has been used to train machine learning (ML) systems to interpret X-ray images, such as for automatic cancer detection (96). The LEAG noted potential for healthcare improvement in this domain, noting also evidence that even simple changes to data provision can improve MH care (97). Companies that provide healthcare themselves have used routinely collected data to predict treatment response from psychotherapy (98–101). |
| **Effective de-identification permits greater use** | The LEAG endorsed the principle that effective de-identification or anonymisation of data can allow public benefits to follow, noting examples such as de-identified intensive care data sets available to approved researchers worldwide (102,103). |
| **Full anonymisation permits public distribution of results** | The LEAG noted the legal principle that truly anonymous data is no longer personal and does not need to be restricted (3,4,104), though this label should not be applied without careful thought (**Table 4**). |
| **Widening participation** | The use of routinely collected health data for recruiting to clinical trials and similar research, subject to further consent (**Table 1**), may allow greater and more diverse and representative participation in research. |
| **Cost effectiveness** | Use or re-use of existing data may reduce the costs associated with new data collection. |
| **Scale** | Use of large-scale data may allow new patterns to be identified in data, including via automated methods. |
| **Potential financial benefits to the NHS and thus patients** | “Will industry pay for data?” |

**Table 4:**
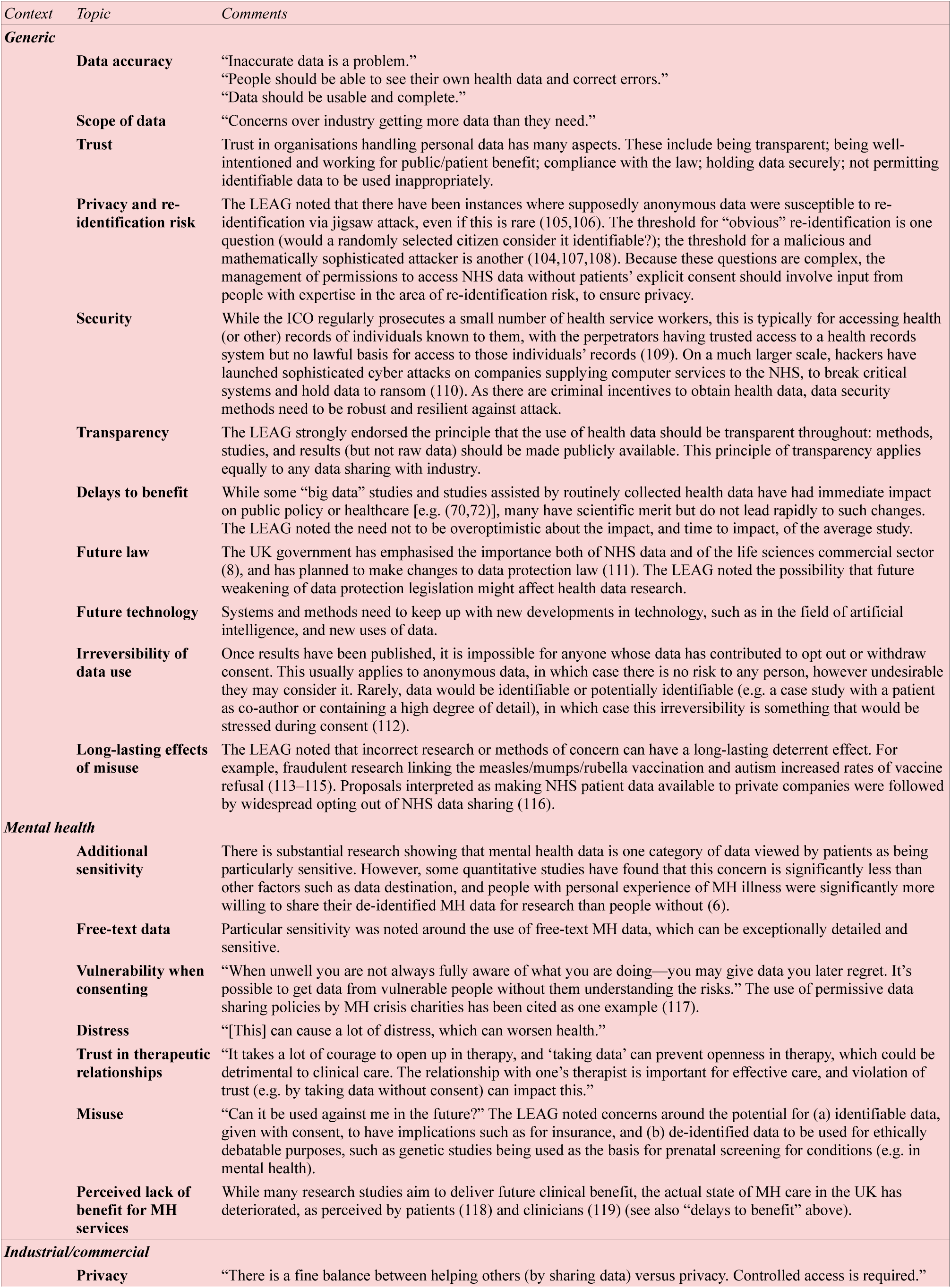
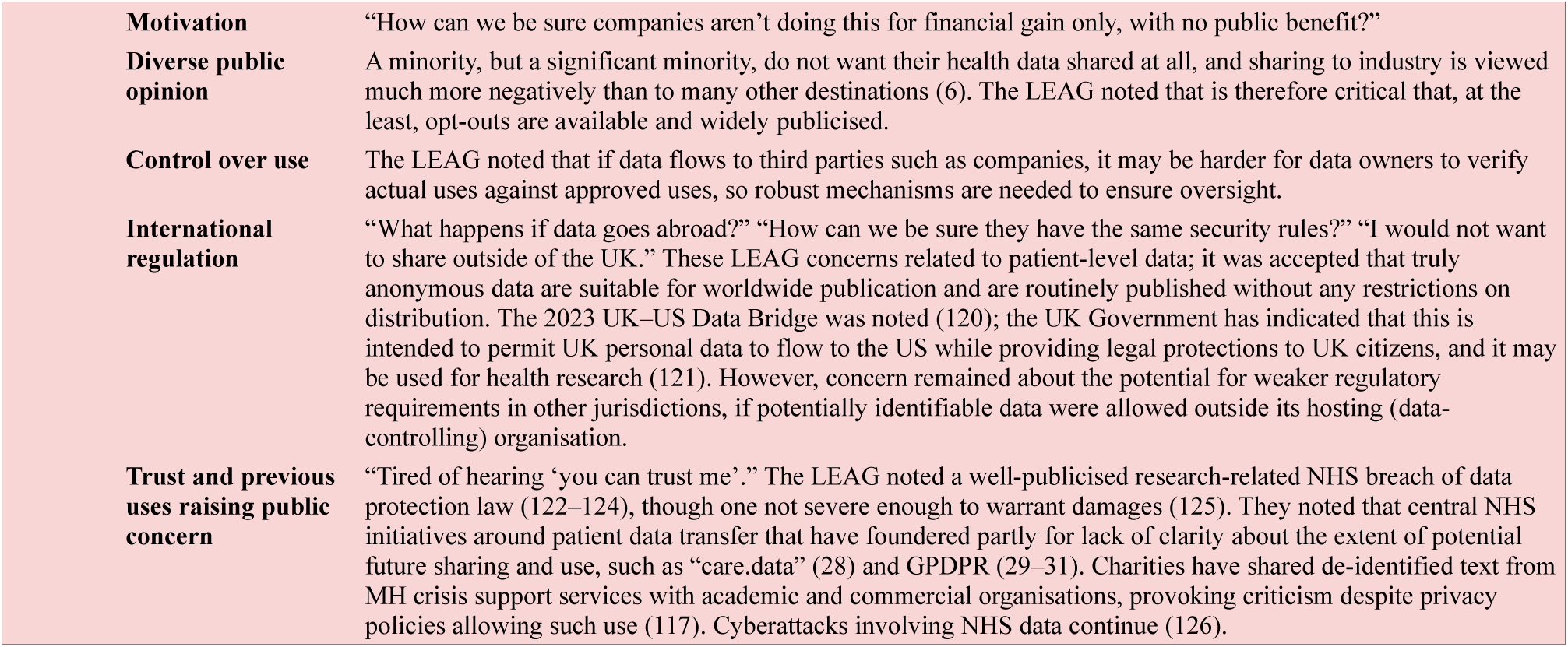
Concerns noted by the LEAG in relation to the use of health data for research: generically, relating to mental health conditions, and relating to industrial/commercial use. For abbreviations, see **Abbreviations**.

Variation was also noted in the financial structures surrounding commercial/industrial health research. Some commercial companies are traditional profit-based entities seeking to develop healthcare, e.g. pharmaceutical or medical device manufacturers; some may be non-profit entities. Benefits to public sector healthcare organisations may also vary, e.g. from free access to software developed from research, through “fee for service” in delivering research, through to participation in commercialisation of new products or technology.

The LEAG considered issues surrounding the process for overseeing and granting access to MH data for research, expressing views in the domains of oversight and accountability; ways to maximise transparency; the need to consider risks and benefits of proposed research (noting that public benefit is necessary but not sufficient, benefits and risks must be considered together, and while most health data research will not be completely risk-free, the approaches taken must seek to minimise risks, allowing only appropriate and necessary risks to be taken in a carefully safeguarded way); and on issues surrounding consent and opt-outs when applicable (**Table 5**). The LEAG noted that existing processes already include significant controls and these normally include PPI, such as when research is conducted under NHS research ethics approvals (53), when UK Confidentiality Advisory Group (CAG) approvals are sought for the use of identifiable patient information for research without consent (54), and within “local” TRE approval processes [e.g. (55)].

**Table 5:**
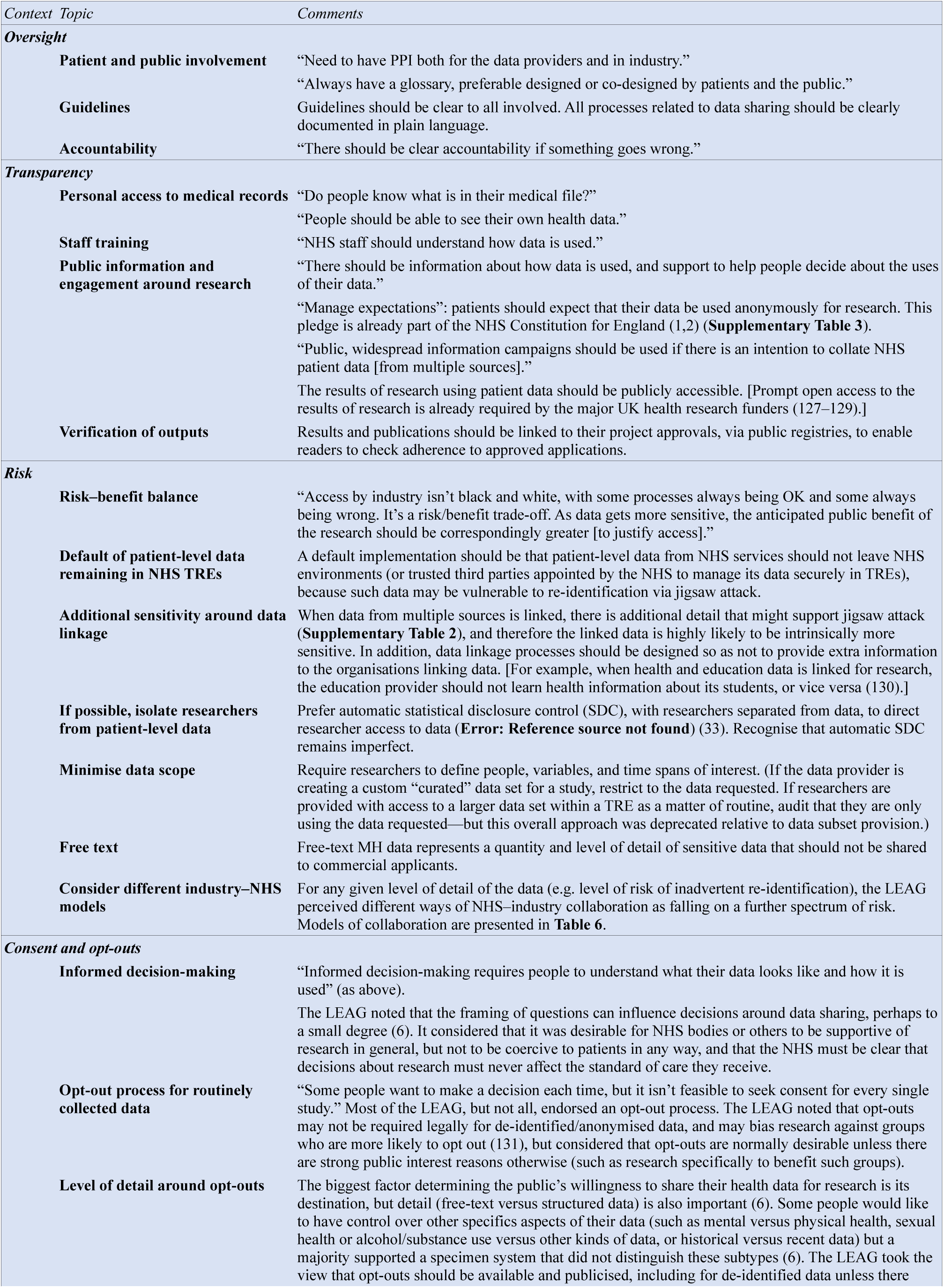
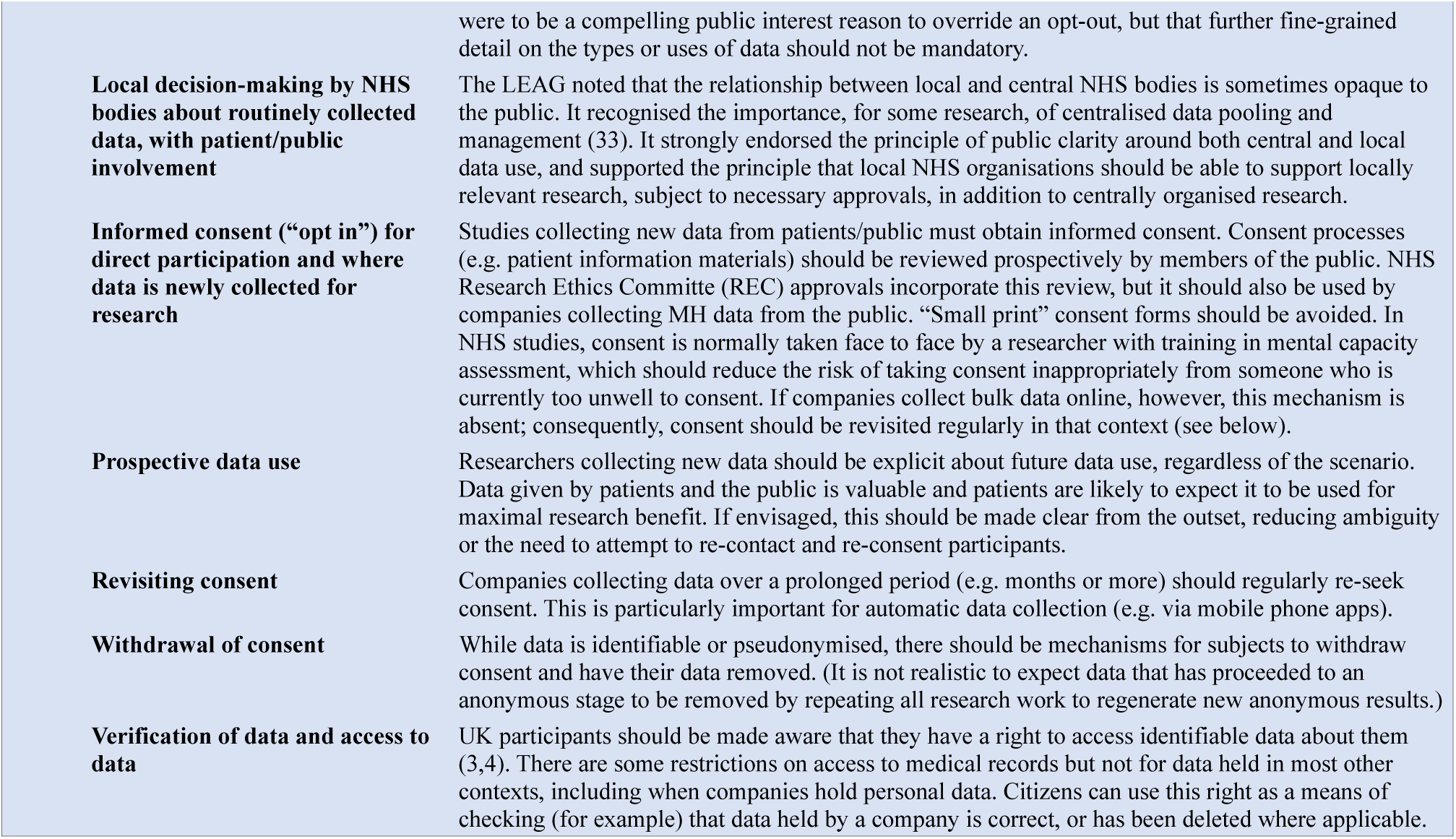
LEAG opinions and perspectives on the MH data access request process for research. For abbreviations, see **Abbreviations**.

Outside the domain of research involving collaboration between industry and health services, the LEAG noted that “health apps”, such as software applications available from public app stores, represent a relatively unregulated area—compared, for example, to software in use within healthcare services for decision support. The domain of “health apps”, and of other online therapy and support services, is an area where data may be collected subject to “small print” consent forms, with relatively little clarity about final uses of the data; the LEAG deprecated such processes in favour of more transparency around data collection and use (**Table 5**).

The LEAG noted that one core principle is that that the anticipated patient/public benefit of research is fundamental to assessing it against any risks posed, such as to privacy. Another core principle is of trustworthiness and transparency, noting that coherent rules and guidelines assist this. The LEAG took the view that, where public benefits are strong, research is conducted transparently, privacy is strongly safeguarded, and the public are appropriately consulted, that potential industrial/commercial users of MH data should be treated by data providers in broadly the same way as health service/academic users, with the exceptions that (a) free-text MH data should not be provided to industrial/commercial applicants, and (b) unconsented patient-level data should not be exported outside NHS-controlled secure data environments.

### 5.3 Guidelines

Recommendations were then produced, revised, and agreed (see **Methods**). The resulting specific recommendations and best-practice guidelines were divided into those for host (data controller) organisations and potential commercial users of MH data.

The guidelines for host organisations (data controllers, and potential data access providers, e.g. NHS bodies) are shown in **Table 6**. In summary, they emphasise general principles of transparency, patient rights and the need to support these via trained staff, PPI, the use of formal governance processes, and the need for awareness of regulatory and legal requirements. Individual applications for data access should be specified in detail and have their risks and potential benefits assessed by an oversight committee including PPI. The guidelines advise that the following be entirely avoided for commercial applicants: (a) high-risk models of data sharing, such as exporting unconsented data outside NHS-controlled environments; (b) the provision of unconsented free-text MH data, even if de-identified. Standard principles must apply as usual, including reseacher training and oversight of researchers, data minimisation, and statistical disclosure control before publishable results leave secure environments. Full details and additional points are in **Table 6**.

**Table 6:**
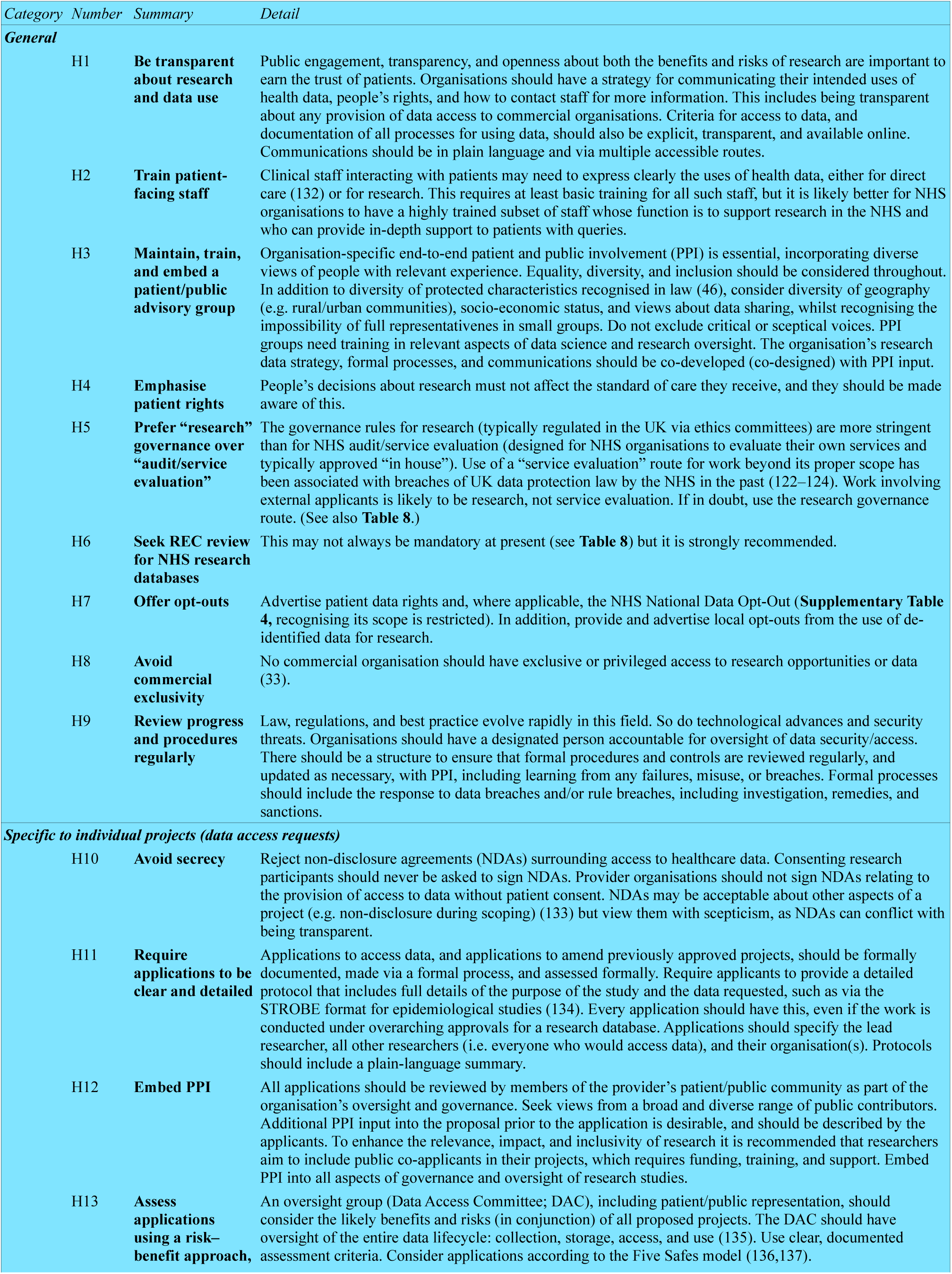
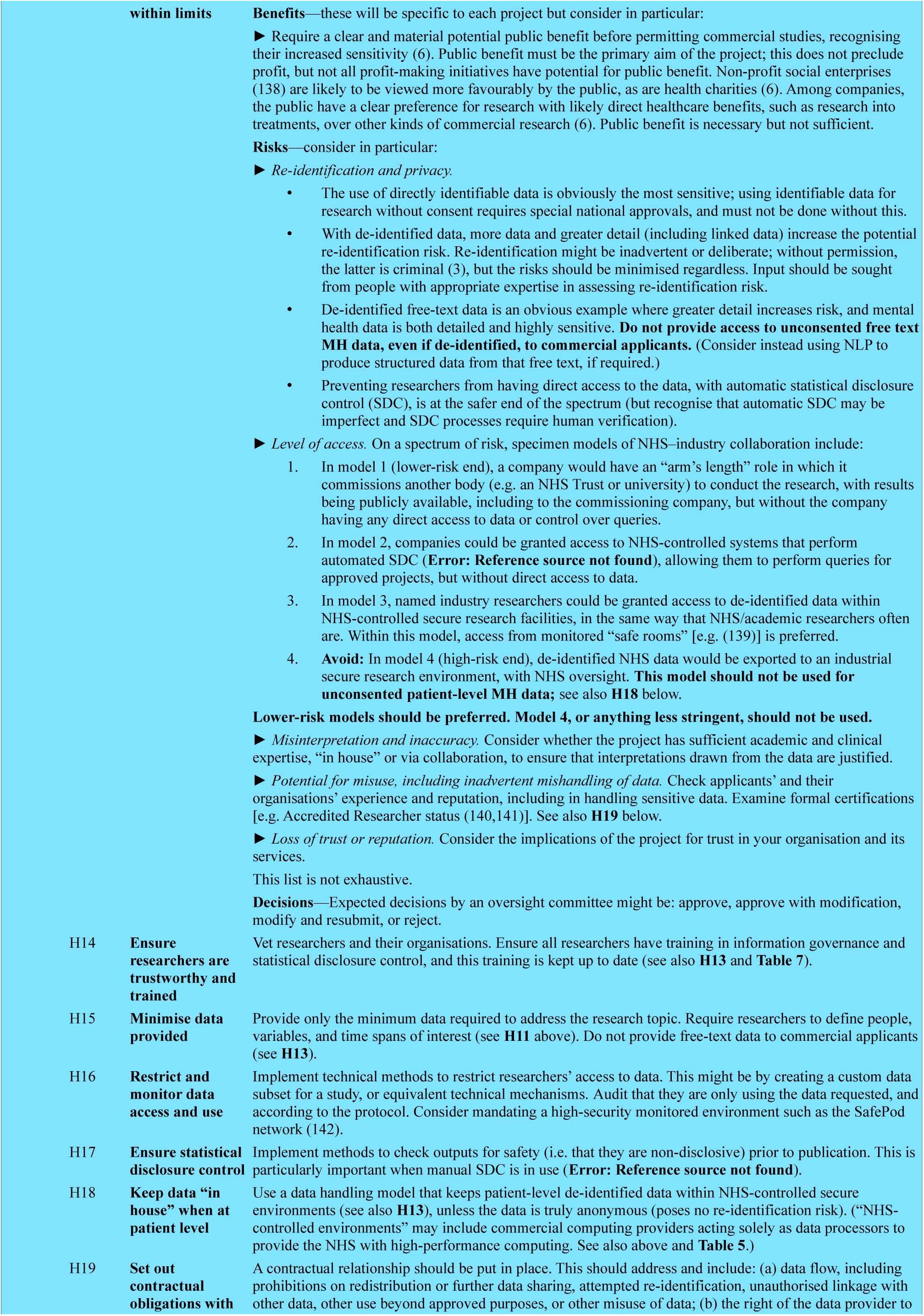
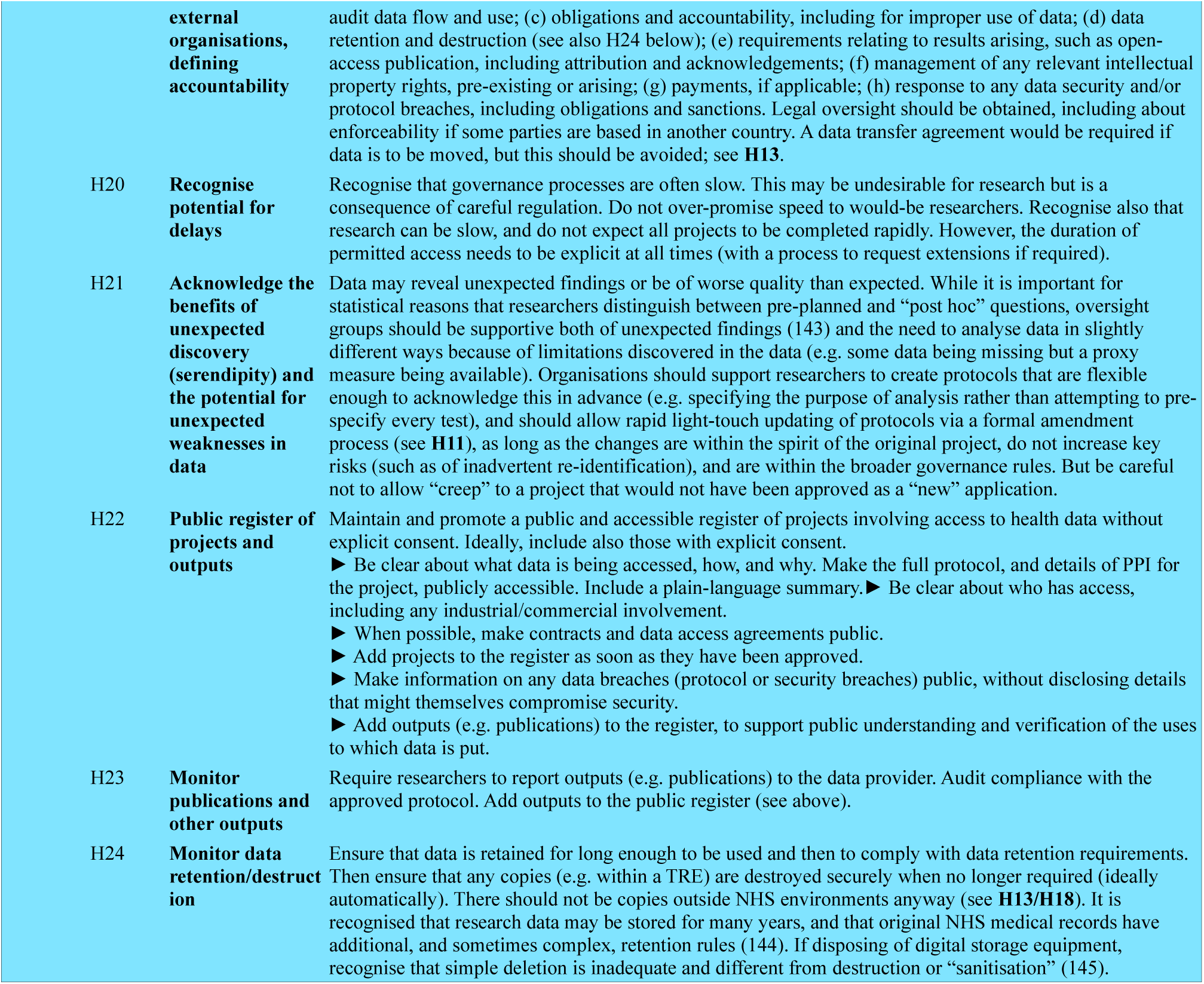
Guidelines for organisations hosting (H) mental health data as data controllers, i.e. potential data access providers (such as NHS organisations), approached by industrial/commercial applicants relating to MH data requests for research. For scope, see **Methods.** For abbreviations, see **Abbreviations**.

| Category | Number | Summary | Detail |
| --- | --- | --- | --- |
| **General** |  |  |  |
|  | H1 | **Be transparent about research and data use** | Public engagement, transparency, and openness about both the benefits and risks of research are important to earn the trust of patients. Organisations should have a strategy for communicating their intended uses of health data, people's rights, and how to contact staff for more information. This includes being transparent about any provision of data access to commercial organisations. Criteria for access to data, and documentation of all processes for using data, should also be explicit, transparent, and available online. Communications should be in plain language and via multiple accessible routes. |
|  | H2 | **Train patient-facing staff** | Clinical staff interacting with patients may need to express clearly the uses of health data, either for direct care (132) or for research. This requires at least basic training for all such staff, but it is likely better for NHS organisations to have a highly trained subset of staff whose function is to support research in the NHS and who can provide in-depth support to patients with queries. |
|  | H3 | **Maintain, train, and embed a patient/public advisory group** | Organisation-specific end-to-end patient and public involvement (PPI) is essential, incorporating diverse views of people with relevant experience. Equality, diversity, and inclusion should be considered throughout. In addition to diversity of protected characteristics recognised in law (46), consider diversity of geography (e.g. rural/urban communities), socio-economic status, and views about data sharing, whilst recognising the impossibility of full representativeness in small groups. Do not exclude critical or sceptical voices. PPI groups need training in relevant aspects of data science and research oversight. The organisation's research data strategy, formal processes, and communications should be co-developed (co-designed) with PPI input. |
|  | H4 | **Emphasise patient rights** | People's decisions about research must not affect the standard of care they receive, and they should be made aware of this. |
|  | H5 | **Prefer "research" governance over "audit/service evaluation"** | The governance rules for research (typically regulated in the UK via ethics committees) are more stringent than for NHS audit/service evaluation (designed for NHS organisations to evaluate their own services and typically approved "in house"). Use of a "service evaluation" route for work beyond its proper scope has been associated with breaches of UK data protection law by the NHS in the past (122–124). Work involving external applicants is likely to be research, not service evaluation. If in doubt, use the research governance route. (See also **Table 8**.) |
|  | H6 | **Seek REC review for NHS research databases** | This may not always be mandatory at present (see **Table 8**) but it is strongly recommended. |
|  | H7 | **Offer opt-outs** | Advertise patient data rights and, where applicable, the NHS National Data Opt-Out (**Supplementary Table 4**, recognising its scope is restricted). In addition, provide and advertise local opt-outs from the use of de-identified data for research. |
|  | H8 | **Avoid commercial exclusivity** | No commercial organisation should have exclusive or privileged access to research opportunities or data (33). |
|  | H9 | **Review progress and procedures regularly** | Law, regulations, and best practice evolve rapidly in this field. So do technological advances and security threats. Organisations should have a designated person accountable for oversight of data security/access. There should be a structure to ensure that formal procedures and controls are reviewed regularly, and updated as necessary, with PPI, including learning from any failures, misuse, or breaches. Formal processes should include the response to data breaches and/or rule breaches, including investigation, remedies, and sanctions. |
| **Specific to individual projects (data access requests)** |  |  |  |
|  | H10 | **Avoid secrecy** | Reject non-disclosure agreements (NDAs) surrounding access to healthcare data. Consenting research participants should never be asked to sign NDAs. Provider organisations should not sign NDAs relating to the provision of access to data without patient consent. NDAs may be acceptable about other aspects of a project (e.g. non-disclosure during scoping) (133) but view them with scepticism, as NDAs can conflict with being transparent. |
|  | H11 | **Require applications to be clear and detailed** | Applications to access data, and applications to amend previously approved projects, should be formally documented, made via a formal process, and assessed formally. Require applicants to provide a detailed protocol that includes full details of the purpose of the study and the data requested, such as via the STROBE format for epidemiological studies (134). Every application should have this, even if the work is conducted under overarching approvals for a research database. Applications should specify the lead researcher, all other researchers (i.e. everyone who would access data), and their organisation(s). Protocols should include a plain-language summary. |
|  | H12 | **Embed PPI** | All applications should be reviewed by members of the provider's patient/public community as part of the organisation's oversight and governance. Seek views from a broad and diverse range of public contributors. Additional PPI input into the proposal prior to the application is desirable, and should be described by the applicants. To enhance the relevance, impact, and inclusivity of research it is recommended that researchers aim to include public co-applicants in their projects, which requires funding, training, and support. Embed PPI into all aspects of governance and oversight of research studies. |
|  | H13 | **Assess applications using a risk–benefit approach,** | An oversight group (Data Access Committee; DAC), including patient/public representation, should consider the likely benefits and risks (in conjunction) of all proposed projects. The DAC should have oversight of the entire data lifecycle: collection, storage, access, and use (135). Use clear, documented assessment criteria. Consider applications according to the Five Safes model (136,137). |

|  |  |  |
| --- | --- | --- |
|  | within limits | **Benefits**—these will be specific to each project but consider in particular:<br>► Require a clear and material potential public benefit before permitting commercial studies, recognising their increased sensitivity (6). Public benefit must be the primary aim of the project; this does not preclude profit, but not all profit-making initiatives have potential for public benefit. Non-profit social enterprises (138) are likely to be viewed more favourably by the public, as are health charities (6). Among companies, the public have a clear preference for research with likely direct healthcare benefits, such as research into treatments, over other kinds of commercial research (6). Public benefit is necessary but not sufficient.<br>**Risks**—consider in particular:<br>► *Re-identification and privacy.*<br>• The use of directly identifiable data is obviously the most sensitive; using identifiable data for research without consent requires special national approvals, and must not be done without this.<br>• With de-identified data, more data and greater detail (including linked data) increase the potential re-identification risk. Re-identification might be inadvertent or deliberate; without permission, the latter is criminal (3), but the risks should be minimised regardless. Input should be sought from people with appropriate expertise in assessing re-identification risk.<br>• De-identified free-text data is an obvious example where greater detail increases risk, and mental health data is both detailed and highly sensitive. **Do not provide access to unconsented free text MH data, even if de-identified, to commercial applicants.** (Consider instead using NLP to produce structured data from that free text, if required.)<br>• Preventing researchers from having direct access to the data, with automatic statistical disclosure control (SDC), is at the safer end of the spectrum (but recognise that automatic SDC may be imperfect and SDC processes require human verification).<br>► *Level of access.* On a spectrum of risk, specimen models of NHS–industry collaboration include:<br>1. In model 1 (lower-risk end), a company would have an “arm’s length” role in which it commissions another body (e.g. an NHS Trust or university) to conduct the research, with results being publicly available, including to the commissioning company, but without the company having any direct access to data or control over queries.<br>2. In model 2, companies could be granted access to NHS-controlled systems that perform automated SDC (**Error: Reference source not found**), allowing them to perform queries for approved projects, but without direct access to data.<br>3. In model 3, named industry researchers could be granted access to de-identified data within NHS-controlled secure research facilities, in the same way that NHS/academic researchers often are. Within this model, access from monitored “safe rooms” [e.g. (139)] is preferred.<br>4. **Avoid:** In model 4 (high-risk end), de-identified NHS data would be exported to an industrial secure research environment, with NHS oversight. **This model should not be used for unconsented patient-level MH data;** see also **H18** below.<br>**Lower-risk models should be preferred. Model 4, or anything less stringent, should not be used.**<br>► *Misinterpretation and inaccuracy.* Consider whether the project has sufficient academic and clinical expertise, “in house” or via collaboration, to ensure that interpretations drawn from the data are justified.<br>► *Potential for misuse, including inadvertent mishandling of data.* Check applicants’ and their organisations’ experience and reputation, including in handling sensitive data. Examine formal certifications [e.g. Accredited Researcher status (140,141)]. See also **H19** below.<br>► *Loss of trust or reputation.* Consider the implications of the project for trust in your organisation and its services.<br>This list is not exhaustive.<br>**Decisions**—Expected decisions by an oversight committee might be: approve, approve with modification, modify and resubmit, or reject. |
| H14 | Ensure researchers are trustworthy and trained | Vet researchers and their organisations. Ensure all researchers have training in information governance and statistical disclosure control, and this training is kept up to date (see also **H13** and **Table 7**). |
| H15 | Minimise data provided | Provide only the minimum data required to address the research topic. Require researchers to define people, variables, and time spans of interest (see **H11** above). Do not provide free-text data to commercial applicants (see **H13**). |
| H16 | Restrict and monitor data access and use | Implement technical methods to restrict researchers’ access to data. This might be by creating a custom data subset for a study, or equivalent technical mechanisms. Audit that they are only using the data requested, and according to the protocol. Consider mandating a high-security monitored environment such as the SafePod network (142). |
| H17 | Ensure statistical disclosure control | Implement methods to check outputs for safety (i.e. that they are non-disclosive) prior to publication. This is particularly important when manual SDC is in use (**Error: Reference source not found**). |
| H18 | Keep data “in house” when at patient level | Use a data handling model that keeps patient-level de-identified data within NHS-controlled secure environments (see also **H13**), unless the data is truly anonymous (poses no re-identification risk). (“NHS-controlled environments” may include commercial computing providers acting solely as data processors to provide the NHS with high-performance computing. See also above and **Table 5**.) |
| H19 | Set out contractual obligations with | A contractual relationship should be put in place. This should address and include: (a) data flow, including prohibitions on redistribution or further data sharing, attempted re-identification, unauthorised linkage with other data, other use beyond approved purposes, or other misuse of data; (b) the right of the data provider to |
|  | **external organisations, defining accountability** | audit data flow and use; (c) obligations and accountability, including for improper use of data; (d) data retention and destruction (see also H24 below); (e) requirements relating to results arising, such as open-access publication, including attribution and acknowledgements; (f) management of any relevant intellectual property rights, pre-existing or arising; (g) payments, if applicable; (h) response to any data security and/or protocol breaches, including obligations and sanctions. Legal oversight should be obtained, including about enforceability if some parties are based in another country. A data transfer agreement would be required if data is to be moved, but this should be avoided; see **H13**. |
| H20 | **Recognise potential for delays** | Recognise that governance processes are often slow. This may be undesirable for research but is a consequence of careful regulation. Do not over-promise speed to would-be researchers. Recognise also that research can be slow, and do not expect all projects to be completed rapidly. However, the duration of permitted access needs to be explicit at all times (with a process to request extensions if required). |
| H21 | **Acknowledge the benefits of unexpected discovery (serendipity) and the potential for unexpected weaknesses in data** | Data may reveal unexpected findings or be of worse quality than expected. While it is important for statistical reasons that researchers distinguish between pre-planned and “post hoc” questions, oversight groups should be supportive both of unexpected findings (143) and the need to analyse data in slightly different ways because of limitations discovered in the data (e.g. some data being missing but a proxy measure being available). Organisations should support researchers to create protocols that are flexible enough to acknowledge this in advance (e.g. specifying the purpose of analysis rather than attempting to pre-specify every test), and should allow rapid light-touch updating of protocols via a formal amendment process (see **H11**), as long as the changes are within the spirit of the original project, do not increase key risks (such as of inadvertent re-identification), and are within the broader governance rules. But be careful not to allow “creep” to a project that would not have been approved as a “new” application. |
| H22 | **Public register of projects and outputs** | Maintain and promote a public and accessible register of projects involving access to health data without explicit consent. Ideally, include also those with explicit consent.<br>▶ Be clear about what data is being accessed, how, and why. Make the full protocol, and details of PPI for the project, publicly accessible. Include a plain-language summary.<br>▶ Be clear about who has access, including any industrial/commercial involvement.<br>▶ When possible, make contracts and data access agreements public.<br>▶ Add projects to the register as soon as they have been approved.<br>▶ Make information on any data breaches (protocol or security breaches) public, without disclosing details that might themselves compromise security.<br>▶ Add outputs (e.g. publications) to the register, to support public understanding and verification of the uses to which data is put. |
| H23 | **Monitor publications and other outputs** | Require researchers to report outputs (e.g. publications) to the data provider. Audit compliance with the approved protocol. Add outputs to the public register (see above). |
| H24 | **Monitor data retention/destruction** | Ensure that data is retained for long enough to be used and then to comply with data retention requirements. Then ensure that any copies (e.g. within a TRE) are destroyed securely when no longer required (ideally automatically). There should not be copies outside NHS environments anyway (see **H13/H18**). It is recognised that research data may be stored for many years, and that original NHS medical records have additional, and sometimes complex, retention rules (144). If disposing of digital storage equipment, recognise that simple deletion is inadequate and different from destruction or “sanitisation” (145). |

The guidelines for commercial applicants (potential data users) mirror those for host organisations, and are shown in **Table 7**. In summary, they emphasise the need for thorough awareness of information governance principles and applicable regulations (including an understanding of data collected with versus without consent for specific research purposes), involvement of patients and the public in commercial research, and training in statistical disclosure control. Fully specified protocols should be prepared for a project application to a hosting (data-controlling) organisation, and all requirements from that host organisation complied with, with transparency to the public throughout. Full details are in **Table 7**.

**Table 7:**
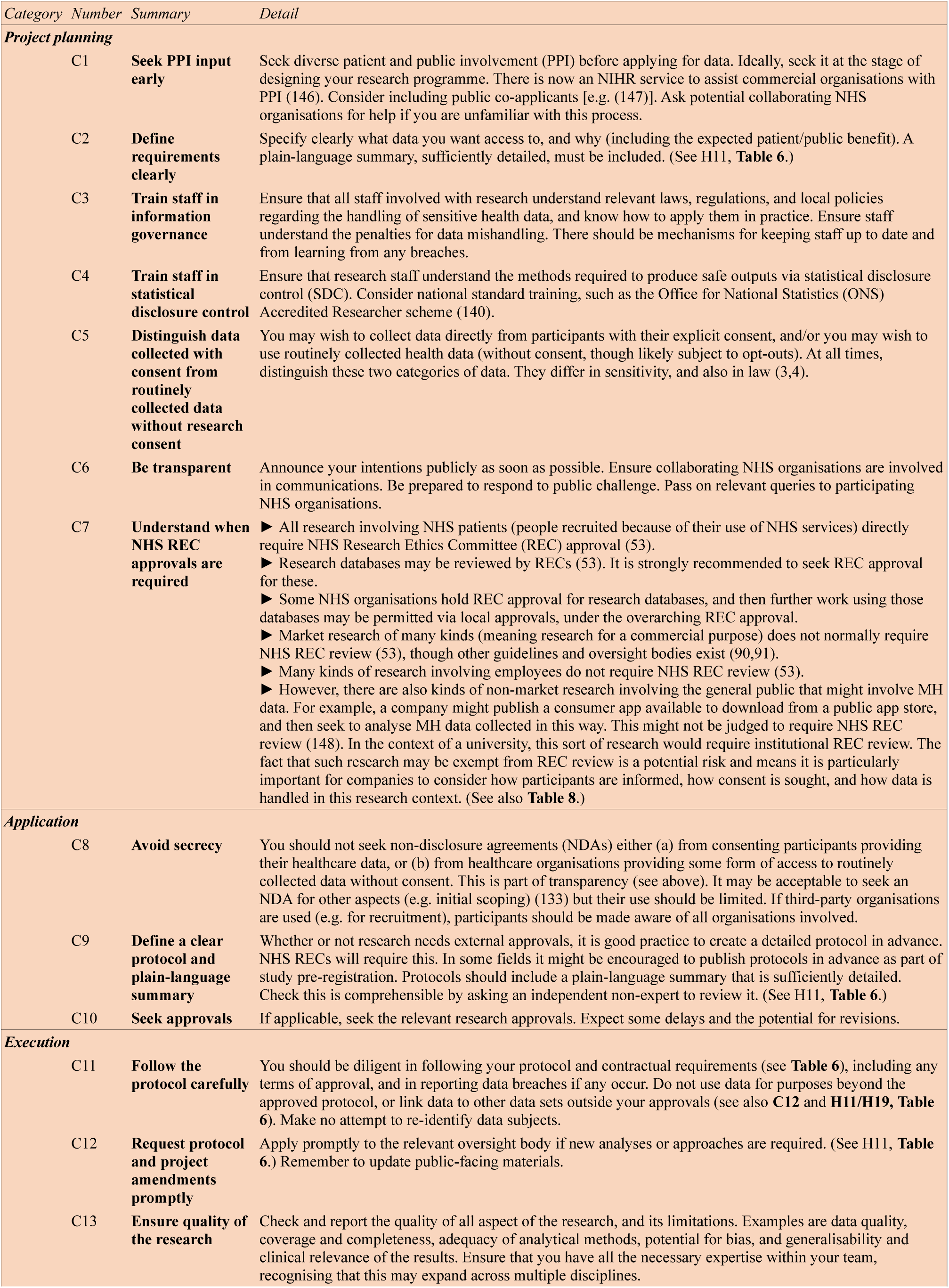
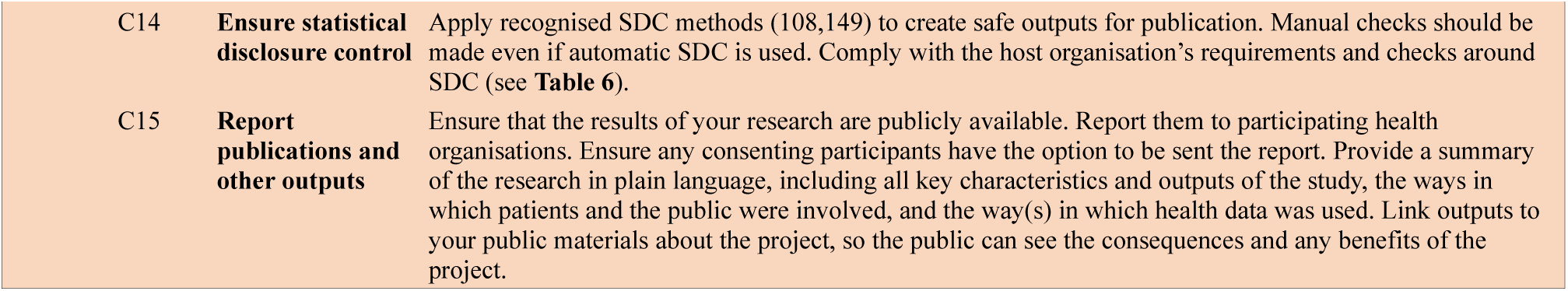
Guidelines for commercial (C) applicants seeking to use mental health data for research (potential data users). For abbreviations, see **Abbreviations**.

| Category | Number | Summary | Detail |
| --- | --- | --- | --- |
| **Project planning** |  |  |  |
|  | C1 | **Seek PPI input early** | Seek diverse patient and public involvement (PPI) before applying for data. Ideally, seek it at the stage of designing your research programme. There is now an NIHR service to assist commercial organisations with PPI (146). Consider including public co-applicants [e.g. (147)]. Ask potential collaborating NHS organisations for help if you are unfamiliar with this process. |
|  | C2 | **Define requirements clearly** | Specify clearly what data you want access to, and why (including the expected patient/public benefit). A plain-language summary, sufficiently detailed, must be included. (See H11, **Table 6**.) |
|  | C3 | **Train staff in information governance** | Ensure that all staff involved with research understand relevant laws, regulations, and local policies regarding the handling of sensitive health data, and know how to apply them in practice. Ensure staff understand the penalties for data mishandling. There should be mechanisms for keeping staff up to date and from learning from any breaches. |
|  | C4 | **Train staff in statistical disclosure control** | Ensure that research staff understand the methods required to produce safe outputs via statistical disclosure control (SDC). Consider national standard training, such as the Office for National Statistics (ONS) Accredited Researcher scheme (140). |
|  | C5 | **Distinguish data collected with consent from routinely collected data without research consent** | You may wish to collect data directly from participants with their explicit consent, and/or you may wish to use routinely collected health data (without consent, though likely subject to opt-outs). At all times, distinguish these two categories of data. They differ in sensitivity, and also in law (3,4). |
|  | C6 | **Be transparent** | Announce your intentions publicly as soon as possible. Ensure collaborating NHS organisations are involved in communications. Be prepared to respond to public challenge. Pass on relevant queries to participating NHS organisations. |
|  | C7 | **Understand when NHS REC approvals are required** | ▶ All research involving NHS patients (people recruited because of their use of NHS services) directly require NHS Research Ethics Committee (REC) approval (53).<br>▶ Research databases may be reviewed by RECs (53). It is strongly recommended to seek REC approval for these.<br>▶ Some NHS organisations hold REC approval for research databases, and then further work using those databases may be permitted via local approvals, under the overarching REC approval.<br>▶ Market research of many kinds (meaning research for a commercial purpose) does not normally require NHS REC review (53), though other guidelines and oversight bodies exist (90,91).<br>▶ Many kinds of research involving employees do not require NHS REC review (53).<br>▶ However, there are also kinds of non-market research involving the general public that might involve MH data. For example, a company might publish a consumer app available to download from a public app store, and then seek to analyse MH data collected in this way. This might not be judged to require NHS REC review (148). In the context of a university, this sort of research would require institutional REC review. The fact that such research may be exempt from REC review is a potential risk and means it is particularly important for companies to consider how participants are informed, how consent is sought, and how data is handled in this research context. (See also **Table 8**.) |
| **Application** |  |  |  |
|  | C8 | **Avoid secrecy** | You should not seek non-disclosure agreements (NDAs) either (a) from consenting participants providing their healthcare data, or (b) from healthcare organisations providing some form of access to routinely collected data without consent. This is part of transparency (see above). It may be acceptable to seek an NDA for other aspects (e.g. initial scoping) (133) but their use should be limited. If third-party organisations are used (e.g. for recruitment), participants should be made aware of all organisations involved. |
|  | C9 | **Define a clear protocol and plain-language summary** | Whether or not research needs external approvals, it is good practice to create a detailed protocol in advance. NHS RECs will require this. In some fields it might be encouraged to publish protocols in advance as part of study pre-registration. Protocols should include a plain-language summary that is sufficiently detailed. Check this is comprehensible by asking an independent non-expert to review it. (See H11, **Table 6**.) |
|  | C10 | **Seek approvals** | If applicable, seek the relevant research approvals. Expect some delays and the potential for revisions. |
| **Execution** |  |  |  |
|  | C11 | **Follow the protocol carefully** | You should be diligent in following your protocol and contractual requirements (see **Table 6**), including any terms of approval, and in reporting data breaches if any occur. Do not use data for purposes beyond the approved protocol, or link data to other data sets outside your approvals (see also **C12** and **H11/H19, Table 6**). Make no attempt to re-identify data subjects. |
|  | C12 | **Request protocol and project amendments promptly** | Apply promptly to the relevant oversight body if new analyses or approaches are required. (See H11, **Table 6**.) Remember to update public-facing materials. |
|  | C13 | **Ensure quality of the research** | Check and report the quality of all aspect of the research, and its limitations. Examples are data quality, coverage and completeness, adequacy of analytical methods, potential for bias, and generalisability and clinical relevance of the results. Ensure that you have all the necessary expertise within your team, recognising that this may expand across multiple disciplines. |

|  |  |  |
| --- | --- | --- |
| C14 | **Ensure statistical disclosure control** | Apply recognised SDC methods (108,149) to create safe outputs for publication. Manual checks should be made even if automatic SDC is used. Comply with the host organisation's requirements and checks around SDC (see **Table 6**). |
| C15 | **Report publications and other outputs** | Ensure that the results of your research are publicly available. Report them to participating health organisations. Ensure any consenting participants have the option to be sent the report. Provide a summary of the research in plain language, including all key characteristics and outputs of the study, the ways in which patients and the public were involved, and the way(s) in which health data was used. Link outputs to your public materials about the project, so the public can see the consequences and any benefits of the project. |

We also make recommendations for NHS executive and regulatory bodies, shown in **Table 8**. In summary, these cover the provision of central systems to simplify choice and consent from the public; clearer and consistent advice to data-controlling (e.g. NHS) organisations around aspects of research governance; support for efforts to de-identify data in situations where it is sometimes not de-identified at present; and continuing to improve transparency through citizens viewing their own health data, as well as through public visibility of research conducted with routinely collected data. For full details, see **Table 8**.

**Table 8:**
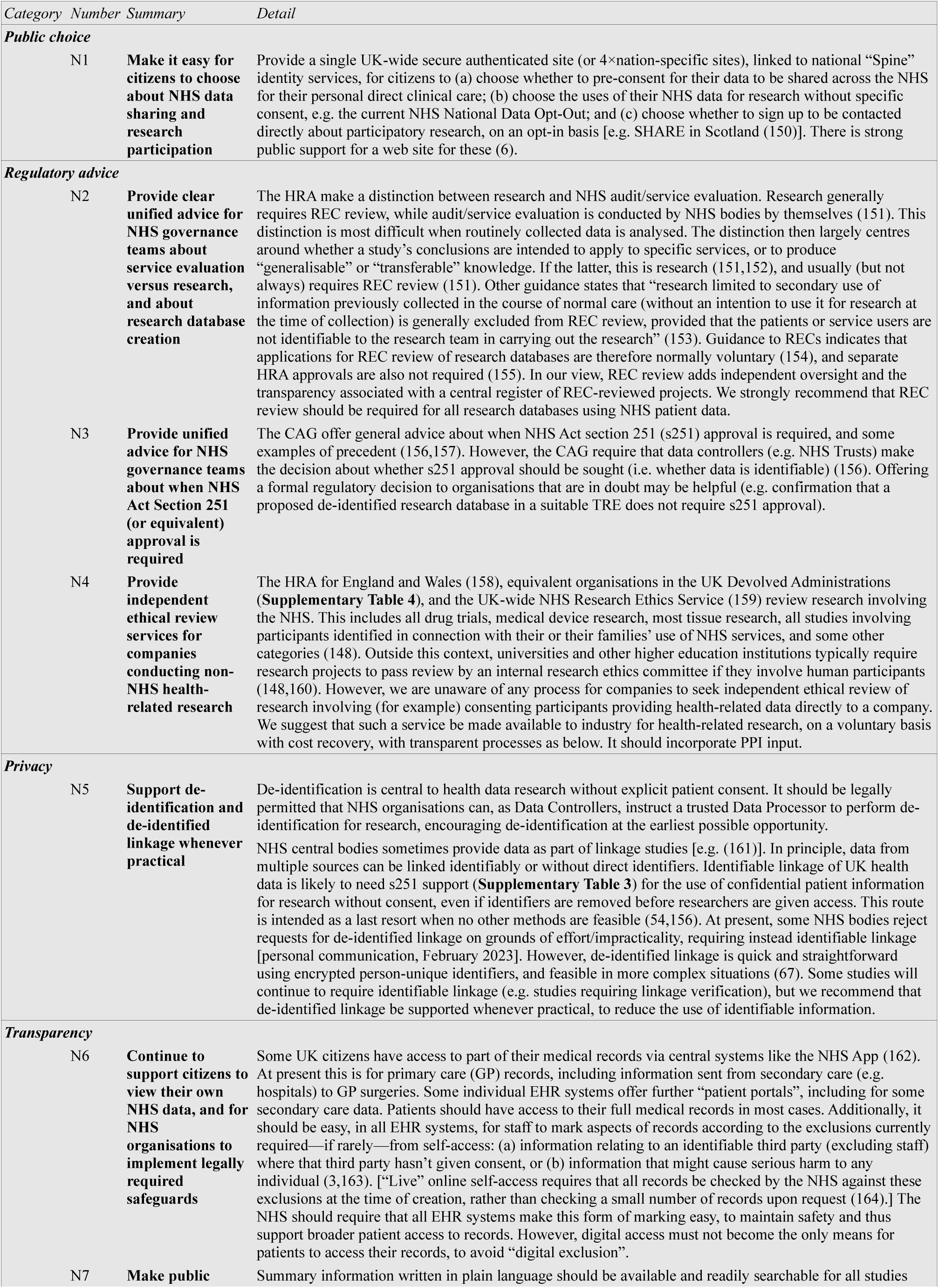
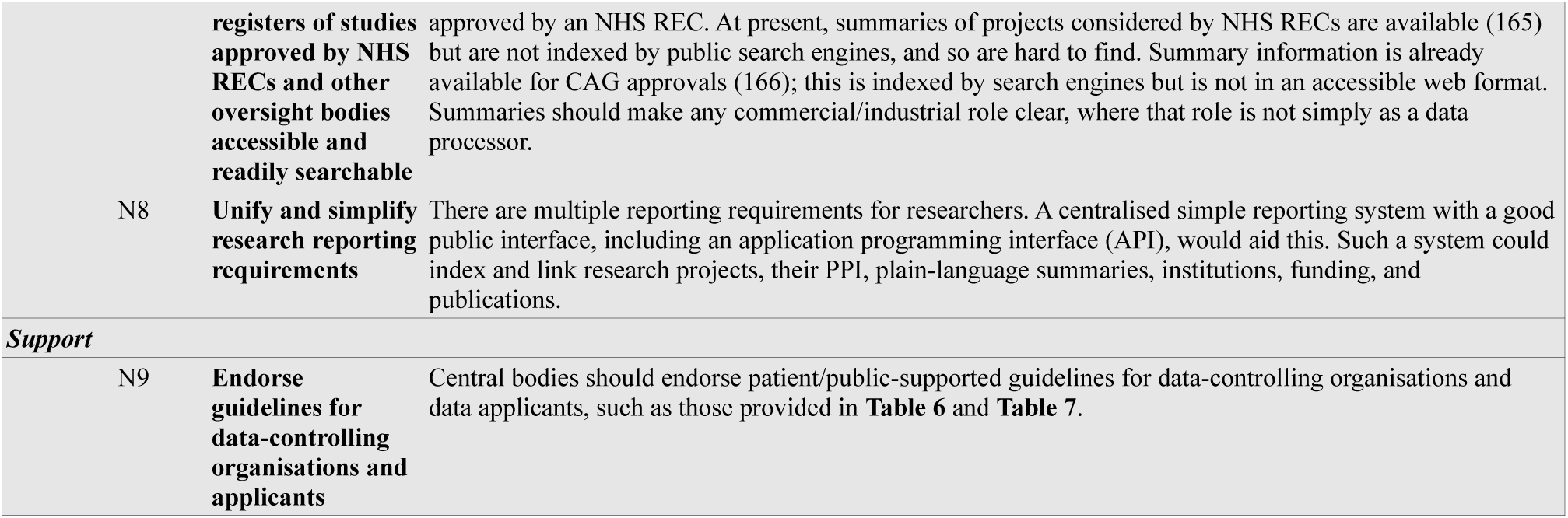
Recommendations for NHS (N) executive/regulatory bodies. Most do not relate to commercial/industrial research specifically but are more generic. For abbreviations, see **Abbreviations**.

## 6 DISCUSSION

To our knowledge, there is no previous set of best-practice recommendations to guide UK public healthcare providers approached by companies about potential access specifically to MH data for research, nor for companies seeking to make use of such data. In the present study, a research advisory group of patient/public members developed such guidance iteratively (**Table 6**, **Table 7**), and we make additional suggestions for executive/regulatory bodies (**Table 8**). We hope these are useful in practice, for these bodies and also for patients and the public, to increase awareness and expectations of good practice.

The guidance includes much that is uncontroversial: it is well established that there should be an emphasis on transparency, data safety and security, statistical disclosure control, and robust oversight (45). The guidance for host (data controller) organisations adds specificity, with organisation-wide guidance such as on staff training; ensuring a strong oversight regime is preferred (e.g. “research” over “service evaluation” approvals when in doubt); providing opt-outs; and avoiding exclusivity with industrial/commercial applicants (**Table 6**). For the consideration of individual proposed projects, the LEAG excluded some specific models of data sharing, namely the transmission of patient-level unconsented data outside NHS-controlled TREs and the provision of free-text data to commercial applicants, even when de-identified (**Table 6**). They set other models on a continuum of risk, advising strongly that lower-risk models should be preferred, and that higher-risk data access approaches can only be justified if there is a compelling public benefit argument and strong patient/public support for a specific project, which was envisaged to be exceptional. Linkage projects were noted to bring additional risks (**Table 6**).

An international study of young people’s preferences around mental health data use has also supported the “secure server” or TRE/SDE framework (56), as did the Goldacre and Sudlow reviews (33,57). The Sudlow review notes the metaphor of SDEs as “reading libraries” rather than “lending libraries”, with person-level data remaining in the secure environment (57). The recommendation to assess potential healthcare benefits versus risks, with detail on some relevant considerations (**Table 6**), reflects the public ambivalence about commercial research and the strong preference, within the sphere of commercial studies, for research with direct healthcare benefits (such as research into treatments). Net public preference is against data sharing to companies for other kinds of research (6). The emphasis on “public benefit” accords with another study noting that the public’s concept of “public good” emphasises research that addresses real-world needs, including those affecting only small numbers of people; that addresses social inequity and inequality; and that minimises harm, e.g. by not perpetuating harmful stereotypes (13). Patient-led research is one way to focus research on these goals (58). The recommendations around transparency accord with further recent guidelines (59).

The additional concern around de-identified free-text data reflects, in part, the difficulties of “perfect” de-identification, which arises from the high level of detail, as well as sensitivity. Free text can have important research uses, both directly via clinical expert review [e.g. (60)] and by conversion to structured data via natural language processing (NLP) [e.g. (61)] before researchers are given access. The latter is one method to help safeguard privacy. A “safe setting” (TRE) is another important safeguard around managing free-text data itself (61), and the guidance reflects that (**Table 6**).

Guidance has perhaps been lacking particularly for commercial applicants seeking to use MH data for research, either via collaboration with healthcare provider organisations such as the NHS or for data collected directly with consent. The LEAG set out concrete recommendations for such organisations (**Table 7**), but also noted that the UK’s health research ethics framework might usefully offer ethical review for some studies that may technically be exempt. An example might be if companies collect and study health data, with consent, direct from users (e.g. users of healthcare “apps”), with no reference to the NHS ( **Table 8**).

The recommendations for executive/regulatory bodies (**Table 8**) also emphasise public information and citizen choice about data sharing and research participation, as well as the provision of clear and consistent guidance to NHS bodies seeking to conduct health data research. An emphasis on public communication is important for a number of reasons: transparency is one, and ensuring trustworthiness another, but potentially also willingness to contribute—the extent to which people agree that their data is valuable to society strongly predicts their support for commercial use of their de-identified health data for health research (19). Transparent communication and public awareness is also essential for methods involving opt-outs or where consent is taken as implied. The recommendation for a central mechanism to control clinical/research sharing of one’s routinely collected data is consistent with the public’s wish that the NHS should be in charge of, and administer the use of, patient data (62,63), and is an explicitly expressed public preference (6).

The strengths of this study include the development of practical guidelines in a somewhat controversial area of MH data science for which they were previously lacking; their development by members of the public with personal or other close experience of mental ill health forming a lived-experience research advisory group; and the inclusion of background information required by that group to consider the questions at hand and thus likely of use in broader public information work. The work also has limitations. While large-scale surveys can measure the ways in which their samples are biased relative to the entire population, and seek to correct for that to some extent [e.g. (6)], the present work involves a smaller number of people, whose views may not be fully representative of those of the UK populace or the subset affected personally by the issues considered. The non-quantitative methods represent in some ways a less rigorous approach to defining consensus than formal Delphi methods (45,51), though they enabled the capture of a broad-ranging set of background information requirements and individual specimen views (developed via rigorous methods) alongside the consensus recommendations, and without predefinition of statements to consider. The guidelines may require revision as use-cases and technologies develop, as for any guidelines.

Future commercial/industrial use of MH data has potential to bring benefits to patients but these potential benefits must be set against potential risks (**Table 3**, **Table 4**) (21,33). PPI is important when designing and considering individual research projects, but PPI can also contribute to framing an approach to this aspect of research in the round. We hope that the present guidelines help to do so.

## 7 DECLARATIONS

### 7.1 Availability of data and materials

Not applicable.

## 7.4 Acknowledgements

We thank Mark Avery, Hannah Clarke, Sue Fletcher-Watson, Mike Garwood, Laura Hannah, and Kay Taylor for helpful comments, and Carrol, P.S., and two other LEAG members for their contributions. In memory of the late Dermot O’Reilly, of Queen’s University Belfast, who also contributed to this work.

## 8 ABBREVIATIONS

ADP: Adolescent Mental Health Data Platform
ADR UK: Administrative Data Research UK (funded by the ESRC)
API: application programming interface
BRC: (NIHR) Biomedical Research Centre
CAG: UK Confidentiality Advisory Group (68). Created in 2013 to replace the National Information Governance Board (NIGB)
CBT: cognitive–behavioural therapy
CDA: confidential disclosure agreement (alternative name for an NDA)
CI: chief investigator
COVID-19: coronavirus disease 2019
CPFT: Cambridgeshire & Peterborough NHS Foundation Trust
CRIS: Clinical Records Interactive Search (software, or the resulting database)
DAC: Data Access Committee
DARE UK: Data and Analytics Research Environments UK (funded by UKRI, HDR UK, ADR UK)
DATAMIND: DATA hub for Mental health INformatics research Development
EHR: electronic health record (also: EPR)
EPR: electronic patient record (also: EHR)
ESRC: Economic and Social Research Council, part of UKRI
EU: European Union
FDP: NHS Federated Data Platform (England)
GP: general practitioner / general practice
GPDPR: General Practice Data for Planning and Research (England)
HDR UK: Health Data Research UK
HRA: UK NHS Health Research Authority
HSC: Health and Social Care (Northern Ireland)
ICO: UK Information Commissioner’s Office (69)
LEAG: DATAMIND Lived Experience Advisory Group (formerly Super Research Advisory Group, SRAG)
MH: mental health
ML: machine learning
MRC: Medical Research Council, part of UKRI
NDA: non-disclosure agreement; see also CDA
NHS: National Health Service (England, Scotland, Wales) or HSC (Northern Ireland)
NIHR: UK National Institute for Health and Care Research
NLP: natural language processing
ONS: UK Office for National Statistics
PPI(E): patient and public involvement (and engagement)
REC: Research Ethics Committee
RECOVERY: Randomised Evaluation of COVID-19 Therapy trial (70)
SAIL Databank: Secure Anonymised Information Linkage Databank (Wales)
SDC: statistical disclosure control
SDE: secure data environment (a broader term for a TRE)
SLaM: South London & Maudsley NHS Foundation Trust
STROBE: Strengthening the Reporting of Observational Studies in Epidemiology
TRE: trusted research environment (see also SDE)
UK: United Kingdom
UKRI: UK Research and Innovation, a public body uniting the UK’s seven research councils (which provide public funding for research)
US: United States of America

---

**Figure 1:**
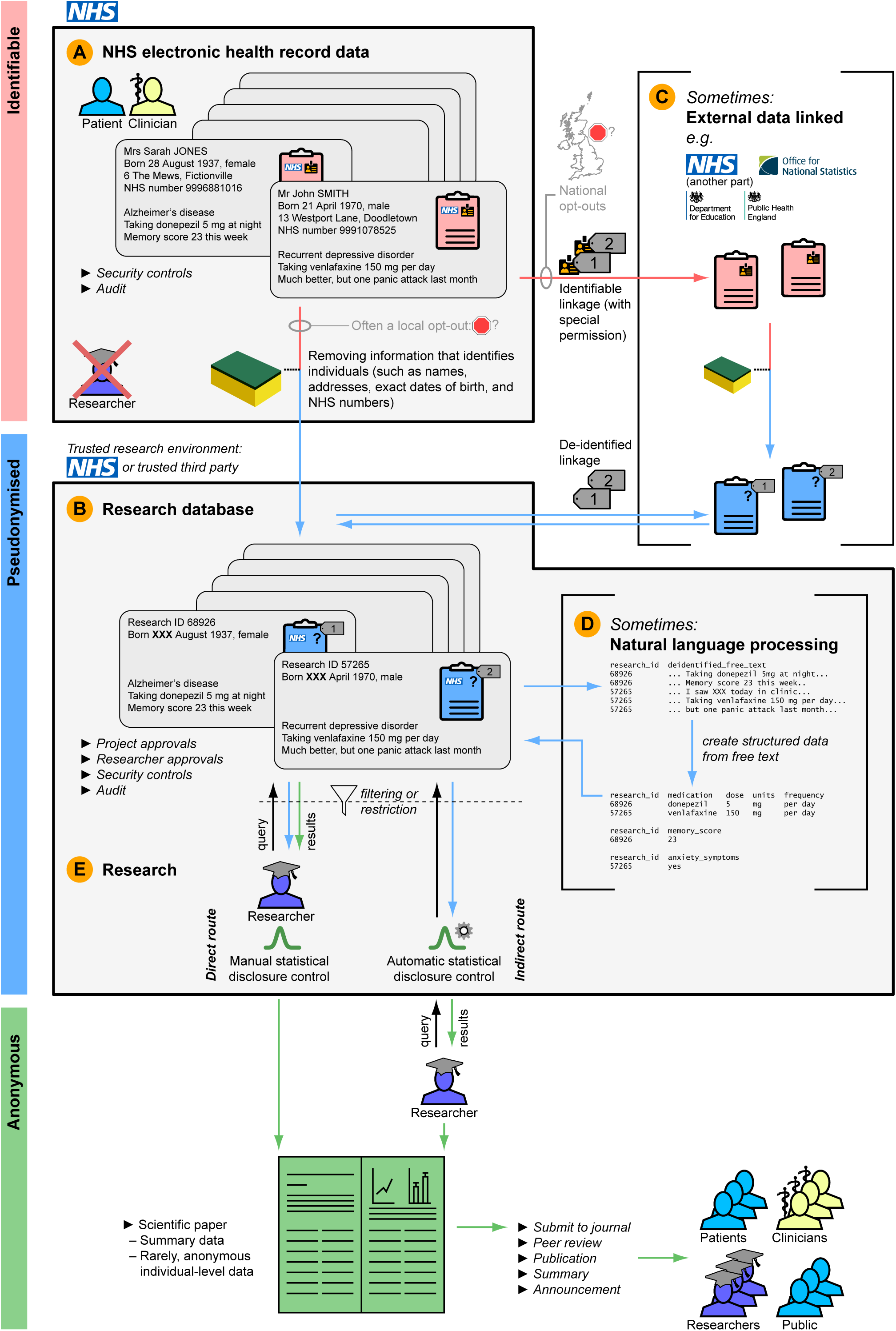
Typical journey of data from NHS electronic health records to research. Data colour coding follows **Supplementary Figure 1**. All examples are fictional and the fictional NHS numbers are from the official test range. **(A)** Typical starting point: identifiable health data held by the NHS (**Supplementary Table 1**). **(B)** De-identification creates a pseudonymised research database within a trusted research environment (TRE) (**Supplementary Table 2**), which might be within the NHS or provided by a trusted service (55,64,66,167). Local opt-outs may apply. **(C)** For some studies, additional data may be linked in, from external sources. There are identifiable and de-identified ways to do linkage ( **Supplementary Table 2**). National opt-outs may apply. **(D)** Sometimes, de-identified free text is passed through natural language processing (NLP) software to extract more structured data (**Supplementary Table 2**). **(E)** Research can then take place, by approved researchers doing approved projects (studies). The researchers might be permitted to work within the TRE, either (a) by making electronic queries of the pseudonymised data (filtered/restricted by those queries and sometimes by other restrictions such as access only to partial data) and ensuring statistical disclosure control is performed (direct route, left), or (b) by using software to isolate them from the pseudonymised data itself (indirect route, right), e.g. (168). The latter is preferable (33). Either way, anonymous results are produced (**Supplementary Table 2**). The researchers write up their findings and submit them for peer review (**Supplementary Table 1**), and then the results are published and disseminated.

