## Supplementary Materials for "Commercial or industrial use of mental health data for research: primer and best-practice guidelines from the DATAMIND patient/public Lived Experience Advisory Group"

#### CONTENTS OF SUPPLEMENTARY MATERIAL

### 1 SUPPLEMENTARY METHODS

#### *1.1 Creation of a glossary of terms*

Initially, the LEAG set out requirements for definitions of a range of terms, and explanations of aspects of health care and scientific practice, that they considered central to public understanding and engagement around this topic. Definitions and a specimen data flow were provided by academic members of the DATAMIND team and edited by the LEAG.

#### *1.2 Methods for LEAG member's review of selected existing guidelines*

The following existing guidelines and commentary articles were identified and reviewed by a LEAG member:

- Ford et al. (2021), "The challenges and opportunities of mental health data sharing in the UK" (1).
- Health Data Research UK (HDR UK) Public Advisory Board (2021), "Building trust in data access through public involvement in governance" (2), alongside some pages of HDR UK's web site (https://www.hdruk.ac.uk/).
- Cancer Research Horizons (2022), "Our guiding principles: commercial data partnerships" (3).
- Data and Analytics Research Environments UK (DARE UK) (2022), "Building a trustworthy national data research infrastructure: a UK-wide public dialogue" (4).
- Content from Understanding Patient Data (https://understandingpatientdata.org.uk/) (to 2025) (5); details below.
- Kirkham et al.'s (2020/2021) "Best Practice for Mental Health Data Science" checklist (6) and the Delphi study underpinning its development (7).

In broad terms, for each document, the reviewer considered:

- The accompanying text and/or supporting information.
- Any information on how the guidelines were developed, including:
    - Whether the specific sensitivity of unconsented MH data was sufficiently considered during guideline development;

    - Whether commercial use of unconsented MH data was specifically considered during guideline development;
    - Whether patients and the public were involved in developing the guidelines, and if so, how?
    - Whether patient and public benefits have been considered.
- The content of the guidelines, including:
    - What are specific public and patient concerns around the sharing of mental health (MH) data, and specifically unconsented MH data? Do the guidelines address these concerns or do they give cause for more concern?
    - What are the reviewer's specific concerns around the sharing of MH data, and specifically unconsented MH data? Do the guidelines address these concerns or do they give cause for more concern? Where relevant, the reviewer specified the concerns and provided reasoning. The reviewer considered also how various concerns and risks can be minimised, and noted that there may well be counter-arguments and/or other mitigations.
    - What are potential risks around the sharing of unconsented MH data and how do the guidelines reduce or mitigate these risks?
    - The appropriateness of each guideline with regards to the sharing of unconsented MH data given the sensitive nature of MH data.
    - Does the guideline rely on a procedural or technical control, or both?
    - Does the guideline place undue reliance on the commercial organisation to reduce risks?
- The wording of the guidelines, including:
    - Whether the guidelines are easy to understand;
    - Any ambiguity or vagueness in the guidelines;
    - The words used and any problematic connotations;
    - Whether necessary terms have been clearly defined;
    - Uses of absolute words (e.g. all, always, everything) or the implication of absolutes;
    - Any contradictions in the guidelines;
    - Any problematic implications of the guidelines.

- The motivation or reasoning behind the guidelines, including:
  - Any reasoning offered and/or underlying motivations for the guidelines;
  - Any problematic assumptions that have been made, on which the guidelines are based;
  - Where findings from prior research studies are offered as reasoning for any guidelines, what are the strengths and limitations of those studies, and are the findings generalisable and/or applicable to unconsented MH data?
  - Where reasoning for a given guideline is included in the wording of the guideline itself, and this reasoning is problematic and/or may impede the implementation of the guideline, this has been specified in the review.
- Completeness of the guidelines, including:
  - Any incomplete information or incompleteness in the guideline(s);
  - Whether the guidelines could be meaningfully/properly implemented as they are or whether additional requirements or guidelines may be needed.
- Inclusions and exclusions for guidelines, including:
  - Any inclusions and any exclusions that apply for any guidelines;
  - For exclusions that apply for any guideline, are these specifically stated or left open to interpretation?
- Whether there are other benefits for certain proposed guidelines, especially if these have not been articulated clearly within the literature reviewed. If so, whether such benefits have been specified in the review of the respective guidelines.
- The structure of the guidelines (e.g. whether the guidelines are categorised, and if so, how?).

Then, in detail for each guideline, in relation to developing guidelines for sharing unconsented MH data with commercial organisations, the reviewer considered the following:

- Elements worth including, such as:
  - Accompanying text or guidelines that address some public and patient concerns or concerns that the reviewer had around the sharing of unconsented MH data.

  - Accompanying text or guidelines that would promote the implementation of procedural, technological or systemic aspects/controls that would help to reduce risks.
  - (In some cases, suggestions for amendments were made.)
- Elements worth including with amendments (with reasoning).
- Elements noted to avoid, with reasoning (e.g. why a particular statement or guideline may not be appropriate in relation to the sharing of unconsented MH data with commercial organisations, even though it may be acceptable for sharing other types of data).
- Any obvious gaps.
- Public involvement approach:
  - PPI in developing the guidelines/recommendations;
  - Consideration of patient and public benefits and concerns;
  - Consideration of PPI in processes concerned with data access, monitoring, and management (e.g. does the guideline itself advise the incorporation of PPI? An example of this might be: data access applications to be reviewed by experts and members of a PPI panel);
  - Consideration of the wider public in the guidelines (e.g., public accountability and scrutiny);
  - Any other indications of the approach towards the public.
- Industry involvement approach:
  - Industry involvement in developing the guidelines/recommendations;
  - Consideration of specific risks presented by industry access to data;
  - Whether specific industry sectors have been considered or whether industry is regarded as one homogeneous entity;
  - Consideration of industry involvement in processes concerned with data access, monitoring, and management (e.g. do guidelines stipulate specific requirements for industry applicants?).
- Strengths and limitations of the guidelines (in summary):
  - Strengths and limitations with regards to how they relate to the sharing of unconsented MH data with industry (e.g. guidelines for sharing consented research trials data may omit various considerations);

   - Strengths and limitations with regards to how the guidelines were developed (e.g., the way in which public contributors were involved);
   - Strengths and limitations with regards to the content of the guidelines;
   - Strengths and limitations with regards to the wording of guidelines.
- Equality, diversity, and inclusion (EDI) approach:
   - EDI considerations for PPI;
   - EDI considerations when developing guidelines;
   - EDI considerations in guidelines.
- Any other considerations or comments.

The results of this process are presented here, and were fed into the development of the guidelines, but are not themselves the guidelines thus developed (for which, see **Results**).

Note that the terms "data host", "data controller", and "data owner" are used interchangeably throughout the review and (unless otherwise stated) all refer to the organisation that owns the data and is responsible for the data.

### 2 SUPPLEMENTARY RESULTS

#### *2.1 Glossary of terms*

We provide these here as a primer:

- Definitions of core terms around health data and health research (**Supplementary Table 1**).
- Definition of technical terms used in MH data science (**Supplementary Table 2**, **Supplementary Figure 1**).
- Definitions around health information governance (**Supplementary Table 3**).
- How do the regulations and processes governing health data research differ across UK nations (**Supplementary Table 4**)?

#### 2.2 LEAG member's review of existing guidelines

##### 2.2.1 Ford et al. (2021)

The LEAG reviewer's broad comments on this opinion paper (1) were as follows:

> *In summary, this document focuses primarily on the challenges faced by researchers when trying to access datasets in the UK (including healthcare data). There is also substantial focus on obtaining access and permissions for data linkage. While they purport to propose solutions from a MH research perspective, the recommendations are general and there is no separate consideration of the higher sensitivity of MH data. The focus is much broader, covering the linkage and analysis of data from across different sectors including healthcare, and "administrative datasets." Arguably, there was not sufficient consideration of the specific sensitivity of MH data. There is also no specific or separate consideration of applicants from industry, and the unique challenges this presents.*

A detailed review is provided in **Supplementary Table 5**.

##### 2.2.2 Health Data Research UK Public Advisory Board (2021)

The reviewer's contextual comments on this document (2) were as follows:

> *HDR UK is a national charity organisation, focused on health data science, funded by ten funders, nine of which are the largest government and charity research funders in the UK: Medical Research Council (MRC), National Institute for Health Research (NIHR), British Heart Foundation (BHF), Cancer Research UK (CRUK), Economic and Social Research Council (ESRC), Engineering and Physical Sciences Research Council (EPSRC), Health and Care Research Wales, Chief Scientists Office (CSO) and Health and Social Care R&D N. Ireland. HDR UK Public Advisory Board (PAB) is a group of members of the public.*
>
> *UK HDR Alliance "is an independent alliance of leading healthcare and research organisations united to establish best practice for the ethical use of UK health data for research at scale" (https://ukhealthdata.org/members/). There are currently 106 Alliance members, including various universities, charities, research programmes, NHS Trusts, and trade associations.*
>
> *In Feb/March 2021, a survey of 45 UK HDR Alliance members was conducted to better understand their current and future plans for "patient and public involvement in the assessment of data access requests and monitoring of data use." Only 20 Alliance member organisations responded (out of 45), with 22 survey responses (responses from different staff at one or two organisations). The survey revealed "only a few examples of excellence in public involvement and transparency" and "a lack of consistency in how data access requests are assessed."*
>
> *The questionnaire covered "patient/public involvement in data access committees," "criteria for assessing data access requests" and "publication and monitoring of data access approvals." Therefore, it did not focus on all aspects relevant to data sharing for research purposes, e.g. technical environment, how*

> *data is accessed/methods of data access, what (type of) data is available for sharing, contractual agreements. Arguably, some of this was implicitly covered when "respondents were asked what criteria they use to assess SAFE use of data (Safe Projects, Safe People, Safe Data, Safe Settings, Safe Outputs) when assessing data access requests."*
>
> *It does not appear that the survey was specifically focused on the sharing of unconsented data, MH data, unconsented MH data, nor on sharing data with commercial organisations. Rather, it appears that it was broader in focus, covering the sharing of health data in general, with any organisation (not only commercial organisations) for planning or research purposes. Thus, there was not sufficient consideration of the specific sensitivity of MH data.*
>
> *This work included a review of: (a) HDR UK's PAB's Paper "Building trust in data access through public involvement in governance" (June 2021). This paper, compiled by PAB, summarises survey findings and offers recommendations. (b) Some pages of HDR UK's website. Note that the survey and the full survey findings have not been reviewed.*

A detailed review is provided in **Supplementary Table 6**.

##### 2.2.3 Cancer Research Horizons (2022)

The reviewer's summary comments on this set of stated principles (3) were as follows:

> *Cancer Research Horizon's (CRH's) guiding principles for commercial data partnerships are principles that are adopted by CRH when establishing Commercial Data Partnerships, essentially when sharing data with commercial entities. There is no separate consideration of different industry sectors, and "industry" appears to generally be regarded as a homogeneous entity. In terms of the data that is in scope for sharing (and that these guidelines pertain to) this is largely consented research data: "Data about a patient generated through clinical trials or research projects performed outside of standard clinical practice." This may sometimes include data "about a patient collected as part of standard clinical practice… where it is linked to data generated in a research project to provide clinical context" or where it has "been built on through curation, annotation or linkage as part of a research project." Thus, clinical data is only included where it provides context or annotation to data generated in research; clinical data that is independent of research data is not included. Hence, these guidelines consider implications specific to sharing data with industry, but do not consider the sharing of unconsented data and the sharing of unconsented sensitive MH data.*
>
> *This work included a review of (3); these guidelines cover the sharing of health data with commercial entities in the UK, but specifically pertain to cancer-related data, primarily obtained via research trials (i.e. consented data). A review was also conducted of CRH's Data Partnerships registry (8,9).*

A detailed review is provided in **Supplementary Table 7**.

##### 2.2.4 DARE UK (2022)

The reviewer's summary comments on this report (4) were as follows:

*Data and Analytics Research Environments UK (DARE UK) is funded by UK Research and Innovation (UKRI), as part of the Digital Research Infrastructure programme, and is a multi-phase programme. DARE UK has been established to design and deliver a novel and innovative, coordinated, and trustworthy, data research infrastructure for the UK, with a specific focus on supporting cross-domain linkage and analysis of sensitive data for public good.*

*Phase 1 of the programme, "Design and Dialogue", is a "listening exercise" led by Health Data Research UK (HDR UK) and Administrative Data Research UK (ADR UK) to understand better what a national data research infrastructure that is efficient, trustworthy and supports research at scale looks like for various stakeholders including the public. In some DARE UK materials, phase 1 is specified as running from July 2021–March 2024; elsewhere it is specified as running from July 2021–Aug 2022. Phase 2 (timelines to be clarified) is "Build, Test and Establish". Phase 3 (timelines to be clarified) is "Deliver, Optimise and Federate".*

*Note that DARE UK does not focus on MH data, and does not even focus on healthcare data, but rather the focus is much broader, covering the linkage and analysis of sensitive data from across different sectors including education, healthcare, and the environment. While this broader focus would mean that MH data would be included, it could also mean that there was not sufficient consideration of the specific sensitivity of MH data. This is especially important as DARE UK's focus is the linkage and analysis of sensitive data from across different sectors, which would potentially include the linkage of sensitive unconsented MH data with other data, thus increasing the sensitivity of the data, privacy concerns and other risks.*

*Alongside the primary Phase 1 output (4), the associated public dialogue workshop materials were also reviewed, together with DARE UK's "Involving the public" information (10). The report (4) is based on a UK-wide public dialogue to understand better:*

► *what a national data research infrastructure that is efficient, trustworthy and supports research at scale looks like for members of the public;*

► *whether public views have changed since the beginning of the COVID-19 pandemic;*

► *gaps in public views, for example regarding methods of data access.*

*The findings from the public dialogue were developed into six sets or areas of recommendations.*

A detailed review is provided in **Supplementary Table 8**.

##### 2.2.5 Understanding Patient Data (2025)

The reviewer's summary comments on these materials (5) were as follows:

*Understanding Patient Data (UPD, https://understandingpatientdata.org.uk/) was set up in 2016 and is currently a hosted organisation of the NHS Confederation. Although it is "funded by Wellcome, the Medical Research Council, the National*

*Institute for Health and Care Research and NHS England," it claims to be independent in decision-making and voicing views. Its remit includes all UK countries as well as international collaborations (https://understandingpatientdata.org.uk/about-us).*

*UPD is focused on routinely collected patient data that is "used for purposes beyond individual care without explicit consent." Its aim is "to make the way patient data is used more visible, understandable and trustworthy, for patients, the public and health professionals." It does this by creating resources, understanding people's views, developing a community of those who care about patient data, influencing and advocating (insights for policy, advice, and consultancy for organisations) and communications and media (https://understandingpatientdata.org.uk/about-us).*

*Although UPD covers the use of unconsented healthcare data, it does not focus specifically on MH data. Similarly, while UPD covers the use of health data beyond individual care, it does not focus specifically on the sharing of health data with commercial organisations. Rather, it is broader in focus, covering the sharing of any routinely collected health data, with any organisation (not only commercial organisations) for planning or research purposes. Thus, there is not sufficient consideration of the specific sensitivity of MH data, nor of the specific issues of sharing sensitive MH data with commercial organisations. Arguably, commercial organisations are considered in research commissioned by UPD (11) but this is alongside other third-party organisations (including charities and academia).*

*It was provisionally decided that we would individually review the following sections of the UPD site, accessible via the tab for "Research and Resources":*

- *Guide to explaining the use of health data;*
- *Trustworthy use of patient data;*
- *Examples of public and patient engagement;*
- *Summary of public attitudes to the use of data;*
- *Public views on third-party use of NHS data.*

*During the course of the review, we decided to omit the "Examples of public and patient engagement" section as it only contained links to other sites and no references to industry use of patient data. I also decided to omit the full findings report for UPD-commissioned research (12), due to resource constraints. Instead, only the report summarising, analysing, and building on findings was reviewed (11). The above-mentioned sections that were included in the review correspond to the following specific content:*

- *(13) [about trustworthy use of patient data] (including videos and textual content)*
- *(14) [guide to explaining how patient data is used]*
- *(15) [summary of public attitudes to the use of data] (including video and textual content).*
- *(16); this document summarises published research from 2010–2018 on UK public attitudes towards the use of patient data for research.*

- *(17); this document is about published research from 2018–2021 on UK public attitudes towards the use of patient data for research.*
- *(18); referenced by UPD with regards to public attitudes to patient data use. Not properly reviewed, but included as a reference in the tables below.*
- *(19); public views on third-party use of NHS data.*
- *(11); summary and analysis of research findings and the wider policy context.*

*I subsequently found other highly relevant content on the UPD site which was included in the review:*

- *(20) ["Can private sector organisations access patient data?"]. Note that this content covers commercial organisations accessing patient data for a range of purposes including for the delivery and provision of healthcare (e.g. pharmacies and private healthcare providers); this content is not limited to commercial entities that access patient data for research purposes. [The present] review is focused on sharing patient data with commercial entities for purposes beyond healthcare delivery and provision.*
- *(21) [frequently asked questions]*
- *(22) [UPD, "About us"]*
- *(23) ["Defining 'public benefit' for data use"]*
- *(24); this document is about published research from 2021–2024 on UK public attitudes towards the use of patient data for research.*

*One piece of UPD content reviewed (11) was a summary of research commissioned by UPD "and NHS England, in partnership with the Ada Lovelace Institute and the Office for Life Sciences (OLS)" (19). "The Ada Lovelace Institute is a… research institute and deliberative body dedicated to ensuring that data and artificial intelligence (AI) work for people and society" (11). The research sought to find out what the public thinks constitutes a fair data partnership between the NHS and third-party organisations. Specifically, the research study asked 'What constitutes a fair partnership between the NHS and researchers, charities and industry on the uses of NHS patients' data and NHS operational data?' (11). Note that this question assumes that there will be (data) partnerships between the NHS and third parties including academia, charities, and industry, and is not questioning whether such partnerships should exist. Rather, the question is focused on the form that such partnerships should take. A mixed-methods deliberative approach was taken and included "discussions with patient advocacy groups, three citizens' juries and a nationally representative survey of over 2,000 people" (19). "Deliberation emphasises logically building people's understanding about a topic, drawing out multiple perspectives and trade-offs rather than driving at a consensus and allowing time for views to be expressed and developed" (4). Note, however, that the citizens juries only ran for 2.5 days (in Sept 2019) which included the time spent on building understanding and expressing views, and arguably, this would not have allowed enough time for jurors to develop sufficient understanding on the topic. The report reviewed builds on the research conducted, has been produced jointly*

> *by UPD and the Ada Lovelace Institute, and it analyses and responds to "the jurors' deliberations and the survey findings" (11).*
>
> *Since many UPD statements refer to or rely on prior research, it is important to be clear that there are substantial limitations of prior research. This includes the limited amount of prior research on public views on the sharing of MH data, and the lack of deliberative research on public views on the sharing of MH free-text data.*
>
> *It is important to recognise that there is substantial content on UPD's website and hence it [was] not possible within this exercise to review all of it. Further, inevitably, the website continues to be updated, with new content added. As all UPD content was not reviewed, please view all gaps identified or "Elements worth including with amendments" with caution and it may well be that those gaps are covered elsewhere on the UPD website and/or content.*

A detailed review is provided in **Supplementary Table 9**.

##### 2.2.6 Kirkham et al. (2020/2021)

The reviewer's summary comments on these materials (6,7) were as follows:

> *This study aimed to co-develop a "best" practice checklist with those with lived experience of MH conditions. The Delphi method, with three rounds in early 2020, was used to create the checklist.*
>
> *The study argues that the use of unconsented or routinely-collected data for MH research could be a way to address the difficulties with recruiting and retaining (a representative sample) of participants for research (7). No doubt there are many cases where the use of existing data would significantly reduce costs and speed up the research process. However, arguably, there will always be a need for research involving participants (e.g. to test new treatments or interventions) and study attrition is not the reason for findings being "based on a sub-sample of those actually living with mental illness" (7) (as the study argues) as arguably by definition all research findings are based on a sub-sample of the total affected population. While research using unconsented or routinely-collected data would arguably be more representative and be based on a greater proportion of the population than any participant study, it would still be based on a sub-sample (for various reasons, including: not everyone gets diagnosed, not everyone gets diagnosed correctly, not everyone is engaged with the health system, not everyone seeks help from the NHS, errors in coding illnesses or patient details, some people opt-out of sharing their health data for secondary purposes).*
>
> *The co-produced checklist is "designed to complement other guidance regarding good practice within data science" (7), including:*
>
> - *The UK Data Ethics Framework (25);*
> - *The UK Government's Code of Conduct for Data-Driven Health and Care Technology (26);*
> - *Development of data governance for the use of clinical free-text data (27).*

*The study states that the checklist encapsulates "the perspective of people with lived experience of mental illness, without making recommendations that contravene existing data science frameworks" (7). This may suggest that the parameters around the study involved not questioning the validity or appropriateness of existing frameworks/codes. While the twenty participants had "personal experience of mental illness", it is important to note that they were required to also have "expertise in data science" (which may include research experience) (7). The reason given for this was so that "the participants themselves were in a position to weigh up the relative merits of the information from both" mental illness and data science perspectives and so that there was reduced "need for researcher involvement in handling potential trade-offs" (7). Arguably, when participants have "expertise in data science" and/or relevant research experience, this means they are significantly more likely to be pro-data research and pro-data sharing for research purposes. Certainly, such participants would not be representative of the wider population living with mental illness. It is highly unlikely that the perspectives of those who are against or who are more cautious about the sharing of mental health data were included. This does not mean that no public participants with data science experience should have been recruited in a study of this nature, but rather that making this a condition of participation is problematic and counteracts one of the advantages of conducting a Delphi study—that of including diverse perspectives and subsequently building consensus through several iterations. It is important to have the involvement of those with diverse perspectives and equal representation of each broad perspective because what "passes" each phase in a Delphi study is based on consensus opinion.*

*The researchers, together with a group of stakeholders, "four people with lived and professional experience of mental illness and two psychiatry researchers", decided the statements that were used in the first phase of the Delphi study (7). Arguably, there was a lack of sufficient involvement of those who only had lived experience of mental illness in putting together the initial list of statements. It is important to note that the initial list of statements may have framed thinking and perspectives on the study, introducing some bias or influence. While study participants could use the free text option to recommend the inclusion of further statements, the relatively homogenous group of participants and the way in which the initial list of statements were compiled, may mean that important public concerns were missed. Scanning the list of statements, it seems that some areas have been omitted, notably:*

- *Auditing/reviewing/monitoring the actual access and use of data;*
- *Whether access should be restricted by type of data, e.g. no access for certain parties to free text data;*
- *Requirements relating to the data access request and approvals process;*
- *Industry/commercial access to data.*

*The co-produced checklists—one for now and one for the future—received "comprehensive support" from participants, both for individual statements and the complete checklists (although note the limitations of the study specified above and below).*

*The guidelines cover (i) both consented (research trials) and unconsented (routinely-collected) MH data; (ii) the sharing of MH data with academics, clinicians, researchers, scientists. Thus, there was no specific consideration of the sharing of unconsented MH data with commercial organisations.*

A detailed review is provided in **Supplementary Table 10**.

### 3 SUPPLEMENTARY TABLES

#### 3.1 Supplementary Table 1: Basic terms about health data and research

| *Area* | *What is meant by:* | *Answer:* |
|---|---|---|
| ***Health services and health data*** | | |
| | **Data** | Data means information. This is a very broad term. In health research, we usually mean information (data) about a person, stored electronically (on computers). (Scientists traditionally speak of data as plural—"one datum", "data were analysed"—but in computing and general use it is often treated as a singular or uncountable term—"data was analysed".) |
| | **Patient/service user** | Someone who uses health care services such as GPs, hospitals, and clinics. |
| | **Clinician** | A member of staff in a health care service (such as a nurse, doctor, or psychologist) who delivers care to patients/service users. |
| | **NHS** | Shorthand for the National Health Service (NHS) (England, Scotland, Wales) or Health and Social Care (HSC) (Northern Ireland), the UK's publicly funded health care systems. |
| | **Electronic health record (EHR)** | A person's health records that are held digitally, on computer (as opposed to on paper). Also known as an electronic patient record (EPR). |
| | **Clinical/medical/ health data** | A person's information about their health care. Data in NHS EHRs may include:<br>• Simple data, in the form of a table, which are easy for computers to work with. Medical records contain data to identify a patient (e.g. name, date of birth, sex/gender, NHS number, address/contact details). They also contain information about health and healthcare processes (why the patient is being seen, any illness or condition they have, and what treatment or care they have received). This information is entered by staff or automatically (e.g. dates of referrals and appointments, codes representing diagnoses or problems or operations, blood test results, and medication prescriptions). Records may also contain information provided directly by patients (e.g. questionnaires or surveys).<br>• Text without a particular structure ("free-text" data), such as notes or correspondence typed in by staff or provided by a patient.<br>• Images (pictures), such as chest X-rays, or CT or MRI scans, where parts of the body can be seen.<br>• Other forms of complex data, such as genetic information (gene sequences from DNA). Genetic sequencing is currently uncommon in the NHS but is likely to increase over time. |
| | **Mental health data** | Medical data relating to mental health (psychological or psychiatric) problems and health care. There is no sharp dividing line between mental and physical health, because physical problems can affect mental health (through their psychological effect, such as stress and worry, or by affecting brain function directly) and mental health can affect physical health. In the present work, however, when "mental health data" is referred to, we mean data relating to psychological/psychiatric health problems. |
| | **Routinely collected health data** | Data that is collected by health services as part of their normal operation, such as when patients go to their GP or a hospital (as opposed to data specially collected for a research study). |
| | **Administrative data** | Data that is collected by public services for administrative or management purposes, both within health services (e.g. the numbers of hospital admissions on a given day) and beyond (e.g. data on air pollution or taxes) (28). |
| ***Health research and its types*** | | |
| | **Medical research** | A process to investigate a health-related question. Research is conducted in "studies" or |

| | | |
|---|---|---|
| | | "projects". |
| | **Public benefit** | When health data is used for research, that research doesn't usually benefit the contributing patients (who are part of the research) directly, but there is an expectation that the research is for "public benefit" in some way in the longer term (18,29). The expected benefits of a research study commonly relate to learning more about the causes, nature, or consequences of a disease/condition, or how best to treat it, so that others can be helped. Examples might include:<br>• Looking at who does and doesn't get a particular condition, to discover what might put people at risk of getting it.<br>• Studying people with a disease or condition in detail, to understand their problems or to develop new ideas about how to help them.<br>• Conducting a trial of a new treatment with volunteers who have a particular condition, to see if it works.<br>• Studying people who have had a certain treatment, to see how well it works or what side effects it has.<br>These days, the published results of research funded by UK public bodies must be made available freely to everyone (30). Nearly all published medical research can be found through PubMed (https://pubmed.ncbi.nlm.nih.gov/). |
| | **Participatory research** | Research in which people take part directly and meet the researchers. This includes studies in which information is collected about people (e.g. completing a questionnaire, having a brain scan) but also studies in which new treatments are tried (e.g. taking a new medicine or a placebo). |
| | **Epidemiological or observational research** | Research in which data about people is analysed. Although this might be participatory research (the researchers meet people and collect data from them), this kind of research often uses routinely collected data from large numbers of people. Using information from a very large number of people often makes the research better able to find answers. However, conclusions from routinely collected data are tentative, because "correlation does not imply causation". If event A is associated (correlated) with event B, is that because A causes B, because B causes A, because X causes both A and B, or because it was a chance finding? Strong conclusions may require randomised controlled trials (see below). |
| | **Clinical trial** | A trial (research study) of a new treatment, such as a medicine. Clinical trials of medicines, known as CTIMPs, have special extra rules (31). |
| | **Randomised controlled trial** | A proper experiment to test an intervention, such as a new medication. In a typical study, people with a condition are randomly assigned to one of two conditions. One group is given the new medication. The other is given a placebo (dummy) medication as the "control" condition. Neither the patients nor their clinicians know which is which (this is called "double-blind" as neither knows). If a difference is found, this is likely to be due to the medication. (Compare epidemiological research, above.) |
| ***Running and overseeing research*** | | |
| | **Chief investigator (CI)** | The investigator (researcher) with overall responsibility for a research study, and the person who seeks ethical approvals. |
| | **Principal investigator (PI)** | The researcher in charge of a study at a particular site (e.g. hospital). For a research study at a single site, this is the same as the chief investigator. |
| | **Research approvals** | All research involving identified NHS patients directly must be approved by an NHS Research Ethics Committee (REC), to ensure it is planned and conducted in an ethical way, and is for public benefit. RECs are independent of the researchers, diverse, and include people who are expert in different aspects of research (expert members) and members of the public ("lay" members) (32). Research using de-identified NHS patient data (research databases) may be reviewed by a REC (29,32). (See **Supplementary Table 2** for an explanation of "de-identified".) NHS RECs are part of the UK Research Ethics Service. Their procedures are set out transparently (29). Some types of research need approvals from other regulatory or NHS organisations as well. (There are other kinds of RECs too: for example, research in a university with healthy volunteers would |

| | |
|---|---|
| | usually be approved by a university REC, not an NHS REC.) |
| **Peer review** | The results of research are normally submitted to a scientific journal as a manuscript or paper. The journal editor sends the paper to other experts in the topic (peer reviewers or referees). The reviewers should consider questions like whether the methods are sound, and whether the conclusions are supported by the results. They may recommend publication (often after requiring amendments) or rejection. Peer review is not a guarantee that the research is correct, but it remains the "gold standard" for research (33–35). |
| ***Participation, involvement, and engagement in research*** | |
| **Mental capacity** | The ability of a person to make an informed decision. In the UK, the law sets out what this means (36). Research involving people whose mental capacity is impaired has to follow special rules to keep participants safe (37). Capacity can fluctuate, and is decision-specific. People must be assumed to have capacity unless demonstrated otherwise (36). |
| **Informed consent** | Consent is permission for something to happen. Normally, medical research studies in which people take part may only involve volunteers who consent (37). Informed consent is an internationally agreed ethical principle of participatory research (37). "Informed" means that before deciding, the volunteer should understand all relevant aspects of the study, including its aims and potential risks and benefits (37). Participatory research involving people who lack mental capacity to consent is only allowed if the research is specifically about the condition that impairs their capacity (e.g. research into dementia) (37). Consent is typically not required for non-participatory epidemiological research using routinely collected data (where researchers use data already collected and people do not participate) if nobody could be identified from the research (38). |
| **Patient/public engagement in research** | "Engagement" means providing information and knowledge about research to patients and the public, including through speaking with researchers, via web sites and papers, or at events where the public are invited to attend (39,40). |
| **Patient/public involvement in research** | "Involvement" means that research is done "with" or "by" patients or members of the public (e.g. as advisors or researchers), rather than "to", "about" or "for" them (39,40). PPI informs research questions and research design with public, patient, and carer opinions (41). PPI does not itself require ethical approval (41). ("Participation" is different; see "Participatory research" above.) |
| **Plain-language ("lay") summary** | A description of something (like a research study's methods or results) that can be understood by people without a background in scientific research and with an average level of literacy. |
| ***Not research, but similar*** | |
| **Clinical audit and service evaluation** | Clinical audit and service evaluation are different from research, but they can look quite similar. "Clinical audit" (often "audit") examines the performance of a health service against agreed standards (42), usually coupled with efforts to improve the service, and repeating the audit regularly to check that services are performing well or improving. Some UK audits are national, meaning lots of NHS organisations participate. For example, the National Clinical Audit of Psychosis examines standards such as whether physical health screening is offered regularly to people with psychosis (43). "Service evaluation" is a broader term meaning measuring the standard achieved by a service (like audit, but without reference to a standard) (44). Audits and service evaluations are conducted by NHS bodies themselves, without the need for research approvals (44). Both involve the analysis of routinely collected data by the organisation that did the collecting. |

For abbreviations, see **Supplementary Abbreviations**.

##### 3.2 Supplementary Table 2: Some common technical terms used in data science

| *Theme* | *What is meant by:* | *Answer:* |
|---|---|---|
| ***Structure: What is the "shape" of the data?*** | | |
| | **Variable** | A quantity that can vary (take different values). Used in computing as the name of a data item, such as "date of birth", "haemoglobin level", or "diagnosis". |
| | **Database** | An electronic collection of data. Most databases are a collection of tables. Each table is a grid with rows and columns, about a single concept (e.g. "referrals"). Every column is a simple variable (e.g. "referral_number", "referral_date"). Each row or "record" relates to a single instance (e.g. one referral). Each row/column intersection ("value" or sometimes "field" or "cell") contains a single value (e.g. "2023-01-01"). Values are sometimes allowed to be missing (blank or "null"). Tables frequently cross-reference each other through relationships, where one record refers to another, often in a different table (a "relational" database) (45,46). Databases can contain any kind of data, including data such as images (e.g. scans) and text (e.g. clinical notes). |
| | **User interface (UI)** | The way a human interacts with a computer. Databases are not very human-friendly, so EHR systems (**Supplementary Table 1**) typically present the information in a clearer way to patients and clinicians, and try to make it easy for humans to enter data. For example, whereas a database might contain information about thousands of patients, an EHR UI would focus on one patient at a time. A good EHR UI would make it easy to find important information quickly, would display alerts prominently (e.g. allergies), would make it quick to enter new information, and might offer "decision support" (e.g. "those two medicines sometimes don't mix well; are you sure you want to prescribe that?"). |
| | **Structured data** | Data that is in a format suitable for computer analysis (see also **Supplementary Table 1**). This typically means a database (see above) of simple values. For example, in an EHR system, there might be tables like "patient", "referral", "diagnosis", and "blood test". The "diagnosis" table might contain columns like "patient number", "diagnosis code", "start date", and "end date". Other kinds of complex structure may also be used (e.g. for genetic information). |
| | **Unstructured data** | "Unstructured" data is something of a misnomer: all data has some structure, as data with no structure whatsoever doesn't convey any information. But researchers tend to use this term for data whose structure is limited or difficult to analyse electronically. That might include free text (see below) or images (e.g. pictures from X-rays or scans, or letters that have been scanned in). |
| | **Free text** | Humans find it easier to write in sentences than to enter information into databases. Patients write to clinicians and provide comments to health services. Clinicians write letters to each other and make notes in health records. This means that EHR systems often contain large quantities of text like "I met Mr Smith today. He has type 1 diabetes." This is known as "free" text because the person is free to write anything, without electronic constraints. Text is easy for humans to understand, but much more challenging to use for research. Free text can be lengthy and extremely detailed, and gives rise to additional concerns around privacy and de-identification (see below). Research projects using EHRs might ignore free text and just use structured data; or employ a human expert to read free text (typically after de-identification—see below); or use natural language processing (see below). |
| | **Natural language processing (NLP)** | Computer software exists to "read" free text written in a natural (human) language, and attempt to extract structured information. For example, there are programs to find medications (47), adverse drug events (48,49), diagnoses (47), blood tests (50), recorded thoughts of suicide (51), "negative" symptoms of schizophrenia (52), and so on. NLP is difficult because grammar is complex. An NLP program to find hopelessness as a symptom of depression might need to distinguish "X is feeling hopeless" from "X used to feel hopeless but is now better", "X's spouse is feeling hopeless", and "X said he is |

| | | |
|---|---|---|
| | | hopeless at football". NLP programs are imperfect, and need checking when they are designed in one context and then used in another, but may still be very useful. NLP is mostly used for research, but as NLP improves, it may become more common in clinical practice (e.g. in GP [general practice] surgeries and hospitals) (53). |
| | **Metadata** | Data about data. An example of metadata is a description of a data table, giving the purpose of the table (e.g. "this table records referrals for psychological therapy"), the type of each variable (e.g. whether it's an integer number, a date, a code with a few possible values, or free text) and their meaning (e.g. "code P means a referral from primary care, code S means a referral from secondary care"). |
| | **Data curation** | Looking after data for other people to work with, like the curator of a museum. This can involve putting data together, quality control (finding and eliminating invalid data), describing it well so that other researchers can understand it (providing metadata), or mapping it to a standard "vocabulary" (e.g. if two databases record problems using different coding systems, can those be mapped to each other?). |
| ***Identifiability: Can people be recognised from the data?*** | | |
| | **Identifiable data** | Data that plainly identifies people, using "direct" identifiers such as names, dates of birth (DOB), addresses, NHS numbers, telephone numbers, and so forth (**Figure 1**). A fictional example: "John Smith, male, DOB 21 Apr 1970, NHS# 9991078525, diagnoses of depression and heart failure" (38). |
| | **De-identification** | Data from which direct identifiers have been removed. Typically done via pseudonymisation (see below) (54). Both structured data and unstructured data (such as text) can be de-identified (55,56). However, it can be much harder to guarantee the perfect de-identification of unstructured data such as free text. |
| | **Pseudonymisation / pseudonymous data** | Data in which direct identifiers have been removed and replaced by a research identifier (ID) "pseudonym", typically gibberish (**Figure 1**; **Figure 2**). Other direct identifiers may be made less accurate and informative (e.g. only year of birth rather than full date). For example: "Research ID *c430c2f7a298b4e7ccd8dd763e1d85f6*, male, born 1948, diagnoses of depression and heart failure." It may be possible and permitted for some people or organisations to re-identify the person, but impossible for others (54,57). For example, the NHS Trust that performed the pseudonymisation could look up that *c430c2f7a298b4e7ccd8dd763e1d85f6* is John Smith, but researchers analysing the data could not. |
| | **Jigsaw attack** | The process of attempting to re-identify someone from de-identified or supposedly anonymous data (58). It involves combining several pieces of information about a person, like the pieces of a jigsaw, to identify them. (A simple fictional example: imagine de-identified health data says "57-year-old woman with diabetes hit by a bus in Ely on 1 Jan 2020". A local newspaper might have reported the accident and given the person's name. Combining the two would identify the person and allow the attacker to discover that she had diabetes.) A jigsaw attack may be performed by someone malicious to learn things about an individual, or by security researchers testing de-identification systems. It is usually a criminal offence, but it can be legal in special circumstances like testing the effectiveness of privacy systems (59). Sometimes it requires surprisingly small amounts of information (60). If data is not about individuals, but about groups of people, jigsaw attacks are much harder or impossible, particularly for large groups. |
| | **Anonymisation / anonymised data** | Data from which all personal identifying information has been removed, and it is no longer possible to trace the information back to an individual (or to link it to other data). Anonymous results are the kind normally made public, e.g. in scientific journals (**Figure 1**). For example, this statement does not allow an individual to be identified: "We studied 1000 people with depression, and 42% had tried cognitive–behavioural therapy." See also statistical disclosure control, below. [In the past this term has sometimes been used to mean the same as "de-identified" (50), but now the term "anonymised" is generally used to describe "strong" identity removal, where there is no possibility of re-identifying someone from the data (54,61).] |

| | | |
|---|---|---|
| | **Statistical disclosure control (SDC)** | The process of ensuring that information about identifiable individuals cannot be extracted from data, even by clever mathematical techniques—that is, the process of ensuring that data is properly anonymised and protects privacy (62). A common rule of thumb is that information about people should be put together into groups of at least a certain size, such as 10 people. Groups smaller than this can be reported as containing "fewer than 10 people" rather than "one person" or "three people". Alternatively, small groups could be put together to form larger groups. This threshold is not an absolute rule, though; sometimes data from individuals couldn't identify anyone. For example, a list of referral dates from a busy health service is unlikely to identify anyone by itself, even if each date relates to a single person. (We have described SDC as being about people, which is the normal concern in health research, but in different contexts it might be about other things, such as not identifying companies from analysis of tax data.) |
| | **Manual versus automatic SDC** | In some settings, researchers work with pseudonymised data, produce anonymous results, and check (and/or someone else checks) that they are truly anonymous—for example, checking that no groups contain information relating to very small numbers of people. In a sense, this is "manual" SDC. In other settings, special software separates the researchers from the data, so the researchers ask questions and get anonymous results without ever seeing the "patient-level" pseudonymised data (63–66). The software can provide mathematical guarantees around privacy protection; this is "automatic" SDC. Human double-checking may still be required after automatic SDC. |
| | **Synthetic data** | Data that is synthesised (made up) so that it looks roughly like real data, but isn't about real people. It can be used to ensure computer programs are working properly before they are used with real data. |
| ***Processes: the safe analysis of potentially sensitive data*** | | |
| | **"Five Safes"** | The Five Safes framework (67,68) refers to:<br>• Safe data: for example, de-identifying data before researchers are given access.<br>• Safe projects: for example, ensuring that projects are reviewed by the data owners and judged to be in the public interest before being approved.<br>• Safe people: for example, ensuring that researchers are appropriately trained and approved to use data safely, and are under contractual and professional obligations of confidentiality when handling the data. Some training schemes are nationally recognised: for example, the Office for National Statistics (ONS) provides Accredited Researcher training (69).<br>• Safe place or setting: for example, that the analysis is conducted in a trusted research environment (see below).<br>• Safe outputs: that any results to be published are truly anonymous (see above). |
| | **Trusted research environment (TRE) or "safe haven"** | A secure computing environment, where data that is too sensitive to be made public can be analysed. This might sometimes mean identifiable data, or more commonly data that is de-identified but where there is some small residual risk of jigsaw attack (see above). A TRE is an example of a "safe place" (see above). TREs deal with what comes in (e.g. data, software for analysis), who gets in (authentication and authorisation), what can be done (projects and permissions), and what goes out (e.g. vetting of anonymous results for publication). Many support high-security "remote desktop" facilities, and some TREs include facilities for closed-circuit television (CCTV) monitoring of researchers (70). |
| | **Data linkage** | Joining (linking) data from more than one source, typically about individuals. For example, to study the relationships between mental and physical health conditions, it might be necessary to link data from NHS mental health services to primary care (GPs) or general hospital data. To study the relationships between health conditions and education, it might be necessary to link data from health services and a government education department. Linkage may be legally complex because it involves data from more than one data controller (**Supplementary Table 4**). Linkage may be based on straightforward rules ("two records with the same NHS number are from the same person") or based on probability ("if two records share the same forename, surname, and date of birth, they are more likely to be from the same person"). Links may be made |

| | |
|---|---|
| | using identifiable data (e.g. NHS number) or de-identified data (e.g. a research pseudonym, see above); see (71). There are good public information resources about data linkage (72). |
| **Federation** | Where multiple databases, often located in different places, can function as one (73). An example is the use of the TriNetX international federated database to investigate mental health outcomes after COVID-19 (74). A researcher might ask an electronic question (query). This is split into multiple queries that are sent securely around the internet (e.g. "how many people in your database had COVID-19, and of them, how many developed depression in the next three months?"). Each database contributes part of the answer (e.g. "at our site, we saw 500 people with COVID-19 and 10 developed depression"), and the parts are assembled before being returned to the researcher (e.g. "overall there were 25,000 people with COVID-19 and 500 developed depression"). This requires all the databases to communicate using agreed standards. |
| ***Analysis: How do researchers extract meaning from the data?*** | |
| **Statistics** | Statistical analysis involves testing hypotheses (research questions) using data, acknowledging that data is usually "noisy" (containing all sorts of sources of variation or error, some of it random, some of it not). It is underpinned by mathematical theory and has been formalised for over a century (75–77). |
| **Quantitative analysis** | Analysis using numbers, i.e. quantities. Typically this means examining the effects of particular circumstances (independent variables, e.g. "was the person given the new drug or a placebo?") on an outcome of interest (dependent variable, e.g. survival) using statistics (78). |
| **Qualitative analysis** | Analysis without numbers, looking at qualities rather than quantities (78). This is often interpretative and exploratory (79). An example: asking a focus group about a topic, and teasing out themes in their responses (thematic analysis) (80). |
| **Mixed methods research** | A mixture of quantitative and qualitative analysis. |
| **Machine learning (ML)** | Machine learning is about teaching computers to "learn": to classify or discover patterns in data, and/or to make predictions about future data. ML algorithms (which are computer programs) can work by themselves ("unsupervised") to discover patterns, or can be trained to classify or predict data automatically based on examples provided by a human ("supervised"). ML systems can perform complex tasks like detecting breast cancer in X-ray images (81). One challenge is that a ML system taught in one context may perform poorly in another (may not transfer well). Another is that a system taught by ML may be like a "black box": it might be difficult to express in commonly understandable terms how it reached its decision, making it harder for researchers to extract knowledge or for clinicians to trust its results. |
| ***Other terms: potential buzzwords*** | |
| **Data science** | Data science is generally seen an interdisciplinary research field concerned with extracting knowledge from data. It covers processes such as storing and processing data (data management, computer science) and analysing data (computer science, statistics). It also requires thinking about the specific problem (e.g. a particular disease or condition of interest) (82); after all, there is no science without data. |
| **Big data** | Jargon used when large quantities of data are being analysed. What's big? That depends on your perspective. It might mean "data from lots of people" (83) (e.g. routinely collected health data from millions of people), or "data that takes up a lot of space" (84) (e.g. DNA sequences, MRI scan images, activity data from mobile phones), or both. The term didn't feature in the international PubMed database before 2003, but was associated with over 25,000 publications in the life sciences up to March 2023. |
| **Artificial intelligence (AI)** | Artificial intelligence is the broad scientific discipline to do with intelligence or intelligence-like behaviour shown by machines, rather than living beings. It encompasses fields such as ML, NLP, and others. Because it is a broad term, and an evolving field, it is also susceptible to hype (85). |

| | |
|---|---|
| **Data mining** | Searching for previously unsuspected structure and patterns in data (86), often using large data sets, and sometimes using ML. Contrast with "hypothesis testing", where you start with a question (hypothesis) and then test it using data. One risk of data mining is of discovering spurious patterns (87). |

For abbreviations, see **Supplementary Abbreviations**.

#### 3.3 Supplementary Table 3: Some jargon used in the areas of NHS information governance and computing

| *Theme* | *What is meant by:* | *Answer:* |
|---|---|---|
| ***UK law and rules*** | | |
| | **Information governance (IG)** | The overall process through which an organisation looks after information (data). It encompasses the organisation's strategies and processes for data collection, storage, security, use, and privacy. |
| | **Data Protection Act (DPA)** | The Data Protection Act 2018 (59) is the UK's principal law governing the handling of data relating to identifiable living people ("personal data"). It implemented UK-specific aspects of the GDPR (see below) and superseded previous UK legislation (88). |
| | **European Union (EU) General Data Protection Regulation (GDPR)** | The 2016 GDPR (89) set out the EU framework for the handling of data relating to identifiable living people. Among many other things, it sets out possible legal grounds for using personal data, such as "the data subject has given consent", "a task… in the public interest", or for "scientific… research". The UK DPA (see above) was framed in its terms and set out UK-specific aspects. When the UK left the EU in 2020, the GDPR remained in UK law as the "frozen GDPR" or "UK GDPR" (90,91). |
| | **Information Commissioner's Office (ICO)** | The UK's independent authority for data protection. The ICO oversees the application of the DPA. |
| | **Data subject** | A person whose personal data is being held by a data controller. |
| | **Data controller** | A data controller is a person or organisation who decides what is done with some personal data (data about identifiable living people) that they hold. NHS organisations such as Trusts and GP surgeries are examples. Compare "data processor". All UK organisations looking after personal data must be registered with the ICO (with a very few exemptions), and this register is public. |
| | **Data processor** | Whereas a data controller decides what is done with data, data processors do what they're told (and only what they're told) by the controller, with the controller's data. For example, it would be typical that an NHS Trust pays a computing company to run its e-mail service or to run an EHR system. In this situation, the NHS Trust is likely to be the data controller, and the computing company the data processor. The data processor isn't allowed to use the controller's data for other purposes. |
| | **Data protection officer** | Where data controllers/processors are public bodies or organisations handling personal data on a large scale, they must (under the GDPR) appoint a data protection officer to advise them on data protection and monitor compliance. Data protection officers are listed on the public register held by the ICO. |
| | **Data transfer agreement** | An agreement or contract between a data controller and another organisation (such as a data processor), governing the transfer of data. |
| | **Data protection impact assessment** | An assessment of the potential risks to data subjects from a proposed type of processing, and the measures planned to address those risks and protect personal data (59). Also known as a privacy impact assessment. |
| ***Special aspects in the NHS context*** | | |
| | **Caldicott Guardian** | All NHS organisations must have a senior professional responsible for safeguarding the confidentiality of patient information (92). This person is known as the Caldicott Guardian, after Dame Fiona Caldicott, who led a series of reports on the protection and use of patient information (93,94) and became the first National Data Guardian (see below). |
| | **National Data Guardian** | The National Data Guardian for Health and Social Care advises the UK government and NHS about the processing of health and adult social care data in England (95). They are independent, by statute, and appointed by the Secretary of State for Health and Social |

| | |
|---|---|
| | Care. |
| **Pledges to use anonymous data for research** | The NHS Constitution for England promises that patients' anonymous data will be used for research and to improve the care of others (96,97). All NHS organisations across the UK are expected to participate in and support health and care research (98). |
| **NHS Act and related confidentiality law** | The use of health information in the UK is governed not only by the DPA but in England by the NHS Act 2006 (99), subsequent laws amending and clarifying it (59,100,101), associated regulations made by government ministers (102), and a duty of confidentiality in health care that comes from common law (which is to say, case law coming from court cases rather than statutes decided by Parliament) (38,103). Note that "consent" is not the basis on which the NHS holds patient records (104,105), which is most obvious when considering the enforced care of patients who may lack capacity (36) or object (106). |
| **NHS Act Section 251 approval** | Research is often conducted with the patient's consent (see **Supplementary Table 1**) or with de-identified data (see **Supplementary Table 2**). In some circumstances, research in England and Wales may be conducted with identifiable patient data without consent, if it is considered to be of sufficient public interest, under section 251 of the NHS Act (99,102). This is permitted by the GDPR and the DPA (38,59,89). This kind of research needs approvals not just from a REC (see **Supplementary Table 1**) but also the Confidentiality Advisory Group (see below) and is subject to a national opt-out (see below). (See **Supplementary Table 4** for equivalent arrangements in Northern Ireland and Scotland.) A well-known example of this sort of research is the National Confidential Enquiry into Suicide and Safety in Mental Health (107), but there are many others. Sometimes very basic identifiable information only, such as names and dates of birth, is used to link data and then removed before researchers are given access, but this may still need special approvals (38,108). There is a public register of approved projects. |
| **Confidentiality Advisory Group (CAG)** | This group, part of the NHS Health Research Authority (HRA), considers applications to use identifiable patient information for research, without consent (see above), in England and Wales. The group advises (and in practice acts on behalf of) the Secretary of State for Health and Social Care. See **Supplementary Table 4** for arrangements in Northern Ireland and Scotland. |
| **Opt in** | Where people have to choose to be "in" to participate, or they are not included (e.g. choosing to volunteer for research). |
| **Opt out** | Where people have to choose to be "out", or else they are included by default (e.g. opting out from having one's anonymous health data used for research) (96,97). |
| **National Data Opt-Out (NDO)** | The NHS National Data Opt-Out allows people to opt out from the use of their personal confidential data (identifiable information) being used for purposes beyond individual care (such as research), where consent has not been given, and NHS Act Section 251 support is the legal basis (109–112). It does not apply to anonymised data, for example, or when NHS organisations process data within their own boundaries (110). There are a number of further situations involving identifiable data where it does not apply (38,109,110). People are frequently unaware of it (38). |
| ***Computing*** | |
| **Cloud computing and cloud storage** | When another organisation provides you with computers in its data centres and looks after them, usually for a fee. Often this organisation is a very large computing services company: examples include Amazon, Google, and Microsoft. Cloud services provide some combination of storing data (big hard disk drives or a fast equivalent) and being able to analyse it (lots of computer processors and memory). The provider typically looks after things like physical security (preventing break-ins), electronic security (only permitting access by authorised users and preventing hacking over the network), and resilience (e.g. keeping regular backups, having redundant devices for when one breaks, having batteries or generators for power cuts, and maybe having redundant data centres in case of disasters). Many cloud providers allow the customer to choose the physical |

| | |
|---|---|
| | location of the data centre (e.g. Cardiff versus California), which may be important for compliance with relevant data protection law. Cloud computing is distinguished from computing "on premises", i.e. physical computers that an organisation (such as an NHS Trust) owns and looks after itself. |
| **Data security standards** | There are many formal standards about data security. NHS organisations typically assess themselves using the Data Security and Protection Toolkit (113). Private companies and data centre providers typically use related but different international standards (114). |

For abbreviations, see **Supplementary Abbreviations**.

##### 3.4 Supplementary Table 4: Relevant NHS, data protection, and research bodies across the four UK nations

| | England | Northern Ireland | Scotland | Wales |
|---|---|---|---|---|
| **Main national health service body** | NHS England (*) | Health and Social Care | NHS Scotland | NHS Wales |
| **Government health department** | Department of Health and Social Care | Department of Health | Health and Social Care Directorates | Department of Health and Social Services |
| **Lead government minister (elected politician)** | Secretary of State for Health and Social Care | Minister of Health | Cabinet Secretary for NHS Recovery, Health and Social Care | Minister for Health and Social Services |
| **Principal data protection laws** | UK GDPR, DPA | UK GDPR, DPA | UK GDPR, DPA | UK GDPR, DPA |
| **Local (e.g. NHS Trust) data protection oversight** | Data Protection Officer; Caldicott Guardian | Data Protection Officer; Caldicott Guardian | Data Protection Officer; Caldicott Guardian | Data Protection Officer; Caldicott Guardian |
| **National data protection oversight** | ICO | ICO | ICO | ICO |
| **Route for applying for research permissions** | IRAS | IRAS | IRAS | IRAS |
| **NHS research ethics service, providing RECs (32)** | RES / HRA | RES / Office for Research Ethics Committees Northern Ireland | RES / Chief Scientist Office | RES / Health and Care Wales Ethics Service |
| **NHS research oversight/support body** | HRA | Health and Social Care | NHS Research Scotland | Health and Care Research Wales |
| **Some additional NHS data laws** | Access to Health Records Act 1990; CPIR; NHS Act 2006 | Access to Health Records Act 1990; Health and Social Care (Control of Data Processing) Act 2016 | Access to Health Records Act 1990; NHS (Functions of the Common Services Agency) (Scotland) Orders 2008, 2014; Public Records (Scotland) Act 2011; Patient Rights (Scotland) Act 2011 | Access to Health Records Act 1990; CPIR; NHS Act 2006 |
| **Additional oversight bodies for use of health data** | CAG (for identifiable data without consent) | HSC R&D Division | Public Benefit and Privacy Panel for Health and Social Care (HSC-PBPP) (115) | CAG (for identifiable data without consent) |
| **National data opt-out** | NDO (110–112) | None yet (116) | None; previously via GPs (117,118); opt-in model for confidential data (119) but with exceptions (120). | None; via GPs (121,122) |

(*) On 13 March 2025, the Government announced that NHS England will be abolished and subsumed into the Department for Health and Social Care (123,124). For abbreviations, see **Supplementary Abbreviations**.

##### 3.5 Supplementary Table 5: LEAG member's review of Ford et al. (2021)

Direct quotes are from the material reviewed. (Not all direct quotes from the primary source document are thus identified.)

| Domain | Reviewer's comments |
|---|---|
| **Elements worth including** | Data custodians to *"Ensure that there is transparent, consistent, and clear information about the access process and what is required at each step."*<br>• This is important, regardless of whether there are many different processes or a single set of UK-wide processes.<br>• Consider provision of such information online and ensuring that this information is easy to find. A single dedicated website for all data access processes—whether limited to providing information or providing a comprehensive service including enabling applications to be submitted.<br>• A single website where information on all access processes is provided might be helpful. Necessary information on changes to application processes, names of datasets, names of data controllers, could all be provided on this site, which would help address the information gaps, communication difficulties (during application processes) and confusion that researchers report.<br><br>Data custodians to *"Maintain regular contact with project leaders, providing updates on data availability and application progress."*<br>• Such updates could be via online application tracking.<br><br>At Government level, there needs to be clarity provided on the remit of *"data controllers, data curators, and data processors."* This would allow for clearer roles and responsibilities.<br><br>At Government level, to *"Identify and share examples of best practice by data controllers and organisations that facilitate the sharing of data."*<br>• However, this should not be limited to researcher feedback and best practice should not be equated to whether the process is quick and simple for researchers.<br>• This would require a comprehensive consideration of what constitutes best practice.<br><br>Secure data environments for access and analysis.<br><br>Vetting of outputs.<br>• Ideally through technical controls? |
| **Elements worth including with amendments** | *"Redesign a proportionate and uniform data access process that balances the risks of privacy breaches with the benefits to science and health policy using the data sensitivity level (low, medium, or high)."*<br>• Must be co-designed with patients and public, including patients who are averse or cautious to sharing MH data. Also, co-design with researchers (potential applicants).<br>• Must consider the identifiability of the data, and not just the "risk of privacy breaches." Due to the level of detail and diversity contained within MH data, it is arguably more identifiable.<br>• Does not define/clarify what is meant by proportionate, benefits, "data sensitivity."<br>• Must explicitly indicate that MH data (and free-text MH data especially) would be considered |

highly sensitive and with a high identifiability level, and must be treated accordingly. It is important to explicitly state this because it does not appear that currently MH data is regarded as highly sensitive in the same way that data pertaining to sex change or fertility treatment is, for example. Arguably, MH data (especially free-text data) should be treated as much more sensitive given the kind of detail it contains, i.e. there is no sensible reason why data about someone undergoing IVF [*in vitro* fertilisation] is considered much more sensitive than data about someone having a MH crisis, disclosing trauma or abuse? It makes no sense, and these appear to be persisting entrenched ideas based on biases which are not being questioned.

- Patient and public benefits are not mentioned. Regardless of whether the patient/public benefits are immediate or delayed, direct or indirect, it is the responsibility of the research team to clearly articulate these. Patient and public benefits cannot be neglected just because researchers find it difficult to clearly articulate the benefits of some types of research (e.g. exploratory research). In any case, the articulation of such benefits would be required for funding applications, so why should it be any different for data access applications?
- There would be benefits to having a single *set* of UK-wide data access processes for health data, with appropriate considerations of sensitive data, such as MH data.
    - One process for data access and a separate more stringent single UK-wide process for applications to link health data with other data? Arguably linked data is more sensitive and more identifiable.
    - Not only would it be fairer for all stakeholders, but it would inevitably make applying for data access easier to understand and navigate (because there would be one set of processes and not because the processes should be simpler and quicker).
    - Easier to make changes to single set of processes in response to changing legislation.
    - Easier to understand changes to a single set of processes.
    - May mean less wastage in research, i.e. fewer abandoned projects, less public funds spent on data access application processes.
- It is not clear why currently it is easier to apply for access to older datasets compared to more recent datasets (e.g. 2007 dataset compared to 2014 dataset). Do more recent regulatory and legal changes not apply to older datasets? It is not clear from this paper why this is so. Would there be value in adopting a single set of UK-wide processes which would apply regardless of the date of the dataset? This would allow research to be conducted on more recent data which would ensure that findings are more relevant, and avoid potential harm from the use of out-of-date data.

Individual data users (researchers) should *"Identify and undertake data security and General Data Protection Regulation training."*

- GDPR training would not cover all the relevant legal and ethical issues for this kind of data, e.g. *"The GDPR sets out lawful bases for processing identifiable data; scientific research does not always need to rely on consent and is potentially exempt from one or more obligations set out by the GDPR, considered on a case-by-case basis when certain criteria can be documented."*
- There is a risk that this type of training becomes a tick-boxing exercise that has very little public benefit, e.g. consider how Equality, Diversity & Inclusion (EDI) training for recruitment and workplaces is mostly tick-boxing and with little positive impact. It would arguably be more effective to require data users to undergo regular (annual?) accredited and assessed training relevant to this specific type of data, covering up-to-date legal and ethical issues, risks, including privacy risks.
- Such training could include up-to-date information on application processes, which may be helpful for students and early career researchers.

Individual data users (researchers) should *"Obtain appropriate supervision from experienced individuals where necessary."*

- Should be mandatory for all researchers with less than a given number of years' experience to have appropriate supervision and a designated supervisor should be indicated on the application form.

Individual data users (researchers) should *"Maintain regular contact with data custodians, providing updates on changing requirements and project circumstances."*

- Regular project progress reports (in plain language) may provide transparency and reassurance that the data is being used for the approved purpose. PPI to check such reports.
- If data requirements change, there should be a *formal process* to apply for increased or modified access (e.g. access for longer term, access to more data, access to alternative data). This should *not* be handled informally through contact with data custodians.

Institutions hosting data users to *"Maintain infrastructure to support the necessary permissions, such as obtaining and maintaining a National Health Service Data Security and Protection Toolkit."*

- There are many, many more considerations for infrastructure beyond supporting the necessary permissions.
- Wherever possible it would be appropriate for technical controls to be built into the infrastructure and environment, e.g. controls around access, outputs, auditing of activity.

Institutions hosting data users to *"Ensure that appropriately trained senior staff have sufficient time to advise, review, and sign off applications."*

- Should make clear when referring to hosting organisations that this means data controllers and data owners, and not organisations that have only been contracted to provide technical storage and processing.
- Should always be a PPI panel to also review data access applications.
- To address delays and increasing delays reported by researchers, complexities, safeguards, and requirements within processes should not be removed. Instead, the following may help to address delays:
  - Adequate staffing for application reviews;
  - A single set of UK-wide processes for data access, e.g. one process for access to MH data, one for access to other health data, one process for data linkage. The burden of learning and navigating many processes would be removed for applicants and staff (e.g. easier on-boarding for new staff, easier to deal with staff turnover). It would also be easier for all concerned to learn about any changes to a single set of UK-wide processes compared to changes to hundreds of processes.
  - Multiple checks and reviews can occur concurrently, where appropriate, e.g. vetting of the organisation, vetting of the researchers, checking the planned use of the data, patient and public reviews. There is no sensible reason why all these should occur in sequence, but could safely be carried out concurrently. Thus, all the necessary complexities, safeguards and requirements would still exist within the process, but the researcher may perceive a more efficient and quick process.
  - A single online portal for all applications (assuming transition to single set of UK-wide processes). Would allow applicants to check the progress of their application online, including any information on delays, without needing to take up staff time. Necessary information on changes to application processes, names of datasets, names of data controllers, could all be provided on this site, which would help address the information gaps, communication difficulties (during application process) and confusion that researchers report. Any questions or information requests from reviewers to applicants, response from applicants could all be provided here, ensuring all information is in one place, allowing multiple staff members to respond and protecting against inefficiencies caused by staff turnover. This may also allow for comparisons between data controllers, and for systemic issues to be identified.

At the Government-level they propose the establishment of *"an All-Party Parliamentary Group to review how to optimise the safe and legal access to data, including a review of how legal requirements are being interpreted."*

- This must include appropriate patient and public involvement.

**Noted to avoid** Providing recommendations or guidelines without indicating clearly and in plain language what is meant by the terms used.

Neglecting to consider patient and public benefits in guidelines and referencing only *"benefits to science and health policy."*

Advocating for a single data access process for all data, without explicitly stating that MH data should be considered as highly sensitive and that the process should appropriately consider this.

In advocating for a single data access process, should not encourage a "race to the bottom," whereby all necessary complexities, safeguards, and requirements are removed. A single UK-wide process should not be assumed to mean the simplest and quickest process.

Should not advocate for all complexities, safeguards, and requirements to be removed. Just because not all researchers may understand why some requirements exist does not mean those requirements are not necessary. The inclusion of necessary safeguards is not necessarily diametrically opposed to process efficiency. It would be far better to consider how both could be achieved rather than sacrificing one to achieve the other (based on assumptions).

There should not be a single data access process for data access and data linkage. Separate consideration of applications for data linkage.

Recommendations for training that could become a tick-boxing exercise with little positive impact.

If data requirements change, this should not be handled informally through contact with data custodians. There should be a formal process to apply for increased or modified access (e.g. access for longer term, more data, or alternative data).

Institutions hosting data users and data custodians to *"Link those applying for data access with those who have had successful applications."*

- May enable those who should not get access to learn how to "game the system."
- Would be far more effective to have a transparent process, with sufficient information, including feedback, provided to applicants.
- May be beneficial to encourage applicants to include PPI in data access applications as this would help with articulating patient and public benefits in plain language (including for applications with delayed benefits).

Data custodians to "Provide additional low-risk, open access datasets that can be shared with institutions for student projects to ensure that we encourage and develop the next generation of data scientists."

- What is low-risk?
- How can unconsented healthcare data, especially unconsented MH data, ever be provided on an open-access basis?
- Research institutions could create dummy dataset for student training.

| | |
|---|---|
| | Advocating for *"the creation of one-off, large, multipurpose research datasets might be an efficient way to meet the needs of the research community."*<br>• This presents various privacy concerns and increased security concerns.<br>• What about the needs of patients and the public? |
| **Gaps** | The consideration of patients and the public in the operation of data access processes, e.g. data access application reviews by patients and the public.<br><br>Neglecting to consider patient and public benefits in guidelines, with exclusive focus on the needs of academic researchers.<br><br>Limited consideration of aspects beyond the access request process, e.g. no consideration of audit/monitoring/review of access. Hence the guidelines do not pertain to other important aspects which would increase trustworthiness of the system.<br><br>No consideration of data misuse, security breaches, privacy violations and any other contract breaches. No information on what steps should be taken in the event of such breaches/violations.<br><br>No stipulation for data access (and other processes related to data, e.g. data monitoring process, data breach response process) to be documented. If processes are documented, that formalises them, helps ensure that all involved understand the steps in the process, ensures consistency and fairness in application, aids transparency, among other benefits. It would also allow for audits to be performed against the documented procedures, e.g. for a random sample of access requests were the documented processes followed?<br><br>The basis for the guidelines is not a consideration of risks, but rather a focus on how to make processes easier and quicker for researchers. Thus, risks have not appropriately been considered. Risks and concerns related to industry access to data have not been considered.<br><br>Other gaps in addition; not all covered. |
| **Public involvement approach** | Have not considered patients and the public in the implementation of data access processes, e.g. data access applications to be reviewed by patients and the public.<br><br>Neglected to consider patient and public benefits in the guidelines, with exclusive focus on the needs of academic researchers.<br><br>Consideration of patients and the public is limited to encouraging the collection of more data from patients and building trust through convincing the public of the overall benefits of sharing data with researchers and showing robust and secure processes.<br>• Patient/public benefits should be demonstrated for each access request (not assumed to exist overall for all research), which should be reviewed by patients and the public.<br>• Trust should not need to be built if there is trustworthiness within the system. This is better achieved through transparency and ensuring effective controls, rather than seeking to convince the public of apparent benefits.<br><br>Lack of PPI in deliberations and in putting together recommendations. |

| | |
|---|---|
| | Lack of public consultation on recommendations. |
| **Industry involvement approach** | No reference to industry involvement in putting together these recommendations.<br><br>No reference to consideration of applicants from industry. Appears to be solely focused on academic researcher applicants. |
| **Equality, diversity, and inclusion considerations** | No EDI considerations. |
| **Other considerations** | – |
| **Summary of strengths and limitations** | Strengths:<br>• Summarises the challenges faced by academic researchers in accessing data. This information can be used to design better processes.<br><br>Limitations:<br>• Exclusive focus on academic researchers;<br>• Exclusion of patient and public perspective and considerations;<br>• Exclusion of industry applicants and considerations related to such applicants.<br>• Various other limitations, as above. |

##### 3.6 Supplementary Table 6: LEAG member's review of Health Data Research UK Public Advisory Board (2021)

Direct quotes are from the material reviewed. (Not all direct quotes from the primary source document are thus identified.) (HDR, or HDR UK: Health Data Research UK.)

| Domain | Reviewer's comments |
|---|---|
| **Elements worth including** | Establish a Data Access Committee (DAC). A data access committee (DAC) is a "committee or an equivalent body that is involved in assessing access requests and overseeing the management and administration of data access." Note that all survey respondees indicated that they already had a DAC or were planning to establish one. This may indicate that there is substantial support for this guideline among health data hosting organisations, although note that the number of respondees is relatively small.<br>• DAC should be independent;<br>• DAC should have "oversight of the entire data lifecycle—collection, storage, access, and use."<br>• There should be patient and public representation on all DACs, making up at least 25% of the DAC members. The number of patient/public members should be proportionate and reasonable considering organisation size and volume of data access requests.<br>• DAC chairs should support meaningful patient/public involvement. This goes beyond a requirement for patient and public involvement to advocating for meaningful involvement and explicit support from the DAC chair.<br>• DAC members should be from diverse backgrounds and recruitment must ensure this.<br>• Training for all members.<br>• Reimbursement/honorarium in line with NIHR guidance.<br><br>Data access request and approval procedures:<br>• Patients and the public should be involved in co-developing data access request and approval procedures. Such procedures are referred to as only "data access approval procedures" but the end-to-end procedure would include the data access request as well; i.e. an application form would be submitted; there would presumably be various requirements/stipulations for the application form.<br>• Procedural documents should be written in plain and accessible language, with clear explanations.<br>• Procedural documents should be publicly available (published online) and these should be easily findable.<br>• Publicly available information should include clear rationale for (DAC) processes. The rationale for processes is not usually required in guidelines, but the reviewer considers that this could be useful for various reasons (not specified in reviewed document): (i) greater transparency; (ii) improve public understanding; (iii) encourage more deliberation on why something is being or is not being done; (iv) may increase procedural adherence as staff may be more likely to follow a process if they understand the reasons for it; (v) may be helpful when procedures are reviewed; (vi) may be helpful for documenting and understanding any changes to procedures.<br><br>Reviewing data access requests:<br>• "Criteria need to be clear and understood by all committee members to ensure consistent application." |

| | |
|---|---|
| | ◦ The criteria for assessing data access requests needs to be documented in plain language and available for all committee members;<br>◦ Appropriate training for all members.<br>• "Disputed decisions" should be "clearly discussed and recorded."<br>• "[The] 'Five Safes' model should be adopted" for assessing requests.<br><br>Transparency and public scrutiny:<br>• All data hosting organisations to maintain a public registry of approved data requests which is made publicly available online. This would be appropriate for the sharing of unconsented MH data.<br>◦ Public registry should be in accessible format;<br>◦ Should include conditions imposed by the DAC;<br>◦ Should include information on how data access will be audited;<br>◦ Should include information on how data access will be monitored.<br>• Data custodian/hosting organisation should include its conditions of data use (which are applicable to all organisations) on its website.<br>◦ Should be in an accessible format;<br>◦ Should include details of what actions will be taken if conditions are breached.<br><br>HDR UK recommendation: "Establish an Alliance forum to encourage shared learning across organisations, drawing on the views and experiences of patient/public members involved in existing approval processes, with a view to securing meaningful public involvement in all stages of the data lifecycle."<br>• While there would inevitably not be a need for a multitude of such forums, it may be beneficial to establish one focused exclusively on MH data. |
| **Elements worth including with amendments** | Data access request and approval procedures: The recommendation on establishing a DAC also includes specific recommendations on the data access processes and other procedures followed. Such procedural recommendations should be specified separately (i.e. distinct from the guideline to establish a DAC). However, the various individual recommendations with regards to the data access request and approval procedures should be included in the MH data sharing guidelines.<br><br>Reviewing data access requests: The recommendations state that it "may not be necessary for every request to be reviewed by patient/public representatives" and that there should be some prioritisation and consideration of context if there is a high volume of requests. Arguably, with regards to unconsented MH data, all data requests should be reviewed by patient/public reviewers. Note that only 9 survey respondents (out of 22) indicated public involvement in reviewing data access requests, meaning that this is a practice that needs to be encouraged across health data hosting organisations that share data.<br>• Arguably, if there is a high volume of requests, there should be a correspondingly high number of patient/public representatives.<br>• Assuming that a high volume of requests would mean more successful applications, which would mean more funding for the hosting organisation, more reviews by more patient/public representatives could be funded.<br>• Deciding that certain data requests do not have to be reviewed by patient/public reviewers is problematic and raises concerns about how this would be decided. The recommendations do state that the method for determining whether a data access request should be subject to patient/public review "should be subject to discussion, active agreement, and review with patient/public members." This may work if there is transparency about how this determination is made and if patient/public members can |

regularly review this. If it is decided that certain data access requests will not be reviewed by patient/public reviewers, it should be publicly indicated (online) which types/categories of applications are not subject to patient/public review and this should be clear on the public registry of requests.

- The recommendations refer to the identification of "precedents that make detailed assessment unnecessary" which is concerning because organisations should not be able to bypass the assessment process. There may indeed be valid precedents in rare cases but since no clarification or examples are given in the HDR recommendations; there is a risk that a recommendation such as this can be open to interpretation, leading to bad practices.
- If it is decided that not all requests for MH data have to be reviewed by PPI representatives, there is a risk that over time, a greater proportion of applications may be regarded as not requiring patient/public review.
- When it is decided that certain access requests do not need to be subject to patient/public review, this can result in certain types of application being wrongly put into this category; e.g. […] that funding applications that are deemed to be too technical (e.g. concerning artificial intelligence [AI]), that do not have an (adequate) plain-language summary, or that have not planned for PPI, are deemed to not require public review. These are exactly the applications that do require public review.
  - The use of AI, including in medical treatment, is a contentious issue. While there are many potential benefits, there are also risks and concerns, including the risk of bias. Thus, there should be transparency around health research projects that use or develop AI and that require access to unconsented MH data; such data access requests should be subject to patient/public review. The tendency of AI health research to not plan for PPI is also concerning and something that can be raised via patient/public review of data access requests.

One HDR recommendation states that "Patient/public representatives should always be involved in data access requests that involve personal and/or sensitive data." Arguably, for unconsented MH data, identifiable data should not be shared with commercial organisations. If/when data is shared, only anonymous data should be shared. All MH data can be classified as "sensitive" and thus there should be patient/public review of all MH data requests.

Access to data (how/where data accessed):

- Data "access should take place only within a Trusted Research Environment" or similarly safe environment.
  - This would be appropriate for unconsented MH data, which by nature is sensitive.
  - Note, however, that the HDR recommendation states that data access should take place within a TRE "wherever possible". This wording is problematic and essentially permits organisations for whom it is not possible to use a TRE (for whatever reason) to use other unsecure means of data access, including direct transfers of data.
  - It would be appropriate to word this more strongly for the MH data guidelines, as arguably a MH data hosting organisation should have devised secure ways of sharing this data before considering any data access requests and there should not really be situations in which such sensitive and unconsented data is accessed outside of secure environments.
  - For unconsented MH data, it would be appropriate to include a guideline prohibiting the direct transfer of MH data to organisations. It may also be an idea to consider what other insecure and problematic data access methods are in use and explicitly prohibit them or at the very least list them as examples of bad practice.

Transparency and public scrutiny: The HDR recommendations state that all data hosting organisations should maintain a public registry of approved data requests which is made publicly available online.

- Arguably, it would be more transparent to include a public registry of all requests, including those that have been rejected (and the reasons for this). This would make for a more trustworthy system and allow greater public scrutiny. The HDR paper includes examples of practices by surveyed organisations, including one organisation that has a public registry of all requests "including those that have been declined and the committee's reasons for refusing access."
    - Perhaps the guideline in relation to a public registry could specify different options which are ranked? For example: minimum implementation (public registry of approved data requests) to excellent implementation (public registry of all requests, including those that have been rejected and the reasons for rejection, that is accessible (e.g. in clear plain language), contains all necessary information and is easily searchable).
    - This kind of grading system might actually inspire organisations to strive for better practices.
    - It may re-frame how the implementation of guidelines is viewed, e.g. the minimum implementation may be viewed as the default rather than "no implementation" of a guideline.
- The HDR recommendations specify that the public registry should include details on conditions imposed by the DAC and information on how data access/use will be audited and monitored. Note that the HDR recommendations do not stipulate that data access should be audited or monitored, only that the details of what auditing and monitoring will be done be specified on the registry. For unconsented MH data, it would be important for data access and use to be audited and monitored, and there should be a specific guideline stating this.
- Additional information that should be included in the public registry, in clear plain language, includes:
    - Whether the access request was reviewed by patient/public reviewers, and if not, why not;
    - Whether approved data access requests were supported by patient/public reviewers, and if not, why not;
    - How data is accessed, information on how this is secure;
    - Duration of data access;
    - Details of the organisation requesting data access;
    - Details of the research project for which data access requested;
    - Details of what data was shared—how much data was shared, what was included in the data, individual or aggregate, identifiability of data, structured data or free text;
        - Note, however, that for unconsented MH data, it would not be appropriate to share identifiable data under any circumstances and it would not be appropriate to share free text as this cannot (currently) be anonymous and may contain highly sensitive and confidential information.
    - Details of any reviews/audits/monitoring of data use and compliance with conditions of use/access;
    - Details of any breaches in conditions (including any data misuse) and actions taken;
    - Details of any security and privacy breaches and actions taken;
    - Information on research outputs.

**Noted to avoid** The recommendations refer to the identification of "precedents that make detailed assessment unnecessary" which is concerning because organisations should not be able to bypass the assessment process. There may indeed be valid precedents in rare cases but since no clarification or examples are given in the HDR recommendations, there is a risk that a recommendation such as this can be open to interpretation, leading to bad practices.

In general, avoid guidelines or wording within guidelines that permit organisations to circumvent good practices in certain situations without clearly defining those situations or circumstances. In the HDR recommendations this occurs in the above-mentioned case (regarding precedents that make assessments unnecessary) and also when hosting organisations are advised to use TREs where this is possible.

Reference to the sharing of personal (i.e. identifiable) data. For unconsented MH data, identifiable data should never be shared with commercial organisations. If/when data is shared, only anonymous data should be shared.

**Gaps**

No reference to information that commercial organisations should provide in data access requests.

No reference to purpose for which data can be shared. No reference to patient/public benefit and impact.

No reference to the type of data available for sharing.

No reference to the degree of identifiability of the data. There is a passing mention to de-identified data in the introduction, but this is contradicted by a recommendation that there always be patient/public reviewers of data access requests which "involve personal and/or sensitive data."

Very limited coverage of the technical environment and how data is accessed/methods of data access—states that data access should be via TRE where possible. Safe environments and secure methods of access need to be clearly defined. Insecure methods should be prohibited.

No reference to the duration of data access and use. Data access and use should not be granted for an indefinite period and both the access request and access approval should explicitly mention the duration. If access is required beyond the approved duration period, then a (formal) request should be made for renewal of access.

Only recommends that information about how audits and monitoring are conducted be specified on the public registry. No requirement for audits and monitoring of data use to be conducted.

There is no reference to patient involvement in monitoring and auditing data use (whether reviewing progress reports for this purpose or conducting audits). It should not be assumed that monitoring and audit activities are a purely technical exercise; they are not.

No reference to any requirement for patient and public involvement at the commercial organisation (from application to project completion).

Other gaps in addition; not all covered.

**Public involvement approach**

The paper has been compiled by HDR UK's Public Advisory Board (PAB), following review of the survey findings. The PAB summarise key findings and offer recommendations.

Recommendations covering PPI and consideration of the wider public:

- There should be patient and public representation on all DACs.
- DAC chairs should support meaningful patient/public involvement.
- Training for all DAC members;
- Reimbursement/honorarium in line with NIHR guidance.

- The criteria for assessing data access requests should be clear and documented in plain language.
- Procedural documents should be written in plain and accessible language, with clear explanations.
- Procedural documents should be publicly available (published online) and these should be easily findable.
- Patients and the public should be involved in co-developing data access request and approval procedures.

The recommendations state that it "may not be necessary for every request to be reviewed by patient/public representatives" which may not be appropriate in the case of unconsented MH data.

There is no stipulation for PPI in the (corporate) organisation that is requesting access to the data.

**Industry involvement approach**

There do not appear to be Alliance members that are commercial organisations. Although there are Alliance members that are trade associations, there is no indication in this paper that trade associations were surveyed. There is also no indication that there was any industry involvement in developing the recommendations.

The survey and the resulting paper of recommendations is relatively broad in focus, covering the sharing of health data in general, with any organisation (not only commercial organisations) for planning or research purposes. Hence, specific risks presented by industry access to data have not been considered.

Specific public concerns about sharing healthcare data with industry have not been considered.

The paper does not specifically focus on the sharing of healthcare data with commercial organisations.

Specific industry sectors have not been considered.

No recommendations stipulating requirements for industry applicants (e.g. what information to provide on application form, requirement to provide plain language information).

**Equality, diversity, and inclusion considerations**

No information provided on the diversity of PAB members.

There is a recommendation for the recruitment of diverse DAC members, but this does not specifically say that the patient/public representatives on the DAC should be from diverse groups.

Recommendation that all data access procedures are written in accessible language.

Recommendation that all publicly available information is accessible.

**Other considerations**

–

**Summary of strengths and limitations**

Strengths:

- Survey based on what hosting organisations already doing or planning and revealed some examples of good practices. This provides insight on the potential acceptability of some

guidelines.
- Recommendations proposed by members of the public.

Limitations:
- Representatives from only 20 organisations responded, which limits the ability to generalise from the findings.
- The survey and the resulting paper of recommendations is relatively broad in focus, covering the sharing of health data in general, with any organisation (not only commercial organisations) for planning or research purposes. Hence, the specific sensitivity of MH data is not considered.
- Focused on public involvement in DAC, review of data requests and monitoring. Many aspects relevant to data sharing not covered.
- Lack of detail.

#### 3.7 Supplementary Table 7: LEAG member's review of Cancer Research Horizons (2022)

Direct quotes are from the material reviewed. (Not all direct quotes from the primary source document are thus identified.)

| Domain | Reviewer's comments |
|---|---|
| **Elements worth including** | Glossary: Include a clear glossary (1–4 pages) with the guidelines. Several terms can have multiple meanings and it is important that the specific intended meaning for certain terms (in the context of the guidelines) is conveyed within the guidelines document. |
| | Glossary definitions are in "lay" (plain) language and clear. |
| | Industry commissioned research through a university or academic research conducted in collaboration with a commercial organisation is distinguished from other commercial research. The former is defined as academic research if the commercial entity has no commercial rights to the research results. It is further clarified that Commercial Data Partnerships do not include academic research. There should be clarity in guidelines on whether academic research commissioned by industry and/or research conducted in collaboration with a commercial organisation would be classed as commercial research. |
| | People affected by cancer were involved in developing CRH's guidelines. For MH data guidelines, people affected by mental illness to be involved. |
| | Commit to regularly reviewing the guidelines with patients affected by mental illness and update the guidelines, as appropriate. |
| | Industry involvement in developing the guidelines. |
| | There is clarity on the kind of data that would potentially be included in Commercial Data Partnerships, and the kind of data that would not be included: *"CRUK are a research funding organisation, not a hospital group and therefore CRH does not have access to Raw Clinical Data outside of the context of providing clinical context or annotation to data generated in research. Therefore, the Commercial Data Partnerships referred to in this document do not generally include the transfer of or access to Raw Clinical Data independent of research data."* Since the MH data referenced/indexed by the DATAMIND catalogue would potentially be more diverse (e.g. routinely collected data, trials data) and given that previous studies have shown that people are more averse to sharing certain types of MH data (e.g. free text versus structured data), there should be clarity on exactly what type of MH data can be shared with commercial organisations. |
| | Different (scoping) statements to address the different concerns and needs of different groups (research partners, commercial entities, public and patients). |
| | Guideline 1.1: CR-UK (CRH) will only enter Commercial Data Partnerships for specific purposes, and they define this purpose as specifically including *"the prevention, diagnosis, treatment or monitoring of cancer and related diseases."* This aids transparency, would aid in determining what data access requests to approve and would help ensure that patient data is used for a purpose that various interested parties would be okay/happy with. So far, while recognising that it is still early stages, in DATAMIND, there has been talk of patient MH data being used for public benefit or for the public good, without defining what that is in any way. |
| | Guidelines 1.2 and 1.3: Each data access request must include a plain-language summary, which |

| | |
|---|---|
| | contains the aims and objectives of the project. Provide details on the intended use of the patient data.<br><br>Guideline 2.1 states that access will only be allowed to pseudonymised or anonymised data. This would be appropriate for MH data given the sensitivity of such data. Access to identifiable MH data or "personal data" should not be granted.<br><br>Guidelines 3.1 and 3.3: *"The originating research institutes, researchers and/or NHS Trusts and their collaborators shall, at all times, retain the rights to continue to use the Patient Derived Data in Academic Research, including sharing with academic collaborators and for the delivery of patient care and can continue to publish the outputs of their research"* and *"The ownership of the IP rights in the Patient Derived Data will remain with CRH and/or the originating institution(s) as per established technology transfer agreements."*<br>• Should be included.<br>• DATAMIND will index various data sets including routinely collected data and data from research studies. It would be appropriate to explicitly state that any commercial organisation that is granted access to any data is not acquiring any intellectual property (IP) rights to the data, nor will they gain any other exclusive rights with regards to access or use of the data.<br><br>Guideline 3.5 states that publication of any outputs of the use of the patient data should recognise the contribution of the relevant organisations and originating researchers (where applicable).<br><br>Guideline 3.6: The commercial organisation is prohibited from publishing any identifiable/personal patient data.<br>• In the case of MH data, identifiable/personal data should not be shared with commercial entities anyway, but this guideline should still be included in case any anonymised or pseudonymised data contains any identifiable patient information whatsoever.<br>• There should be additional technical controls in place to ensure that any data outputs from the Trusted Research Environments (TREs) are suitably anonymised to limit reliance on the commercial entity and to increase trustworthiness within the system.<br><br>Guideline 4.1 refers to ensuring that partnerships with commercial organisations are "mutually beneficial" and financially fair. It refers to mechanisms such as cost sharing, cost recovery, access fees, annual fees, annual fees, revenue shares, milestones, and royalties. These should be considered with regards to the sharing of MH data. As with CRH, for any revenue generated from MH data partnerships, this should be shared with the research and/or NHS organisations who (co-)own the data.<br><br>Guideline 4.2 refers to tailoring financials for each commercial partnership, which may also be appropriate for MH data. However, to ensure fairness and transparency, there should be pricing models/lists, and any financial arrangements should be open to public scrutiny.<br><br>CRH categorises its guidelines into four categories. The MH data guidelines should be categorised into distinct sections. More thought needed on appropriate categories for MH data guidelines. |
| **Elements worth including with amendments** | Co-develop the guidelines with people affected by MH (which we are hopefully going to do) rather than in consultation with people affected by MH.<br><br>Commit to reviewing the guidelines with patients affected by MH every six months or annually (at least) and update the guidelines, as appropriate. Would suggest 6-monthly to annual reviews, due to the high sensitivity of MH data and the rapidly accelerating speed of developments in data science and artificial intelligence. Additionally, the frequency of the reviews themselves, should be up for review, so that this can change, as necessary. |

It appears that industry consultations and patient consultations were separate, and that there were no joint collaborative consultations. There would be benefits in involving all parties (industry, patients, CR-UK, academics) in joint collaborations, in addition to separate consultations.

CRH is open to sharing all data it has the rights to share, with commercial organisations. While this may be appropriate for CRH, given the nature of the data (i.e. primarily consented data from cancer trials) it may not be the appropriate approach for sharing MH data with commercial entities (for which there was no explicit consent to share). There needs to be clarity around and categorisation of the different types of MH data available, which should inevitably be co-produced with patients, including consideration of the specific concerns that patients and the public have around the sharing of this data. One concern may be that free-text MH data may never be fully anonymised. Subsequently, there needs to be clarity around what types of MH data can be available to share with commercial entities—and this does not have to include all the available MH data. Currently, it appears that considerations take an "all or nothing" approach and if MH data is going to be shared, it appears that all of it is available to be shared by default.

Includes different (scoping) statements for different interested parties. Given the findings of prior studies, specifically that minority groups are more averse to sharing MH data, it may be appropriate to include a further statement to address concerns expressed specifically by this group.

Guideline 1.1: *"CRH will only enter into a Commercial Data Partnership and enable access to Patient Derived Data for a defined purpose, which must include the prevention, diagnosis, treatment or monitoring of cancer and related diseases."*

- Implies that the purpose may include other things if this essential criterion is met.
- However, it then states that any use of patient data "beyond this approved purpose shall be explicitly prohibited." Hence, this is ambiguous and open to interpretation. This guideline could be worded better, although the essence of what it includes should be included in the guidelines produced by DATAMIND.
- Unless there is a clearly defined purpose for which MH data can be used by commercial entities, transparency and accountability cannot be ensured. A clearly defined purpose would also aid in determining what data access requests to approve and would help ensure that patient data is used for a purpose that various interested parties would be okay with.
- Of course, when a specific purpose is defined for data use, there will be requests for which there is some uncertainty or ambiguity about whether they fall within scope. (In the case of CRH's defined purpose for Commercial Data Partnerships, what about: [i] research which seeks to rule out cases of cancer rather than diagnose cancer? [ii] research which seeks to better understand illnesses which arise in patients post-cancer treatment? [iii] research on cancer processes and pathways that patients follow including how treatment decisions are made?) In some cases, the data request may clearly be out of scope, and yet may present potential benefits that were not previously considered.
- It may be appropriate to be clear on the defined purpose here, and perhaps include an option later (specific guideline) for considering some requests that may fall outside of the specific defined purpose. There should be a formal process for considering requests that fall outside of the defined purpose.
- If another guideline will be included to consider requests that fall outside of the defined purpose, it may not be appropriate to include an explicit statement here prohibiting the use of data beyond the approved (defined) purpose. Perhaps it would be better to include instead a separate guideline prohibiting the use of data beyond the purpose that is approved for each individual access request.
- The "defined purpose" can be reviewed when the guidelines are regularly reviewed.

Guideline 1.2: *"CRUK is funded by public donations and therefore public transparency is integral to*

*CRH's activity. As a result, CRH will require a lay summary to be made available for public scrutiny, consisting of non-confidential information relating to the aims and objectives of each Commercial Data Partnership we enter into."*

- Could be worded better to clearly clarify whether the lay (plain language) summaries will be reviewed by patients and the public when reviewing the access request, or whether they will also be made publicly available for public scrutiny (i.e. not just by those involved in PPI).
- Could clearly indicate what a lay/plain-language summary is, and whether a request will be considered without an adequate plain-language summary. The absence of a sufficiently good plain-language summary can hinder the lived experience reviewer from determining the public and patient benefits of the research. It is not the job of the lived experience reviewer to critique the plain-language summary and to suggest alternative simpler language, but rather to use the plain-language summary to determine whether the access request should be supported. Also, a plain-language summary should not be an abstraction of the scientific description with all detail removed, resulting in inadequate information to make a decision. Instead, the plain-language summary needs to include all relevant information in clear language.
- Given the sensitivity of MH data, and the fact that such data is largely unconsented for sharing, maintaining a public registry of all requests in plain language, regardless of whether such requests are approved or not, would greatly aid transparency and perhaps help build trust. To be clear, it should be the responsibility of commercial entities to provide the data access request in plain language that is clear and sufficiently detailed (before any request can be considered), and this request could be made publicly available on the public registry. The registry could include details on the requesting entity, summary of the proposed project, plain-language summary, and outcome of the request (approved/declined). If requests are likely to contain information which may compromise competitiveness (which may well be the case) then there could be a (specified and suitable) delay between receiving the request and the information being made publicly available on the registry. However, the aim would be to publicly log all requests, regardless of whether they are approved. Given all these additions, 1–2 additional guidelines may be needed.

Guideline 1.3: *"CRH will involve patients in the decision-making process when entering into Commercial Data Partnerships and prospective partners shall provide sufficient information to allow a comprehensive review to be performed, including a detailed overview of the intended use of the data."*

- This could be clearer in specifying that the "sufficient information" provided should be in plain language. Patients and the public are hindered from being meaningfully involved in decision-making if detailed and sufficient information is not provided in plain language. It is not enough to simply state that patients will be involved in decision-making if this is not properly enabled, including through the requirements imposed on commercial organisations. Otherwise, the involvement of patients and public is just a tick-boxing exercise as opposed to meaningful involvement.
- Not sufficiently clear on how patients will be involved in decision-making.
    - How many patients will be involved, what proportion of the decision-making panel will be patients (and the public) and what proportion will be researchers/clinicians?
- Given the sensitivity of MH data, this should be clearer for MH data guidelines, and patients should be equally involved in making decisions on access requests.

Guideline 3.2: *"CRH will consult the lead PI who generated the data in the decision-making process when entering into Commercial Data Partnerships and prospective commercial partners shall provide sufficient information to allow a comprehensive review to be performed by CRH, including a detailed overview of the intended use of the Patient Derived Data."*

- The essence of guidelines 1.2, 1.3 and 3.2 should be included in DATAMIND's guidelines, i.e. that patients and the PI (or data controller, as appropriate) should be meaningfully involved in decision-making regarding the data access request and that the commercial applicant is required to provide sufficient information, including in plain language, on the aims and objectives of the proposed project and details on the intended use of the data.

- However, CRH's guidelines do not explicitly state that such information would be required as part of the data access application, and in fact guideline 1.2 implies that such information would only be required in plain language once the partnership has been entered into (i.e. once data access has been granted).
- These guidelines could be improved by explicitly adding that unless plain-language summaries are provided that are clear, sufficiently detailed and that contain the aims and objectives of the proposed project and details of the intended use of the data, the request will not be considered. This would emphasise how important public transparency and public involvement is.

Guideline 1.4: *"Upon completion of a Commercial Data Partnership involving the direct transfer of Patient Derived Data… If the commercial partner wishes to conduct further research using the dataset it must request renewed access to the Patient Derived Data for the new purpose."*

- Guideline 1.4 says that in cases involving the direct transfer of data, the commercial partner should request renewed access if it wants to conduct further research using the data. With MH data, there should not be any direct transfer of data and all such data should be accessed in a safe environment such as a TRE. However, even when data is accessed within a TRE, that data should only be used for the purpose for which access has been granted, and if the commercial entity wants to conduct further research beyond the approved purpose, access should be requested formally again.
- This aspect of the guideline should be amended so that it applies to data accessed within "safe" environments such as TREs (rather than directly transferred data).

Guideline 1.5 states that the commercial entity must provide project progress updates, including progress and outcome reports to enable CRH to *"manage and inform its internal business processes."*

- There should be a requirement to provide progress updates, and this can be one way to help ensure that the data is used for the approved purpose. This will help ensure greater transparency and accountability. This may also enable the system to be more trustworthy and provide reassurance to the public that there are formal mechanisms to check that the data is being used for the approved purpose.
- Frequency of progress updates needs to be mutually agreed during the application process (as this may vary by project) and this should be mentioned in the guideline.
- There will need to be appropriate allocation of resources to ensure that progress reports can be properly reviewed and actions taken, as appropriate.
- Will need an additional guideline concerning the review of progress reports.
- Although this guideline can potentially promote greater transparency and accountability, CR-UK's motivations and explicitly stated reasons for including it are problematic and may lead to problems with compliance—why should a third-party commercial organisation be concerned with CR-UK's internal business processes?
- While progress reports may be useful for managing and informing the data controller's internal business processes, this should not be referenced in the guideline, or this should not be the only explicitly stated reason for the guideline.

Guideline 1.6: *"CRH will share non-confidential, lay summaries of progress/outcome reports with patient involvement groups to inform further partnering activity and the evolution of policies and principles in line with our public & patient involvement process."*

- The sharing of plain-language summaries of project progress/outcome reports with PPI groups would ensure greater public accountability and transparency. Patient and public review of plain-language summaries of progress/outcome reports can be used to ensure that the data access and use is in line with approved purposes.
- More information is needed on whether this is a formal PPI review panel and the frequency of reviews.

- The guideline does not impose a requirement on the commercial organisation to produce these plain-language summaries. Producing such plain-language summaries would take work and require sufficient funding, and so from a financial sustainability perspective, it would be appropriate to place this requirement on the commercial entities rather than the data controller. Furthermore, the commercial organisations would be better placed to explain their projects in plain language.
- Although this guideline can potentially promote greater transparency and accountability, CRH's motivations and explicitly stated reasons for including it are problematic and may lead to problems with compliance—why would a third-party commercial organisation be concerned with CRH's "further partnering activity" or the evolution of CRH's "policies and principles"? If reasons such as this need to be declared, there needs to be due consideration on where such reasons need to be declared (e.g. would this be more appropriate in an access agreement?) and how the stated reasons can impact compliance.

Guideline 1.7: *"The commercial partner's use of the Patient Derived Data shall be transparent and auditable, and CRH will seek to obtain the right to audit to ensure that the principles of the partnership and this document are being maintained."*

- Rather than placing the requirement to be "transparent and auditable" on the commercial entity, it would be appropriate to ensure this through technical controls, i.e. ensure that all data use is auditable within the TRE.
- If direct data transfers are prohibited (as they should be) and all access is via a "safe" environment such as a TRE, then audit controls could be built into the environment. This would ensure greater transparency and help ensure that misuse is less likely.
- The right to audit data access, use and outputs should be part of any robust contract with all commercial organisations, and this should be stipulated in the guidelines. Hence, there should not be need for the data controller to "seek the right to audit."
- The frequency of audit is not mentioned and there is no mention of PPI in the audit. At least one audit should be conducted for each "partnership" and patients and the public should be on the audit team. It should not be assumed that this is a purely technical activity that does not require PPI. PPI on such audits would ensure that: (i) the aim of each audit is relevant and meaningful, taking consideration of patient and public concerns; (ii) what is audited is in line with the aim of the audit; (iii) how the audit is performed and the methods used are appropriate; (iv) there is public scrutiny of the results of the audit; (v) appropriate actions are taken after the audit.
- With regards to audit activities, is this something that an independent organisation could offer to ensure this is consistent nationally? Note that any audit would still be performed in line with local rules, i.e. what is allowed locally. Additional independent audit would offer more reassurance, making the system more trustworthy. This may help identify patterns in issues and the concentration of issues with regards to commercial use of MH data (e.g. are there particular types of issues that re-occur? Are there issues in particular organisations, industries, locations? Are there increased issues for commercial partners of particular data controllers?). Another suggestion—annual ratings for data controllers/owners based on audit performance of commercial partners, as a way to encourage better practices and to stop the sharing of unconsented MH patient data where audit performance is particularly poor?

Guideline 2.1 states that CR-UK will ensure that commercial partners have *"adhered to all relevant regulations and ethical approvals"* before access is granted. Beyond this, other vetting of commercial organisations would be advisable. More work is needed on what form this should take, but there should be stringent vetting procedural controls, a formal (documented) process for this, and patients should be involved in all vetting. This is important given the sensitivity of MH data and the fact that sharing of this data is unconsented. Vetting may include who/what the organisation is, what the organisation does, is the organisation who they say they are, what their primary activity is, what other activities they do, whether they engage in any controversial activities, whether they engage in any controversial sharing or uses of data. Many organisations who request data access may be smaller organisations, start-ups or organisations that are not as well known.

Even if a TRE is used, there would be data outputs and information can be committed to memory and reproduced. Hence, even if there is no direct transfer of patient (identifiable) data, the commercial organisation should be required to "adhere to all relevant regulations including UK GDPR and the Data Protection Act 2018" and this should be explicitly mentioned in the guidelines.

Guideline 2.4 states that commercial organisations must limit access to the data to employees who reasonably need access for the project.

- It should not be left entirely up to commercial organisations to determine who has access and how many employees have access to the data.
- In the case of (unconsented) MH data, it would be appropriate to require commercial organisations to specify individual employees who will require access in the access request, indicating why they would require access. Access could then only be granted for those employees who are deemed to reasonably require access (e.g. separate login on TRE for each approved employee), and this would enable better auditing of data use. Given the highly sensitive nature and unconsented status of the data, there appears to be no good reason why hundreds or even tens of employees at a single commercial organisation should have access to the data.
- Commercial organisations would still be required to ensure that only approved employees use the login credentials (although note that if all data use is logged and subject to audit and monitoring, employees may be less likely to allow other employees to use their logins). Commercial organisations should also be required to request removal of access promptly for employees who leave or change roles.
- If access is needed for additional employees, this should be requested through a separate (formal) process (which would not be as lengthy as a full access request process as data access for the project would already have been granted).

Guideline 2.5 states that the commercial organisation must maintain security of the data, notify CR-UK of any security breaches, and take steps to remedy such breaches.

- Use of TREs would reduce requirements around maintaining security, as commercial organisations would not be responsible for the storage and management of the data, but it would not eliminate such requirements.
- The commercial organisation would have to ensure that only authorised personnel gain access to the TRE (and hence the data).
- Even with the use of TREs, there are always security risks, and hence it would be appropriate to include a guideline requiring the commercial organisation to promptly notify the data controller of any security breach and take all and any reasonable steps to remedy such breaches.
- May be appropriate to specify the time-frame within which the commercial entity should notify the data controller after becoming aware of a security breach.

Guideline 2.6: *"CRH will require the commercial partner to make no efforts to identify individuals that are the subject of the Patient Derived Data."*

- Should be worded more strongly e.g. employees at the commercial organisation are strictly prohibited from seeking to identify/re-identify any patient from the data, and any attempt to do so would constitute a breach of the commercial organisation's contract with the data controller, as well as violation of laws and regulations.
- In the context of CRH's guidelines, this may be redundant at times because CRH also shares identifiable/personal data (which should not be shared for MH data).

Guideline 2.7: *"CRH will require the commercial partner not to further sub-license the Patient Derived*

*Data to a 3rd party without CRH's consent."*

- This should be worded more strongly. Something along the lines of: the commercial organisation is strictly prohibited from sub-licensing or sharing the data with any third-party, and any attempt to do so would constitute a breach of the commercial organisation's contract/agreement with the data controller as well as violation of laws and regulations.
- All sub-licensing of unconsented MH data should be prohibited, regardless of whether consent is sought beforehand.
- All organisations that will be working on the project and potentially accessing the data should be properly declared on the data access application form (as co-applicants, if applicable) with relevant and sufficient details provided. (Sometimes a project may involve multiple organisations working together, and/or an organisation that is primarily responsible for the research project may contract out certain parts of the work to other organisations.) This would allow all such organisations to be properly vetted. Essentially, all third-parties should be subject to the formal data access application and approval process, including review by patients and the public.
- If data access has already been granted for a particular project, and a third-party commercial entity requires access to the same data for the same project, there should be a separate appropriate approvals process for this, which includes PPI.
- There should be technical controls embedded in the safe environment to prevent or significantly minimise the risk of sub-licensing of MH data. There should also be "detect" controls in the safe environment to enable detection of any sub-licensing which has occurred (as "prevent" controls cannot account for all risks and preventative controls can be circumvented).

Guideline 3.2 states that CR-UK will consult the lead principal investigator (PI) (who generated the data) when considering a commercial data access request. DATAMIND will index various data sets including routinely collected data and data from research studies. For the latter, it would be appropriate to involve (rather than consult) the lead PI (and relevant persons in the affiliated organisation) in the decision-making process. For the former, it would be appropriate to involve (rather than consult) the relevant data owner (e.g. NHS England, specific NHS Trust) in the decision-making process. Refer to further improvements to guideline 3.2 above (with guidelines 1.2 and 1.3).

Guideline 3.5: *"Publication of any outputs… should correctly recognise the contribution of CRH, CRUK, involved institutions and the originating researchers."* Should also recognise the contribution of the patients, on whose data, the work is based.

Guideline 3.6: *"For Anonymised Patient Derived Data, the commercial partner shall, at no times, publish or make available on an unsecured source/server or public repository, the licensed data, without approval from CRH and the originating researchers and only if accompanied by an appropriate data access statement, process or agreement. The commercial partner will not publish Patient Derived Data that constitutes Personal Data."*

- In the case of MH data, there should not be any direct data transfers, and all approved data access should be via a TRE. (If there are no direct data transfers, the commercial organisation would not be able to make such MH data available on an unsecured server or public repository.)
- Even if a TRE is used, there would still be data outputs and information can be committed to memory and reproduced. So, an amended version of this guideline could be included to ensure that no data accessed is made available on an unsecured server or public repository.
- There should be additional technical controls in place to ensure that any data outputs from the TREs are suitably anonymised. However, the guideline should still prohibit the publication of all data unless it (a) is anonymised to a publishable standard, and (b) has had prior review and approval by the data owner/controller.

Guideline 4.5 states that if the commercial entity *"generates enriching data that improves the utility of the Patient Derived Data and has broad applicability, CRH may request that such additional enriching data is provided back to CRH so that it can be made available for other academic and commercial users."*

- This may be considered unfair if the commercial entity paid for data access.
- However, if all work on the data is done within a TRE, wouldn't all enriching data generated reside within the TRE? And if this is the case, would such enriching data be owned by the originating data owner?

**Noted to avoid** Explicit statements or even implications that commercial organisations are key to unlocking value in patient healthcare data: *"We believe that, due to their unique skill set, resources and experience taking products to market, collaborating with commercial organisations is key to unlocking the impact of this Patient Derived Data, and CRH want to enable this through establishing Commercial Data Partnerships."*

- This undermines the work of other types of organisations (Universities, charities, public sector organisations, social enterprises) when working with patient health data, depicts commercial organisations as solely beneficial actors, and shows too much "keenness" to sell patient data to commercial organisations.
- Commercial organisations are not a homogeneous group, and there are both benefits and disadvantages of collaborations with such organisations, which will vary depending on the specific organisation and the specific project.
- Context matters: Statements such as this may not be hugely problematic within CRH Data Partnership guidelines, since the data in question would mostly be collected via cancer research trials, and the patients would have explicitly consented to sharing their data, but such statements would absolutely not be appropriate for unconsented MH data, given that prior research has already found that MH data is regarded as more sensitive, and that patients are more adverse to sharing such data with industry. Consider how such statements would come across to patients and the public.

The glossary definition for pseudonymised hides the risks of re-identification, which may especially be a concern for MH data.

It is not clear whether academics were consulted in the development of the guidelines.

CRH is open to sharing all data it has the rights to share, with commercial organisations.

- This may be appropriate for CRH, given the nature of the data (i.e. primarily consented data from cancer trials) but it will not be an appropriate approach for sharing MH data with commercial entities (for which there is no explicit consent to share).
- There needs to be clarity around and categorisation of the different types of MH data available, which should inevitably be co-produced with patients, including consideration of specific concerns that patients and the public have around the sharing of this data.
- Subsequently, there needs to be clarity around what types of MH data can be available to share with commercial entities and this does not have to include all the available MH data.

CRH does not distinguish between the data that it could share with entities from different industry sectors, and seems to regard all of industry as one homogeneous entity. While this may be okay for CRH and cancer trials data, this may not be appropriate for MH data, given views expressed in prior studies, specifically that patients and the public may be more in favour of sharing such data with commercial entities in certain sectors over others. Further in-depth mixed methods research may need to be undertaken on perspectives on sharing MH data with organisations in different industry sectors.

Guideline 1.4 states that in cases involving the direct transfer of data, the commercial partner should delete the data after completion of the partnership. Given the sensitivity of MH data, arguably such data should not be directly transferred to any organisation, and all access should be via a TRE. There should not be reliance on the commercial (or any other) organisation to delete the data after use. It would be appropriate for there to be technical controls in place to ensure that data is accessed in a "safe" environment and that no copies of patient data are taken outside of that environment (except for data that is anonymised and to a standard suitable for publication).

Guideline 1.4 also states that in certain circumstances, the commercial entity *"may request the retention of a single static copy"* of the patient data to comply with laws and regulations.

- Given the sensitivity of MH data and given that the data is unconsented for non-clinical use, unless the data is anonymised to a standard suitable for publication, there should not be retention of patient data in this way by commercial organisations.
- There is an increased risk of re-identification for MH data given the type and amount of information included.
- If laws and regulations require such data to be retained, can this be arranged within the TRE? i.e. for the data analysis to be saved within the TRE and associated with the project in question.

Guideline 1.7 states that the commercial entity's use of the data must be *"transparent and auditable."*

- Direct data transfers should be prohibited and all access should be via a "safe" environment, e.g. TRE.
- There should not be reliance on the commercial entity to ensure that their use of data is transparent and auditable.
- It would be appropriate to ensure that all data use is auditable within the TRE through technical controls, i.e. audit controls could be built into the environment. This would help ensure: (i) greater transparency; (ii) misuse is less likely; (iii) greater trustworthiness within the system; (iv) that audits can be conducted in a consistent way across commercial organisations (as otherwise audits would rely on what is auditable at each commercial entity, which would vary between entities).

Guideline 2.2 states that when data is anonymised (no mention of pseudonymised), CRH will either provide access to the data via direct transfer or via a TRE.

- Unconsented MH data should never be directly transferred to any organisation, including (and perhaps especially) commercial organisations.
- There are degrees of anonymity and identifiability, and even when there is an attempt to anonymise data, there is always a risk that some personal information remains in the data (e.g. on an image, in free text) and even if this is not the case, there is always some risk of re-identification. Arguably for MH data there is an increased risk of identification due to the type of data and diversity and level of detail contained within the data. Hence, it would be inappropriate to directly transfer MH data to commercial organisations, even if that data is apparently "anonymised."
- Direct transfers also present substantially increased risks of data misuse and unauthorised access.
- There would be no guarantee that unauthorised copies of the data have not been made or that the data has been deleted when required (at project completion or due to a contract breach).
- This guideline also states that, for direct data transfers, CR-UK *"will work with the data owner… to ensure that it is shared through a secure medium."* Secure mediums are not explicitly defined, and if this is left to individual staff members (without explicit guidance) insecure mediums can be used, i.e. there are plenty of people, including NHS clinicians, who believe email to be a secure means of communication. Note also that prior research has found that public perceptions and confidence of technological capability within the NHS is low.
- Guideline 2.2 also states that, for direct transfers, the commercial entity must ensure that the

data is "kept in a secure environment." "Secure environment" is not defined and open to interpretation. Reliance should not be placed on commercial organisations to ensure this.

- This guideline also states that the commercial entity should protect the data from damage or loss. There should not be reliance on the commercial entity to ensure the data is not damaged or lost. However, as such, if one copy of such a dataset is damaged or lost, that may not present a huge public concern (as there would presumably be other copies of the same data, i.e. the data would not be lost). There are other serious concerns that are not mentioned in the guideline, nor do there appear to be any explicitly stated measures to reduce these risks: unauthorised access to the data, misuse of the data, unauthorised transfers of the data to other organisations, unauthorised copies being made of the data, unauthorised linking of the data, plus more.
- If all access is via a TRE, then the data can be retained in a secure environment, which would address many issues.

There is a contradiction in the guidelines whereby guideline 2.1 states that CRH would only enable access to pseudonymised or anonymised data, but then guideline 2.3 refers to the sharing of identifiable or personal data (that too via direct transfer). Guideline 2.3: In the case of identifiable or personal data, CRH's "preference" is to enable access via a TRE, but where this "is not possible", direct transfers will be used to share the data if CRH "believe there is the potential for significant patient benefit" from the commercial partnership.

- There should not be direct transfers of identifiable or personal MH data to commercial (or any research) organisations.
- It is not sufficient for there to be potential "patient benefit", and in any case, this "patient benefit" is undefined and open to interpretation.
- In this guideline, again, the medium through which the data will be shared is not defined, and there is the risk that an insecure medium may be used.
- The commercial entity is required "to securely store and manage the data and adhere to all relevant regulations including UK GDPR and the Data Protection Act 2018."
- For sensitive MH data, which is unconsented for the purpose of sharing, it would be appropriate to ensure the secure storage and management of the data via technical controls in the TRE, rather than relying on commercial organisations to do this.

Guideline 2.7: CR-UK permits commercial organisations to sub-license patient data to third-party organisations if CR-UK's consent is sought beforehand.

- This would be completely inappropriate for sensitive and unconsented MH data.
- It would cause the entire vetting and data access process to be by-passed, and instead consent would be sought via an inconsistent, ad hoc, and inadequate process that would not ensure transparency, accountability, security, or the appropriateness and public acceptability of access.
- All organisations that will be working on the project and potentially accessing the data should be properly declared on the data access application form (as co-applicants, if applicable) with relevant and sufficient details provided. (Sometimes a project may involve multiple organisations working together, and/or an organisation that is primarily responsible for the research project may contract out certain parts of the work to other organisations.) This would allow all such organisations to be properly vetted. Essentially, all third-parties should be subject to the formal data access application and approval process, including review by patients and the public.
- If data access has already been granted for a particular project, and the third-party commercial organisation requires access to the same data for the same project, there should be a separate appropriate approvals process for this, which includes PPI.
- The use of TREs would reduce the risk of a commercial partner providing access to the data to other third-party organisations (i.e. the data could not be directly transferred, but login credentials might be shared). There should be technical controls to prevent sub-licensing of MH data and to detect any instances of sub-licensing which have occurred (as "prevent"

controls cannot account for all risks and preventative controls can be circumvented).

Guideline 3.4 implies that for approved commercial data requests (partnerships) CRH would seek to enable, where possible, collaboration with the originating researchers.

- This is applicable to research trials data rather than unconsented data.
- It is not clear what this collaboration would be for and it could be more clearly stated that the collaboration would be on the proposed research project.

Guideline 4.3 states that if there is potential for *"significant patient benefit", "time-limited or field-restricted exclusivity would be considered"* and Guideline 4.4 states that such commercial exclusivity would not apply to raw clinical data and would not prevent academics from continuing to use the data.

- Unsure about this one, but leaning towards avoiding it.
- I would be wary of any MH research that promises (overnight) "significant patient benefit."
- For routinely collected (unconsented) MH data, it would be inappropriate to grant exclusivity to a commercial entity.
- Could be done for some research study data (where appropriate) but any exclusivity granted should be for a limited time and the commercial entity should pay accordingly for the exclusivity.

Categorisation of the guidelines into: (i) Transparency & Accountability; (ii) Data Security & Management; (iii) Protecting Academic Research; (iv) Fair Partnership. These categories do not reflect the main concerns with regards to the sharing of unconsented MH data and as such may not be meaningful for this type of data.

**Gaps**

No reference to the duration of data access and use. Data access and use should not be granted for an indefinite period and both the access request and access approval should explicitly mention the duration. If access is required beyond the approved duration period, then a (formal) request should be made for renewal of access.

There is no reference to patient involvement in monitoring and auditing data use (whether reviewing progress reports for this purpose or conducting audits). It should not be assumed that monitoring and audit activities are a purely technical exercise; they are not.

There is no reference to patient involvement in vetting of commercial organisations beyond reviewing the access request.

No mention of any registries in the guidelines, although CR-UK do appear to maintain some kind of online registry of commercial partners. Beyond a public registry of approved partners (for whom access has been granted), it may be appropriate to maintain a public registry of all access requests in plain language for greater transparency and public scrutiny (with minimal information to address commercial confidentiality and competitiveness concerns).

No mention of situations in which more than one commercial organisation is working on the same project from the onset (i.e. multiple commercial organisations requiring access to same data for same project) or if further commercial organisations become involved in a project later and require access to the data later. The former would require a more comprehensive data access approvals process (with appropriate allocations of time and resources) while the latter would require a separate process dedicated to vetting commercial organisations and considering whether they (and specific employees) require access to the data in question, without any need to consider the project (as this would already have been approved).

| | |
|---|---|
| | There is no mention of what steps will be taken in the event of a breach/violation of the guidelines. How and how soon would all access to the data by the commercial organisations in question be revoked and how would this be ensured?<br><br>Detail on monitoring processes is missing. What this would entail, who would perform monitoring, what access would be required, frequency of monitoring, what would happen with the outputs of monitoring processes, what would happen if guideline violations are detected.<br><br>Detail on auditing processes is missing. What this would entail, who would perform auditing, what access would be required, frequency of auditing, what would happen with the outputs of auditing processes, what would happen if guideline violations are detected.<br><br>No reference to any requirement for patient and public involvement at the commercial organisation (from application to project completion).<br><br>Any and all EDI considerations have been completely omitted.<br><br>Addressing the issues in research culture, e.g. the reproducibility and replicability problems with research. Such problems directly affect patients and the public, and can cause much harm rather than the research being for public benefit. There have been specific issues with commercial organisations misrepresenting the benefits and side effects of [specific psychotropic] medications, which has harmed [many] patients.<br><br>Stipulation to use SDE/TRE for all data access by commercial entities.<br><br>Technical controls are not referenced and there appears to be undue reliance and perhaps even blind faith (?) on the commercial organisation to comply with requirements. While this may not be a concern for the kind of data shared by CRUK (consented cancer trials data), it would not be appropriate for the sharing of unconsented MH data.<br><br>Should be additional technical controls in place to ensure that any data outputs from TREs are suitably anonymised.<br><br>Safe environments and secure methods of access need to be clearly defined.<br><br>Consideration of misuse of data.<br><br>Other gaps in addition; not all covered. |
| **Public involvement approach** | Developed through consultation with people affected by cancer.<br><br>The guidelines will be reviewed in consultation with patients annually (at least) and updated as appropriate.<br><br>Patients and public involved in data strategy, after which the guiding principles were developed.<br><br>The involvement of patients and the public in developing the guidelines has been largely consultative and the guidelines were not co-developed with patients and the public. |

| | |
|---|---|
| | Each data access request must include a plain-language summary, which will be reviewed by patients and the public. Patients will be involved in the decision-making process for considering data access requests. However, there is no indication of what would happen if a plain-language summary is not included or if the plain-language summary is inadequate. Note that if there is no adequate plain-language summary and if the information provided is not accessible, PPI becomes more of a tick-boxing exercise.<br><br>Plain-language summaries of project progress/outcome reports will be shared with PPI groups "to inform further partnering activity" and the review of policies and guidelines. It is unclear what is meant by this and there is no indication that there will be PPI beyond review of data access requests, e.g. no indication of PPI in monitoring.<br><br>Information that the commercial entity is required to produce (plain-language summaries etc.) as part of their application or for monitoring purposes is required to be of a non-confidential nature. |
| **Industry involvement approach** | Guidelines were developed through consultation with the public and private sectors, although it is not clear what sectors of industry were involved. Industry regarded as single homogeneous entity, with no recognition of differences between different industry sectors.<br><br>Industry applicants required to provide a plain-language summary with their access request.<br><br>Industry applicants who have been granted access required to provide progress reports.<br><br>Various requirements around storage, management and deletion of data placed on commercial organisations, which should NOT be necessary if TREs/secure environments used. Over-reliance and undue reliance on commercial organisations to comply with requirements, which would not be appropriate for unconsented sensitive MH data. There should be more technical controls within the environment that enable greater trustworthiness within the system.<br><br>Various requirements on commercial organisations around reporting of data breaches and remedying.<br><br>Requirements on commercial organisations to specify individual employees who will require access and ensure only authorised employees have access<br><br>Requirements on commercial organisations to ensure data use for approved purpose only.<br><br>Measures and rules for commercial organisations regarding publication—anonymisation of data, credits.<br><br>Reference to collaborations with academic researchers.<br><br>Requirements on commercial organisations to submit to audits and monitoring. Requirement to be auditable. This should not be necessary if TREs used and if there are appropriate technical controls within the system.<br><br>Not to undertake any prohibited actions. |
| **Equality, diversity, and inclusion** | Not considered. |

| | |
|---|---|
| **considerations** | |
| **Other considerations** | Considerations for MH data:<br>• For data for which access is not possible via a secure and safe environment (e.g. a TRE), do not include it on the DATAMIND catalogue unless and until this is possible—and include a guideline for data controllers to not make such data available unless and until it can be made available via a secure and safe environment (with appropriate technical controls).<br>• Maintain public registry of all requests in plain language, regardless of whether such requests are approved or not, to aid transparency. If requests are likely to contain information which may compromise competitiveness (which may well be the case) then minimal information could be provided on the public registry and/or there could be a (specified and suitable) delay between receiving the request and the information being made publicly available on the registry. |
| **Summary of strengths and limitations** | Strengths:<br>• Relevant stakeholders, including patients and the public involved in decision-making for access requests and in reviews of guidelines.<br>• Range of issues considered including security, privacy, revenue-generation, data use, PPI, collaborations, third parties, legal issues, audits.<br><br>Limitations:<br>• Does not cover MH data or routinely collected (unconsented) data.<br>• No systematic approach to ensure that direct data transfers not required.<br>• PPI mostly limited to review of access requests.<br>• No EDI considerations.<br>• Undue and disproportionate reliance on commercial organisations to comply with requirements, which does not build trustworthiness into the system.<br>• Contradictory and ambiguous language in guidelines. |

#### 3.8 Supplementary Table 8: LEAG member's review of DARE UK (2022)

Direct quotes are from the material reviewed. (Not all direct quotes from the primary source document are thus identified.)

| Domain | Reviewer's comments |
|---|---|
| **Elements worth including** | Glossary: Include a clear glossary (1–4 pages) with the guidelines. Several terms can have multiple meanings and it is important that the specific intended meaning for certain terms (in the context of the guidelines) is conveyed within the guidelines document.<br><br>Glossary definitions are in plain language, although some definitions and distinctions could be clearer e.g. "the governance that exists around it" for data research infrastructure, how public engagement is different to public involvement and the definition for a SafePod.<br><br>Purpose of data use: The purpose of the use of sensitive data is important. It is important that use of such data is for the "public benefit." However, anything can be framed as being for the public good. Public contributors should be involved in determining what they believe is for the public good. It is important that the benefits of any research can be shared across a diverse population. More research could be conducted on what constitutes "public good."<br><br>De-identified data: Emphasis on the use of de-identified data for projects which are in the public interest.<br><br>Transparency of data use: Proactive transparency around all processes concerned with the use of sensitive data. Transparency over what data is being used, who is using it, what the data is being used for, and how it is being used so that the public is aware. This is also important because what is regarded as being for the public good is subjective and anything can be potentially framed as being for the public good. Making this information publicly available, beyond the public contributors on the data access request/approval panel, allows for greater reassurance. Public contributors and the public at large should be kept informed of research that uses sensitive data and the findings of that research. Reporting of research outcomes/findings back to public contributors. The risks and benefits of each research project using sensitive data should be communicated.<br><br>PPIE with diverse communities: Increased engagement with diverse communities, including engagement with specific groups who may have more concerns about the use of their sensitive data due to prior experiences or their current situation, e.g. ethnic minorities, refugees, and migrants.<br><br>Recruitment of public contributors: Proactive and determined recruitment of public contributors, including those members of the public who do not know about PPIE, using a range of mediums, especially off-line methods, and reaching out to the public rather than expecting the public to find them.<br><br>EDI considerations for public contributors: There should be recruitment of diverse public contributors, including different ethnicities, ages, socioeconomic backgrounds, nationalities, those who are digitally excluded, those who are socially and geographically excluded and specific intersectional groups (e.g. older ethnic minorities). There are other considerations beyond these (e.g. religion, neurodiversity, gender)—see EDI section of this review. There should be targeted recruitment of groups that have been historically excluded, but it is important to recognise that people are more than their demographic information. In the same way that PPI should not be a tick-box exercise, EDI should not be a tick-boxing exercise, and people should be regarded as individuals rather than as a means to satisfy criteria. |

| | |
|---|---|
| | Meaningful public involvement: Public involvement should not be a tick-box exercise, it should be meaningful, and public contributors should be kept informed of the research and its findings (including the benefits for the public). Public contributors should be allowed to develop the required understanding (through training, for example) for meaningful contribution.<br><br>Researcher attitudes: Researchers should not be condescending towards members of the public, and should not regard them as being intellectually inferior or incapable. Researchers should be trained and "equipped with the necessary skills to engage with the public."<br><br>Public involvement on data access decision-making panels: There should be public involvement on decision-making panels for data access. There should be a simple application process for such public involvement roles, shorter duration public involvement opportunities (e.g., for those who cannot commit for longer periods) and payment/honoraria for time spent.<br><br>Transparency on data breaches and data misuse: Information on data breaches, data misuse, frequency of occurrence, and the consequences of such incidents, should be made available to the public.<br><br>Vetting of individual researchers: Individual researchers who request access to data should be vetted.<br><br>Monitoring of what researchers do with the data: Researcher use of data should be monitored and there should be an independent monitoring body.<br><br>Data sharing across the four nations and internationally: Consistent governance processes across the four nations, but there may be the need for some differences in processes based on national differences. There should be national core values and national standards.<br><br>Developing national standards: Public contributors from all four nations should be involved in developing national standards.<br><br>Standardisation and centralisation: Centralised processes can aid transparency and trustworthiness.<br>• Centralised process(es) for data access.<br>• Centralised approach for researcher training and approval.<br><br>Clear information on the data access request and approval process made available to the public.<br><br>Data access request/approval panels should consider potential harmful uses of data and the effect on specific groups, when considering who gets access and for what purpose.<br><br>Regular reviews of security processes. |
| **Elements worth including with amendments** | Visualisation to accompany glossary: Alongside a glossary, we should include a simple visual depiction that shows the various parts and how they relate to each other. Equally, depending on how many terms we need to define, we could include the definitions on the visualisation itself.<br><br>A modified deliberative approach (addressing more than TREs and case studies) would allow for public contributors' understanding to be logically built and for various aspects of the topic to be explored.<br><br>Transparency of data use: Transparency over what data is being used, who is using it, what data is being used for, how it is being used and what is being done to ensure biases are addressed. A public registry |

with supporting information in plain-language format and an awareness campaign may help with transparency. Research outputs made publicly available.

Public involvement on data access decision-making panels: Public involvement on decision-making panels for data access. Simple and inclusive application process, inclusive and flexible opportunities for involvement (e.g. flexible time commitment, breaks) and remuneration for time spent. Should there be different data access decision-making panels, with involvement of contributors from affected groups on each panel?

A requirement for meaningful and diverse PPIE in all research using sensitive data.

Transparency on data breaches and data misuse: Information on data breaches, data misuse, frequency of occurrence, and the consequences of such incidents, should be made available to the public.

- There should be a mandatory requirement for users of sensitive data (i.e. in this case, commercial organisations) to report such incidents to the respective data controller and an independent monitoring organisation. This information should be made publicly available by the data controller / independent monitoring organisation via a public registry. The public registry entry could include subsequent measures taken to mitigate negative consequences and any remedial actions. There should not be a culture of cover-up, as appears to be the case in many aspects of the healthcare system, as this will erode trust.
- There could be a means for researchers to confidentially report potential misuse at their organisation (i.e. whistleblowing process).
- There could be an additional process that seeks to investigate any incident, how it occurred, and what could be learnt from it. Additional training requirements could be identified and necessary changes could be made to processes and systems to ensure that the risk of such incidents re-occurring are minimised.

Vetting of individual researchers: Individual researchers who request access to data should be vetted. More trust placed on systems rather than individual researchers, with references made to regular security updates and reviews of governance structures.

- Rather than specifying piecemeal actions, it might be better to develop an overarching approach that places more reliance on systems controls, i.e. rather than relying on individual researchers not misusing data, there should be system controls that make this very difficult to do. Such controls can include both procedural and technical controls within the environment.

Data access methods: It was felt that it is safest when researchers access data at a secure room at a TRE, followed by access in a safe room at an approved university, followed by access via a SafePod based at a university. The option regarded as being the least safe was researchers accessing data via a remote secure connection, with some participants being "deeply resistant" to this. Most participants in the follow-up workshop were supportive of researchers accessing data via a remote secure connection if researchers had been vetted and activity was monitored. However, only ten participants were involved in the follow-up workshop, there was lingering reluctance to trust individual researchers and two members were against this. There were specific worries around access via remote secure connection such as researchers being able to take photos of data.

- There are various other risks with data access via a remote secure connection. One such risk is increased risk of unauthorised individuals gaining access to the data if the researcher is accessing the data at home or at a public (working) space (which should be considered given increased prevalence of employees working from non-traditional workplaces since the COVID pandemic). Remote secure connections encompass several possibilities and this should not be assumed to mean data access from a traditional workplace like an office.
- Access for researchers/staff at commercial organisations should be in a secure environment such as safe room or SafePod at a university/hospital/TRE. There should not be the option for

| | |
|---|---|
| | staff at commercial organisations to access sensitive MH data via a remote secure connection.<br><br>TRE regulation: Independent regulatory body for the governance of TREs. This may be out of scope for this work, but this could be a recommendation for government or executive/regulatory bodies.<br><br>A single centralised process would arguably not be sufficient and it may be advisable to have a set of centralised processes:<br>• Separate centralised process for access to MH data;<br>• Separate centralised process for access to other health data;<br>• Other centralised processes for access to other types of data;<br>• Centralised process for permissions for data linkage.<br><br>Different access requirements for different data: Vary access requirements depending on the sensitivity of the data and associated risks.<br>• However, rather than being a subjectively applied set of requirements, there should be clearly defined categories that allow for different requirements for different and clearly defined categories of data (e.g. more stringent controls and monitoring for access to MH free-text data by permitted types of organisations). For certain types of data (e.g. MH free-text data) access by certain types of organisations (such as commercial organisations) should not be allowed.<br><br>Commercial access to sensitive data: There was concern amongst participants about commercial and government access to sensitive data. For commercial access, there were concerns that profit would be prioritised over serving the public good, misrepresentations of findings and the reluctance to share the findings of commercial research.<br>• There could be a requirement for open research—to make all methodologies, data, and findings publicly available, so that this can be scrutinised and to aid with reproducibility and replicability.<br>• A requirement on all commercial organisations to contribute funding (for completed projects) for a university to scrutinise the research and aim to reproduce the findings.<br>• Resources to check the outputs/findings of commercial research using sensitive data for bias?<br>• It should not be left entirely up to commercial organisations to determine who has access and how many employees have access to the data. This is something that should be included and justified on the data access request and subject to approval. |
| **Noted to avoid** | Transparency at an individual level—individuals being informed every time their sensitive data is stored, transferred, or used for research. This would be impractical for both data controllers and members of the public. It would result in many individuals receiving too many notifications. It would also place a huge technical burden in terms of the number of notifications that would have to be sent out (with energy and financial costs) and the risk of and legal implications in case notifications were not sent/received due to technical issues.<br><br>Information about sensitive data use in research should be given at the point at which people "give" their data. However, this would not be practical and place an enormous burden on public services. It is also not clear what this would achieve if people have no choice over the use of that data, i.e. would they decide not to use healthcare services, even in an emergency, because they did not want their data to be used in this way? Even if patients are given information on their right to opt out of data use, would it be appropriate to give this information when a patient is at A&E or when a patient is receiving a cancer diagnosis?<br><br>For GPs to inform individuals about the use of their data for research. This is not practical and would place too much of a burden on GP practices. While this would be unsustainable and unfeasible at an |

| | |
|---|---|
| | individual-level, it could be possible to implement awareness campaigns using GP surgeries.<br><br>Data misuse helpline—it is not clear what this would be for, who would use it and how practical this would be to implement.<br><br>Addressing individual concerns about data use (for members of the public) may not be practical. However, of course, researchers should be able to report concerns about data use at their organisation.<br><br>Merging of all sensitive data storage so that there is centralised storage for all sensitive data—this would be inappropriate and much riskier, presenting huge privacy and security risks.<br><br>Independent regulatory body for the governance of TREs. As above: this may be out of scope for this work, but this could be a recommendation for government or executive/regulatory bodies.<br><br>At home access to sensitive data via a secure remote connection. This would not be appropriate given the increased sensitivity of MH data, the fact that it is unconsented for non-clinical use, and the reluctance among a large proportion of the public to share such sensitive data with commercial organisations. For MH data, access for researchers/staff at commercial organisations should be in a secure environment such as safe room or SafePod at a university/hospital/TRE.<br><br>Varying access requirements depending on the urgency of the research.<br><br>In the follow-up workshop, participants were against different requirements for different types of sensitive data. However, it may not be appropriate to treat all types of data in the same way, especially for commercial access. Arguably MH data is more sensitive and there should be stringent controls around access.<br><br>In the follow-up workshop, participants decided that there should not be different criteria for different types of organisations (academic, government or industry). However, only ten participants attended the follow-up workshop and this view was in sharp contrast to the views expressed in the initial workshops. Note also that the ten participants were chosen by DARE UK.<br><br>Checking all research outputs/findings—there may not be sufficient resources for this. However, if all research findings and data are made public, this would allow for scrutiny of that data.<br><br>Tours of TREs for the general public may not be advisable—for practical reasons, the lack of resources to offer this and the increased security risks posed. Video tours of TREs (at undisclosed locations) could be offered. |
| **Gaps** | No mention of what a data access request should include (beyond the reason for access) and no mention that information in the request should be provided in plain language (for public reviewers).<br><br>Whether patients and the public will be equal partners in the decision-making process (i.e. will their opinion be given equal weight?). This may be more appropriate for unconsented MH data, while it would not be appropriate for all types of data.<br><br>No explicit mention of access to or use of identifiable data, or is this implied? DARE UK is focused on data linkage, but it appears that they have completely skipped the potential use of identifiable data for data linkage. Access to identifiable MH data or "personal data" should not be granted. |

| | |
|---|---|
| | Duration of access. Data access and use should not be granted for an indefinite period and both the access request and access approval should explicitly mention the duration. If access is required beyond the approved duration period, then a (formal) request should be made for renewal of access.<br><br>No clarity on the kind of sensitive data that would be in scope—it appears that all sensitive data is in scope.<br><br>No mention of regular project progress updates.<br><br>No mention of regularly reviewing standards for data access.<br><br>No mention of regular monitoring, beyond apparent (real-time or continuous?) monitoring of researcher use of data.<br><br>No mention of regular audits of data access, use, or outputs.<br><br>There is no reference to patient involvement in monitoring and auditing data access and use (whether reviewing progress reports for this purpose or conducting audits).<br><br>There is no mention that the commercial organisation is prohibited from publishing any identifiable/personal patient data.<br><br>There is no mention of what steps will be taken in the event of a breach/violation of the data agreement or contract. How and how soon would all access to the data by the commercial organisations in question be revoked and how would this be ensured?<br><br>No reference to any requirement for patient and public involvement at the commercial organisation (from application to project completion).<br><br>It would be appropriate to explicitly state that any commercial organisation that is granted access to any data is not acquiring any intellectual property (IP) rights to the data, nor will they gain any other exclusive rights with regards to access or use of the data.<br><br>Addressing the issues in research culture, e.g. the reproducibility and replicability problems with research. Such problems directly affect patients and the public, and can cause much harm rather than the research being for public benefit. There have been specific issues with commercial organisations misrepresenting the benefits and side effects of [certain psychotropic] medications […].<br><br>There is no mention of fees for commercial access to sensitive data.<br><br>There is no distinction between commercial organisations from different industry sectors.<br><br>Other gaps in addition; not all covered. |
| **Public involvement approach** | Apparently public involvement and engagement is embedded throughout the programme (which would therefore include all programme phases).<br><br>There is recognition of the fact that public support should not be taken for granted—the specifics of a |

given project affect public expectations.

Board representation: Four members of the public sit on *"DARE UK Programme Board and Scientific and Technical Advisory Group, to provide valuable input into decision-making processes."* This is insufficient for a programme of this magnitude (https://dareuk.org.uk/involving-the-public/). See also EDI limitations.

Delivery public advisor: There is a Delivery Team Public Advisor, who is consulted on *"day-to-day activities to ensure public involvement is embedded across all aspects"* of the work. For a programme of this scale, would a single "Delivery Team Public Advisor" be sufficient?

Planned in-person workshops in four UK cities: Initial workshops for members of the public were planned as in-person workshops in four cities across the UK. These were only changed to online workshops due to COVID-19. Hence, it is unlikely that many people from outside of the four cities would have been recruited. If online (or hybrid) workshops had been planned from the outset, this would have allowed:

- For recruitment from more places across the UK, including from rural locations. Due to the planned in-person workshops, participants were mostly recruited from four cities.
- Those with disabilities/caring responsibilities or those who could not travel for other reasons to participate also.
- For recruitment of a larger number of participants (easier to organise more online workshops, travel costs do not have to be reimbursed so more resources are available).

Lack of adequate EDI and accessibility consideration for participants.

Local participant recruitment strategy in four cities by lay community recruiters: In each of four cities across the UK, a lay person was recruited by a local charity to take on the role of "community researcher."

- These "researchers" tailored recruitment strategies to localities. While involving members of the public in recruitment (and implementing local recruitment strategies) can be effective, care still needs to be taken to ensure that a sufficiently diverse sample can be recruited.
- Outsourcing to a community recruiter should not be seen as a way to circumvent all other EDI considerations, but rather this should be seen as one means of helping to achieve EDI objectives.
- Would have been appropriate to have a national recruitment strategy to ensure that many different groups were considered at national and local level.

Problematic exclusion criterion for participants: excluded participants who had been involved in PPIE activities on research data, apparently "to capture a range of different views on the subject—including both spontaneous/uninformed views and, as the workshops progressed, informed views."

- Unlikely that participants were going to develop any sufficiently "informed" views anyway given the limited duration and scope of the workshops.
- Excluding those who had participated in (minimal) PPIE activities on research data, but potentially including those with extensive PPIE experience in clinical research (which uses and depends on research data) may result in the involvement of more participants who are in favour of sharing sensitive data.
- Participants who had not been involved in PPIE activities focused on research data may still have experience in working with sensitive data, e.g. they may work in data analysis.
- Hence, this seems like an arbitrary exclusion criterion that may not have been particularly effective.

| | |
|---|---|
| | Insufficient number of participants:<br>• 44 participants—not enough participants given the scale of the programme, and DARE UK agrees that *"44 people cannot represent the diversity of views across England, Northern Ireland, Scotland and Wales."*<br>• 44 people would also not allow for sufficiently diverse recruitment.<br><br>Public involvement in Workshop design: The design of the workshops (initial and follow-up) was decided by 13 PPIE professionals and only one public contributor. More public contributors should have been involved in designing the workshops. However, community researchers/recruiters were able to provide feedback on the workshop activities and this feedback was used to refine the activities.<br><br>A deliberative approach was taken—*"Deliberation emphasises logically building people's understanding about a topic, drawing out multiple perspectives and trade-offs rather than driving at a consensus and allowing time for views to be expressed and developed."* This involved building understanding on a topic, *"followed by interactive activities and group reflection time on that topic,"* all of which is then repeated for the next topic, and so on. However, having reviewed the workshop materials and exercises, it appears that the way in which the exercises were framed and designed was to almost "direct" participants towards particular "outcomes" or "insights."<br><br>How were the key topics to be covered in the workshop decided? PPIE involvement in deciding key topics?<br><br>Follow-up workshop: Only 10 participants were included in the follow-up workshop to check whether participants' views had been accurately captured (with the opportunity for further deeper discussions) and to decide tangible actions/recommendations. Arguably, all 44 participants should have been involved in the follow-up workshop. No information on how participants for the follow-up workshop were selected, and beyond the fact that the 10 participants were from across the four nations, no demographic information has been provided.<br><br>Addressing concerns of PPI tokenism: Keeping participants informed of impact.<br><br>Payment: Rates of reimbursement were in line with expectations (e.g. £150 for full-day), but payments were in "digital vouchers" rather than cash which may have deterred some people from becoming involved, e.g. someone with a lower household income may not be able to justify spending a whole day on this activity and paying tax in cash for a voucher that they may not use.<br><br>Awareness campaigns recommended: Mixed-media awareness campaigns for the use of sensitive data in research. Some participants felt a lack of control and disempowered due to not knowing that sensitive data collated via public services was even used for research. Must be a more determined and proactive effort to inform people. Messaging and mediums used must cater for a diverse population, including those who are digitally excluded, those who do not have contact with healthcare services and those who are geographically isolated. The findings do not mention those who are socially excluded, but this is an important group too. National awareness campaigns can be locally tailored and targeted. There should be meaningful transparency, with the public provided with relevant details clearly, and not overloaded with irrelevant information.<br><br>Needs to be proactive recruiting of public contributors by researchers, especially members of the public who do not even know about PPIE. |
| **Industry involvement approach** | Industry involvement was separate and not part of the public dialogue. The document reviewed only constitutes the output from workshops with members of the public. |

| | |
|---|---|
| **Equality, diversity, and inclusion considerations** | Board representation: Four members of the public sit on "DARE UK Programme Board and Scientific and Technical Advisory Group." The members are not sufficiently diverse or representative of the general public:<br>• [This number prohibits representativeness of UK demographics.]<br>• Their involvement in research, healthcare, and data science, may mean they are not able to provide a critical perspective (one is a health services researcher, one is the COO of a company focused on data science, and one currently works in healthcare having previously worked in the NHS).<br>• No or inadequate EDI considerations.<br>Can they represent or understand the perspectives of members of the public who have concerns about sharing sensitive data?<br><br>Accessibility of workshops: Initial public workshops were planned as in-person workshops in four cities across the UK. These were only changed to online workshops due to COVID-19. If online (or hybrid) workshops had been planned from the outset, this would have allowed:<br>• For recruitment from more places across the UK, including from rural locations. Due to the planned in-person workshops, participants were mostly recruited from four cities.<br>• Those with disabilities/caring responsibilities or those who could not travel for other reasons to participate also.<br>• For recruitment of a larger number of participants (easier to organise more online workshops, travel costs do not have to be reimbursed so more resources are available).<br><br>A range of dates for online workshops, and allowing participants to choose dates and times, would have allowed more diverse people to become involved.<br><br>Accessibility of full-day workshops: Organising full-day workshops would have excluded people who could not manage to attend for the full-day due to health-related reasons, caring responsibilities, socioeconomic and/or other reasons.<br><br>Requiring participants to learn so much in one session before sharing more "informed" views is not reasonable and can exclude people who are neurodiverse, who learn differently and people from different socioeconomic groups.<br><br>Payment: Rates of reimbursement were in line with expectations (e.g. £150 for full-day), but payments were in "digital vouchers" rather than cash which may have deterred some people from becoming involved, e.g. someone with a lower household income may not be able to justify spending a whole day on this activity and paying tax in cash for a voucher that they may not use.<br><br>Participant recruitment: Local participant recruitment strategy in four cities by lay community recruiters: In each city, a lay person was recruited by a local charity to take on the role of "community researcher." These "researchers" tailored recruitment strategies to localities.<br>• Targeted recruitment from only four cities rather than from across the UK (as suggested) and rural areas were neglected.<br>• No information about the "community researchers" is provided. Some groups have been completely excluded among participants [e.g. of some ethnicities] and some are under-represented [e.g. of other ethnicities]. It is not enough to simply "outsource" local recruitment to "community recruiters" and believe that by doing this alone a diverse population will be recruited, as recruitment may be focused on certain demographics, depending on the background of the community recruiter.<br>• Many aspects of diversity have been completely ignored—religion, disability status, marital |

status, sexual orientation, neurodiversity, among others.

- Need appropriate oversight to ensure that sufficiently diverse groups are targeted. Minorities are not a single homogeneous group.
- Outsourcing to a community recruiter should not be seen as a way to circumvent all other EDI considerations, but rather this should be seen as one means of helping to achieve EDI objectives.
- Opportunity was advertised in "local community hubs"—there needs to be consideration of what community hubs and groups are being targeted.
- Would have been appropriate to have a national recruitment strategy to ensure that many different groups were considered at national and local level.
- Problematic exclusion criterion: excluded participants who had been involved in PPIE activities on research data, apparently "to capture a range of different views on the subject—including both spontaneous/uninformed views and, as the workshops progressed, informed views."
    - Unlikely that participants were going to develop any sufficiently "informed" views anyway given the limited duration and scope of the workshops.
    - Excluding those who had participated in (minimal) PPIE activities on research data, but potentially including those with extensive PPIE experience in clinical research (which uses and depends on research data) may result in the involvement of more participants who are in favour of sharing sensitive data.
    - Participants who had not been involved in PPIE activities focused on research data may still have experience in working with sensitive data, e.g. they may work in data analysis.

  Hence, this seems like an arbitrary exclusion criterion that may not be particularly effective.

Insufficient number of participants:

- 44 participants—not enough participants given the scale of the programme, and DARE UK agrees that *"44 people cannot represent the diversity of views across England, Northern Ireland, Scotland and Wales."*
- 44 people would also not allow for sufficiently diverse recruitment.

EDI considerations for workshop participants:

- Limited number of participants so insufficient diversity and representation.
- Almost equal participation from across the four nations, but arguably there should have been many more participants from England (given population distribution), although it is great that they were able to recruit participants from all four nations.
- Participation from across all age groups from 18 to 75+.
- Female, male and non-binary participants were included.
- Some ethnicities have been excluded […]. Ethnic minorities are not a single homogeneous group.
- No consideration of religion, disability status, marital status, sexual orientation, socioeconomic status, neurodiversity, and geographic area (beyond residency in one of the four nations).
- It appears that intersectional characteristics were not considered.
- Many more potential considerations—minority groups are not a single homogeneous group.

Workshop(s) design:

- Effective use of smaller groups: Splitting participants into smaller groups of 4–5 would have allowed for more opportunity to contribute and may have included those who feel uncomfortable contributing in larger groups. May have helped to mitigate the impact of the "tendency towards agreement rather than disagreement" in groups, which can "obscure the variety of views" in the group (more smaller groups can mean that a variety of views are expressed across groups).

- Lack of Inclusion for follow-up workshop to decide recommendations: Only 10 participants were included in the follow-up workshop to check whether participants' views had been accurately captured and to decide tangible actions. Beyond the fact that the 10 participants were from across the four nations, no demographic information has been provided. All 44 participants should have been involved in deciding recommendations.

| | |
|---|---|
| **Other considerations** | Regarding the sharing of sensitive data internationally, more information needs to be provided on how this would be carried out in practice, before a determination can be made on the potential risks of this, e.g. would access be granted via a SafePod at a University in another country? |
| **Summary of strengths and limitations** | Strengths:<br>• *"The DARE UK programme is being designed... with involvement from researchers, technologists and the public embedded throughout."* Although see PPI/EDI sections for limitations of public involvement.<br>• Emphasises the importance of transparency.<br><br>Limitations:<br>• A deliberative approach was taken—*"Deliberation emphasises logically building people's understanding about a topic, drawing out multiple perspectives and trade-offs rather than driving at a consensus and allowing time for views to be expressed and developed."* However, having reviewed the workshop materials and exercises, it appears that the way in which the exercises were framed and designed was to almost "direct" participants towards particular "outcomes" or "insights." A "deliberative" approach gives connotations of deeper thinking and thoughtful insights, but to what extent was this true? To what extent was critical thinking encouraged and facilitated?<br>• The key topics that were explored in the workshops:<br>◦ Trustworthiness (What would enable you to feel trust? How can trustworthiness be demonstrated? What is most important for trust? / What does your trust most rely on?);<br>◦ Data access (For whom? Approval processes? Purposes of projects? Data access methods? Remote access to TRE? Views changed due to pandemic? Data sharing across four nations?);<br>◦ Balancing risks and benefits (Risk minimisation? Articulating benefits to public? Ensuring research positively impacts people it represents? (representative research) Tiers of risk?);<br>◦ PPIE (How should public be involved? Keeping public informed? Language to use/avoid).<br>◦ The participants did not decide the topics. How were these key topics decided? PPIE involvement in deciding key topics?<br>• The only technical presentation was on TREs, and all other presentations were case studies. Hence, it is debatable how informed the participants were on key topics related to the use of sensitive data in research prior to discussions. Different case studies were presented to different groups (e.g. racial bias in court outcomes, air pollution, increased deaths at home during COVID-19, health and attainment of pupils, data linkage in children with type 1 diabetes), and this may have impacted what perspectives participants shared, i.e. participants' views may have been biased depending on the case study.<br>• The results and recommendations have not been analysed and broken down by specific demographics or by the specific case studies that were presented.<br>• Lack of Inclusion for follow-up workshop on recommendations: Only 10 participants were included in the follow-up workshop to check whether participants' views had been accurately captured and to decide tangible actions. Beyond the fact that the 10 participants were from across the four nations, no demographic information has been provided. All 44 participants should have been involved in deciding recommendations.<br>• No partnership with or involvement of organisations with opposing views (e.g. organisations |

who have petitioned against previous data sharing projects) so both viewpoints were not presented to the participants.

- For one exercise participants ranked data projects by level of potential public benefit and risk. However, depending on how a project is framed, anything could potentially be depicted as providing public benefit (e.g. […] Was the public good exaggerated for [certain psychotropic medications]?)
- (Not all covered.)

##### 3.9 Supplementary Table 9: LEAG member's review of Understanding Patient Data (2025)

Direct quotes are from the material reviewed. (Not all direct quotes from the primary source document are thus identified.) Some duplication reflects the relevance of comments to multiple topics.

| Domain | Reviewer's comments |
|---|---|
| **Elements worth including** | **Be trustworthy rather than trying to build trust:**<br>• UPD advocates for an approach focused on being trustworthy rather than developing public trust: *"Start with being trustworthy"* rather than trying to build trust (13).<br>○ It is not the public's responsibility to trust organisations who use their data, and members of the public cannot be criticised for not "correctly" trusting such organisations; the public should not be expected to trust "by default".<br>○ UPD argues that people's willingness to trust is influenced by their own subjective experiences and values (13). However, arguably, an individual's willingness to trust an organisation's data approach would also be influenced by how trustworthy the organisation is, the trustworthiness of the data approach and system, and the effectiveness of the organisation's communication of the system's trustworthiness. Some may claim to trust an organisation's data approach and system, without considering any information or evidence of trustworthiness—is that trust or indifference? Is that rational, thoughtful, and mindful trust or is it blind trust? Some may not have the willingness or capacity (e.g. due to other priorities) to ask questions or process information.<br>○ By ensuring that the onus is on ensuring that there is trustworthiness within the system, the organisation can consider what it can control to make the system more trustworthy:<br>▪ *"What can be done to make people, systems, and institutions worthy of people's trust?"*<br>▪ What *"practices and characteristics"* can be developed and what behaviours can be adopted to make a system/organisation more trustworthy?<br>▪ How can the organisation/system provide *"adequate, useful, and simple"* evidence that it is trustworthy (13)?<br>○ Interestingly, note that an approach focused on being trustworthy rather than building public trust is an inherently different approach. The "building public trust" approach typically has a narrative along the lines of "we know we are already trustworthy and we know we are already doing things properly, but how can we convince the uninformed public of this?" In contrast, the "being trustworthy" approach would have a narrative along the lines of "what can we do and what can we continue to do to be trustworthy and to demonstrate that we are trustworthy?" While UPD does not state the following, arguably it is a:<br>▪ Shift *towards* focusing on what risks exist and how they can be reduced or mitigated;<br>▪ Shift *away* from being over-confident and complacent about the existing systems in place to address any risks;<br>▪ Shift *towards* a continuous, ongoing, holistic consideration of risks, and how such risks can be reduced or mitigated. There are a broad spectrum of risks and the nature of, probability of, and potential impact of risks can change over time. An attack by a malicious external entity is not the only type of risk, and there are many types of risks; e.g. (a) someone with approved access, who is well-intentioned or who believes they are well-intentioned may perform risky actions when accessing patient data; (b) someone may inadvertently but irresponsibly share personally identifiable patient data with a third-party.<br>▪ Shift *away* from assuming, at any point in time, that there are no/few risks left to consider or taking an approach focused on a limited set of risks; |

    - Shift *towards* being open to considering patient/public concerns, the basis for the concerns, and how concerns might be addressed. There are a broad spectrum of risks and concerns, and there is a need to consider risks and concerns from a diverse range of perspectives. This may include a PPIE approach that proactively and meaningfully involves public participants with diverse views on patient data, including those who are cautious about or against the sharing of patient data with third-party organisations. It involves being accepting of diverse views, of proactively recruiting public participants with opposing views, and allowing those with opposing and diverse views and concerns to share them. It involves actively engaging with opposing and diverse views; it involves considering such views thoughtfully and seriously rather than dismissing them.
    - Shift *away* from (condescendingly) characterising one of the main issues as being an uninformed public with unfounded and unreasonable concerns. While some members of the public may be less informed, public concerns should not be automatically dismissed on the presumption that the concerns have no basis. Diverse patient and public perspectives can reveal risks and concerns that have not been (sufficiently) considered by experts/professionals/researchers. Some public concerns may be probable, some may not be probable, but may be possible. The challenge is not one of educating the public to the "one correct" way" of thinking.
    - Shift *towards* providing useful information and evidence to the public about what is being done to increase trustworthiness in the system. There is a distinction between what measures are being taken and the effective communication of this. If the latter is not done, then the public would not know what measures are being taken.
    - Shift *away* from communications for the purpose of persuasion.
  - There are some indications that public support for commercial access to health data (in general) has declined over time—53% supportive in 2016 versus 39% supportive in 2018 (16).
  - Much prior research has found significantly less trust among the public for commercial organisations with health data even if such data is de-identified (16).
  - Most organisations *"are trusted less with data than they are trusted overall"* (16).
  - Important to acknowledge that many *"people are uncomfortable with the idea of private sector organisations accessing health data"* and prior research has found that a sizeable proportion of the public do not want commercial entities to access patient data under any circumstances (16,18,20). By private sector organisations UPD is referring to for-profit (non-state, non-voluntary) entities.
  - Some prior research on public views has found a lack of understanding of commercial access to and use of health data (16). Meaningful transparency is an important aspect of being trustworthy.
  - Encouraging genuine dialogue rather than just "transmitting information" can help increase trustworthiness (16). This may involve listening to concerns, and then engaging and responding to concerns. Often communications focus on the importance of health data sharing for research without actively engaging with concerns expressed. Genuine dialogue may ensure that concerns are not side-stepped or dismissed.
  - The nature of specific research and whether specific entities are trusted by the public affect how the public feels about sharing health data (16).
  - Prior research has found that four conditions people would want in place before companies can have access to patient data is that:
    - Data is stored securely;
    - Identifying information is removed from data;
    - Data cannot be passed to third parties;
    - Sanctions and fines are imposed if companies misuse data (16).

    These can be ensured via technical and procedural controls to make the system more trustworthy.

- Do not generalise about public views—these can vary across people, demographics, time, and contexts (15–17). Also, public views can vary across specific types of data—e.g. while someone may be supportive of the sharing of consented patient data obtained from cancer trials, they may be against the sharing of routinely collected unconsented free-text MH data.

**What data?**

- Be clear about the kind/type of data (being considered, being shared, in an access request etc):
  - Is it individual or aggregated data? Is it about one person or many?
  - What is the identifiability of the data? Identifiable, pseudonymised, or anonymised?
  - What type of information does it include? E.g. diagnoses, medications.
  - Where possible, indicate when, where, and how data was collected, e.g. routinely collected or trials data (14)?
  - Being identified from data is a common concern. When explaining identifiability be accessible but accurate (14).
  - See "Elements worth including with amendments" for more on identifiability of data and free-text MH data.

**Motivation:**

- Important to be clear on why the data is needed. This may include:
  - Being transparent about the purpose(s) for which MH data can potentially be shared;
  - Requiring commercial applicants for data access to specify why they require access to specific data;
  - Being transparent on a public registry about specific purposes for which patient MH data has been shared.
  - See "Elements worth including with amendments" for further comments.
- Patient data to only be used where there is public benefit (13,14,16). Arguably, "public benefit" is an elusive concept that requires further definition. See also "Elements worth including with amendments".
- The use of patient data to improve healthcare should be prioritised over profit. There should be public and patient benefit over profit and other interests, regardless of what organisation is using the data (whether academic, commercial or charities) and this intention should be proven consistently over time (13). Please see "Elements worth including with amendments" on the elusiveness of the term "public benefit."
- Such benefits should be shared across the population and *"avoid making health inequalities worse"* (16). See also "Elements worth including with amendments" for further comments on considering the impact on health inequalities when assessing data access requests.
- Data host/controller organisations should be able to sincerely answer questions about motives.
- When communicating, rather than persuading, the aim should be to inform, allowing people to reach their own conclusions, i.e. organisations should not sell their data approach to the public (13). This ties in with taking an approach focused on being trustworthy rather than trying to build trust (see comments above).
- There is low awareness of how patient data is used to improve healthcare services via research and people often cannot relate the benefits of use of data in research to themselves or their loved ones.
  - While this can occur due to lack of public understanding of data science research, for some sub-groups it may also be due to the struggles they experience within the healthcare system. For those who struggle to access standard healthcare in a timely manner, or expected emergency care when required, they may understandably not

consider healthcare research and innovation. Additionally, those who suffer health inequalities—inequalities in access to healthcare in a timely manner and inequalities in outcomes—may benefit less or may understandably believe that they would benefit less from research and innovation, [e.g.] someone who has been repeatedly mis-diagnosed […] has inevitably not been receiving state-of-the-art new treatments.
    - When describing public benefit of data use, this should be accessible and clear, preferably with the use of examples; show the public benefit in an accessible way (14).

**Transparency and "meaningful transparency":**

- Acknowledgement of and openness about risks and uncertainty; do not deny that risks exist, be open about them and share steps being taken to mitigate those risks (13,14). Explain *"how patient data is kept safe and secure"* through "the Five Safes framework" (14).
- Openness about the risks of using patient data and also indicating the risks of not using patient data for research. *"Acceptable balance between risks and benefits will vary"* between people (14).
- Meaningful transparency even if all the information is not known at the start or if the context/situation changes over time; when a situation is developing rapidly… *"be clear about the principles you're working to and what could change"* (13).
- Openness about who can access patient data, what they can access, why they might access data, how they can access data, how data is used and what data access controls there are in place (11,14). Important to the public: *"who sees what data and under what circumstances and how is that controlled"* (13).
- Clear information about what is allowed and what is not allowed with regards to the sharing of patient data, e.g. is there a particular type of data that will not be shared, or particular purposes for which data access will not be granted? If there are strict rules, what are the exceptions to those rules (14)?
    - An overly simplistic approach which does not distinguish between different types of data, and essentially allows all data to potentially be shared, gives cause for concern. Arguably, it strongly implies a lack of sufficient deliberation on the different types of data and the different concerns related to different types of data. It implies that the default position of sharing all data was taken without deliberation of whether all data should be shared, especially with commercial organisations.
    - If different types of data have been carefully considered, and some types of data will not be shared with commercial organisations (e.g. free-text MH patient data) then it would be appropriate to indicate this. Being too simplistic in communication can give cause for concern. Instead, including an appropriate level of detail can be helpful.
    - Clearly indicating what specific data is available for sharing and why, and what specific data is not available for sharing and why, can imply a more deliberative approach with consideration of patient/public concerns.
    - Please see "Elements worth including with amendments" for further comments on clarification of the sharing of different types of data.
- *"Decisions about third party access to NHS data should go through a transparent process and be subject to external oversight"* (11,19).
    - While UPD refers to the need for the data access process to be transparent, it does not explicitly recommend that the end-to-end data access process should be documented, including in an accessible format, and be made publicly available (online). It would be appropriate to include an MH data guideline stating this.
    - There should be transparency about who is involved in making decisions about granting access to patient data and how these decisions are made (11,14).
    - For decisions about third-party data access, it is not clear what form the "external oversight" should take and what specifically should be subject to external oversight, e.g. should local data access processes be subject to an external and independent audit? See

"Elements worth including with amendments" for further comments.
  - A public registry of approved data access requests might allow for some public scrutiny (see below also). However, the level of public scrutiny might depend on what information is provided on the public registry.
  - Please see "Elements worth including with amendments" for further comments on the data access process and governance and monitoring.
- Make relevant information (that is important to people) easily available, findable, accessible, and understandable (13).
- Ensure that *"accessible information is provided at the right time and in the right context for patients and the public"* (11).
- Do not overload people with too much information or "technical language"—this is not meaningful or relevant (13). In UPD commissioned research which included a citizens' jury, jurors *"felt it was crucial that information about partnerships is understandable and with an appropriate level of detail. Providing dense technical information, redacted legal documents or vague assurances of privacy to patients is not meaningfully transparent and could further undermine trust by giving the impression that there is something to hide"* (11).
- Important to consider how information is communicated. Consider local/regional communications and engagement initiatives about the use of health data, tailoring information to local context (11).
- Rather than just "transmitting information" encourage genuine dialogue (16). This may involve listening to concerns, and then engaging and responding to concerns. Often communications focus on the importance of health data sharing for research without actively engaging with concerns expressed. Genuine dialogue may ensure that concerns are not side-stepped or dismissed.
- While UPD does not appear to explicitly state that a glossary should be provided, they do define terms clearly on their site. Hence, include a clear glossary with the guidelines. Several terms can have multiple meanings and it is important that the specific intended meaning for certain terms is conveyed within the guidelines document.
- Provide information on patient rights to access own data and information.
- Clearly explain patient rights and choices about how patient data is used—consider applicability of the choice, national differences, specific terms, impact of choices and possible exceptions. Ensure this information is accessible, and *"easy to understand and act upon"* (14).
- Open research—methodologies, code, and data should be made publicly available to enable research to be scrutinised and for reproducibility (13). This may also help towards addressing some issues with MH research and issues in research culture.
- Based on the UPD content reviewed, there is no guidance on implementing a public registry of approved data access requests. However, UPD does refer to the fact that the NHS already *"publishes details of every organisation that uses NHS data"* (21).
- See "Meaningful Transparency" in "Elements worth including with amendments" for further comments including on a public registry of data access requests.

**Competence:**
- Right people—ensure that staff working with the data have the skills, *"expertise, competencies, tools and methodologies"* to do this safely, securely, effectively, only for intended purposes, and to complete the proposed research (13).
  - Appropriate to include this in the MH data guidelines. Regardless of the implementation of other technical and procedural controls, taking measures to also ensure staff competence provides greater assurance and increases the trustworthiness of the system.
  - Note, however, that UPD is referring generally to anyone using patient data, rather than commercial entities specifically.
  - In the case of commercial data access applicants, the host organisation's data access

process could require that the commercial entity names the staff who will work with the data on the application. The job titles and experience of the staff could be specified, with the host organisation then vetting named staff.
  - Where multiple commercial organisations will be working on a single project, the names, and details, including affiliated organisation, of all staff across all organisations who will access the data could be provided.
  - The capacity of the host organisation to thoroughly vet commercial entity staff will be limited, and there will be reliance on the commercial entity to ensure staff granted access to the data are appropriate and can be trusted. It may be appropriate to include this condition in the data partnership agreement terms.
  - If there are technical controls in place to ensure that data is only accessed via a SDE [Secure Data Environment], that only approved data can be accessed, that identifiable data cannot be accessed, that data access and use can be audited, that all data outputs (from the secure environment) are suitably anonymised and restricted, and that data cannot be shared with third parties, this would mean that undue reliance is not placed on staff (competence) at commercial organisations. Procedural controls can also be used to limit undue reliance on staff at commercial organisations, e.g. external audit and monitoring processes.
  - Staff at commercial entities do not have to be granted access to all types of MH data. For certain types of highly sensitive and private MH data (e.g. free-text MH patient data) access does not have to be granted to commercial entities. If there is a good and valid reason for accessing such data, one solution may be to require commercial organisations to conduct the research in collaboration with a university, with access to the free-text MH patient data only being granted to the university. Even when access to such data is granted to academic researchers this should be done via an interface whereby there is no direct access to the data, and only questions can be asked to a system which then processes the free-text MH data to return non-identifiable answers to the researcher. This limits undue reliance on staff competence (whether commercial or academic staff).
  - UPD says teams should ensure that data is only used for *"intended purposes."* Since the intended purpose may be inappropriate and/or differ from approved purposes, it would be better to state that patient MH data should only be used for approved purposes by the commercial organisation.
- Aside from staff competence at commercial organisations, there is a need to consider staff competence within the data host/controller organisation.
  - Prior studies have found that the public have concerns about *"lack of in-house technical capacity and expertise"* within the NHS to effectively manage risks, and to effectively and safely use and share data (16).
  - Concerns have also been raised about the lack of NHS in-house understanding and skills to negotiate fair terms for data partnership agreements with commercial entities and how this may result in various issues including commercial exploitation of patient data (11,15).
  - Consequently, studies have found that the public *"place a high priority on independent oversight, transparency, and accountability"* (16).
  - An overarching governance framework, oversight organisation(s), a *"coordinated national strategy and guidance",* monitoring activities, appropriate reactive activities for issues, free expert advice, and a central registry, may help to provide oversight, transparency, and accountability (11,19). See also "Governance and Monitoring" in "Elements worth including with amendments".
- EDI considerations for staff—staff should be representative of the population (13).
- Staff should have *"communication and engagement skills"* so they can explain "how data is used," what the public benefit impact is, and to be open about risk management. The existence of risks should not be denied; it is important to be open about risks and to share

steps being taken to mitigate those risks (13).

- See also "Competence" in "Elements worth including with amendments".

**Data access processes:**

- Clear, consistent rules and processes rather than *"a patchwork of rules, criteria and processes"* to determine data use to enable greater transparency, fairness, and to inspire public confidence (13). However, UPD does not explicitly state that data access processes should be formalised, documented, and made publicly available (which should be the case).
- *"Involve members of the public in setting the rules and frameworks for decisions about data,"* including *in "deciding which uses of data are or are not permitted"* (13).
- Clear rules on what is allowed and what is not allowed with regards to the sharing of patient data, e.g. is there a particular type of data that will not be shared, or particular purposes for which data access will not be granted? If there are strict rules, what are the exceptions to those rules (14)?
- People may want reassurances, including that *"a patient's full record will only be seen by healthcare [staff]"*. Do not share any patient's full health record and only share the minimum amount of data necessary for the approved purpose (14).
- See "Elements worth including with amendments" for further comments.

**Governance and monitoring:**

- *"Someone has responsibility for oversight about the purposes for which data can be used"* (14).
    - It would be appropriate for a designated person or panel to be responsible for overseeing the overall purpose(s) (i.e. the defined purpose(s)) for which data can be shared at each data controller. This individual/job role and/or panel should be named on data access documentation, which is made publicly available.
    - However, arguably sufficient detail has not been provided by UPD on what responsibilities this role/panel would have, nor on the level of PPI for this.
    - Please see "Motivation and Purpose for Data Use" and "Governance and Monitoring" in "Elements worth including with amendments" for further comments.
- *"A single point of guidance and oversight should be established to set consistent rules across the NHS for third party access to data"* (19). External independent oversight (16).
    - A *"coordinated national strategy and guidance"* (11);
    - *"Consistent rules across the NHS for third-party access to data"* (11);
    - Implement *"a clear, overarching governance framework for NHS health data use"* (11). There could be a single governance and oversight organisation or this function could be *"spread across governance and oversight structures including the National Data Guardian, existing regulators and the new National Centre of Expertise"* (11). Even if this function is carried out by various local/regional bodies, *"all data partnerships must adhere to the same standards of accountability"* (11).
    - Please see "Governance and Monitoring" in "Elements worth including with amendments" for further comments.
- National-level governance and oversight for data partnerships (which may be implemented by local/regional organisations) and this should include:
    - *"Proactive activities: establishing principles and ensuring a good governance framework"* (11).
    - *"Monitoring activities: auditing and reporting"* (11).
    - *"Reactive activities: firefighting, public investigation and regulatory action where needed"* (11).
    - Lack of detail provided by UPD; please see "Governance and Monitoring" in "Elements

worth including with amendments" for further comments.

- *"Must be robust measures in place to reduce the risks as much as possible"* (14).
- Access controls to control *"who can access what data, when and how"*—these can be technical controls and procedural controls enforced by people. Important to the public: *"who sees what data and under what circumstances and how is that controlled"* (13,14).
- Ensure there are mechanisms to learn and adapt over time to contextual, technological, regulatory, and other changes, e.g. how can processes be changed? (13)

**Accountability:**

- Provide information on *"who is responsible for what decisions"*;
- Provide clarity on what happens if something goes wrong;
- *"It needs to be clear which person, or group of people, is ultimately responsible if something goes wrong"* (13).
- Offence to re-identify anyone from data granted access to (14). However, note that this may be difficult, if not impossible, to detect, unless there are appropriate preventative and detection controls in place.
- Make clear what sanctions/remedies will be imposed if something goes wrong;
- Enforce sanctions/remedies when something goes wrong;
- Prior research has found that one condition people would want in place before companies can access patient data is that sanctions and fines are imposed if companies misuse data (16). Thus, it is important to be transparent when things go wrong (including about incidents of data misuse by companies) by clearly communicating what happened and what subsequent actions (including imposition of sanctions/remedies) were taken.
- *"Public and democratic accountability"* through meaningful transparency and independent scrutiny when something goes wrong, in addition to legal protections/mechanisms (13).

**Elements worth including with amendments**

**Types of private organisations that can access patient data:**

- Currently, *"different types of private sector organisations may be allowed to use patient data, under strict conditions, for a range of purposes"* and organisations include software providers, pharmaceutical companies, analytics services, pharmacies, digital developers and technology companies, private healthcare providers and insurance companies (20). UPD states that commercial entities can mostly *"only access pseudonymised data for the purposes of helping deliver an NHS service or to undertake approved research projects"* (20).
- For purposes beyond the direct and present delivery/provision of healthcare, there is a need for more clarity on what types of commercial organisations can potentially access patient data. Specifically, there is a need for clarity on whether distinctions are being or will be made between organisations in different industry sectors.
- Further clarity is required on what types of commercial entities can potentially access patient MH data, and whether distinctions will be made between entities in different industry sectors.
    - Explicitly ask host organisations to specifically consider and decide, with meaningful patient and public involvement, their data access approach for different industry sectors and sub-sectors. When host organisations make decisions on the types of commercial organisations that can potentially access patient MH data, they should carefully and seriously consider diverse public concerns and opposition to certain types of organisations accessing this data. This should not be limited to the often-repeated (or parroted) concerns over insurance or marketing organisations gaining access to this type of data.
    - Any changes to the data access approach for different industry sectors or sub-sectors should be made with meaningful patient and public involvement.
    - Ask host organisations to provide clarity and to be (publicly) transparent on whether

distinctions will be made between different industry sectors when considering access requests for patient MH data. Host organisations should make this information publicly available in accessible form.

**Types of data/what data?**

- UPD advocates for being clear on the kind/type of data being shared (e.g. individual or aggregate; identifiability; information included in the data).

*Identifiable data*

- Prior research has found that one condition people would want in place before companies can have access to any patient data is that identifying information is removed (16).
    - UPD states that although the NHS will not share *"personally identifiable data for marketing or insurance purposes"* without explicit consent, personally identifiable patient data may be shared with other commercial organisations *"if there is a clear health benefit"* (20). Here, UPD may be referring to the sharing of personally identifiable patient data with private companies that are part of healthcare provision such as pharmacies or private healthcare providers, but they do not make this clear; there is no clarity on the situations in which personally identifiable patient data is currently being shared with companies. Sharing such data for the direct and current (or present, immediate) provision or delivery of healthcare is different to sharing it for research or other purposes.
    - For MH data, it should be made clear in the guidelines that personally identifiable patient data should not be shared with companies for research purposes or for any purpose beyond the direct and present delivery and provision of healthcare.
    - For unconsented MH data it would be completely inappropriate to share identifiable patient data beyond direct healthcare provision, and this should be explicitly prohibited. Being identified from data is a common concern (14). Such concerns are likely increased for patient MH data.
    - UPD refers to not sharing the full health record of any patient (14). Due to the nature of MH data (see below comments on free-text MH data) sharing a greater amount of MH data for a single patient may increase its identifiability (more so than other types of health data). Thus, in addition to indicating that the minimal amount of data necessary should be shared, it would be appropriate to also include a guideline indicating that the full MH record of any patient should not be shared. This should be the case regardless of claimed non-identifiability, as it would arguably be unnecessary and violate privacy to share the full MH record of any patient.
- Many *"people are uncomfortable with the idea of private sector organisations accessing health data"* and prior research has found that a sizeable proportion of the public do not want commercial organisations to access patient data under any circumstances (16,18,20,24). Previous research has found increased concerns with regards to specific types of sensitive data, including MH data (16). Thus, sharing such data should be handled with caution and this should be appropriately reflected in the guidelines. If guidelines are too relaxed or accommodating, for example, by permitting the sharing of personally identifiable MH data with commercial entities, this will massively reduce trustworthiness in the system.

*Data linkage between patient MH data and other data*

- Based on the content reviewed, UPD does not cover data linkage in any detail.
    - There is a cursory mention to the potential benefits of linking data in the summary report of research commissioned by UPD: *"The UK's health services steward large amounts of health data which has huge potential to lead to health improvements if it*

*can be collated, linked and analysed"* and how new *"techniques and processes—such as cloud computing and machine learning… make it possible to collect, use and link more data."*
  - There is no reference to the risks and concerns of data linkage.
  - Only one UPD guideline could be found which referenced data linkage: When communicating data linkage it is important to *"explain the reasons for linking the data and the process of doing it very clearly"* (16).
  - UPD does not cover data linkage for more highly sensitive data such as MH data.
- Given the nature, higher sensitivity, and potential higher identifiability of MH data (see also below comments on free-text MH data) arguably commercial third parties should not have direct access and ability to link patient MH data with other types of data.
  - Include guideline prohibiting commercial entities from linking MH data with other data.

*Free-text data*

- UPD omits any mention of whether the data is structured or free-text information (14).
- The sharing of free-text data is a particular concern for patient MH data, due to:
  - **The diversity of information contained within this data:**
    - Due to the nature of psychotherapy, patients may share any information about their life, e.g. education, employment, relationships, fears, abuse, trauma, dreams, nightmares, financial situation, sexuality, gender identity, plus more. This is in sharp contrast to the limited and largely symptomatic information shared for physical conditions.
    - Many patients may have concerns about this much information about them being shared with others.
    - It is difficult to think of any other example in any context where this quantity and diversity of extremely private and sensitive information would be shared with any number of third parties. There are no other precedents where this much extremely private and sensitive information is shared with third parties without explicit consent. This is being normalised, when it is anything but normal. Even in the legal system, a victim of abuse or rape can be granted anonymity. But if a patient discloses abuse/trauma when seeking help for mental illness in the NHS, why are they not entitled to privacy and for their details to be kept strictly confidential?
    - How would researchers feel about information about their most private experiences, together with diverse information about practically every aspect of their lives, being accessed and read by any number of third parties?
  - **The kind[s] of sensitive and highly personal information it likely contains:**
    - Patients may share extremely personal information with therapists, including about trauma and/or abuse;
    - Patients may reasonably expect such information to be kept private and not shared;
    - When patients are experiencing mental illness, and especially when they are experiencing severe mental illness or crisis, they may share information that they would not share otherwise. An individual in crisis may say or do things they would not say or do otherwise.
    - As such, many MH patients are vulnerable and there are serious ethical concerns about sharing highly sensitive personal information that such patients provide in confidence or that is recorded about patients when they are in very poor health;
    - Arguably sharing such free-text information (without their explicit and informed consent) is not treating patients with (basic human) dignity;
    - Many MH patients may experience the sharing of such information to be a (further)

violation and this can have an adverse impact on the patient.

    - There are no other precedents where this much extremely private and sensitive information is shared with third parties without explicit consent. This is being normalised, when it is anything but normal. Even in the legal system, a victim of abuse or rape can be granted anonymity. But if a patient discloses abuse/trauma when seeking help for mental illness in the NHS, why are they not entitled to privacy and for their details to be kept strictly confidential?
    - Knowledge that such information may be shared may impact what patients are willing to share with clinicians, which can impact their care and the effectiveness of therapy.
  - **Sharing this information undermines the practice of psychotherapy and risks adversely affecting therapist-client relationships, thereby affecting the effectiveness of psychotherapy treatment:**
    - An important and essential basis of the therapist-client relationship is for the therapist to maintain patient (client) confidentiality. By sharing such free-text information, the essential basis for an entire therapeutic discipline is being undermined. This is being done by only considering the apparent benefits of sharing such data (for research), and without considering the impact on the discipline and practice of psychotherapy. There is a reason why confidentiality is such an important foundation of the therapeutic relationship. Why is this confidentiality being considered obsolete now after the increased use and application of data science and AI?
    - The fact that an essential foundation of psychotherapy can be compromised through the destruction of confidentiality, and yet other aspects of psychotherapy are rigidly maintained (e.g. rigid rules on session duration, no contact or support outside of sessions) can cause patients to lose faith in psychotherapy;
    - Patients should not have to filter or moderate what they say to their therapist over concerns it will be shared with any number of third parties for non-clinical purposes. Therapy should be a safe space where the patient can be vulnerable and talk about anything. Knowledge that such information may be shared with third parties may impact what patients are willing to share with clinicians, which can impact the effectiveness of therapy.
  - **It significantly increases the risk of identifiability—there are concerns as to the extent to which patient MH free-text data can be truly be anonymous; in fact, it is highly unlikely that such free text can be anonymous and patients will be personally identifiable:**
    - Prior research has found that one condition people would want in place before companies can have access to any patient data is that identifying information is removed from data before it is accessed (16).
    - It is unclear how true/complete anonymity would be achieved in practice for patient MH free-text data. There are various issues, and any of these, or a combination of them may be used to identify patients:
      - Given the different structural formats of free-text data and the different types of identifiers they potentially contain, how would it be ensured that all identifiers are actually removed? Free-text data would contain free-text input on the system, letters and other documents from clinicians, free-text correspondence to patients (e.g. emails) and free-text correspondence from patients. For example, (1) in some correspondence a patient may be referred to by their full name (as on official documents and the NHS system) while on others they may be referred to by the name they want to be called; (2) there may be various references to patients' digital addresses/accounts/identities in emails from patients. It may be difficult to identify and remove all potential identifiers from emails or other free-text correspondence from patients.
      - MH free-text data can contain anniversary and milestone dates, which can be

used to identify patients. This is more a concern for MH free-text data because it is known that anniversaries and milestones can impact MH and hence patient MH records may include references to these. While an algorithm may change a patient's DOB, would the algorithm go through the patient's entire MH record and change other dates as well? If so, how would it know what dates should be changed versus which should be kept as they are? Presumably dates of correspondence (letters, emails) would not be changed, and if these refer to an anniversary last Tuesday (even if the date of the anniversary is not specified) this could easily be deduced. Inevitably, all or a lot of free text from a patient's MH record could contain substantial such information.

- A single unique experience or a combination of several experiences may be enough to identify patients.
- There may be reference to a patient's protected characteristics and these can be used to help identify a patient (in combination with other data). For example, a patient may belong to a religious group with relatively few followers and their religion may be specified in their record, along with their age, sex, race/ethnicity, and marital status, which together with other details from their record and publicly available information, could be used to identify a patient. Further, there are a diversity of ways in which aspects of each protected characteristic could be referenced, making it difficult for an algorithm to remove all such instances.
- A combination of information which may not be deemed to be "highly sensitive" within a patient's MH record could be used to easily identify a patient based on publicly available information on sites such as LinkedIn, e.g. text about education history, employment history, profession.

- **Legitimate and justifiable patient fears over discrimination/prejudice/stigmatisation:**
  - Discrimination, prejudice, and/or stigmatisation towards mental illness is unfortunately prevalent in society. Many MH patients may have experienced this due to their illness(es) and/or experiences. They may have even experienced discrimination, prejudice, and/or stigmatisation from MH clinicians and from other clinicians when accessing the healthcare system for physical symptoms and illnesses.
  - Such patients may justifiably have concerns about free-text data from their MH record being provided to various third-party organisations, and the wide-ranging adverse consequences this may have for them. Note the earlier comments about the virtual impossibility of MH free-text data being truly anonymous.
    - As an example, in some industry sectors there is prevalent partiality towards "resilient" employees with "thick skin"; the environment is cut-throat, and the willingness to be manipulative appears to be regarded as a sign of higher capability, and hence rewarded and respected. Such environments can include consultancies whose work includes data analysis. In such environments, "mental illness" is not regarded as illness but a weakness of character, and this may especially be the case if a patient's (past/current/future) colleagues/bosses were to gain access to free-text notes on the patient's suffering, episodes of crying and going into crisis. Even suicidal thoughts/tendencies are not viewed with compassion across society and colleagues can view such thoughts/tendencies with disdain.
- **Potential impact on patient interactions and relationships,** including safeguarding concerns due to predators potentially accessing highly sensitive patient information without the patient's knowledge. If free-text MH data is shared with third-party organisations, virtually anyone could know immensely private information about any MH patient and the patient may interact with any of those people at any point in their life and not know that they know all this about them. At best this may place the patient at a disadvantage, and at worst it may expose them to seriously unethical and predatory

practices. This may well give rise to serious safeguarding concerns. This is not an insignificant concern to be dismissed; it is extremely serious.

- Note the earlier comments about the virtual impossibility of MH free-text data being truly anonymous.
- Researchers from any organisation (commercial or other) do not live on a separate planet—they live amongst patients.
- Apart from the imbalance of power between therapist and patient, this is one of the main reasons why it would be considered unethical for a therapist and patient to have any non-clinical relationship: the therapist knows a substantial amount about the patient, which the patient has divulged by allowing themselves to be vulnerable, but the patient knows virtually nothing about the therapist. However, at least the patient *knows* that the therapist knows all this about them. If free-text MH data is shared with any number of third parties, any MH patient would not realistically know which of the people they interact with have read their MH record. This is a serious concern and risk.
- It is completely inappropriate to dismiss this concern by the assertion that the probability of the patient interacting with someone who is not their clinician and who has read their MH record is low.
  - Consider the regulations and limitations on the number of children a sperm donor may give rise to. In many countries there are strict limits to reduce the risk of accidental inbreeding or consanguinity between donor offspring. Limits can be as "low" as 3 children per donor or 3 families to receive sperm from a single donor in countries with populations of several million. In China where the population is over 1.4 billion, only 5 families may use the sperm from a single donor.
  - In contrast, there are no limits at all on how many individuals and third parties, a patient's MH free-text data can be shared with. Therefore, if free-text MH data is shared with commercial third parties, patients may be much more likely to unknowingly come across someone who has read their MH record. Currently, the probability of this occurring and the potential wide-ranging and adverse impacts of this are not being taken seriously at all. Note that the probability of this occurring would very likely rapidly increase over time, especially with the increasing use and application of data science and AI, with diverse applications being made for patient data access.
- It is also not appropriate to dismiss this concern by claiming that all researchers or other professionals from all organisations who access patients' MH free-text data are well-meaning, well-intentioned, and would never allow anything they have read in any patient's MH record to influence their behaviour.
  - Some professionals who request access to patient MH free-text data will unfortunately be actively prejudicial and discriminatory people, and even predatory. If you want to know the prevalence of prejudicial and discriminatory behaviour among "professionals," do not ask other clinicians/researchers/professionals, but ask MH patients and vulnerable people.
  - Some clinicians are predatory and cause intentional harm to patients even though they have been vetted by various processes. Most such cases of deviant clinicians are consistently missed and dismissed by their clinician colleagues. If you want to know the prevalence of this, do not ask other clinicians, but ask patients. Granting such deviants non-clinical access to patient MH free-text data would cause much harm. Right now, there are criminals that are registered and practicing as clinicians even though their criminal history is known and even though their crimes include abuse of children and sexual assault. It is not alarming that some criminals who have committed such crimes were doctors—that is to be expected, as there will be criminals in any profession. The

alarming issue is that the clinicians that are known to have committed serious criminal offences have in many cases been deemed fit to continue to practice. Further, these are only the clinicians who are known to have committed criminal offences, and there will be substantially more whose criminal behaviour remains unknown. There appear to be serious issues with the vetting and accountability processes. Yet, researchers appear to have absolute (over-)confidence in vetting processes for granting clinicians/researchers non-clinical access to patient MH data. How and why should patients and the public have confidence in such vetting processes?

- All professionals are first and foremost human beings, and all human beings have "issues", some amount of unconscious bias, and use various psychological shortcuts. A relatively small proportion of all people are prepared to habitually (i) question their own assumptions, biases, and the psychological short cuts their mind takes; (ii) think critically; (iii) reflect, and be open-minded about revising ideas, opinions, and conclusions. Many people are not aware of the assumptions they make, their biases, or even their prejudices. Therefore, even those who are not predatory or explicitly discriminatory, may be negatively influenced by patient MH free-text data, and cause harm as a consequence.

▪ This can impact the patient in terms of concerns they have about who knows what and their ability to build trust with those they interact with;

▪ This can adversely impact the patient by exposing them to disadvantages, prejudices, various harms, and even predatory practices. In many cases, the patient may know they are suffering because of the behaviour of a "professional" but they may not know how or why the individual has been able to do what they have. Note the potential for additional and aggravated harm because the patient would not be able to prove that the wrongdoer/deviant accessed free-text MH data about them.

▪ How would you (the reader) feel about:
  - Going to a job interview where the interviewer knows about your most private experiences (through reading your MH record) and not just what information you provided on your application form, while you know nothing about them? You would strongly suspect you are being discriminated against based on information that they should not know, but how would you prove this?
  - Going on a first date with someone who already knows about your most private experiences (through reading your MH record) but you know nothing about them? They would know about all your weaknesses and how to exploit them. You would not know that they know all this.

▪ You would have lost the right to keep certain information private. You would have lost the right to decide what to disclose and when to disclose it. How can this be considered appropriate in a civilised democratic society? How can it be considered appropriate to expose vulnerable people to these kinds of risks?

○ **It potentially containing misrepresentations and misleading information which can be very upsetting for patients.** The amount of misrepresentation, and wrong/misleading information, may be substantially higher for MH data compared to other types of health data due to the nature of the information, how it is subject to more subjective interpretation and bias of the healthcare professional, and the way in which such data is collected and added to patient records, e.g. routine blood test results or scans for a cardiology or oncology patient are more likely to be accurate than a MH professional's summary of a patient's educational and work history and a MH professional's summary of a patient's experience of trauma. Note also that it is likely much easier for a patient to question and challenge other types of health data (and get scans or tests repeated) compared to MH data. Many MH patients may not even know what is being (subjectively) recorded on their MH record, whereas they may well have copies of their blood test and scan results.

- Due to the unfortunate inadequacies of the MH system, and the time-limited treatment offered to patients, some patients may have provided free-text information which has been stored on their MH record. **This may include free-text correspondence from the patient while the patient is in a state of significant distress or crisis.** Patients who have provided such free-text information due to inadequacies, limitations, and failings in the MH system, would already have suffered due to such inadequacies/limits/failings. To then use their free-text correspondence, without explicit consent, for anything other than their clinical care, would arguably be exploitative and unethical.
- **The increasing likelihood of identifying patients from their own free-text correspondence.** Each person has their own way of writing, their signature, e.g. the vocabulary they use, the lexical/semantic field they use, their sentence structure, their writing style. It is possible to identify individuals from a piece of writing, and the accuracy with which this can be done will no doubt rapidly increase with the advancement of AI. If free-text correspondence from patients is shared with commercial entities, how would patient anonymity be assured? Note also the earlier comments about the virtual impossibility of MH free-text data being truly anonymous.
- **Legitimate concerns over what happens to patient data when/after it is fed into various large language models (LLMs) and other types of AI systems for non-clinical purposes.** While there may be restrictions in place with regards to the use of publicly available LLMs for clinical purposes, it is not clear what restrictions are in place when patient data is used for non-clinical purposes.
  - There are valid concerns over the potential retention of patient data when it is input into LLMs and other types of AI systems: (i) would it be retained within such systems? (ii) who would have access to the data? (iii) could it be regurgitated and shared more widely in the future?
  - Even if patient data is not entered into publicly available LLMs, private versions of LLMs may be used (which may nonetheless pose various concerns depending on the specific technology) and commercial entities may request access to patient MH data in order to train their own in-house developed LLM or other AI system on the patient data. In such cases, how would it be determined what patient data is retained within such systems without undertaking comprehensive testing? It seems highly unlikely that sufficient resources would be available to comprehensively test all such LLMs and AI systems: how can this be ensured and adequately funded? What would happen when the LLM or AI system leaves the secure environment?

- Serious consideration needs to be given to whether patient MH free-text data should be directly shared with commercial organisations. Arguably, the costs outweigh the potential long-term benefits. Currently, the vague but apparently irrefutable notion of "public benefit" is overshadowing all sensible, measured, and specific consideration of the sharing of unconsented patient MH free-text data. The assumed "public benefits" are causing certain practices to be approved without serious and adequate consideration of all the reasons why it would be completely inappropriate to share patient MH free-text data with commercial organisations.
  - There are particular and serious concerns regarding the sharing of patient MH free-text data with commercial entities (see above for comprehensive but not exhaustive list);
  - Most studies that say there is public support for sharing health data also say that this support is contingent on "public benefit" being obtained (15,16).
    - In such studies, "public benefit" is not defined. Yet, this argument is used to argue or even lobby for greater sharing of patient data, without distinguishing between different types of patient data.
    - Arguably, there is a lack of (unbiased) research, using sufficiently deliberative methods, on public views on the sharing of specific types of sensitive health data with commercial organisations. Note, that most studies do not specifically consider MH data, and none adequately cover patient MH free-text data. **Indeed, no prior research on public views can be shown to strongly support the sharing of**

**patient MH free-text data with commercial organisations.**

- While UPD states that people *"become more supportive of sharing patient data during the course of qualitative and deliberative studies"*, some deliberative research found that public views were actually more nuanced (16). In two 2017 citizens' juries, while jurors became more supportive of sharing patient data for public benefit in general, they were less supportive of sharing data for specific purposes (16). This is interesting, and suggests that while the public is supportive when the abstract and non-specific notion of public benefit is mentioned, they are more discerning and against the use of public health data in various specific contexts.
- Prior research is limited and there *"is limited evidence about how people would weigh up the benefits and risks for the kinds of data access agreements NHS bodies are making"* (11).

- Additionally, while recognising that it is not possible to know in advance all the possible ways in which such data could be utilised, there are valid questions around what publicly beneficial use this kind of detailed and highly sensitive information would have for the majority of commercial organisations, e.g. a pharmaceutical firm does not need to know patients' histories of trauma to develop new medications. Even for commercial uses of this type of data that can be "anticipated" there are valid questions over usefulness and acceptability, e.g. one commercial use might be to develop an AI that provides therapeutic support, but even in this situation, providing free-text data from patients' records does not help to better train an AI on how to interact with patients in real-time. Further, how acceptable is this? (i) how acceptable is it to use such private information to train an AI? (ii) how acceptable would an AI therapist be?
  - This does not discount the possibility that some limited data in free-text MH data might be useful for some commercial entities in some contexts, but it is difficult to imagine that a patient's entire free-text MH record would be required by any commercial entity. Further, there are more acceptable ways to preserve patient confidentiality and provide strictly limited data from patients' free-text MH data (see below).
- Nonetheless, in recognition of the additional value that free-text data offers over structured data for some types of research, and that there may be specific publicly beneficial commercial research for which this data may be useful, there is a need to consider whether this data should be shared with commercial organisations in such contexts, and if so, how this data should be shared. Prior research has found that it is more acceptable to share such data with academic researchers than commercial organisations. The potential for such highly sensitive and highly private information, which is likely identifiable, to be shared, without explicit consent, with any number of commercial organisations is highly unsettling.
  - For commercial projects that do need access to MH free-text data, one solution may be to require them to conduct the research in collaboration with a university, with access to the MH free-text data only being granted to the university.
  - Arguably, even when access to this data is granted to academic researchers this should be done via an interface whereby there is no direct access to the data, and only questions can be asked to a system which then processes the free-text data to return non-identifiable answers to the researcher.
  - An additional possibility is to curate the data such that some data from free-text information is converted and stored in structured fields.
- It is imperative to consider:
  - What possible valid and acceptable reason would a commercial organisation have for this kind of free-text information? Just because a commercial entity requests access to patient MH free-text data, that does not mean that it has a valid reason for access.
  - If there is a valid reason, do the costs outweigh the potential benefits? Even when there is a valid reason, that does not necessarily mean that access must be granted. There are various valid reasons for not approving all kinds of requests on a daily basis.
  - If there is a valid reason for access to this specific type of data, are there ways to

provide the limited information that the commercial organisation requires without providing direct access to patient MH free-text information? For example, partnership with a university, whereby a researcher at a university asks questions through an interface (i.e. does not read all free text) and a non-identifiable answer is returned after processing the free-text data, which the researcher can then share, as appropriate, with the commercial entity.
  - Is there really such an urgency to share unconsented patient MH free-text data with commercial entities (for apparent public benefits) that we cannot first ensure that access is restricted and granted only via a controlled interface and that too via a university?
- There needs to be clarity around and categorisation of the different types of MH data available, and clarity around what specific types of MH data can be shared with commercial entities, and clarity around what specific types of MH data will not be shared with commercial entities. There should be due consideration of the specific concerns that affected patients and the public have around the sharing of different types of MH data.
  - An overly simplistic approach which does not distinguish between different types of data, and essentially allows all data to potentially be shared, gives cause for concern. It strongly implies a lack of sufficient deliberation on the different types of data and the different concerns related to different types of data. It implies that the default position of sharing all data was taken without deliberation of whether all data should be shared, especially with commercial organisations.
  - Categorisation of different types of MH data available:
    - One categorisation is inevitably structured and free-text MH data. However, there may be other/further ways to categorise this data, based on patient and public concerns.
    - PPI in categorisation process.
  - More deliberative and relevant research to better understand patient concerns on the sharing of specific types of MH data:
    - Consider whether there can be in-depth, focused, deliberative consideration by patients and the public on sharing of free-text MH data, including examples of the kind of highly sensitive information such free-text data may contain. UPD states that citizen *"participation, particularly through deliberative methods such as citizens' juries, should form a substantial component of NHS efforts"* (11). Specifically, deliberative *"participation methods can help embed citizen engagement into governance mechanisms"* and involving *"people early on ensures decision-making is informed by public views, values, concerns and expectations"* (11). Arguably, due to the lack of deliberative research on this, patient views, and concerns on the sharing of MH free-text data have not been sufficiently explored and considered.
    - Deliberative research, such as citizens' juries, should also proactively involve organisations and persons who take a more cautious approach to health data sharing to ensure that jurors are able to consider a range of perspectives and to ensure that such research is truly unbiased.
    - Previous research on public views on the sharing of this data appears to be quite abstract and non-specific, and questions to the public often appear to be framed to evoke a "fear of missing out" (FOMO) on apparent, unquestionable, future, unspecified public benefits (e.g. x% *"of people in the study would rather commercial companies access data than miss out on research benefits"*) (16). Such wording could imply bias in the research. Aside from other significant limitations of such research, arguably, the public participants were not asked to consider, in detail, the kind of highly sensitive information such patient MH free-text data may contain.
    - Public participants who participated in previous research may never have disclosed highly sensitive information during psychotherapy and/or to MH services and may not be representative of the people affected by the sharing of such information:

- If public participants can self-identify as having a mental illness, without having received a clinical diagnosis, how representative are they of affected individuals?
- If participants have received a clinical diagnosis of mental illness, but never received intensive therapy or disclosed highly sensitive information during psychotherapy and/or to MH services, how representative are they of affected individuals?
- Would people who self-select to participate in such research likely have different views to affected patients who do not self-select?
- Would affected patients who have suffered different types of abuse and/or trauma have different perspectives on the sharing of MH free-text data?
- Is it acceptable for those who self-identify as having mental illness and those who have never disclosed highly sensitive and private information during psychotherapy and/or to MH services to talk on behalf of a group that is affected by the sharing of such information? For any other demographic, this would not be considered acceptable.
- A valid question is why researchers for prior studies seem to be indifferent about whether participants have shared highly sensitive and private information during psychotherapy and/or to MH services and whether participants would be directly impacted by the sharing of patient MH free-text data. Surely that is important to know before asking participants for their views?

- Subsequent decision-making, with patient involvement (especially affected patients), on what specific types of MH should be available for sharing:
  - There needs to be clarity on what types of MH data can be available and what types of MH data will not be available for sharing with commercial entities. All MH data does not have to be available for sharing. An overly simplistic approach which does not distinguish between different types of MH data, and allows all MH data to potentially be shared, gives cause for concern. It strongly implies a lack of sufficient deliberation of the different types of data and the different concerns related to different types of MH data. It implies that the default position of sharing all data was taken without deliberation on whether all data should be shared, especially with commercial entities. This may also raise concerns about whether there has been a lack of sufficient deliberation regarding other aspects.
  - Would different types of MH data potentially be available to organisations from different industry sectors?
  - Provide clarity online in an accessible way on what is and is not allowed.

**Motivation and purpose for data use:**

- UPD content often refers to patient data only being used where there is public benefit without defining what is meant by "public benefit" (13,14,16).
  - A lot of prior research on public views on patient data contains statements to the effect of "if there can be public benefit" or "if some lives can be saved, I would support patient data being shared" (23).
  - There are a lot of things that can provide public benefits and usually decisions are made by not only considering the "public benefits".
    - For example, cutting money laundering and fraud would provide "public benefit" including various potential MH benefits (e.g. fewer victims of fraud, more money available for public spending including on MH services). To cut money laundering and fraud, could we all provide government agencies access to our phones and bank accounts? It would not cost us anything, the data accessed would only be used for "public benefit" and the data would only be accessed by specific government agencies. Would the same people who support the sharing of all patient MH data to many third parties because it would apparently provide a "public benefit"

allow/approve government agency access to our phones and bank accounts? Why? Why not? Would they decline this because it would invade their privacy? By what reasoning is what they spend on groceries more private than patients' experiences of trauma or abuse? Would they decline this because data on their phone and in their bank account is *their* data, whereas extremely sensitive MH free text data does not relate to them at all and is data relating to *other* people?

- Arguably, "public benefit" is an elusive concept that requires further definition. Indeed, *"there isn't a shared understanding or consistent approach to evaluating public benefit, how to account for it in applications, and what (or who) we should draw on to define it"* (23).
- Note, further, that *"Public benefit is a necessary but not sufficient criteria"* (18). Furthermore, the willingness *"to share health records for the public benefit is not unconditional"* (18).
- Note that prior dialogues *"and surveys have tended to examine support for benefits to the NHS, rather than a more generalised conception of public benefit"* (18). Hence, it would be misleading and inaccurate to refer to public support for the sharing of patient data with commercial organisations where there is potential for public benefit *in general*.
- A report based on UPD commissioned research states that all access to patient data by commercial organisations must be for improvements to health and care (11,19). This is essentially a re-iteration of other findings, albeit stated slightly more precisely. Since any project can potentially be framed as a public benefit and many projects can potentially be framed as a way to improve health and care, there is a need for more guidance on this.
- UPD references a study on "public benefit" (18). A comprehensive review of this study and any subsequent resulting guidelines/standards may be appropriate, but unfortunately this was outside the scope of this review.
- Whilst understanding that all potential publicly beneficial uses of such data (resulting in improvements to health/care) cannot be foreseen, there is a need for specific guidance to help ensure that data is not misused and public trust is not abused. This can also help with assessing access requests, and enable greater transparency and accountability. During two 2017 citizens' juries, while members became more supportive of sharing patient data for public benefit in general, they were less supportive of sharing data for specific purposes (16). Hence the specific purpose matters and it would be more productive, transparent, and trustworthy to define a set of specific purposes for which MH data can potentially be shared.
- Currently, as per Principle 2 of the Code of Conduct, there is *"a requirement for data users to articulate what outcomes and benefits are anticipated from the development of a data-driven tool or technology"* (11). Currently, data may be shared where there is a health benefit and the NHS is prohibited from sharing data for *"solely commercial purposes"* (21). Arguably, a more specific purpose is required.
- The report based on UPD-commissioned research does provide further information on the purpose for which health data should be shared with third parties (11). The report states that jurors (in the citizens juries) felt that *"improved health outcomes should always be the primary purpose of using health data"* whereby improved health outcomes can constitute:
  - *"Direct impact on patient outcomes"*, e.g. new medicines/treatments, improved diagnostics, earlier disease detection.
  - *"Indirect impacts"*, e.g. *"providing additional revenue for the NHS"*, reducing costs and reducing waiting times (11).
    - However, this guidance is contradictory. On one hand, they state that the exploitation of *"health data solely for commercial gain"* would be contrary to public expectations, that improvements to health and care *"must be prioritised over financial incentives"* and that jurors (in citizens' juries) felt *"improved health outcomes should always be the primary purpose of using health data"* (11,19). On the other hand, they have a problematic definition of "improved health outcomes" which could lead to uses of health data solely for commercial gain.
    - Arguably, based on the above-mentioned definition of improved health outcomes, a project could meet the requirement for "improved health outcomes" by using the

health data for a range of purposes unrelated to improving actual health outcomes, if the project provides some "additional revenue for the NHS." That is, the definition frames "additional revenue for the NHS" as one possible "improved health outcome", i.e. a solely financial outcome would cease to be a solely commercial purpose.

    - The conditions or purposes for data use need to be more carefully worded in any guidelines to avoid such contradictions and "loopholes". Guideline wording must ensure carefully that data is only used for intended purposes (i.e. purposes supported by the public) and that there is consistent application across data access requests.
    - At times, the wording and tone in the report, suggest that patient data is viewed as being *"post-Brexit… valuable national assets"* (11). Such wording can encourage undue focus on financial benefits or revenue generation, encouraging (cash-strapped) NHS Trusts to take a more relaxed or accommodating approach to sharing patient data, which in turn may lead to practices which undermine public trust and potentially lead to misuse of patient data.

- Include guideline for data owner/host/controller to define an acceptable purpose for data access, preferably co-defined with patients and the public. All data access requests would be reviewed against the defined purpose.
- Include guideline for the defined purpose to be regularly reviewed with PPI.
- Include guideline that the defined purpose should require an improvement in health and care to be the primary purpose of any project requesting MH data access, whereby "improvements in health and care" should be clearly defined and exclude financial benefits. The improvement to health and care should not be a relatively minor or incidental purpose, nor a potential side effect of another primary purpose which is used to gain access to such data. Note, however, that the stipulation for a primary purpose which involves an improvement in health and care does not preclude indirect improvements in health and care rather than direct improvements. For example, a project that seeks to improve the health data infrastructure would not result in direct tangible patient benefits, but it would enable future improvements in health and care, and as such the primary purpose of the project would be improvements in health and care. That is also not to say that data partnerships should not generate revenue for the NHS.
- Include guideline for appropriate independent experts (including in the subject matter of the respective project) to review each data access request. It should not be taken for granted that appropriate independent experts will review data access requests. Such experts to determine the following (among other determining factors):
    - whether proposed work would potentially result in an improvement in health and care;
    - the relative importance of the work for health and care;
    - the viability of the proposed project;
    - whether this work has already been done;
    - whether important aspects have been omitted;
    - whether the data requested is necessary for the work;
    - whether the data requested would result in bias and whether different or additional data might be necessary to address concerns of bias;
    - what data and the minimum amount of data necessary for the project.
- Include a guideline stipulating that once a defined purpose is developed, the data controller uses *"a case-by-case approach to public benefit assessment so that the definition can be tested against potential unforeseen consequences or harms to individuals and society"* (17,18). This is important and possibly a step that is often omitted. This guidance is not in the report on the commissioned research, but is in non-UPD research referenced by UPD elsewhere. Obviously, if unforeseen harmful consequences are identified, the defined purpose should be amended and re-tested.
- While the primary purpose of a project (for which there is a data access request) should be

an improvement in health and care, it is recognised that tangible or visible patient benefits may not always be directly (and immediately) realised, for a number of reasons (mentioned below) (11). One broad reason for why tangible patient benefits may not be directly realised may be that benefits are indirect and expected over the longer-term due to the nature of the research. Another broad reason for why tangible patient benefits may not be directly realised may be failure of the project or failure to utilise research outputs. There is a distinction between the below-mentioned cases (i.e. where planned benefits are not directly realised) and projects for which the primary purpose is not an improvement in health and care from the outset. Granting access to patient data for the latter would arguably undermine public trust.

- *Indirect and long-term benefits expected:*
  - Some early stage or exploratory research may not result in tangible improvements for patients, e.g. novel data uses and data-driven technologies (11).
  - It may be a long process to *"prove the clinical utility and effectiveness of even highly promising algorithms"* (11).
  - Some projects may focus on improving the health data infrastructure, which would not lead to immediate patient improvements, but which would nonetheless be important for enabling future improvement in health and care, e.g. *"some data partnerships may need to focus on data curation"* (11). In such projects, the primary purpose would still be an improvement in health and care, even though such improvements would be indirect and realised in the future.

- *Project failure or failure to utilise project outputs:*
  - Issues with integrating new technology outputs (from projects) into the NHS (11).
    - For projects where there are tangible and implementable project outputs, the failure to implement these within the NHS ultimately means a failure to realise patient benefits. This does not mean that the NHS should implement all such project outputs. Indeed, there may be many cases where there are good and valid reasons for not implementing such outputs. Instead, this means that it may not be in the public interest for access to patient data to be granted for projects for which there is a high likelihood of outputs not being implemented by the NHS.
    - To reduce the risk of this, should every data access request proposing implementable outputs be required to conduct a preliminary organisational readiness or acceptability assessment of the NHS's readiness or willingness to implement proposed outputs?
      - If so, this assessment should be completed with the appropriate NHS body, but without involvement of the data host/controller to avoid any conflict of interest.
      - There should be a recommendation for the appropriate NHS commissioning body to have a preliminary NHS acceptability assessment process. This should not be as long or detailed as the standard process for new healthcare innovations, and it might not be appropriate for it to be legally binding. The process may need to consider the acceptability of the project outputs at the projected date of availability rather than the immediate acceptability. The guideline should include some explanation on the rationale behind it.
      - Such a preliminary acceptability assessment may help identify project outputs which are highly unlikely to be implemented in the NHS.
      - For data access requests proposing implementable outputs, the data access review process should consider the score/result from the NHS acceptability assessment, and if the result indicates that the project outputs are unlikely to be implemented within the NHS, it may be advisable to reject the access request.
    - Arguably, it is irrelevant if a project claims it will provide public benefit if the project outputs are unlikely to be implemented within the NHS anyway. Note that

prior dialogues *"and surveys have tended to examine support for benefits to the NHS, rather than a more generalised conception of public benefit"* (18). Hence, it would be misleading and inaccurate to refer to public support for the sharing of patient data with commercial organisations where there is a general potential for public benefit.

- Various practical challenges with the project;
  - Given that private patient data would be shared and used, there should be an effort to minimise risk of project failure or failure to utilise research outputs. It is notable and concerning that over 85% of data science projects apparently fail. In cases where patient data is used this should not be in vain, i.e. such private data should not be shared without taking appropriate and adequate measures towards minimising the failure of the project, otherwise it is arguably not ethical to share such data.
  - Some measures/guidelines to minimise risk of failure are proposed in this review: (a) an NHS preliminary acceptability assessment, for relevant projects; (b) project progress reports (see below). More consideration is needed on other appropriate measures.
- The research findings.

- Issues such as this can make it difficult to determine the potential public and patient benefits for a data access request, which can make it difficult to assess such requests fairly. This has to be duly considered in any guidelines.
- The report categorises health data use into three broad categories:
  - *"Broadly acceptable use cases with clear public benefit, minimal privacy risks and low risk of controversy;*
  - *Grey area/mid-range use cases, for which there will be a diversity of views and perspectives;*
  - *'Red line' use cases highly likely to be considered unacceptable, such as using data for insurance or marketing purposes, or clear commercial exploitation"* (11).
    - Given the high sensitivity of MH data, are these broad categories sufficient? For example, for MH data, there may be fewer cases that fall into the first "broadly acceptable" category and there may be a need for more categorisation and guidance on "mid-range" or "grey" cases. See also comments above on the sharing of unconsented MH free-text data.
    - Given the above-mentioned difficulties in assessing data requests, especially when potential patient/public benefits are not direct, immediate and/or tangible, there is a need for more guidance on:
      - How to determine whether the primary purpose of a project is an improvement in health and care;
      - The distinction between projects for which the primary purpose is not an improvement in health and care and those for which improvements in health and care may be indirect and/or long-term outcomes. There is a need for information on and examples of the latter, and how such projects may enable or lead to improvements over the longer term, e.g. how/why data formatting, standardisation and interoperability might lead to health/care improvements over the long term.
- Prompt questions and/or a template may be useful for reviewers to use while determining whether an access request meets the defined purpose for data access.
- Access requests that fall outside the defined purpose, but that potentially have "public benefit" can be considered within a separate formal approval process.

**Transparency and "meaningful transparency":**

- Based on the UPD content reviewed, there is no explicit guidance on implementing a public registry of approved data access requests. Although, UPD refers to the fact that the NHS already *"publishes details of every organisation that uses NHS data"* no detail is provided on how to access this information (21). This reviewer was unable to check and verify this.
    - If more patient data will be shared with third parties in coming years, and if decisions on commercial data partnerships will be made by local host organisations, there may be need for more guidance on how this information can be reliably tracked. If there is currently a central publication or registry of "every organisation that uses NHS data" how is this information currently tracked and centrally updated? What established procedures exist?
    - Include guideline for each host organisation to establish and maintain an (online) public registry of approved data requests. Information in public registry to be in plain language and accessible. Information provided on public registry to include:
        - Name (registered and trading name) of commercial organisation gaining data access;
        - Industry sector(s) that organisation operates in;
        - Main activities of the organisation requesting data access;
        - Details of the research project for which data access requested;
        - Whether access request was reviewed by patient/public reviewers, and if not, why not;
        - Whether approved data access requests were supported by patient/public reviewers, and if not, why not;
        - Details of what data was shared—how much data was shared, what was included in the data, individual or aggregate, identifiability of data, structured data or free text;
        - How data is accessed, information on how this is secure;
        - Duration of data access;
        - Details of conditions imposed for data access and use;
        - Details of any reviews/monitoring of data use and compliance with conditions of use;
        - Details of any breaches in conditions (including any data misuse) and actions taken;
        - Details of any security and privacy breaches and actions taken;
        - Information on research outputs.
    - Given the specific sensitivity of MH free-text data, each registry entry should clarify whether MH free-text data was accessed, why it was necessary for the project in question, and how it was accessed (i.e. via an interface that is only used by an academic researcher?). This would ensure public scrutiny of the sharing of MH free-text data and ensure that such information is not just shared by default, and without appropriate and valid reasons. It may also provide an opportunity for the public to better understand why this kind of data may be useful for commercial organisations in specific contexts, and what stringent controls are in place to prevent direct access by commercial entities. As mentioned previously, patient MH free-text data should not be shared directly with commercial entities. See comments above on the sharing of unconsented MH free-text data.
- Currently, there is a lack of transparency over terms negotiated for data partnership agreements, which allows companies to *"approach multiple NHS providers with terms favourable to them"* (11). Arguably, the terms negotiated for each data partnership should also be made available on the public registry. It might be appropriate for the full terms and a plain language summary of the terms to be provided on the public registry.
- UPD states that *"Decisions about third party access to NHS data should go through a transparent process and be subject to external oversight"* (11,19). Previous research has found that the public want external independent oversight, including on the purposes for

which data can be used (14,15). UPD also refers to prior research indicating that the public want to have a *"say in how patient data is used, so collective mechanisms oversight are important"* (13). Beyond the use of PPI in assessing access requests to achieve "collective mechanisms [of] oversight" there is a lack of clarity on what other "external oversight" there should be. UPD refers to the establishment of a *"single point of guidance and oversight"* in order *"to set consistent rules across the NHS for third party access to data"* (19).

  - More clarity is needed on what specifically should be subject to external oversight, e.g. should local data access processes be subject to an external and independent audit? Should this check whether the data host organisation's documented data access process was properly followed or whether data access should have been granted? In other words, should "compliance with procedures" be audited or should "the appropriateness of decisions to award data access" be audited? Should the way in which data access processes are designed be subject to review and oversight?
  - More clarity is needed on how such external oversight should be conducted in practice.
  - Please see below also for further details regarding oversight and governance.

**Competence:**

- UPD states that organisations should ensure that staff working with the data have the skills, *"expertise, competencies, tools and methodologies"* to do this safely, securely, effectively, only for intended purposes, and to complete the proposed research (13).
  - It is appropriate to include this in the MH data guidelines. Regardless of the implementation of other technical and procedural controls within the system, taking measures to also ensure staff competence provides greater assurance and increases the trustworthiness of the system.
  - Note, however, that UPD is referring generally to anyone using patient data, rather than commercial entities specifically. See Data Access Process (below) for staff competence considerations for commercial applicants.
  - When considering staff competence within the data host/controller organisation UPD refers to concerns that have been expressed about lack of:
    - *"in-house technical capacity and expertise"* within the NHS to effectively manage risks, and to effectively and safely use and share data (16).
    - NHS in-house understanding and skills to negotiate fair terms for data partnership agreements with commercial entities and how this may result in various issues including commercial exploitation of patient data (11,15).
  - Thus, UPD states that the public *"place a high priority on independent oversight, transparency, and accountability"* (16). An overarching governance framework, oversight organisation(s), a *"coordinated national strategy and guidance"*, monitoring activities, appropriate reactive activities for issues, free expert advice, and a central registry, may help to provide oversight, transparency, and accountability (11,19). See also "Governance and Monitoring" (below) for further details.
- However, whilst independent monitoring may retrospectively find issues, it is also important to reduce the occurrence of issues due to lack of competence. UPD makes no explicit mention of appropriate staff (and public/patient reviewers) at the host organisation being:
  - Trained on the various data access processes (data access process, process for change(s) to data access and/or use, monitoring/review processes, processes to respond to breaches/misuse, plus more);
  - Trained on criteria to consider when making assessments;
  - Provided with documented data access processes.
- Staff competence can also include professional ethics and ethical conduct in general. This is not explicitly mentioned by UPD.
- Measures should be taken to ensure ethical conduct of staff at the host organisation. This may include:

    - Appropriate vetting of staff;
    - Regular ethics training;
    - Promptly removing staff for whom concerns have been expressed from roles which involve the sharing or oversight of patient data.
- Measures should be taken to ensure ethical conduct of staff at commercial entities that gain access to patient data. Even if patient MH free-text data is not shared with commercial entities, structured MH data is still highly sensitive. Therefore, precautions should be taken to ensure that staff that gain access to this data can be trusted. Measures taken may include:
    - Requiring disclosure of relevant concerns or complaints against staff on data access applications;
    - Requiring disclosure of relevant concerns or complaints against staff who have already been granted data access;
    - Strong actions/penalties against staff who are found to act inappropriately with any patient data.

**Data access processes:**

*Requirements for the data access process*

- Beyond ensuring PPI in assessing data access requests, UPD does not mention specific requirements for the data access process.
- UPD does not indicate what information commercial applicants for patient data access should be required to provide on applications. Some of the information required from commercial applicants on data access applications may include (among other details):
    - Lead applicant details;
    - Name (registered and trading name) of commercial organisation requesting data access;
    - Industry sector(s) that organisation operates in;
    - Main activities of the organisation requesting data access;
    - Details of other organisations (where multiple organisations working on project);
    - Details of the research project for which data access requested;
    - Prior experience relevant to research project for which data is being requested;
    - Study plan and methodology;
    - Expected outputs and impact;
    - Data requirements; Details of what data is being requested, how much (volume of) data requested and why;
    - Duration of data access requested; research study start and end dates;
    - Names, job titles and experience of staff (at commercial entity or entities) for whom data access is requested;
    - Plain language summary of research project, including project aim and objectives, expected outputs and impact, what data is requested and why;
    - Anticipated risks and mitigation plan;
    - Security measures. Note that undue reliance on the commercial entity to ensure data security should be limited via the use of an SDE and various technical and procedural controls—see other comments on this below.
- While UPD refers to ensuring that the staff working with the data have the skills, *"expertise, competencies, tools and methodologies"* to do this safely, securely, effectively, only for intended/approved purposes, and to complete the research they are setting out to do, it does not detail how this could be ensured for staff at commercial organisations (13).
    - Regardless of other technical and procedural controls within the system, taking measures to also ensure staff competence provides greater assurance and increases the

trustworthiness of the system.

  - Note UPD is referring generally to anyone using patient data, rather than commercial entities specifically.
  - In the case of commercial data access applicants, the host organisation's data access process could require that the commercial entity names the staff who will work with the data on the data access application. The job titles and experience of the staff could be specified, with the host organisation then vetting named staff.
  - Even if patient MH free-text data is not shared with commercial entities, structured MH data is still highly sensitive. Therefore, precautions should be taken to ensure ethical conduct and that staff at commercial entities that gain access to this data can be trusted. Measures taken may include:
    - Requiring disclosure of relevant concerns or complaints against staff (for whom data access is requested) on data access applications;
    - Requiring timely disclosure of relevant concerns or complaints against staff who have already been granted data access;
    - Strong actions/penalties against staff who are found to act inappropriately with any patient data.
- The capacity of the host organisation to thoroughly vet commercial entity staff may be limited, and there will be some reliance on the commercial organisation to ensure that staff who are granted access to the data are appropriate and can be trusted. It may be appropriate to include wording to this effect in the terms of agreement for the data partnership, with data access being conditional on the terms being upheld.
- Technical and procedural controls can be implemented to ensure that undue reliance is not placed on staff (competence) at commercial organisations (see "Elements worth including", and above and below in "Elements worth including with amendments", for further comments).
  - Staff at commercial entities do not have to be granted access to all types of MH data. For certain types of highly sensitive and private MH data (e.g. patient free-text MH data) access does not have to be granted to commercial entities. If there is a good and valid reason for accessing such data, one solution may be to require commercial organisations to conduct the research in collaboration with a university, with access to the MH free-text data only being granted to the university. Even when access to such data is granted to academic researchers this should be done via an interface whereby there is no direct access to the data, and only questions can be asked to a system which then processes the free-text MH data to return non-identifiable answers to the researcher. See above for further comments.
- UPD does not mention what should occur if an applicant is found to have provided false information on their data access application form either at assessment phase or after gaining data access. See Accountability below.
- While UPD refers to the need for the data access process to be transparent, it does not explicitly recommend that the end-to-end data access process should be documented, including in an accessible format, and be made publicly available (online). It would be appropriate to include an MH data guideline stating this.

*PPI in assessing data access requests*

- There should be public involvement in *"decisions about third party access to NHS data"* which is backed by survey results and citizen juries (11,19). However, the report (based on UPD-commissioned research) suggests taking a proportionate approach, categorising health data use into three broad categories:
  - *"Broadly acceptable use cases with clear public benefit, minimal privacy risks and low risk of controversy;*
  - *Grey area/mid-range use cases, for which there will be a diversity of views and*

*perspectives;*
    - *'Red line' use cases highly likely to be considered unacceptable, such as using data for insurance or marketing purposes, or clear commercial exploitation"* (11).
- The report argues that taking *"a proportionate approach might mean focusing citizen involvement on cases in the 'grey area'"* (11).
    - Arguably, with regards to unconsented MH data, all data requests should be reviewed by patient/public reviewers. If there is a high volume of requests, there should be a correspondingly high number of patient/public representatives.
    - Public support for sharing data depends on the nature of the specific research (16). During two 2017 citizens' juries, while members became more supportive of sharing patient data for public benefit in general, they were less supportive of sharing data for specific purposes (16). Furthermore, whether a specific organisation is trusted by members of the public is an important consideration in how the public feels about sharing health data (16). This suggests that it would be important for all MH data access requests to be reviewed by patients and members of the public.
    - Deciding that certain data requests do not have to be reviewed by patient/public reviewers is problematic and raises concerns about how this would be decided.
        - Who decides the specific definition of each category? i.e. what is a "grey area" category?
        - Who decides how cases are allocated to categories, and which cases belong to which categories?
        - If all data access requests will not be reviewed by patient/public reviewers, and if only access requests in certain "use case" categories will be reviewed by patient/public reviewers then:
            - There should be PPI in determining the "use case" categories and the method/process for allocating cases to categories.
            - There should be public transparency on how the determination is made.
            - There should be regular PPI in reviewing the categories and the method/process for allocating cases to categories.
- Prior research is limited and there *"is limited evidence about how people would weigh up the benefits and risks for the kinds of data access agreements NHS bodies are making"* (11).
    - Greater transparency on what benefits and risks were considered may be appropriate;
    - Regular feedback sessions on patient and public reviewers' experiences may be helpful;
    - Important to proactively consider how patient and public involvement can add value rather than be a tick-boxing exercise;
    - The methods, processes, and tools (e.g. prompts, templates, forms) for conducting such reviews should be regularly reviewed, and updated, as appropriate.

*Impact on health inequalities to be considered when assessing data access requests*

- When considering partnerships between the NHS and third parties, UPD-commissioned research found public perspectives such as improvements to health and care *"must aim to improve health and care for everyone"*; improvements to health and care must be *"distributed across the country, to prevent making health inequalities worse"*; benefits should be shared across the population and *"avoid making health inequalities worse"*; improvements *"should be fairly distributed across the NHS"* (11,15,19). UPD does not sufficiently analyse and question such perspectives. While it is important to consider impact on health inequalities, the above specified wording is problematic:
    - Potential impact on health inequalities should be considered when assessing data access requests.
    - Note there are many health inequalities related to aspects other than geographical location. Hence it is inappropriate and insufficient to only consider geographical

location when attempting to address health inequalities.

- More thought is required on the intended and expressed meaning. There is a difference between equality and equity which is not appropriately recognised in the statements. There is a need for more careful wording, with greater clarity, in statements about the impact on health inequalities.
- It is not appropriate or realistic for every project requesting data access to aim to improve *"health and care for everyone"*.
  - To tackle health inequalities, it is necessary to focus on those who are adversely affected; resources and improvements will have to be distributed unequally, with more directed at those who are adversely affected, to reduce health inequalities and to ultimately strive for equal health outcomes. Therefore, for projects that aim to tackle an aspect of existing health inequalities, the benefits from such partnerships will not be equally distributed across the NHS or country.
  - Projects will be necessarily focused on specific solutions for specific health challenges. Hence any given project cannot possibly *"improve health and care for everyone"*.
  - Some projects may focus on rare diseases or illnesses affecting certain demographics. Note also that certain demographics may well be concentrated in certain geographic areas. In such cases, the benefits from such data partnerships will not be equally distributed across the NHS or country.
- It would be inappropriate and limiting to consider whether benefits only pertain to a local area or demographic, and it would instead be more appropriate and fairer to consider the potential impact of the data partnership on existing health inequalities: will the data partnership likely exacerbate existing health inequalities, have no difference, or reduce health inequalities? Important considerations (among others) for projects making data access requests:
  - Bias: What will be done to reduce bias? This is especially a concern for AI projects.
  - Affected demographics: What are the affected demographics for the illness being researched by the project? Does the project design, methodology or approach exclude or not sufficiently consider any affected demographic groups? How well does the requested data match the affected demographics?
- Currently, there is insufficient evidence on the actual impact of data partnerships on health and care. *"Monitoring and evaluation will be critical to ensure that they are not making health inequalities worse or increasing regional disparities in skills, expertise and data use"* (11). See also comments for monitoring (below).
- In the UPD-commissioned research, there were suggestions by several jurors (and via the survey) that decision making about data partnerships should occur at *"national level to ensure an even distribution of outcomes, benefits and rewards across the country"* (11). Concerns were raised about benefits from data partnerships not being fairly distributed across the NHS and across the country. In particular, concerns were raised about poorer and rural areas being left behind or excluded, e.g. some technologies may be developed without due consideration of such populations. However, the UPD report argues that local regions are *"best placed to make decisions for their local populations, and disease specialists best placed to make decisions for the communities under their care."* Therefore, there are clear advantages to local decision-making. However, it is important for all decision-making regarding data partnerships to take account of fair distribution of benefits and to ensure that such partnerships do not result in further health inequalities.

*Formal process for change(s) in data access and/or use*

- UPDs' comments on modification to data access and use imply a lack of sufficient consideration of the risks. UPD says that any *"major alterations in purpose or scope should trigger the data applicant to go back to the data assessor as a minimum to acknowledge the* *change or to seek approval to proceed on the basis of the new focus for the work"* (18).

- It would be inappropriate for the commercial entity to simply inform the data assessor/owner of the change in purpose or scope (albeit as a minimum). Just because the original purpose or scope was approved, it should not be assumed that any changes in purpose or scope would be approved.
- Beyond changes in purpose or scope, there may be other changes including (but not limited to) changes in what data is required, the duration for which it is required, how the data is used (even if the purpose remains the same) and which staff require access to the data. All such changes would be changes in data access and/or use.
- Any such changes should not be requested or approved informally. UPD's statements on data access/use modification imply a lack of recognition of the importance of a formal process. Any changes to data access and data use should be handled via formal processes. That is, the commercial organisation should be required to formally request modified data access and/or use and this should be subject to formal approval.
- Process(es) for change(s) in data access and/or use should be clearly documented, including in accessible format, and publicly available.
- There should be other procedural and technical controls in place to ensure that commercial organisations are severely restricted in modifying data access or data use without formal approval. This may take the form of technical controls in Secure Data Environments (SDEs) such as access only being permitted for a specific subset of a dataset. There may be regular review, monitoring, or audit processes to check what and how data is being accessed and used. There should be clearly defined formal processes, with clearly defined consequences, for addressing any unapproved data access or use.
- UPD's statements also imply that changes in data access and use would be approved. There is almost a presumption of retrospective approval for changes in project scope. Just because the original data access application was approved, it should not be assumed that any changes in data access/use will be approved. NHS Trusts/other data owners of patient data should not feel obligated, coerced, or forced into approving modified data access just because a project has already commenced. Data owners should not be liable for any losses incurred by the commercial entity if modified access requests are not approved.
  - It may be appropriate for data hosts to explicitly state this in data access agreements or contracts agreed *prior* to granting any data access. This could include wording to the effect of:
    - Data access and use is only granted on the basis of the initial approved application and agreed terms of access and use;
    - Commercial organisations are required to formally request modified data access or use (if required) via the referenced formal processes;
    - If the commercial organisation is found to engage in unapproved data access or use, they will be subject to the relevant formal process, with potential removal of all data access;
    - Commercial organisation should be aware that there is no guarantee of applications for modified data access or use being approved and projects should not be commenced on the presumption that (subsequent) requests for access/use modifications will be approved;
    - If applications for modified data access or use are not approved, the data host/controller is not liable for any losses incurred by the commercial entity.
  - Unless this is clearly stated from the outset, and implemented, by the data host/controller, there is a risk of commercial entities intentionally mis-stating their requirements or intentions in the initial data access application to gain data access with the understanding that modifications will be granted later. Some commercial organisations may game the system to gain certain levels of access. Unless there are appropriate contractual clauses, and procedural and technical controls in place, such

commercial organisations may end up (through (repeated) subsequent data access modification application(s)) gaining data access and using data in a way which would never have been approved if it had been requested in the original data access application.

*Commercial organisations prohibited from sharing data with third-party organisations*

- Currently, prior to getting data access, companies sign a contract which *"usually"* states *"that data cannot be passed to any third parties, unless explicitly approved in the application"* (20). UPD provides the example of some analytics companies that *"work with a number of different clients and may want to use the same data with different organisations"* (20). Apparently, such companies may be allowed to share data with third parties *"if it has been specifically agreed"* and the purpose has been approved in the application (20).
  - Based on the use of the word "usually," it appears that such contracts do not always prohibit companies from sharing patient data with any third parties. Thus, this should be explicitly stipulated in the guidelines for MH data.
  - Arguably, rather than allowing companies to share such data with (unnamed) third parties, if access is required by multiple organisations, for a single project, this should be explicitly requested in the application and access should be approved, as appropriate. This would allow all third-party organisations who request access to such data to be appropriately vetted by the host organisation, with PPI.
    - The formal data access process should allow for access to be requested for multiple organisations, as appropriate;
    - The data access application assessment process should vet each named organisation on each application;
    - If data access is approved for a project, but not for certain organisations on the application there should be mechanisms in place to handle this and ensure that those (unapproved) organisations do not gain access..
  - If there is a need for data to be shared with a third-party after the project has already commenced, the formal process for modification to access should be followed.
  - MH data guidelines should stipulate that all access to MH data should be via SDEs which would make it technically more difficult for companies to share data with third parties.

Other aspects of data access:

*Secure data transfer/access and storage*

- Currently, commercial organisations are required to store patient data securely, *"with controlled access and robust IT systems to keep data safe, and there are strong sanctions if data is misused"* (20). Arguably:
  - Commercial entities should not have sole responsibility for secure storage of patient data.
  - This may result in the use of insecure methods of data transfer to the commercial entity.
  - Access to such data should be via secure environments, which would address issues of both insecure data transfer and storage.
  - UPD notes that the NHS is moving away from data transfer to third parties *"to a system of data access where NHS data is stored in a Secure Data Environment (SDE) that can be accessed by approved users but data cannot leave the platform"* (20).
  - For MH data, especially, there should not be direct transfer of data and access should be via an SDE only.
  - Within the SDE there should be access controls to control who has access to the data,

when and how. While a combination of technical and procedural controls may be used, where possible, it may be appropriate to implement technical controls so that undue reliance is not placed on people not doing the wrong things or not making mistakes.

- Even when access is via an SDE, and there are various technical and procedural controls, companies should still be required to ensure that only authorised staff have access, that data is not misused, and that all aspects of the access agreement are upheld.

Governance and monitoring:

*Oversight for defined purpose(s) for data sharing*

- *"Someone has responsibility for oversight about the purposes for which data can be used"* (14).
  - It would be appropriate for a designated person or panel to be responsible for overseeing the overall purpose(s) (i.e. the defined purpose(s)) for which data can be shared. This individual/job role and/or panel should be named on data access process documentation, which should be made publicly available.
  - However, arguably sufficient detail has not been provided by UPD on what responsibilities this role/panel would have, on the level of PPI for this oversight or other requirements:
    - If a panel has oversight for this, there should be patient/public contributors on the panel.
    - The roles and responsibilities for this role/panel should be clearly defined and documented, and made publicly available.
    - This individual/panel may be responsible for initially defining the purpose(s) for data access. This should be done with PPI.
    - The defined purpose(s) should be clearly documented and made publicly available.
    - All processes associated with oversight of the defined purpose should be formally defined, documented, and publicly available.
    - They should use *"a case-by-case approach to public benefit assessment so that the definition can be tested against potential unforeseen consequences or harms to individuals and society"* (17,18). This should be repeated every time the defined purpose(s) is changed.
    - They should have oversight for ensuring that approved data requests are in line with the defined purpose(s) for data access and that any exceptions are appropriate.
    - They should have responsibility for periodically reviewing the defined purpose(s).
    - Please see above "Motivation and Purpose for Data Use" for further comments on defined purpose(s) for data access.

*Project progress reports:*

- Publish *"transparent and publicly accessible updates on the progress of the partnership towards achieving predicted benefits"* (11).
  - There should be a requirement to provide project progress updates, and this can be one way to help ensure that the data is used for the approved purpose. This will help ensure greater transparency and accountability. This may also enable the system to be more trustworthy and provide reassurance to the public that there are formal mechanisms to check that the data is being used for the approved purpose.
  - It is notable and concerning that over 85% of data science projects apparently fail. In cases where patient data is used this should not be in vain, i.e. such private data should not be shared without taking appropriate and adequate measures towards minimising the risk of failure of the project. A requirement to produce project progress reports is one way to check that the project is on track to achieve its objectives.

  - There should be a minimum of one project progress report per year, or a minimum of one project progress report per partnership if the project duration is for less than one year. The specific frequency of progress updates needs to be mutually agreed during the application process (as this may vary by project) and this should be mentioned in the respective guideline.
  - The specific requirements to produce project progress reports, including the frequency and contents of such reports, to be explicitly stated in every data partnership agreement.
  - Project progress reports should be subject to PPI review.
  - Project progress reports to be written in plain language. Producing plain language reports would take work and require sufficient funding, and so from a financial sustainability perspective, it would be appropriate to place this requirement on the commercial entities rather than the data controller. Furthermore, the commercial organisations would be better placed to explain their projects in plain language.
  - The content of such reports may include (among other aspects):
    - Project summary;
    - Project plan with stages/phases/milestones, including what milestones have been achieved so far;
    - How the data has been used;
    - Whether the project is on course for completion as per the application;
    - Whether the anticipated benefits are as expected;
    - Any notable issues that have occurred and how these were addressed;
    - Any further anticipated risks (for the remainder of the project) and mitigation plans for identified risks.
  - There will need to be appropriate allocation of resources to ensure that progress reports can be properly reviewed and actions taken, as appropriate.
  - Will need an additional guideline concerning the review of progress reports.
  - Key information in plain language, after redacting confidential information and with the commercial organisation's agreement, should be provided on the public registry, for greater transparency.

*Data Partnership Agreement/Contract Terms:*

- Currently, companies *"have to sign contracts that set out what they can and cannot do with the data"* (20). Inevitably, there are other considerations for such agreements beyond data use, e.g. agreed monitoring, audits and reviews, terms of payment.
- UPD does not specify requirements for the terms of data partnership agreements.
- MH data guidelines should clearly stipulate key requirements for the process of determining the terms and the terms themselves. Requirements for the process of determining the terms may include that the data host organisation seek expert advice beforehand and comply with national guidance (see "Governance and Monitoring" below). The terms themselves may include the following (among other aspects):
  - Approved purpose for data access;
  - That staff gaining access to the data at the commercial entity have been appropriately vetted by the commercial entity;
  - That the commercial entity will produce project progress reports at agreed intervals;
  - That the commercial entity agrees to specified monitoring, audits, and reviews;
  - List of prohibited actions, including a prohibition on making any attempt to identify any individuals from the data;
  - That if the project scope, or data access/use requirements change, the appropriate formal process for modification of data access and use will be followed;
  - That there is no guarantee of applications for modified data access or use being

approved and projects should not be commenced on the presumption that (subsequent) requests for access/use modifications will be approved. If applications for modified data access or use are not approved, the data host/controller is not liable for any losses incurred by the commercial entity;
  - Terms regarding payments;
  - What will occur in the event of data misuse or a breach of any of the terms.
- Payments: Currently the NHS *"is not allowed to sell data for profit but operates on a cost recovery basis"* which means it *"is allowed to charge for the cost of processing and delivering the service, but not for data itself"* (20,21). This charge can vary and individual *"NHS Trusts will enter into different arrangements when working in partnership with companies, depending on their requirements and the services that are offered"* (20). There needs to be more discussion on how the NHS and patients can benefit from this resource (20). NHS England's recent Value Sharing Framework (July 2023) states that *"the NHS should seek a share of commercial value arising from the use of NHS data, proportional to the NHS's contribution to the project"* (20). Arguably, more detail is needed on how this would work in practice and what other payment structures should be considered.

*Overarching governance framework and oversight organisation(s) for patient data use*

- Concerns and issues include:
  - Research has found that the public is concerned about exploitation of the NHS by commercial organisations and believes that NHS organisations require consistent *"support and guidance to negotiate fair terms for agreements with third parties"* (11,19).
  - There are public concerns about focus on *"short-term revenues at a local level"* while neglecting *"long-term potential benefits for future generations across the health system"* (11).
  - NHS trusts may not have in-house understanding and skills to negotiate fair terms;
  - Lack of NHS capability may result in lack of fairness between data access applicants as larger companies may be better placed to negotiate more favourable terms;
  - Important to ensure that *"partnerships between NHS and third parties are, and are seen to be, fair"* (11). Therefore, it is important to ensure that fairness is achieved in actuality and appearance.
  - There is a need to consider value more broadly (beyond monetary value) for all parties in the context of such agreements;
  - Currently, there is a lack of transparency over terms negotiated for data partnership agreements, which allows companies to *"approach multiple NHS providers with terms favourable to them"* (11).
- The following may help to address the above-mentioned concerns:
  - *"A single point of guidance and oversight should be established to set consistent rules across the NHS for third-party access to data"* (19).
    - A *"coordinated national strategy and guidance"* (11);
    - *"Consistent rules across the NHS for third-party access to data"* (11);
    - Implement *"a clear, overarching governance framework for NHS health data use"* (11). There could be a single governance and oversight organisation or this function could be *"spread across governance and oversight structures including the National Data Guardian, existing regulators and the new National Centre of Expertise"* (11). Even if this function is carried out by various local/regional bodies, *"all data partnerships must adhere to the same standards of accountability"* (11).
  - National-level governance and oversight for data partnerships (which may be implemented by local/regional organisations) and this should include:
    - *"Proactive activities: establishing principles and ensuring a good governance framework;"*

- The report recommends building *"sustainable mechanisms for embedding citizen views and values into its guidance and advice as it changes over time"* (11). This is important. However, there is no mention of PPI in developing and establishing the initial principles, guidance, and advice. There should be PPI in establishing initial national principles.
- There is no mention of PPI in designing and establishing the governance framework and this work should incorporate PPI.
- There is no mention of dissemination activities and ensuring staff at data host organisations are adequately informed, which should be ensured.

▪ *"Monitoring activities: auditing and reporting"* (11).

- There is a lack of clarity on what specifically should be subject to external monitoring and how such monitoring should be conducted in practice.
- Should local data access processes be subject to an external and independent audit? Should this check:
  - How the data access process was designed? (who was involved, what was considered, whether there was PPI).
  - The design of the data access process? What steps and controls it contains? i.e. should the fairness and appropriateness of the data access approvals process be subject to external independent monitoring and oversight?
  - Whether the data host organisation's documented data access process was properly followed? i.e. should the consistent and fair implementation of the data access approvals process be subject to external independent monitoring and oversight?
  - Whether data access should have been granted? i.e. should actual decisions about granting data access be subject to external independent monitoring and oversight?
- What about an external review of terms negotiated for data partnership agreements?
- Should the "defined purpose" for which data can potentially be shared, set locally by each data host organisation, be subject to external monitoring?
- Should patient and public concerns and views help determine what should be monitored regularly?
- What would be monitored to check compliance with existing regulations and what organisations would be responsible for conducting such monitoring?
- One specific monitoring activity mentioned by UPD is *"reviewing, auditing and evaluating NHS data partnerships, to assess whether they are delivering the anticipated benefits"* (11). There is a lack of clarity on whether such monitoring would check whether a given project achieves its objectives (i.e. the benefits) or whether the benefits are realised within the NHS. The latter would involve checking whether project outputs have been successfully implemented within the NHS and what the impact of the implementation is. This is important because currently, there is insufficient evidence on the impact of data partnerships on health and care.
- For projects delivering benefits over the longer term or projects delivering indirect benefits, how would this be measured in practice?
- There is no mention of PPI in monitoring activities.
- *"Monitoring and evaluation will be critical to ensure that they are not making health inequalities worse or increasing regional disparities in skills, expertise and data use"* (11). Thus, it is important that monitoring includes a review of the impact on health inequalities. Currently, there is insufficient evidence on the impact of data partnerships on health and care.
  - While the report states that the *"Academic Health Sciences Networks and*

*the Health Data Research UK Health Data Hubs are well-placed to understand where data partnerships are emerging at local and regional levels"*, it does not explicitly recommend national-level monitoring and evaluation.

- Arguably, while decision-making on data partnerships should occur at a local level, monitoring, and evaluation (of impact) should occur centrally at national level, to ensure fairness, independence, trustworthiness, and more effective and consistent monitoring of impact.
- There is no mention of what should occur following monitoring activities. Would monitoring reports be publicly available? Would reports make recommendations? What would happen if findings give cause for concern?

- *"Reactive activities: firefighting, public investigation and regulatory action where needed"* (11). Please see Accountability below for further comments.

- Multidisciplinary team at the National Centre of Expertise (or other governance bodies) to provide free advice to NHS organisations (11).
- NHS organisations to seek legal advice and guidance from the National Centre of Expertise recently setup by NHSX.
- Meaningful national-level and local-level transparency:
  - *"A central register of data partnerships."* This should provide *"Information about all NHS health data partnerships"* including *"short, accessible explanations that summarise the partnerships' purposes, the data involved and how decisions have been made."* For this to work, reporting requirements for all NHS organisations would need to be established and imposed (11).
  - Each NHS organisation should also provide information about its data partnerships on its website. *"82% of people expect the NHS to publish information about health data partnerships"* (19).
  - A stipulation for applicants to declare on access request applications what other NHS data they currently have access to and the terms of those agreements, in much the same way that applications for funding specify what other funding has already been granted to the (academic) applicant.

*Auditing and monitoring by the data host organisation*

- Specific auditing and monitoring activities by each data host (controller) organisation are not mentioned. Arguably, national-level monitoring activities will be insufficient as they cannot audit and monitor each individual data partnership. This would need to be done by each data host organisation. See "Obvious gaps" (below).

*Learning and adapting processes:*

- Mechanisms to learn over time (e.g. new technologies, laws etc) and adapt governance processes and systems accordingly (13).
  - Beyond encouraging learning and adaptation when technologies change or other changes occur, there should be structured processes established for this, e.g. regular reviews of processes and systems.
  - There should be formal processes to investigate breaches and/or failures and to learn from them, to prevent them occurring again, or at least to reduce the likelihood of them occurring again.

Accountability:

- Please see "Accountability" in "Elements worth including" for what UPD proposes.
- Process(es) to address any prohibited actions need to be clearly defined and documented, and where appropriate these should be publicly available (this may exceptionally not be

appropriate for processes that address specific external malicious threats). There will be a need for formal processes that investigate breaches/failures/prohibited actions and formal processes that address found breaches/failures/prohibited actions.

- While UPD refers to imposing sanctions if things go wrong, they do not recommend that the data controller comprehensively and systematically consider what actions should be explicitly prohibited for organisations that have been granted data access. These will not always be obvious and it would be appropriate for the data controller to determine systematically a list of prohibited actions.. Please see above for various actions that may be prohibited, including (but not limited to) sharing data with third parties, unapproved data access or use, attempting to re-identify any individual from the data, providing false information on the data access application form, any other breaches in the data partnership contract.
- Clearly include prohibited actions in documentation—e.g. in process documentation, in principles for data access, and in data partnership agreement terms.
- Solely prohibiting certain actions does not increase the trustworthiness of the system; prohibition must be accompanied by preventative controls, detection controls and accountability measures for increased trustworthiness.
  - Even when certain actions are prohibited, all reasonable preventative measures should be taken to significantly reduce the likelihood of prohibited actions being performed. This may include the implementation of various technical controls in the secure environment.
  - Even where preventative controls exist, there is always a risk of prohibited actions being performed. Consider how these would be detected if performed. What detection controls are required?
  - What accountability should there be when prohibited actions are detected?
- While UPD indicates the importance of accountability and the need to enforce sanctions if things go wrong, they do not provide any details on the type of sanctions, remedies or penalties that may be imposed. Some guidance may be appropriate here.
  - For many "things that go wrong" or prohibited actions it may be necessary to immediately remove all data access.
  - What other sanctions/remedies/penalties may be imposed? Monetary fines? Legal action? Reporting the entity to a central registry of organisations banned from gaining further patient data access? Public naming of organisation? The latter may be appropriate for transparency and also to prevent the entity gaining access to other sensitive data, however, it may not be implemented by the data controller for fear of public criticism (over why access was granted in the first place). Can the independent oversight organisation(s) mandate that data controllers take certain actions such as publicly naming offending entities to ensure "public and democratic accountability"?
  - There should be PPI in deciding the range of sanctions, remedies and penalties that can be imposed, and in deciding what should be imposed in any given situation.
  - What other steps may be necessary to limit damage? E.g. what about data which the entity has already obtained or what about other NHS data that the entity has access to elsewhere?
- There is a lack of consideration of accountability for the data controller. When external independent monitoring uncovers an issue, how should the data controller be held to account? While UPD refers to *"firefighting, public investigation and regulatory action where needed"* when external independent monitoring uncovers an issue, there is a lack of detail and clarification, i.e. who should be subject to public investigation—the commercial entity, the data controller, or both? Accountability for the data controller needs to be comprehensively considered: in what situations might it be appropriate? What about when external independent monitoring uncovers a pattern of issues at a data controller? Depending on the issues, beyond reporting them to the Information Commissioner's Office (ICO) (and other appropriate regulators), what other actions should be taken to safeguard patient MH data? Are there situations in which a data controller can be banned from sharing patient MH

data with third parties?

**Noted to avoid** Consideration of industry:

- Treating all industry as a homogenous entity. Instead, do acknowledge that there are different industry sectors.
  - Explicitly ask host organisations to specifically consider and decide, with meaningful patient and public involvement, their data access approach for different industry sectors and sub-sectors.
  - Seriously consider diverse public concerns and opposition to certain types of organisations accessing this data. Do not limit this to the often-repeated (or parroted) concerns over insurance or marketing organisations gaining access to this type of data.
  - Ask host organisations to provide clarity and to be (publicly) transparent on whether distinctions will be made between different industry sectors when considering access requests for patient MH data.

Types of data:

- Making unclear and concerning statements about sharing personally identifiable patient data. UPD says that personally identifiable patient data may be shared with non-marketing and non-insurance commercial organisations *"if there is a clear health benefit"* (20). Here, UPD may be referring to the sharing of personally identifiable patient data with private companies that are involved in healthcare provision (e.g. pharmacies or private healthcare providers) rather than commercial organisations accessing patient data for research purposes, but they do not make this clear.
  - Personally identifiable MH data should not be shared with companies for research purposes or for any purpose beyond the direct and present delivery and provision of healthcare.
  - When referring to personally identifiable MH data take care to provide clarity on who can access this, for what purpose and under what circumstances. Ensure that statements about personally identifiable MH data are not open to interpretation and do not leave the public with further concerns. Be transparent.
- Omitting the consideration of data linkage, especially the data linkage of highly sensitive data, such as MH data, with other types of data.
  - Omitting the risks and concerns of data linkage, and the increased risks and concerns of data linkage of highly sensitive MH data with other types of data.
- Omitting the specific and appropriate consideration of free-text MH patient data.
  - The content of free-text MH data is one of the most distinguishing aspects of MH data when compared to other types of healthcare data. Therefore, if this is not sufficiently or appropriately considered, with due consideration of concerns of affected patients, in guidelines for sharing MH data, this would be a serious failing.

Making statements with generalisations about public views, drawing bold conclusions from prior research without acknowledging the limitations of that research and without appropriately recognising the distinctions that may apply to different types of data, especially to free-text MH patient data:

- UPD states that *"care should be taken when generalising about peoples' views"* and that people's *"attitudes are rarely fixed and do not always represent particular demographics, or apply equally across different contexts"* (16). Despite this, UPD sometimes makes statements which generalise about public views, including drawing conclusions from prior research and wrongly applying them across different types of data including those for which such conclusions do not apply.
- Although UPD is clear that many people are uncomfortable about and have concerns about *"companies accessing their health data,"* they are arguably too quick to draw conclusions and generalise on other public views despite the limitations of previously conducted

research. For example, they say people *"are generally comfortable with anonymised data from medical records being used for improving health, care and services, for example for research, provided there is a public benefit"* (16). Elsewhere, UPD acknowledge that there is inadequate research on public views with regards to specific types of health data, especially more sensitive health data such as MH data (16). Furthermore, there is a lack of adequate research on public views specifically with regards to the sharing of free-text MH data. Despite this, conclusions drawn from research are being wrongly used to show support for and argue for increased sharing of all health data even though MH data and especially free-text MH data is much more sensitive.

  - Do not generalise based on previous research;
  - Do not wrongly use findings from prior research to support or argue for the sharing of free-text MH data.
  - Be clear about gaps in prior research and be clear on what is different about free-text MH data.

- When UPD states that prior research shows that the public is generally in favour of health data being used for research (albeit not by commercial organisations) they are referring to the sharing of anonymised or de-identified data (14,16). Arguably, free-text MH patient data cannot be considered anonymous. Hence, no prior research on public views can be shown to strongly support the sharing of free-text MH data with commercial organisations.
  - Do not generalise based on previous research;
  - Do not wrongly use findings from prior research to support or argue for the sharing of free-text MH data.
  - Be clear about gaps in prior research and be clear on what is different about free-text MH data.
- UPD states that *"the more informed people feel, the more they are likely to support"* the sharing of health data for research (16). However:
  - Such support may remain general in nature and without reference to specific types of sensitive data or specific purposes, as found by more deliberative research e.g. during two 2017 citizens' juries, while members became more supportive of sharing patient data for public benefit in general, they were less supportive of sharing data for specific purposes (16).
  - For previously conducted citizens juries (whether the 2017 citizens juries, the UPD-commissioned 2019 citizens juries, or other) it is unclear which experts were involved (i.e. which experts presented to and were questioned by the jury) and whether any organisations or persons who take a more cautious approach to health data sharing were involved.
  - No prior research on public views can be shown to strongly support the sharing of free-text MH patient data with commercial organisations.
  - Previous research on public views on the sharing of MH data appears to be quite abstract and non-specific and questions to the public often appear to be framed to evoke a "fear of missing out" (FOMO) on apparent, future, unspecified public benefits (e.g. x % *"of people in the study would rather commercial companies access data than miss out on research benefits"*) (16). Aside from other significant limitations of such research, arguably, the public participants were not asked to consider, in detail, the kind of highly sensitive information such free-text data may contain.
  - In previous research, the public participants who participated may never have disclosed highly sensitive information during psychotherapy and/or to MH services and may not be representative of the people affected by the sharing of such information:
    - If public participants are allowed to self-identify as having a mental illness, without having received a clinical diagnosis, how representative are they of affected individuals?
    - If participants have received a clinical diagnosis of mental illness, but never received intensive therapy or disclosed highly sensitive information during

psychotherapy and/or to MH services, how representative are they of affected individuals?

  - Would people who self-select to participate in such research likely have different views to affected patients who do not self-select?
  - Would affected patients who have suffered different types of abuse and/or trauma have different perspectives on the sharing of MH free-text data?
  - Is it acceptable for those who self-identify as having mental illness and those who have never disclosed highly sensitive and private information during psychotherapy and/or to MH services to talk on behalf of a group that is directly affected by the sharing of such information? For any other demographic, this would not be considered acceptable.
  - A valid question is why researchers for prior studies seem to be indifferent about whether participants have shared highly sensitive and private information during psychotherapy and/or to MH services and whether participants would be directly impacted by the sharing of free-text MH patient data. Surely that is important to know before asking public participants for their views?
- Do not assume support for the sharing of free-text MH patient data with commercial organisations, especially from the people most impacted by the sharing of such data.

Problematic wording and tone emphasising financial benefits:

- At times, the wording and tone in the report, suggest that patient data is viewed as being *"post-Brexit… valuable national assets"* (11). Such wording can encourage undue focus on financial benefits or revenue generation, encouraging (cash-strapped) NHS trusts to take a more relaxed or accommodating approach to sharing patient data, which in turn may lead to practices which undermine public trust and potentially lead to misuse of patient data. Avoid wording and tone which depicts patient data as financial assets.

Contradictory purpose for which data can be shared, which may lead to sharing MH data solely for commercial gain:

- For the purpose for which health data should be shared with third parties, UPD's guidance is contradictory.
- On one hand, UPD states that the exploitation of *"health data solely for commercial gain"* would be contrary to public expectations, that improvements to health and care *"must be prioritised over financial incentives"* and states that jurors (in the citizens juries) felt that *"improved health outcomes should always be the primary purpose of using health data"* (11,19). On the other hand, UPD has a problematic definition of "improved health outcomes" which could lead to uses of health data solely for commercial gain. This is because the definition frames "additional revenue for the NHS" as one possible "improved health outcome", i.e. a solely financial outcome would cease to be a solely commercial purpose.
  - Ensure that the defined purpose for data access/use does not include financial benefits;
  - Ensure PPI in defining purposes for data use;
  - Specifically consider MH data use cases;
  - Ensure that the defined purpose for data access/use is carefully worded to avoid contradictions and obvious loopholes. Test any proposed defined purpose, with patients and the public, to identify potential contradictions and loopholes;
  - Rather than distinguishing between direct and indirect purposes (whereby indirect purpose may include revenue for the NHS) consider distinguishing between primary and secondary purposes, whereby it is mandatory for projects to meet the primary purpose. Primary purposes may include direct and indirect improvements to health and care, whereas secondary purposes may include revenue for the NHS.

Omitting consideration of project failure and the NHS's likelihood to implement project outputs when considering data access requests:

- Why consider whether a project will provide patient benefits if those benefits will not be realised within the NHS?
- It is notable and concerning that over 85% of data science projects apparently fail. In cases where patient data is used this should not be in vain, i.e. such private data should not be shared without taking appropriate and adequate measures towards minimising the failure of the project, otherwise it is arguably not ethical to share such data.
- Failures may be project failure or failure to utilise research outputs.
    - For projects where there are tangible and implementable project outputs, the failure to implement these within the NHS ultimately means a failure to realise patient benefits. This does *not* mean that the NHS *should* implement all such project outputs. Instead, this means that it may not be in the public interest for access to patient data to be granted for projects for which there is a high likelihood of outputs not being implemented by the NHS.
    - To reduce the risk of this, should every data access request proposing implementable outputs be required to conduct a preliminary acceptability assessment of the NHS's readiness or willingness to implement proposed outputs? Please see "Elements worth including with amendments" for further detailed comments on this.
    - For data access requests proposing implementable outputs, the review process should consider the score/result from the preliminary NHS acceptability assessment, and if the result indicates that the project outputs are unlikely to be implemented within the NHS, it may be advisable to reject the data access request.

Insufficient meaningful transparency and enablement of public scrutiny:

- Omitting the sufficient consideration of public registries of (approved) data access requests and what information such registries may contain.
- Lack of clarity on external oversight of decisions on commercial access to patient MH data, e.g. should local data access processes be subject to an external and independent audit?

Omitting consideration of the requirements for the data access process:

- Beyond ensuring PPI in assessing data access requests, UPD does not mention specific requirements for the data access process.
- UPD does not indicate what information commercial applicants for patient data access should be required to provide on applications. Please see "Data Access Processes" in "Elements worth including with amendments" for further comments.
- While UPD refers to ensuring that the staff working with the data have the necessary competence, it does not detail how this could be ensured for staff at commercial organisations. Please see "Data Access Processes" in "Elements worth including with amendments" for further comments.
- While UPD refers to the need for the data access process to be transparent, it does not explicitly recommend that the end-to-end data access process should be documented, including in an accessible format, and be made publicly available (online).

Neglecting the specific consideration of MH data for use cases and how data access requests should be reviewed:

- If all data access requests will not be reviewed by patient/public reviewers, and if only access requests in certain "use case" categories will be reviewed by patient/public reviewers then:
    - There should be PPI in determining the "use case" categories and the method/process for allocating cases to categories.

  - There should be public transparency on how the determination is made.
  - There should be regular PPI in reviewing the categories and the method/process for allocating cases to categories.

Problematic consideration of health inequalities:

- UPD says that when considering partnerships between the NHS and third parties, improvements to health and care *"must aim to improve health and care for everyone"*; improvements to health and care must be *"distributed across the country, to prevent making health inequalities worse"*; benefits should be shared across the population and *"avoid making health inequalities worse"*; *"Improvements should be fairly distributed across the NHS"* (11,15,19).
- While the potential impact on health inequalities should be considered when assessing data access requests, it is not appropriate or realistic for every project requesting data access to aim to improve *"health and care for everyone."*
- To tackle health inequalities, it is necessary to focus on those who are adversely affected; resources and improvements will have to be distributed unequally, with more directed at those who are adversely affected, to reduce health inequalities and to ultimately strive for equal health outcomes. Therefore, for projects that aim to tackle an aspect of existing health inequalities, the benefits from such partnerships will not be equally distributed across the NHS or country.
- Projects will be necessarily focused on specific solutions for specific health challenges. Hence any given project cannot possibly *"improve health and care for everyone."*
- Some projects may focus on rare diseases or illnesses affecting certain demographics.
- It would be inappropriate and limiting to consider whether benefits only pertain to a local area or demographic, and it would instead be more appropriate and fairer to consider the potential impact of the data partnership on existing health inequalities: will the data partnership likely exacerbate existing health inequalities, have no difference, or reduce health inequalities? The benefits from a project may not be shared across the population, but may help to reduce health inequalities.

National decision-making on data access requests:

- Suggestions by several jurors (and via the survey) that decision making about data partnerships should occur at *"national level to ensure an even distribution of outcomes, benefits and rewards across the country"* (11).
  - In the citizens' juries and survey, concerns were raised about benefits from data partnerships not being fairly distributed across the NHS and across the country. In particular, concerns were raised about poorer and rural areas being left behind or excluded, e.g. some technologies may be developed without due consideration of such populations.
  - However, the report argues that local regions are *"best placed to make decisions for their local populations, and disease specialists best placed to make decisions for the communities under their care."* Therefore, there are clear advantages to local decision-making. However, it is important for all decision-making regarding data partnerships to take account of fair distribution of benefits and to ensure that such partnerships do not result in further health inequalities. See further comments on considering the potential impact on health inequalities when assessing data access requests and also further comments on monitoring the impact of data partnerships.

Inappropriate and inadequate consideration of modifications to data access and use:

- UPDs comments on modification to data access and use imply a lack of sufficient consideration of the risks. UPD says that any *"major alterations in purpose or scope should trigger the data applicant to go back to the data assessor as a minimum to acknowledge the* *change or to seek approval to proceed on the basis of the new focus for the work"* (18).

- ◦ It would be inappropriate for the commercial organisation to simply inform the data assessor/owner of the change in purpose or scope (albeit as a minimum). Just because the original purpose or scope was approved, it should not be assumed that any changes in purpose or scope would be approved.
- ◦ Beyond changes in purpose or scope, there may be other changes including (but not limited to) changes in what data is required, the duration for which it is required, how the data is used (even if the purpose remains the same) and which staff require access to the data. All such changes would be referenced as changes in data access and/or use.
- ◦ Any such changes should not be requested or approved informally. UPD's statements on data access/use modification imply a lack of recognition of the importance of a formal process. Any changes to data access and data use should be handled via formal documented processes and subject to formal approval. That is, the commercial organisation should be required to formally request modified data access and/or use.
- ◦ UPD's statements also imply that changes in data access and use *would* be approved. There is almost a presumption of retrospective approval for changes in project scope. Just because the original access application was approved, it should not be assumed that any changes in data access or use will be approved. NHS Trusts and/or other data owners of patient data should not feel obligated, coerced, or forced into approving modified data access just because a project has already commenced.

- Please see "Data Access Processes" in "Elements worth including with amendments" for further comments.

A relaxed, informal, or ad hoc approach to commercial organisations sharing MH data with third-party organisations:

- UPD says that companies sign a contract which *"usually"* states *"that data cannot be passed to any third parties, unless explicitly approved in the application"* (20). Based on the use of the word "usually," it appears that such contracts do not always prohibit companies from sharing patient data with any third parties.
  - ◦ Data partnership contracts should prohibit companies from sharing any patient data with any third-party organisations.
  - ◦ Rather than allowing companies to share such data with (unnamed) third parties, if access is required by multiple organisations, for a single project, this should be explicitly requested in the application and access should be approved, as appropriate. This would allow all third-party organisations who request access to such data to be appropriately vetted by the host organisation, with PPI.

Other aspects of data access:

- Currently, the responsibility for securely storing patient data apparently lies with commercial organisations. They are required to ensure *"controlled access and robust IT systems to keep data safe"* (20). Arguably:
  - ◦ Commercial organisations should not have sole responsibility for the secure storage of patient data.
  - ◦ This may result in the use of insecure methods of data transfer to the commercial organisation.
  - ◦ Access to such data should be via secure environments.
  - ◦ UPD says the NHS is moving away from data transfer to third parties *"to a system of data access where NHS data is stored in a Secure Data Environment (SDE) that can be accessed by approved users but data cannot leave the platform"* (20).
  - ◦ For MH data, especially, there should not be direct transfer of data and access should be via an SDE.

Governance and monitoring: data partnership agreement/contract terms:

- Complete omission of requirements for data partnership agreement terms.
- Insufficient information on payments and payment structures that can be considered by data controllers.

Accountability:

- Insufficient information or requirements for data controllers with regards to how to implement accountability processes and measures, e.g. no requirement for data controllers to undertake comprehensive and systematic consideration of *what* actions should be prohibited, no requirement for formal documented processes on *how* to address prohibited actions, no requirement for detection controls to detect prohibited actions, lack of detail on sanctions/penalties/remedies.
- Lack of consideration of how to limit damage.
- Lack of distinction between accountability for commercial entity and accountability for data controller.

Note also that UPD covers all patient data and does not specifically focus on MH data, which is highly sensitive.

**Gaps**

- Requirement for data host to provide clarity on types of commercial organisations that can potentially access patient MH data.
- Categorisation of different types of MH data and clarity on what types of MH data can be available and what types of MH data will not be available for sharing with commercial entities.
- Considerations of different industry sectors and the data access approach for different industry sectors.
- Ambiguity and lack of clarity on the sharing of personally identifiable patient data. Lack of explicit prohibition on the sharing of personally identifiable patient data with private-sector organisations for any purpose beyond the direct and present delivery and provision of healthcare.
- Guideline advising against sharing of full MH record of any patient.
- Specific and appropriate consideration of free-text MH patient data, and the substantial concerns specific to the sharing of such data.
- Adequate acknowledgement of the gaps in prior research, e.g. (i) lack of (unbiased) research, using sufficiently deliberative methods, on public views on the sharing of specific types of sensitive health data, including MH data, with commercial organisations; (ii) lack of deliberative research, involving public participants who have shared highly sensitive information during psychotherapy and/or to MH services, on their views on the sharing of free-text MH data.
- Requirement for data host to define a purpose for data access, preferably co-defined with patients and the public, against which data access requests can be assessed. Requirement for the defined purpose to include a **primary** purpose which requires an improvement in health and care of any project requesting MH data access, whereby *"improvements in health and care"* should be clearly defined and **exclude** financial benefits.
- Requirement for defined purpose to *"be tested against potential unforeseen consequences or harms to individuals and society"* (17,18). Such testing to include PPI. Requirement to check for contradictions and obvious loopholes in the defined purpose.
- Requirement for the defined purpose to be regularly reviewed with PPI. Any changes in the defined purpose to be tested again, with PPI, *"against potential unforeseen consequences or harms to individuals and society"*.
- Requirement for appropriate independent experts to review data access requests.

- Consideration of project failure and the NHS's likelihood to implement project outputs when considering data access requests
  - Why consider whether a project will provide patient benefits if those benefits will not be realised within the NHS?
  - Over 85% of data science projects apparently fail. Where patient data is used this should not be in vain, i.e. such private data should not be shared without taking appropriate and adequate measures towards minimising the failure of the project, otherwise it is arguably not ethical to share such data.
  - Failures may be project failure or failure to utilise research outputs. For projects proposing tangible and implementable project outputs:
    - It may not be in the public interest for access to patient data to be granted where there is a high likelihood of outputs not being implemented by the NHS.
    - Should the organisation making the data access request be required to conduct a preliminary acceptability assessment of the NHS's readiness or willingness to implement proposed outputs?
    - The data access request review process should consider the score/result from the preliminary NHS acceptability assessment, and if the result indicates that the project outputs are unlikely to be implemented within the NHS, it may be advisable to reject the data access request.
- Public registry of approved data access requests for each data host and what information to include on such public registries.
- For the terms negotiated for each data partnership to be made available on the public registry.
- How to ensure staff competence (with regards to working with sensitive data and professional ethics) at commercial organisations.
- Technical controls to ensure that undue reliance is not placed on staff competence, ethics, or the absence of human error at commercial organisations.
- For staff and public/patient reviewers at data host to be trained on all data access processes and criteria for making data access request assessments. For staff and patient/public reviewers at data host/controller to be provided with documentation of formal processes.
- What information commercial applicants should be required to provide on data access applications.
- Requirement to document the end-to-end data access process, including in an accessible format, and to make this publicly available.
- If categories will be used to determine what data access applications will be reviewed by public and patient reviewers, there should be PPI in determining the categories and the method/process for allocating cases to categories. Public transparency on how determinations are made.
- Regular PPI in reviewing the categories and the method/process for allocating cases to categories.
- The data access request application form (and associated guidance) to be regularly reviewed and updated as appropriate, e.g. to ensure sufficient and relevant information is provided by applicants, or in response to issues identified across multiple applications.
- The methods, processes, and tools (e.g. prompts, templates, forms) for conducting data access application reviews should be regularly reviewed, and updated, as appropriate.
- Clarity on assessing impact on health inequalities when assessing data access requests
  - To address health inequalities, benefits from data partnerships may well need to be distributed unequally and may not be *"shared across the population"*;
  - What specifically should be checked when assessing data access applications when assessing impact on health inequalities?
- Formal process for change(s) in data access and/or use.

- PPI in assessing change(s) in data access/use.
- Consideration of changes other than changes in project scope.
- Requirement to document the end-to-end process for requesting and reviewing change(s) in data access and/or use, including in an accessible format, and to make this publicly available.
- Procedural and technical controls to ensure that commercial organisations are severely restricted in modifying data access or data use without formal approval.
- Sufficient consideration of risks related to change(s) in data access and/or use.
- Consideration of change(s) in data access and/or use in data partnership agreement terms.
- Prohibition on commercial organisations sharing data with third-party organisations.
- Measures and rules regarding publication—anonymisation of data.
- Formal data access process should allow for access to be requested for multiple organisations (when multiple organisations working on a project).
- When access is required by multiple organisations for a given project, the formal processes should be followed (formal data access process or formal process for change(s) in data access and/or use).
- Technical controls to make it technically more difficult for companies to share patient data with third parties.
- All access to MH data to be via SDEs. Access controls within SDE.
- No direct transfer of MH data to commercial organisations.
- Specific requirements for project progress reports—what such reports should contain, frequency.
- PPI in project progress report reviews.
- Requirements for the terms of data partnership agreements.
- Guidance on payments and payment structures.
- PPI in establishing the national-level governance framework and oversight.
- PPI in developing the initial national-level principles, guidance, and advice.
- Clarity on what specifically should be subject to external monitoring/review/audit and how such monitoring should be conducted in practice, e.g. should local data access processes be subject to an external and independent audit? Should this check:
  - How the data access process was designed?
  - The design of the data access process?
  - Whether the data host organisation's documented data access process was properly followed?
  - Whether data access should have been granted?
- Reference to PPI in monitoring/auditing/reviewing activities.
- Clarity on monitoring and evaluating the impact of data partnerships.
- What should occur following monitoring activities: Would monitoring reports be publicly available? Would reports make recommendations? What would happen if findings give cause for concern?
- Stipulation for commercial applicants to declare on access request applications what other NHS data they currently have access to and the terms of those agreements (similar to how funding applicants have to declare other funding on funding applications). While it is currently not required for applicants to declare other (NHS/public) data access on data access request applications, this information could be very valuable, e.g. in informing appropriate responses to prohibited actions.
- Specific auditing and monitoring activities by each data host/controller.
- Audit and monitoring of each data host/controller (by a national organisation?).
- Formal processes for learning and adapting processes, as appropriate.

    - Formal learning processes for changes in technologies/systems/laws/policies/other changes.
    - Processes that enable learning from things that go wrong with appropriate actions taken and changes made (to processes and systems). These should include learning from breaches, failures and entities performing prohibited actions.
- Reference to any requirement for patient and public involvement at the commercial organisation (from application to project completion).
- Detection controls.
- Processes to investigate breaches/failures/suspected prohibited actions.
- Processes to respond to breaches/failures/prohibited actions that are found. The steps that should be taken in the event of a breach/violation of the data partnership terms.
- Guidance on sanctions, penalties, and remedies.
- Consideration of how to limit damage in the event of breaches/failures/prohibited actions.
- Distinction between accountability for commercial entity (granted access to data) and accountability for data controller.
- Requirement to document all processes related to data access including processes to respond to breaches (this may exceptionally not be appropriate for processes that address specific external malicious threats).
- Requirement of regular risk assessments to ensure that robust preventative measures are taken and/or updated as appropriate to reduce risks as much as possible.
- Other gaps in addition; not all listed.

**Public involvement approach**

This aspect of the review includes:

- UPD's apparent approach towards the public;
- Consideration of patient and public benefits and concerns; consideration of public/patient impact;
- PPI in developing the guidelines/recommendations;
- Consideration of PPI in processes concerned with data access, monitoring, and management, e.g. do guidelines consider PPI?

*UPD's approach towards public*

Be trustworthy rather than trying to build trust:

- UPD advocates for an approach focused on being trustworthy rather than developing public trust: *"Start with being trustworthy"* rather than trying to build trust (13).
    - It is not the public's responsibility to trust organisations who use their data, and members of the public cannot be criticised for not "correctly" trusting such organisations; the public should not be expected to trust "by default".
    - UPD argues that peoples' willingness to trust is influenced by their own subjective experiences and values (13). However, arguably, an individual's willingness to trust an organisation's data approach would also be influenced by how trustworthy the organisation is, the trustworthiness of the data approach and system, and the effectiveness of the organisation's communication of the system's trustworthiness. Some may claim to trust an organisation's data approach and system, without considering any information or evidence of trustworthiness—is that trust or indifference? Some may not have the willingness or capacity (due to other priorities) to ask questions or process information.
    - By ensuring that the onus is on ensuring that there is trustworthiness within the system, the organisation can consider what it can control to make the system more trustworthy:
        - *"What can be done to make people, systems, and institutions worthy of peoples'*

*trust"*?
    - What *"practices and characteristics"* can be developed and what behaviours can be adopted to make a system/organisation more trustworthy?
    - How can the organisation/system provide *"adequate, useful, and simple"* evidence that it is trustworthy (13)?
  - Interestingly, note that an approach focused on being trustworthy rather than building public trust is an inherently different approach. The "building public trust" approach typically has a narrative along the lines of "we know we are already trustworthy and we know we are already doing things properly, but how can we convince the uninformed public of this?" In contrast, the "being trustworthy" approach would have a narrative along the lines of "what can we do and what can we continue to do to be trustworthy and to demonstrate that we are trustworthy?" While UPD does not state the following, arguably it is a:
    - Shift *towards* being open to considering patient/public concerns, the basis for the concerns, and how concerns might be addressed. There are a broad spectrum of risks and concerns, and there is a need to consider this from a diverse range of perspectives.
    - Shift *away* from characterising one of the main issues as being an uninformed public with unfounded and unreasonable concerns. While some members of the public may be less informed, public concerns should not be automatically dismissed on the presumption that the concerns have no basis. Diverse patient and public perspectives can reveal risks and concerns that have not been (sufficiently) considered by experts/professionals/researchers. Some public concerns may be probable, some may not be probable, but may be possible. The challenge is not one of educating the public to the "correct" way of thinking.
    - Shift *towards* providing useful information and evidence to the public about what is being done to increase trustworthiness in the system. There is a distinction between what measures are being taken and the effective communication of this. If the latter is not done, then the public would not know what measures are being taken.
    - Shift *away* from communications for the purpose of persuasion.

Engage in genuine dialogue rather than transmitting information or persuading, and ensure meaningful transparency:

- UPD encourages organisations to engage in genuine dialogue rather than just "transmitting information" to help increase trustworthiness (16). This may involve listening to concerns, and then engaging and responding to concerns. Often communications about a data approach focus on the importance of health data sharing for research without actively engaging with concerns expressed. Genuine dialogue may ensure that concerns are not side-stepped or dismissed.
- Do not overload people with too much information or "technical language"—this is not meaningful or relevant (13).
- When communicating, rather than persuading, the aim should be to inform, allowing people to reach their own conclusions, i.e. organisations should not sell their data approach to the public (13). This ties in with taking an approach focused on being trustworthy rather than trying to build trust (see comments above).
- Acknowledgement of and openness about risks and uncertainty; do not deny that risks exist, be open about them and share steps being taken to mitigate those risks (13,14).
- Be able to sincerely answer questions about motives.

Do not generalise about public views:

- UPD advises against generalising about public views on patient data, arguing that views can vary across people, demographics, time, and contexts (15–17).

- UPD does not appear to explicitly acknowledge that public views can also vary across specific types of data—e.g. while someone may be supportive of the sharing of consented patient data obtained from cancer trials, they may be against the sharing of routinely collected unconsented free-text MH data. However, UPD recognises the insufficient research on public views for specific types of patient data, including more sensitive data such as MH data (16).
- Despite UPD's advice not to generalise about public views, there are several instances of UPD making statements with generalisations about public views, drawing bold conclusions from prior research without acknowledging the limitations of that research and without appropriately recognising the distinctions that may apply to different types of data, especially to free-text MH data. For example, they say people *"are generally comfortable with anonymised data from medical records being used for improving health, care and services, for example for research, provided there is a public benefit"* (16). However, this does not necessarily apply to the sharing of much more sensitive free-text MH data with commercial organisations:
    - There is a lack of adequate research on public views specifically with regards to the sharing of free-text MH data.
    - Previous research on public views on the sharing of MH data appears to be quite abstract and non-specific and questions to the public often appear to be framed to evoke a "fear of missing out" (FOMO) on apparent, future, unspecified public benefits (e.g. x % *"of people in the study would rather commercial companies access data than miss out on research benefits"*) (16). Aside from other significant limitations of such research, arguably, the public participants were not asked to consider, in detail, the kind of highly sensitive information such free-text data may contain;
    - In previous research, the public participants who participated may never have disclosed highly sensitive information during psychotherapy and/or to MH services and may not be representative of the people affected by the sharing of such information:
        - If public participants are allowed to self-identify as having a mental illness, without having received a clinical diagnosis, how representative are they of affected individuals?
        - If participants have received a clinical diagnosis of mental illness, but never received intensive therapy or disclosed highly sensitive information during psychotherapy and/or to MH services, how representative are they of affected individuals?
        - Would people who self-select to participate in such research likely have different views to affected patients who do not self-select?
        - Would affected patients who have suffered different types of abuse and/or trauma have different perspectives on the sharing of MH free-text data?
        - Is it acceptable for those who self-identify as having mental illness and those who have never disclosed highly sensitive and private information during psychotherapy and/or to MH services to talk on behalf of a group that is affected by the sharing of such information? For any other demographic, this would not be considered acceptable.
        - A valid question is why researchers for prior studies seem to be indifferent about whether participants have shared highly sensitive and private information during psychotherapy and/or to MH services and whether participants would be directly impacted by the sharing of free-text MH patient data. Surely that is important to know before asking participants for their views?
- Support for the sharing of free-text MH patient data with commercial organisations, should not be assumed, especially from the people most impacted by the sharing of such data.
    - Prior studies have found that many people are uncomfortable about and have concerns about *"companies accessing their health data"* (16).
    - Arguably, free-text MH patient data cannot be considered anonymous.

    - **No prior research on public views can be shown to strongly support the sharing of free-text MH data with commercial organisations.**

*Consideration of patient and public benefits and concerns; consideration of patient/public impact*

Omits consideration of free-text (MH) data:

- UPD omits consideration or mention of whether patient data is structured or free-text information (14).
- UPD therefore neglects to consider the particular, wide-ranging, and substantial concerns related to the sharing of patient MH free-text data. See "Elements worth including with amendments" for details.
- **No prior research on public views can be shown to strongly support the sharing of free-text MH patient data with commercial organisations.**

Patient/public benefit:

- UPD states that patient data should only be used where there is public benefit (13,14,16). However, "public benefit" is an elusive concept that requires further definition.
- The report based on UPD-commissioned research does provide further information on the purpose for which health data should be shared with third parties (11). However, the guidance is contradictory. On one hand, it states that the exploitation of *"health data solely for commercial gain"* would be contrary to public expectations, that improvements to health and care *"must be prioritised over financial incentives"* and states that jurors (in the citizens juries) felt that *"improved health outcomes should always be the primary purpose of using health data"* (11,19). On the other hand, they have a problematic definition of *"improved health outcomes"* which could lead to uses of health data solely for commercial gain. See "Elements worth including with amendments" for further details.

*PPI in developing the guidelines/recommendations*

- Some of UPD's guidelines have been developed via UPD-commissioned research which included citizens' juries, public involvement, and public surveys.
- Arguably, possibly due to the lack of specific focus on MH data, and MH free-text data, there has been a lack of involvement of affected patients in developing the guidelines/recommendations, e.g. patients who have disclosed highly sensitive information during psychotherapy and/or to MH services do not appear to have been involved. Please see above ("Do not generalise about public views").

*Consideration of PPI in processes concerned with data access, monitoring, and management - guidelines which reference PPI or the public*

- *"Involve members of the public in setting the rules and frameworks for decisions about data,"* including *in "deciding which uses of data are or are not permitted"* (13).
- PPI in assessing data access requests (11,19).
- The report (based on UPD commissioned research) suggests taking a proportionate approach, categorising health data use into three broad categories:
    - *"Broadly acceptable use cases with clear public benefit, minimal privacy risks and low risk of controversy;*
    - *Grey area/mid-range use cases, for which there will be a diversity of views and perspectives;*
    - *'Red line' use cases highly likely to be considered unacceptable, such as using data for*

*insurance or marketing purposes, or clear commercial exploitation"* (11).

The report argues that taking *"a proportionate approach might mean focusing citizen involvement on cases in the 'grey area'"* (11).

- Arguably, with regards to unconsented MH data, all data requests should be reviewed by patient/public reviewers. If there is a high volume of requests, there should be a correspondingly high number of patient/public representatives.
- Public support for sharing data depends on the nature of the specific research (16).
- Deciding that certain data requests do not have to be reviewed by patient/public reviewers is problematic and raises concerns about how this would be decided.
  - Who decides the specific definition of each category? i.e. what is a "grey area" category?
  - Who decides how cases are allocated to categories, and which cases belong to which categories?
- If all data access requests will not be reviewed by patient/public reviewers, and if only access requests in certain "use case" categories will be reviewed by patient/public reviewers then:
  - There should be PPI in determining the "use case" categories and the method/process for allocating cases to categories.
  - There should be public transparency on how the determination is made.
  - There should be regular PPI in reviewing the categories and the method/process for allocating cases to categories.

- While UPD appears to take a generally positive approach to involving the public and patients, they do not explicitly mention PPI in many guidelines and recommendations. See also "Gaps" for omissions with regards to PPI.

**Industry involvement approach**

This aspect of the review includes:

- References to industry access to patient data;
- Consideration of different industry sectors;
- Industry involvement in developing guidelines;
- Guidelines that reference requirements for industry applicants for data access.

*References to industry access to patient data*

Industry access to personally identifiable data:

- Lack of clarity and concerning statements about the sharing of personally identifiable patient data with industry. UPD says that personally identifiable patient data may be shared with non-marketing and non-insurance commercial organisations *"if there is a clear health benefit"* (20). Here, UPD may be referring to the sharing of personally identifiable patient data with private companies that are involved in healthcare provision (e.g. pharmacies or private healthcare providers) rather than commercial organisations accessing patient data for research purposes, but they do not make this clear.
- UPD states that prior research has found that one condition people would want in place before companies can have access to any patient data is that identifying information is removed from data before it is accessed (16). Being identified from data is a common concern (14). Such concerns are likely increased for patient MH data. It is highly unlikely that MH free-text data can be truly anonymous and patients will be identifiable.

Public views on industry access to patient data:

- UPD acknowledges that many *"people are uncomfortable with the idea of private sector organisations accessing health data"* and prior research has found that a sizeable proportion of the public do not want commercial organisations to access patient data under any circumstances (16,18,20).

  - UPD states that there are some indications that public support for commercial access to health data (in general) has declined over time—53% supportive in 2016 versus 39% supportive in 2018 (16).
  - Much prior research has found significantly less trust among the public for commercial organisations with health data even if such data is de-identified (16).
  - Previous research has found increased concerns with regards to specific types of sensitive data, including MH data (16).
  - Note also the above-mentioned comments and concerns about the sharing of MH free-text data. **No prior research on public views can be shown to strongly support the sharing of free-text MH patient data with commercial organisations.**

Purpose for industry access:

- A report based on UPD commissioned research states that all access to patient data by commercial organisations must be for improvements to health and care (11,19). Since any project can potentially be framed as a public benefit and many projects can potentially be framed as a way to improve health and care, there is a need for more guidance on this.
- Note that prior dialogues *"and surveys have tended to examine support for benefits to the NHS, rather than a more generalised conception of public benefit"* (18). Hence, it would be misleading and inaccurate to refer to public support for the sharing of patient data with commercial organisations where there is potential for public benefit in general.
- The report based on UPD-commissioned research does provide further information on the purpose for which health data should be shared with third parties (11). However, the guidance is contradictory. On one hand, it states that the exploitation of *"health data solely for commercial gain"* would be contrary to public expectations, that improvements to health and care *"must be prioritised over financial incentives"* (11,19). On the other hand, they have a problematic definition of *"improved health outcomes"* which could lead to uses of health data solely for commercial gain. See "Elements worth including with amendments" for further details.

Industry partnership terms:

- UPD states that there is a lack of transparency over terms negotiated for data partnership agreements, which allows companies to *"approach multiple NHS providers with terms favourable to them"* (11).
- UPD states that research has found that the public is concerned about exploitation of the NHS by commercial organisations and believes that NHS organisations require consistent *"support and guidance to negotiate fair terms for agreements with third parties"* (11,19).
- UPD states that lack of NHS capability may result in lack of fairness between data access applicants as larger companies may be better placed to negotiate more favourable terms.
- UPD states that it is important to ensure that *"partnerships between NHS and third parties are, and are seen to be, fair"* (11). Therefore, it is important to ensure that fairness is achieved in actuality and appearance.

*Consideration of different industry sectors*

- UPD appears to treat all industry as a homogenous entity and does not acknowledge or distinguish between different industry sectors.
- UPD provides unclear information on the types of commercial organisations that can currently access patient data for purposes beyond the direct and present delivery/provision of healthcare.

- UPD does not specify any requirement for host organisations to specifically consider, with PPI, their data access approach for different industry sectors.
- Further clarity is required on what types of commercial organisations can potentially access patient MH data, and whether distinctions will be made between entities in different industry sectors. There should be a requirement for host organisations to make this information publicly available in accessible format.

*Industry involvement in developing guidelines*

- It appears that there was no industry involvement in developing the guidelines/recommendations.

*Guidelines that reference requirements for industry applicants for data access*

- UPD states that organisations should ensure that staff working with the data have the skills, *"expertise, competencies, tools and methodologies"* to do this safely, securely, effectively, only for intended purposes, and to complete the proposed research (13).
  - Note, however, that UPD is referring generally to anyone using patient data, rather than commercial entities specifically.
  - UPD does not provide any guidance on how staff competence should be ensured for staff at commercial entities.
- UPD says that any *"major alterations in purpose or scope should trigger the data applicant to go back to the data assessor as a minimum to acknowledge the change or to seek approval to proceed on the basis of the new focus for the work"* (18). The commercial organisation should be required to formally request modified data access and/or use and this should be subject to formal approval.
- UPD states that, currently, prior to getting data access, companies sign a contract which *"usually"* states *"that data cannot be passed to any third parties, unless explicitly approved in the application"* (20). Based on the use of the word "usually," it appears that such contracts do not always prohibit companies from sharing patient data with any third parties.
  - Data partnership contracts should prohibit companies from sharing any patient data with any third-party organisations.
  - Rather than allowing companies to share such data with (unnamed) third parties, if access is required by multiple organisations, for a single project, this should be explicitly requested in the application and access should be approved, as appropriate. This would allow all third-party organisations who request access to such data to be appropriately vetted by the host organisation, with PPI.
- Publish *"transparent and publicly accessible updates on the progress of the partnership towards achieving predicted benefits"* (11). While implied, it is not explicitly stated that the commercial entity should be responsible for producing project progress reports.
- UPD states that currently companies *"have to sign contracts that set out what they can and cannot do with the data"* (20).
  - Inevitably, there are other considerations for such agreements beyond data use, e.g. agreed monitoring, audits and reviews, terms of payment, accountability measures.
- UPD states that, currently, the responsibility for securely storing patient data lies with commercial organisations. They are required to ensure *"controlled access and robust IT systems to keep data safe"* (20). Arguably:
  - Commercial organisations should not have sole responsibility for the secure storage of patient data.
  - This may result in the use of insecure methods of data transfer to the commercial organisation.

  - Access to such data should be via secure environments. This should especially be the case for access to MH data.
- While UPD refers to governance and monitoring activities, there is a lack of detail on what specifically should be subject to monitoring.
- Offence to re-identify anyone from data granted access to (14).
  - However, note that this may be difficult, if not impossible, to detect, unless there are appropriate preventative and detection controls in place.
- Sanctions and fines to be imposed if companies misuse data (16). UPD provides a lack of specific guidance on this.
- Notable lack of guidelines that reference requirements for industry applicants, e.g. what information commercial entities should provide on data access application forms.
- See "Elements worth including with amendments" and "Gaps" for further comments.

**Equality, diversity, and inclusion considerations**

Do not generalise about public views:

- UPD discourages organisations from generalising about public views about patient data, arguing that views can vary across people, demographics, time, and contexts (15–17).
- UPD omits the potential for views to change depending on the type of data, which can also be the case, and this may be linked to individual characteristics—for example, those who have shared sensitive information with MH services may have different views on the sharing of free text MH data compared to the sharing of cancer data.

Accessible information:

- UPD encourages organisations to ensure that publicly provided information is easily available, findable, accessible, and understandable (13).

EDI considerations for staff:

- Emphasises EDI considerations for staff—staff should be representative of the population (13).

Consideration of health inequalities:

- Encourages consideration of impact on health inequalities when assessing data access requests, which is an important equity consideration.
  - However, UPD makes problematic statements with regards to this. UPD says that when considering partnerships between the NHS and third parties, improvements to health and care *"must aim to improve health and care for everyone"*; improvements to health and care must be *"distributed across the country, to prevent making health inequalities worse"*; benefits should be shared across the population and *"avoid making health inequalities worse"*; *"Improvements should be fairly distributed across the NHS"* (11,15,19).
  - It is not appropriate or realistic for every project requesting data access to aim to improve *"health and care for everyone"*.
  - To tackle health inequalities, it is necessary to focus on those who are adversely affected; resources and improvements will have to be distributed unequally, with more directed at those who are adversely affected, to reduce health inequalities and to ultimately strive for equal health outcomes. Therefore, for projects that aim to tackle an aspect of existing health inequalities, the benefits from such partnerships will not be equally distributed across the NHS or country.
  - It would instead be more appropriate and fairer to consider the potential impact of the data partnership on existing health inequalities: will the data partnership likely exacerbate existing health inequalities, have no difference, or reduce health inequalities? The benefits from a project may not be shared across the population, but may help to

| | |
|---|---|
| | reduce health inequalities.<br>◦ UPD does not stipulate how data access requests should be assessed for impact on health inequality. Potential "health inequality" considerations (among others) for projects making data access requests:<br>▪ i. Bias: What will be done to reduce bias? This is especially a concern for AI projects.<br>▪ ii. Affected demographics: What are the affected demographics for the illness being researched by the project? Does the project design, methodology or approach exclude or not sufficiently consider any affected demographic groups? How well does the requested data match the affected demographics?<br>• Emphasises importance of monitoring the impact of data partnerships, specifically that monitoring *"and evaluation will be critical to ensure that they are not making health inequalities worse or increasing regional disparities in skills, expertise and data use"* (11).<br><br>Lack of consideration and involvement of affected patients with regards to guidelines pertaining to the sharing of free-text MH data; for example, patients who have disclosed highly sensitive information during psychotherapy and/or to MH services do not appear to have been involved.<br><br>Lack of specific and targeted recruitment of patients with more cautious views on sharing patient data for involvement activities, including the citizens' juries.<br><br>Lack of involvement of experts with more cautious views on sharing patient data during citizens' juries.<br>• It is highly unlikely that individuals or organisations that take a more cautious approach to the sharing of patient data were involved in presenting information or their views during the citizens' juries.<br><br>Lack of explicit statements on diversity and inclusion with regards to guidelines which incorporate PPI. |
| **Other considerations** | – |
| **Summary of strengths and limitations** | Strengths and limitations of *UPD-commissioned mixed-methods deliberative research,* which included *"discussions with patient advocacy groups, three citizens' juries and a nationally representative survey of over 2,000 people"* (19). The research sought to find out what the public thinks constitutes a fair data partnership between the NHS and third-party organisations.<br><br>Strengths:<br>• Advantages of a deliberative method: *"Deliberation emphasises logically building people's understanding about a topic, drawing out multiple perspectives and trade-offs rather than driving at a consensus and allowing time for views to be expressed and developed"* (4).<br>• Study has breadth and depth due to the methods chosen.<br><br>Limitations:<br>• Note, however, that the citizens juries only ran for 2.5 days (in Sept 2019) which included the time spent on building understanding and expressing views, and arguably, this would not have allowed enough time for jurors to develop sufficient understanding on the topic.<br>• The research question is focused on the form that data partnerships should take and it therefore assumes that there will be (data) partnerships between the NHS and third |

parties including academia, charities, and industry, and is not questioning whether such partnerships should exist.

- Lack of focus and coverage of the sharing of specific types of sensitive health data, including MH data, and specific types of MH data, including MH free-text data, with commercial organisations.
- The public participants were not asked to consider, in detail, the kind of highly sensitive information such patient MH free-text data may contain.
- It is highly likely that the public participants who participated in this research may never have disclosed highly sensitive information during psychotherapy and/or to MH services and are not representative of the people affected by the sharing of such information.
- It is highly unlikely that individuals or organisations that take a more cautious approach to the sharing of patient data were involved in presenting information or their views during the citizens' juries.

*Strengths and limitations of UPD content overall:*

Strengths:

- Comprehensive information available;
- Information in range of different formats.

Limitations:

- There is a lack of specific consideration of MH data and the specific concerns this presents (see other comments on free-text MH data). This means that many relevant issues and guidelines have been omitted.

##### 3.10 Supplementary Table 10: LEAG member's review of Kirkham et al. (2020/2021)

Direct quotes are from the material reviewed. (Not all direct quotes from the primary source document are thus identified.)

| Domain | Reviewer's comments |
|---|---|
| **Elements worth including** | **Security, safe settings:** *"Ensuring that data are accessed in safe settings using clear and efficient procedures"* (6).<br>• It would be appropriate for data to be accessed in safe settings, although do note amendments under "Elements worth including with amendments."<br><br>**Security breaches and near misses:** *"planning in advance to prevent data breaches, using a recording process for data breaches, and reporting near misses"* (6).<br>• It is important to ensure security planning and preventative measures to reduce risks including the risk of data breaches. It is important for breaches and near misses to be reported.<br>• However, this guideline needs to be significantly expanded—see "Elements worth including with amendments".<br><br>**Security:** *"providing appropriate training and supervision for data users, and carrying out criminal record checks where relevant"* (6).<br>• This guideline has been included on the "future" checklist, meaning that it has not been designated for guiding current data access practices but is instead for the research community to help make implementable through the development of new protocols. It is not clear why this is.<br>• Appropriate training and supervision should be provided for data users now, especially when sensitive unconsented MH data is accessed. It is unclear why a provision cannot be made for this now, and why this is something that can only be delivered in the future.<br>• For those who make requests to access unconsented MH data, it would certainly be safer for criminal record checks to be carried out. While this may require additional resources (time, funding), such checks are arguably justified given the additional concerns around the sharing of unconsented MH data:<br>◦ It contains highly sensitive and highly personal information, including about trauma and abuse. Even where free text MH data is not shared, and only structured data is shared, it would be highly sensitive and personal.<br>◦ It contains information which increases the risk of identifiability. This may be the case even where only structured data (and not free text data) is shared. Further, note that there will be variations in what data is stored in structured fields, and this may change over time.<br>◦ Legitimate and justifiable patient fears over discrimination/prejudice/stigmatisation.<br>◦ Potential impact on patient interactions and relationships, including safeguarding concerns due to predators potentially accessing highly sensitive patient information without the patient's knowledge. Unless the data is completely anonymous, with no risk of identifiability at all, a patient may interact with someone who has read their MH record without knowing this, while the individual they are interacting with may know highly private and sensitive details about the patient. This may place the patient at a disadvantage, and/or expose them to seriously unethical and predatory practices. This is not an irrational concern, but may conceivably occur given that there are currently no limits on how many individuals a patient's MH record may be shared with.<br>◦ Legitimate patient fears over experiences and feelings talked about in therapy being exposed. Such patient fears may include people who the experiences/feelings were about |

finding out (e.g. patients may have talked about family members in therapy and they may reasonably not want them to know what was said). Patients may also have concerns about such content being shared more widely and people they know finding out about confidential matters. Patients may reasonably believe that sharing of MH data (with other organisations for non-clinical purposes) may lead to increased risks of therapy content being exposed, whether through cybersecurity threats, human error, or otherwise.

- ○ Legitimate concerns over what happens to patient data when/after it is fed into various Large Language Models (LLMs) and other types of AI systems for non-clinical purposes. Even if patient data is not entered into publicly available LLMs, private versions of LLMs may be used (which may nonetheless pose various concerns depending on the specific technology) and commercial entities may request access to patient MH data in order to train their own in-house developed LLM or other AI system on the patient data. In such cases, how would it be determined what patient data is retained within such systems without undertaking comprehensive testing?
- ○ Please see review of UPD content (**Supplementary Table 9,** "Elements worth including with amendments") for further information on concerns regarding the sharing of unconsented free text MH data.

**Ensuring privacy and anonymity for published data:** Statistical disclosure control should be incorporated *"by following rules designed to prevent identification of individuals"* (6).

- See also "Elements worth including with amendments".

**Transparency and scrutiny of research:** *"allowing other researchers to check analyses wherever possible (in addition to peer review)"* and *"providing access to synthetic data where real data cannot be shared, in order to allow other researchers to check analyses and conclusions"* (6).

- This would benefit patients by enabling better scrutiny of research.

**Transparency and data quality:** *"monitoring data quality and taking account of the origin and quality of data when drawing conclusions"* (6).

- Appropriate to include.
- Unsure why this guideline is categorised under "transparency" (rather than research quality, for example) unless it is specifically advising transparency (in reporting) on the limitations of research findings due to data quality.
- When considering the quality of data, consider representativeness and missingness of data. Consider also the potential impact of bias on data quality.

**Patient and Public Involvement:** *"incorporating the views of people with lived experience throughout the course of each project, and providing nuanced and high-quality public communication of findings"* (6).

- See also "Elements worth including with amendments".

**Elements worth including with amendments**

**Security, safe settings:** *"Ensuring that data are accessed in safe settings using clear and efficient procedures"* (6).

- In these guidelines, there is a lack of clarification over what is meant by "safe settings."
- This guideline should be included but with more clarity on what is meant by "safe settings."
- Do "safe settings" mean a virtual secure environment such as a TRE/SDE? Or do they mean a physical secure setting such as a SafePod which provides access to the TRE/SDE? The latter would be preferable for access to unconsented MH data.
- Procedures should be clear and efficient, but security should not be compromised for increased efficiency.

**Security breaches and near misses:** *"planning in advance to prevent data breaches, using a recording process for data breaches, and reporting near misses"* (6).

- There needs to be more information on planning to prevent data breaches—who is responsible for this planning? The data host/controller or the commercial organisations that are granted access to the data? If there is no direct transfer of data and if data access is via a TRE/SDE, and access to the TRE/SDE is via a safe room (e.g. SafePod), who would be responsible for security planning?
- All access to data should be via a TRE/SDE. There should be prevent and detect technical controls embedded within the TRE/SDE to minimise risks and detect incidents. There should also be procedural controls in data access processes and audit/review/monitoring processes. While this is not included for guiding current data access practices, there are guidelines on the "future" checklist (for the research community to help make implementable through the development of new protocols) which advise *"developing robust systems to prevent data leaks and record data breaches and near misses"* and *"incorporating inspection processes to ensure ongoing compliance with good data practice"* (6). It is not clear why both should not be guidelines for current data access practices. Clearly, there should be robust preventative controls (both technical and procedural) in place to reduce the risk of breaches, near misses and misuse. It is also not clear how breaches and near misses would be detected (which would need to occur for them to be reported) if the guidelines effectively defer detection controls and processes to an unspecified future date. Hence, there should currently be robust and effective technical detection controls and inspection processes.
- Unclear who would be responsible for reporting data breaches and near misses, and to whom.
- Other sets of guidelines do not refer to "near misses" and it is important to report on, understand and learn from near misses to improve security and minimise the risk of future breaches. Unclear from this guideline how near misses would be detected (via technical controls, review/monitoring procedures, or both?). The fact that "inspection processes" (review/monitoring procedures) have been designated as a future guideline suggests there would be reliance on technical detection controls.
- There is no mention of "data misuse" (by authorised personnel), which is another risk. There should be (technical and procedural) prevent controls to reduce the risk of data misuse and (technical and procedural) detection controls to detect occurrences of misuse.
- Even where there are technical and procedural detection controls, they will not necessarily detect all breaches, near misses or occurrences of misuse. Thus, it would be appropriate to have a formal whistleblowing process for researchers to confidentially report any breaches, near misses and potential misuse at their organisation with regards to data access.
- There is no mention of investigative processes, nor of learning from breaches and near misses, and modifying systems and processes accordingly.
  - There should be a formal response process for investigating incidents (including near misses), including how each incident occurred, and what could be learnt from it. There should be emphasis on learning from incidents.
  - For near misses, inevitably the depth of investigations may vary depending on the nature of the near miss.
  - Where appropriate, additional training requirements could be identified and necessary changes could be made to processes and systems to ensure that the risk of incidents re-occurring or the risk of incidents occurring for the first time (in the case of near misses) are minimised.
  - It needs to be clear who will be responsible for response/investigative processes.
  - There should be subsequent checks that identified actions have been implemented.
- No mention of accountability measures for when things go wrong in guidelines for current practices, which should be clearly specified.
  - While accountability measures are not included for guiding current data access practices, there is a guideline on the "future" checklist (for the research community to help make implementable through the development of new protocols) which advises *"responding*

*proportionately to inappropriate data use with measures such as training or temporary or long-term suspension of access"* (6). It is not clear why this should not be a guideline for current data access practices. Note also that a range of things can go wrong beyond "inappropriate data use."
    - There need to be clear indications of what sanctions will be imposed and in what situations. Procedures should clearly specify what steps will be taken in the event of a security breach or incident of data misuse, including in what situations data access would be revoked for organisations and how this would be enforced.
- No mention of transparency. There needs to be transparency to allow public scrutiny and accountability. Information on all breaches and incidents of data misuse should be made available to the public (obviously without further compromising security) including details on the impact of the incident and any remedial actions taken. There should not be a culture of cover-up, as appears to be the case in many aspects of the healthcare system. Depending on the nature of the near miss, it may not be appropriate to report this as this may compromise security (e.g. in the case of near misses for which preventative solutions cannot be immediately implemented).
- There is a culture of cover-up within many NHS trusts whereby the reputation of institutions is prioritised over patients, patient safety and transparency (125,126). When serious harms to patients are covered up, how can patients have faith that security breaches (and other incidents) with regards to patient data will be publicly reported? If the loss of lives is covered up, will the loss of megabytes be reported? For incidents pertaining to patient data, there is a need for a guideline to address this cover-up culture. For example, to reduce the risk of cover-ups, a guideline may mandate that the consequences for not reporting breaches, near misses or data misuse in a timely manner, that later come to light (e.g. via an audit, monitoring, or employee whistle-blowing) should be relatively severe. Depending on the party responsible (for the incident and/or cover-up), such consequences may include (among other penalties) the revocation of data access, restrictions on future patient data access, higher fines, and legal action.

**Ensuring privacy and anonymity while working with data:** *"using de-identified data, except where identifiable information (including information about protected characteristics) is essential to beneficial outcomes"* (6).

- Given the high sensitivity of MH data, arguably commercial organisations should not be provided with any identifiable data, regardless of the purported public benefit. Hence, for commercial access to MH data, only de-identified data should be accessed and used.
- Even in cases requiring data linkage (when this is absolutely necessary) there should not be any need for identifiable data to be shared and de-identified data linkage should be possible.
- There is a guideline on the "future" checklist (for the research community to help make implementable through the development of new protocols) which advises *"building plans for de-identified data collected by researchers, scientists and clinical services to be made available for analysis on an open-access basis"* (6). Unconsented MH data should never be made available on an open-access basis. Even when MH data is de-identified, there is always some identifiability risk, especially for free text MH data.
- Given the substantial concerns regarding the sharing of free text MH data, commercial entities should not be given access to this type of data. Please see review of UPD content (**Supplementary Table 9,** "Elements worth including with amendments") for further information on concerns related to the sharing of unconsented free text MH data.

**Anonymity, statistical disclosure control, and research outputs (findings, published data):** There are a few guidelines pertaining to this, including (i) *"researchers, scientists and clinical services making de-identified data and findings (including null results) open-access where possible. It also involves awareness of the risk that qualitative data (such as free text) could contain identifiable information"*; (ii) statistical disclosure control should be incorporated *"by following rules designed to prevent identification of individuals"*; and (iii) *"incorporating statistical disclosure control based on* *principles, such as the principle that no individual may be identified, with training and external oversight"* (6).

- There should be open-access publication of research. This would benefit patients by enabling better scrutiny of research and helping to address issues of reproducibility and replicability in research.
- However, due care must be taken with the publication of patient MH data. There should be a strong requirement to ensure that all published data is anonymous to an expected publishable standard.
- The reference to "statistical disclosure control" would include methods to prevent identification of individuals at various stages of the data lifecycle including in published outputs (statistical output control).
- It is unclear why (iii) is designated as a future guideline rather than a current guideline. It should absolutely be ensured that no individual can be identified from published research data in current data access practices. (iii) may be designated as a future guideline because it is currently difficult to ensure perfect de-identification throughout the data lifecycle. However, this is not clear, and because statistical disclosure control includes statistical output control, this may be interpreted to mean that the latter is not currently mandated.
- Commercial entities should not have access to unconsented free text MH data, and therefore commercial entities should not have access to publish such data anyway. Please see review of UPD content (**Supplementary Table 9,** "Elements worth including with amendments") for further information on concerns regarding the sharing of unconsented free text MH data.
- Where automatic statistical output control (technical controls to ensure that data output from TRE/SDEs is suitably anonymised) is used, all outputs should also be checked manually. Automatic statistical output control should be assistive and procedural controls must ensure that undue reliance is not placed on automatic checking.
- There should be appropriate staff training and oversight to ensure the effective implementation of statistical disclosure control.

**Anonymity and data linkage:** There are two guidelines pertaining to this: (i) *"developing effective measures, including secure linking systems, to protect against inappropriate identification and misuse"* and (ii) *"facilitating research linking mental health data with other sources of public data, such as education or welfare data, in order to provide new information of public benefit about mental health"* (6).

- (i) is designated as a future guideline rather than a current guideline, and it is unclear why.
- There should be effective measures taken currently to significantly reduce the risk of inappropriate identification and data misuse for all MH data access.
- While data linkage of MH data with other public data may indeed generate new insights about MH, it is important to recognise that these guidelines do not specifically relate to commercial access to MH data (and hence commercial access to data linkage of unconsented MH data with other unconsented public data).
- There should be due consideration that unconsented MH data is highly sensitive and with a higher degree of identifiability than physical health data (even when de-identified, as perfect de-identification cannot be assured and de-identified data is not the same as anonymous data). If unconsented MH data is linked with other unconsented public data, the degree of identifiability would increase. Thus, commercial organisations should not have access to link unconsented MH data with other unconsented public data unless absolutely necessary and in the public interest to do so, determined with PPI. All such commercial linkage projects should be disclosed on a public registry in an accessible way, to enable public scrutiny.
- It is also important to recognise that allowing commercial organisations to potentially link unconsented MH data with other unconsented public data would make this practice significantly more widespread as there are an enormous number of commercial organisations. Thus, any commercial request for MH data linkage must be rigorously considered, and only approved where it is appropriate to do so. There should be a complete public registry of all

commercial linkage projects linking unconsented MH data with other unconsented public data, to enable public scrutiny and appropriate public challenge.

- In some cases, it may be appropriate to only approve data linkage access where commercial organisations partner with a university, whereby only the university has access to link unconsented MH data with other unconsented public data.
- Where data linkage is approved, there should not be any need for identifiable data to be shared and de-identified data linkage should be possible. It is currently possible to perform de-identified data linkage, and this should be mandated.

**Patient and Public Involvement:** *"incorporating the views of people with lived experience throughout the course of each project, and providing nuanced and high-quality public communication of findings"* (6).

- This pertains to PPI throughout the research project for which data access is approved.
- There is a similar guideline in the future guidelines list which combines a few different topics, causing some confusion: *"incorporating the views of people with lived experience throughout the course of each project. At the same time the principles of open access should be followed by publicly pre-registering studies and providing accessible online information of each overarching request to use data, each output, and any null results"* (6). This single guideline refers to PPI on research projects, as well as the importance of open-access research, and transparency over projects with data access. Aside from the different topics, the various aspects of this guideline are for different audiences—for example, it would be for the data access applicant to ensure PPI on the research project, while it would be for the data host/controller to ensure transparency over who is awarded data access [*"providing accessible online information of each overarching request to use data"* (6)]. Ensure each guideline addresses a distinct area and is intended for a specific audience. Ensure each guideline is unambiguous and clear.
- There are no guidelines that refer to PPI in decision-making on MH data access requests, nor with regards to any data access process.

**Noted to avoid** **Requiring lived experience members involved in guidelines development to have data science experience:** Requiring all lived experience participants involved in developing guidelines to also have "expertise in data science" (or research experience) as was the case in this study (7).

- The reason given for this was so that "the participants themselves were in a position to weigh up the relative merits of the information from both" mental illness and data science perspectives and so that there was reduced "need for researcher involvement in handling potential trade-offs" (7).
- Arguably, when participants have "expertise in data science" and/or relevant research experience, this means they are significantly more likely to be pro-data research and pro-data sharing for research purposes. Such participants may have been less likely to critically analyse and consider various issues related to the sharing of unconsented MH data. Certainly, such participants would not be representative of the wider population living with mental illness.
- It is highly unlikely that the perspectives of those who are against or who are more cautious about the sharing of mental health data were included. This may mean that important public concerns were missed.
- It is important that lived experience members with a diverse range of perspectives on data sharing are included.

**Separate "today" and "future" checklists/guidelines.**

- While this can be useful for examining change around practices and for understanding the expectations those with lived experience have for future practices, it can also cause confusion when guidelines are implemented. For example, the distinction between what applies now and what is an expectation for the future can become blurred.

- Under "How to use the checklist" it does state that researchers *"conducting new studies should consider each item on the today checklist"* while the research community should *"aim to develop and disseminate new protocols to meet the criteria for best practice in the future"* (6). Therefore, arguably, there are clearly differentiated purposes for each checklist, whereby the "today" checklist is for guiding current data access practices, while the "future" checklist is for influencing the development of tools/protocols/methods which enable the "future" guidelines to be implementable. However, these different purposes are not clearly demarcated on the guidelines themselves. If someone were to only read the guidelines and miss the section on "How to use the checklist", it would also not be apparent why "future" guidelines are designated for the future and why they are not immediately implementable.
- It is not clear what is meant by "the future". Does this mean in 6 months, 1 year, 5 years, or another time frame? Unless some kind of timeline is specified for the implementation of the future guidelines, it is unclear what benefits their inclusion provides.
- What if there are changes in technology, or changes in the law, or changes in public expectations? Surely these would impact any guidelines for the future?
- To avoid confusion, there should be a clear set of guidelines that apply now, that are publicly available. When there are changes to the guidelines, the revised guidelines should clearly state the date from which they apply.
- Any information about expectations for the future can be maintained in internal documents, separate from current guidelines. Such a document may be useful for guiding further revision of the guidelines, but should not pre-empt future versions of the guidelines. Alternatively, if future guidelines are to be publicly available, there should be a clear, unambiguous, and explicit indication of their purpose and intended audience.

**Incomplete guidelines that are heavily complemented by other/existing frameworks or guidelines.**

- These guidelines are sparse, contain little detail, and are *"designed to complement other guidance regarding good practice within data science"* (7), including:
  - The UK Data Ethics Framework (25);
  - The UK Government's Code of Conduct for Data-Driven Health and Care Technology (26);
  - Development of data governance for the use of clinical free-text data (27).
- While it is perhaps inevitable for all guidelines to complement some existing guidance, care should be taken to ensure that all key principles and guidelines are contained within the new guidelines' documentation, and where applicable, references can be made to other existing guidelines or frameworks. Such references should be in the main body of the guidelines rather than in the footnotes or small print. Otherwise, there is a risk of having fragmented guidelines scattered in various places that no one treats as a unified whole, which may impact implementation. For example, a given organisation may only refer to the new guidelines and not all the existing frameworks and guidelines that the new guidelines are supposed to complement. Further, when references to complementary guidance are not made in the main guidelines text, there is a lack of indication of what is considered particularly important and relevant in the existing guidance and frameworks.
- Confusion can arise when new guidelines appear to contradict existing guidance which they are apparently supposed to complement. For example, in this case, the UK Data Ethics Framework advises transparency and accountability, among other criteria for projects using public data, but arguably this may be eroded if unconsented patient MH data is provided on an open-access basis, as apparently proposed by these guidelines (25).
- There are other cases where existing guidance may not be completely relevant anymore—for example, the UK Government's Code of Conduct for Data-Driven Health and Care Technology refers to data linkage using identifiable data, and it should be possible to carry out data linkage using non-identifiable data (26).

*"Data should be accessible to a range of people who conduct research, including authorised academics and clinicians"* (6).

- While there is nothing inherently incorrect about this statement, it does not appear appropriate as a guideline. To include a guideline which advises that a diversity of applicants gain data access risks undermining and potentially pre-empting certain outcomes from the formal access request approvals process. Data should only be accessible to those whose formal access requests have been appropriately approved, with PPI, based on assessment criteria.
- This guideline appears under the category "security" and it is not clear why this is; there is a lack of clarification on why this would be a "security" guideline.

**Data management plans:** *"creating data management plans and ensuring that these are adhered to at all times"* (6).

- A data management plan is a formal documented plan that outlines how data will be handled during and after a research project (essentially throughout the research lifecycle). Such a plan may include aspects such as data collection, data storage, and data security.
- It is not clear what is intended by this guideline and for whom it is intended, in the context of data access to unconsented patient MH data. For example, a commercial applicant for access to unconsented patient MH data would not be responsible for data collection, storage, or security.
- The data host/controller would be responsible for ongoing data management that would not be limited to a single research project and hence the term data management plan (which pertains to a single research project) may be misleading when applied to a data controller.
- More clarity would need to be provided and unambiguous terms used, before the usefulness of this guideline can be determined.

**Remote access to data:** *"providing digital controls to allow remote access from private settings, using procedures that are robust and easy to follow"* (6).

- This guideline has been included on the "future" checklist, meaning that it has not been designated for guiding current data access practices but is instead for the research community to help make implementable through the development of new protocols.
- While all access to data would be remote in that researchers should obviously not have direct server access, there are conceivably different risks posed by data access from different remote settings. The term "private settings" in this guideline suggests at-home access. Arguably, at-home access would introduce more risks such as other non-authorised people gaining access to data and connected devices getting lost or stolen. Further, there would be fewer deterrents to a user taking photos of data at home (as there is no one around them to observe their behaviour) for the purpose of re-identifying individuals compared to at an NHS site or workplace.
- While it is appreciated that SafePods are relatively expensive, such a physical secure setting which provides access to the virtual TRE/SDE, would reduce risks and provide reassurance. For example, users would not be able to take photos of data (not without this being recorded) and access to the pods themselves would be controlled.
- Given the sensitivity of unconsented MH data, at-home (remote) access to data should not be provided. In the future, there may be technical controls which can allow at-home access in a way that reduces risks.

**Access to identifiable data:** *"using de-identified data, except where identifiable information (including information about protected characteristics) is essential to beneficial outcomes"* (6).

- Given the high sensitivity of MH data, commercial organisations should not be provided with any identifiable data, regardless of the purported public benefit. Hence, for commercial access to MH data, only de-identified data should be accessed and used.

**Open access to unconsented MH data:** *"building plans for de-identified data collected by researchers, scientists and clinical services to be made available for analysis on an open-access basis"* (6).

- Unconsented MH data should never be made available on an open-access basis.
- Even when MH data is de-identified, there is always some identifiability risk, especially for free text MH data. It is not possible to provide assurance of de-identification; "perfect" de-identification in all cases would be impossible. Note also that de-identification is not the same as anonymisation.
- Even if free text MH data is not shared, it would not be at all appropriate for unconsented MH data which is highly sensitive and private to be made available on an open-access basis. De-identified data would not be the same as anonymous data. The knowledge of their highly private MH data being shared on an open-access basis would have an adverse impact on many patients. Further, there would be no vetting whatsoever of any party that gains data access, which could cause many potential harms.

**Transparency and withdrawal of consent:** *"ensuring that researchers have a process in place for responding to withdrawal requests and that they provide transparency on whether, how and when participants can withdraw their data"* and *"appointing a qualified, independent arbiter to arbitrate on complex questions relating to consent and data withdrawal"* (6).

- These guidelines appear to be for research trials data, providing a means for participants who have explicitly provided consent for data sharing to withdraw that consent.
- These guidelines would not apply for unconsented MH data, as in most cases affected patients would not even know that their data is being used in research, and they would not have provided explicit consent (and hence consent cannot be withdrawn).

**Anonymity and data linkage:** *"facilitating research linking mental health data with other sources of public data, such as education or welfare data, in order to provide new information of public benefit about mental health"* (6).

- While data linkage of MH data with other public data may indeed generate new insights about MH, it is important to recognise that these guidelines do not specifically relate to commercial access to MH data (and hence commercial access to data linkage of MH data with other public data).
- There should not be undue eagerness to facilitate commercial access to data linkage of unconsented MH data with other unconsented public data. A guideline worded in this way may pre-empt decision-making on data access approval processes, leading to inappropriate access.
- Any commercial request for MH data linkage must be rigorously considered, and only approved where it is appropriate and in the public interest to do so, determined with PPI.
- There should be due consideration that unconsented MH data is highly sensitive and with a higher degree of identifiability than physical health data (even when de-identified, as perfect de-identification cannot be assured and de-identified data is not the same as anonymous data). If unconsented MH data is linked with other unconsented public data, the degree of identifiability would increase.
- It is also important to recognise that allowing commercial organisations potentially to link unconsented MH data with other unconsented public data would make this practice significantly more widespread, as there are an enormous number of commercial organisations.
- All approved commercial linkage projects linking unconsented MH data with other unconsented public data should be disclosed on a public registry in an accessible way, to enable public scrutiny and appropriate public challenge.

**Developing innovative de-identification methods:** *"developing methods for de-identification, including innovative ways to mask identifiable information"* (6).

- This is designated as a future guideline rather than a current guideline.

- While innovative de-identification methods may enable better de-identification throughout the data lifecycle and potentially enable access to more data, it is not in scope for guidelines aimed at host organisations and commercial applicants, nor for guidelines focused on the current sharing of MH data.

**Monitoring quality of data repositories:** *"incorporating oversight of data repositories in order to monitor data quality and respond to public enquiries"* (6).

- Not in scope for [DATAMIND LEAG] guidelines focused on the current sharing of MH data.

**Participant consent:** *"ensuring that participants have as much control over consent as possible"* (current guideline) and *"exploring alternative models of consent, which may involve moving away from individualised models of consent"* (future guideline) (6).

- These guidelines appear to be for research trials data, with the current guideline concerned with participants who provide explicit (individualised) consent for data sharing.
- This would not be relevant for the sharing of unconsented MH data.

**Reducing stigma:** *"active commitment to reducing the stigma associated with mental illness and its research, and to increasing public understanding of science"* (6).

- This guideline has been included on the "future" checklist, meaning that it has not been designated for guiding current data access practices but is instead for the research community to help make implementable through the development of new protocols.
- While there would inevitably be benefits to this, it is not really in scope for current guidelines relating to data access/use practices.

**Researcher knowledge:** *"ensuring that data users understand the underlying data collection tools as well as the socio-cultural context in which studies are designed and findings are disseminated"* (6).

- This appears to be more relevant for research trials. Not sure if relevant in the context of DATAMIND LEAG guidelines.

**Gaps**

- Glossary—for terms used in the guidelines, clarifying what they mean in the context of the guidelines.
- Clarity on what constitutes a safe setting.
- Consideration of data misuse.
- Technical and procedural preventative and detection controls embedded within TRE/SDE to minimise risks and detect incidents.
- Whistleblowing process.
- Investigative process for breaches, near misses and misuse—to investigate any incident, how it occurred, and what could be learnt from it.
- Learning from incidents and using this to improve processes and systems.
- Accountability measures, whether revocation of data access, other sanctions, legal consequences, public accountability, or other measures.
- The need for transparency and making information on data breaches, near misses and data misuse available to the public (obviously without further compromising security). This could be via a public registry.
- Addressing cover-up culture to enable greater transparency on incidents.
- Sufficient coverage of free text MH data and concerns with regards to sharing such data.
- Clear guideline on statistical disclosure control which is applicable for current data access.
- Requirement for all automatic statistical output control outputs to be manually checked.
- Consideration of risks and concerns relating to data linkage of unconsented MH data with

| | |
|---|---|
| | other unconsented public data.<br>• Adequate coverage of risks and concerns related to identifiability.<br>• Opt-out for use of unconsented MH data in research.<br>• Requirement for data access processes to be formally documented and publicly accessible.<br>• Requirement for PPI in data access processes, including decision-making on access requests and monitoring of data use.<br>• Other gaps in addition; not all listed. |
| **Public involvement approach** | **Participation and drop-out:** 20 participants with lived experience of mental illness were involved in all three stages of the Delphi study. 30 participants were recruited, but there was drop-out of 10 participants.<br>• This may have been due to the Covid-19 pandemic and lockdown.<br>• Note also, that participants were given 8–11 days to complete each phase of the Delphi study, and given that 80% of participants were suffering from mental illness at the time of the study, the short window of completion for each phase may have affected participation.<br>• It is important to take proactive measures to ensure inclusion and continued involvement of lived experience members.<br><br>**Requiring lived experience members involved in guidelines development to have data science experience:** While the twenty participants had "personal experience of mental illness", it is important to note that they were required to also have "expertise in data science" (which may include research experience) (7).<br>• The reason given for this was so that "the participants themselves were in a position to weigh up the relative merits of the information from both" mental illness and data science perspectives and so that there was reduced "need for researcher involvement in handling potential trade-offs" (7).<br>• Arguably, when participants have "expertise in data science" and/or relevant research experience, this means they are significantly more likely to be pro-data research and pro-data sharing for research purposes. Such participants may have been less likely to critically analyse and consider various issues related to the sharing of unconsented MH data. Certainly, such participants would not be representative of the wider population living with mental illness.<br>• It is highly unlikely that the perspectives of those who are against or who are more cautious about the sharing of mental health data were included. This may mean that important public concerns were missed.<br>• It is important that lived experience members with a diverse range of perspectives on data sharing are included.<br>• This does not mean that no public participants with data science (or research) experience should have been recruited in a study of this nature, but rather that making this a condition of participation is problematic and counteracts one of the advantages of conducting a Delphi study—that of including diverse perspectives and *subsequently building* consensus through several iterations. It is important to have the involvement of those with diverse perspectives and equal representation of each broad perspective because what "passes" each phase in a Delphi study is based on consensus opinion.<br><br>**Involvement and inclusion of those with diverse perspectives:**<br>• In this project, it is highly unlikely that the perspectives of those who are against or who are more cautious about the sharing of mental health data were included.<br>• Some characteristics of some mentally ill people may mean that they are well placed to consider a diverse range of risks for a given endeavour:<br> ○ Some people with mental illness may have substantial experience of considering risks for diverse situations. |

    - A relatively large proportion of people in therapy (50%) have been found to be highly sensitive people, which means that they perceive a lot more than the average person and process what they perceive very deeply.
- Thus, involving such individuals would not only enable the views of affected individuals to be shared, but may enable previously neglected risks to be considered which may help ensure appropriate safeguards are put in place.
- However, diversity is not the same as inclusion. It is insufficient to only recruit diverse members with lived experience; adequate measures must be taken to actively include such members.
- Some mentally ill people may have been (previously) extensively criticised or shamed for considering risks. There is continued stigma around mental illness, and this includes an aversion towards behaviours that include identifying risks or issues, and different ways of thinking. There is a lot of pressure to be agreeable and always in agreement with others, to the extent of repressing actual thoughts. This can mean that those with lived experience feel that they cannot share what they really think and feel about a given issue. Hence, in addition to involving those with lived experience, it is important that:
    - Those with a diverse range of opinions on data sharing are proactively welcomed, recruited, and included;
    - An inclusive and safe space is created to enable those with marginalised or dissenting perspectives on data sharing to express such views;
    - There is mindfulness around the difficulties that some members may feel in sharing their perspectives, with proactive measures taken to encourage such members to share.

**Development of initial list of Delphi statements:** The researchers, together with a group of stakeholders, *"four people with lived and professional experience of mental illness and two psychiatry researchers"*, decided the statements that were used in the first phase of the Delphi study (7).

- Arguably, there was a lack of sufficient involvement of those who only had lived experience of mental illness in putting together the initial list of statements.
- It is important to note that the initial list of statements may have framed thinking and perspectives on the study, introducing some bias or influence. While study participants could use the free text option to recommend the inclusion of further statements, the relatively homogenous group of participants and the way in which the initial list of statements were compiled, may mean that important public concerns were missed.
- Indeed, scanning the initial list of statements, it seems that some areas have been omitted, notably:
    - Auditing/reviewing/monitoring the actual access and use of data;
    - Whether access should be restricted by type of data e.g., no access for certain parties to free text data;
    - Requirements relating to the data access request and approvals process;
    - Industry/commercial access to data.

The study states that the checklist encapsulates *"the perspective of people with lived experience of mental illness, without making recommendations that contravene existing data science frameworks"* (7). This may suggest that the parameters around the study involved not questioning the validity or appropriateness of existing frameworks/codes.

Guidelines referencing the public or PPI:

- Prioritising *"the health and benefit of people with lived experience"* in determining the use of MH data for research (6).
- Incorporating *"the views of people with lived experience throughout the course of each project, and providing nuanced and high-quality public communication of findings"* (6).

| | |
|---|---|
| | • Guidelines relating to consent and withdrawal (of consent) for participants in research trials (not applicable for unconsented MH data).<br>• Guideline referencing responding to public enquiries.<br><br>There are no guidelines that refer to PPI in decision-making on data access requests, nor with regards to any data access process. |
| **Industry involvement approach** | No industry involvement in developing guidelines.<br><br>No specific consideration of industry access to unconsented MH data and the unique risks and concerns this may present.<br><br>No reference to requirements for commercial organisations in the guidelines. For example, there is no guideline on what information a commercial organisation should provide when requesting MH data access. |
| **Equality, diversity, and inclusion considerations** | Those with lived experience of mental illness were involved in developing the guidelines.<br><br>Inclusion and exclusion criteria for study participants mean it is highly unlikely that the perspectives of those who are against or who are more cautious about the sharing of mental health data were included. Hence, lived experience members with a diverse range of perspectives on data sharing were not included.<br><br>Apart from guidelines advising the involvement of those with lived experience of mental illness on data research projects, there are no guidelines pertaining to EDI. |
| **Other considerations** | – |
| **Summary of strengths and limitations** | Strengths:<br>• Those with lived experience of mental illness were involved in developing the guidelines. 80% of participants were "living with a mental illness at the time of the study" (7).<br>• Guidelines specifically focused on MH data.<br><br>Limitations:<br>• The lived experience participants were required to also have "expertise in data science" (or research). Thus, they would not be representative of the wider population living with mental illness. It is highly unlikely that the perspectives of those who are against or who are more cautious about the sharing of MH data were included.<br>• No specific consideration of the risks and concerns related to commercial access to unconsented MH data.<br>• Omission of important public concerns with regards to the sharing of unconsented MH data.<br>• Lack of detail for each guideline.<br>• Insufficient coverage of free text MH data and concerns with regards to sharing such data, especially with commercial organisations.<br>• Lack of detail for each guideline.<br>• (Not all covered.) |

#### 4 SUPPLEMENTARY ABBREVIATIONS

| | |
|---|---|
| # | number |
| ADR UK | Administrative Data Research UK (funded by the ESRC) |
| AI | artificial intelligence |
| BHF | British Heart Foundation |
| CAG | UK Confidentiality Advisory Group (127). Created in 2013 to replace the National Information Governance Board (NIGB). |
| CCTV | closed circuit television |
| CI | chief investigator |
| COVID-19 | coronavirus disease 2019 |
| CPIR | The Health Service (Control of Patient Information) Regulations 2002 (102) |
| CRUK | Cancer Research UK |
| CSO | UK Chief Scientific Officer (or Chief Scientist Office) |
| CT | computerised tomography (a form of scan) |
| CTIMP | Clinical Trial of an Investigational Medicinal Product |
| DARE UK | Data and Analytics Research Environments UK (funded by UKRI, HDR UK, ADR UK) |
| DATAMIND | DATA hub for Mental health INformatics research Development |
| DNA | deoxyribonucleic acid |
| DOB | date of birth |
| DPA | UK Data Protection Act 2018 (59) |
| EDI | equality, diversity, and inclusion |
| EHR | electronic health record (also: EPR) |
| EPR | electronic patient record (also: EHR) |
| EPSRC | Engineering and Physical Sciences Research Council, part of UKRI |
| ESRC | Economic and Social Research Council, part of UKRI |
| EU | European Union |
| FOMO | fear of missing out |
| GDPR | EU General Data Protection Regulation (2016) (89) |
| GP | general practitioner / general practice |
| HDR UK | Health Data Research UK |
| HRA | UK NHS Health Research Authority |
| HSC | Health and Social Care (Northern Ireland) |
| HSC-PBPP | Public Benefit and Privacy Panel for Health and Social Care (Scotland) |
| ICO | UK Information Commissioner’s Office (128) |
| ID | identity/identifier |
| IG | information governance |
| IRAS | Integrated Research Application System (129) |
| LEAG | DATAMIND Lived Experience Advisory Group (formerly Super Research Advisory Group, SRAG) |
| LLM | large language model |
| MH | mental health |
| ML | machine learning |
| MRC | Medical Research Council, part of UKRI |
| MRI | magnetic resonance imaging (a form of scan) |
| NDO | NHS National Data Opt-Out |

| | |
|---|---|
| NHS | National Health Service (England, Scotland, Wales) or HSC (Northern Ireland) |
| NIHR | UK National Institute for Health and Care Research |
| NLP | natural language processing |
| ONS | UK Office for National Statistics |
| PI | principal investigator |
| PPI(E) | patient and public involvement (and engagement) |
| R&D | research and development |
| REC | Research Ethics Committee |
| RES | Research Ethics Service (formerly National Research Ethics Service), hosted by the HRA and the UK Devolved Administrations |
| SDC | statistical disclosure control |
| SDE | secure data environment (a broader term for a TRE) |
| TRE | trusted research environment (see also SDE) |
| UI | user interface |
| UK | United Kingdom |
| UKRI | UK Research and Innovation, a public body uniting the UK's seven research councils (which provide public funding for research) |
| UPD | Understanding Patient Data |

### 6 SUPPLEMENTARY FIGURES

#### *6.1 Supplementary Figure 1: Types of information found in medical records and corresponding research databases*

Types of information found in medical records and corresponding research databases, with UK legal status (focusing on England and Wales; see also **Supplementary Table 4**). Typically, researchers seek to operate with pseudonymised or anonymised health data, prior to publishing aggregated anonymous data (see **Figure 1**). (All examples are fictional and the fictional NHS numbers are from the official test range.) Modified from (38).

(Figure overleaf.)

Supplementary Figure 1

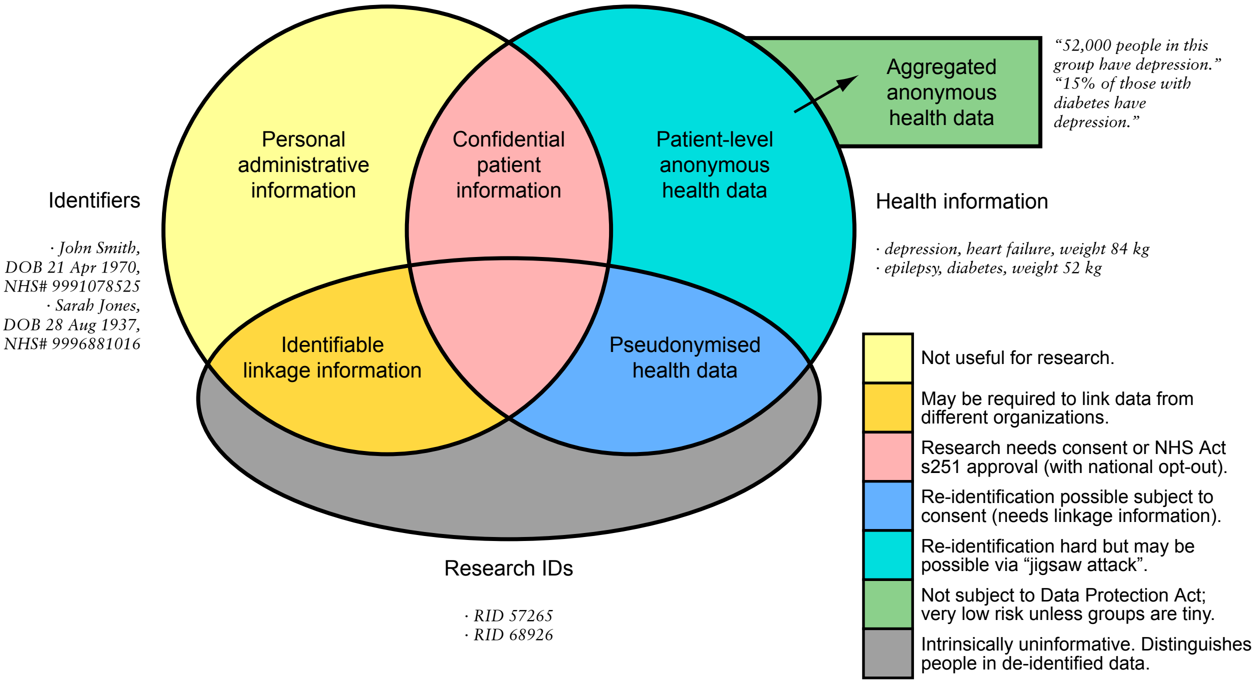
